# High seroprevalence but short-lived immune response to SARS-CoV-2 infection in Paris

**DOI:** 10.1101/2020.10.25.20219030

**Authors:** François Anna, Sophie Goyard, Ana Ines Lalanne, Fabien Nevo, Marion Gransagne, Philippe Souque, Delphine Louis, Véronique Gillon, Isabelle Turbiez, François-Clément Bidard, Aline Gobillion, Alexia Savignoni, Maude Guillot-Delost, François Dejardin, Evelyne Dufour, Stéphane Petres, Odile Richard-Le Goff, Zaineb Choucha, Olivier Helynck, Yves L. Janin, Nicolas Escriou, Pierre Charneau, Franck Perez, Thierry Rose, Olivier Lantz

## Abstract

Although the COVID-19 pandemic peaked in March/April 2020 in France, the prevalence of infection is barely known. Herein, we assessed using high-throughput methods the serological response against the SARS-CoV-2 virus of 1847 participants working in one institution in Paris.

In May-July 2020, 11% (95% CI: 9.7-12.6) of serums were positive for IgG against the SARS-CoV-2 N and S proteins and 9.5% (CI:8.2-11.0) were pseudo-neutralizer. The prevalence of immunization was 11.6% (CI:10.2-13.2) considering positivity in at least one assays. In 5% (CI:3.9-7.1) of RT-qPCR positive individuals, no systemic IgGs were detected. Among immune individuals, 21% had been asymptomatic. Anosmia and ageusia occurred in 52% of the IgG-positive individuals and in 3% of the negative ones. In contrast, 30% of the anosmia-ageusia cases were seronegative suggesting that the true prevalence of infection may reach 16.6%. In sera obtained 4-8 weeks after the first sampling anti-N and anti-S IgG titers and pseudo-neutralization activity declined by 31%, 17% and 53%, respectively with half-life of 35, 87 and 28 days, respectively.

The population studied is representative of active workers in Paris. The short lifespan of the serological systemic responses suggests an underestimation the true prevalence of infection.

## INTRODUCTION

Severe acute respiratory syndrome coronavirus 2 (SARS-CoV-2) causing the coronavirus disease 2019 (COVID-19) emerged in 2019 in China [1-3] before being detected in a patient living in the Paris conurbation in December 2019 [4]. The virus spread exponentially from January 2020 leading to a risk to saturate the intensive care units of Paris conurbation hospitals. Accordingly, on March 17^th^, a lockdown was imposed by the French authorities to slow down virus progression. To date, the exposure of the French population remains poorly documented. In contrast with RT-qPCR assays which are positive for only 2-3 weeks after infection [5], a more efficient way to monitor virus propagation is a serological study of representative populations since specific and lasting antibodies are generated in the great majority of infected subjects [6, 7]. However, studying the anti-SARS-CoV-2 serological response of large cohorts is challenging and require robustness, sensitivities and large dynamics range of the measurement methods exceeding the performance of currently marketed serological assays.

Here, we developed two original bioluminescence-based serological assays allowing a high throughput assessment of the specific antibody responses to the Spike (S) and Nucleoprotein (N) proteins of SARS-CoV-2 and their ability to neutralize the virus fusion with a permissive human cell line.

We monitored individual serology against SARS-CoV-2 in a large cohort of workers in three institution sites following the March-April 2020 peak of the COVID-19 pandemic in Paris (France) and over the next six months. More than half of Institut Curie workers (n=1847), a hospital and research center specialized in oncology, volunteered for this Curie-O-SA longitudinal serological study. The participants have been marginally in contact with COVID-19 patients and are domiciled in the Paris conurbation. This cohort is thus representative of an urban population of healthy active adults of a big metropolitan area. We found a high prevalence of immunization but rather short-lived responses.

## METHODS AND MATERIALS

### PARTICIPANTS AND WEB-BASED QUESTIONNAIRE

This study was registered and received ethical approval by the Comité de Protection des Personnes Méditerranée III (2020.04.18 bis 20.04.16.49458, 27/4/2020) registered in the clinical trial database (NCT04369066). Following informed consent, 18 years of age or older volunteer participant outside of any SARS-CoV-2 acute infectious episode in the last 7 days, working at one of the three Institut Curie locations (Paris, Orsay or Saint Cloud) completed a web-based questionnaire (Ennov Clinical) detailed in **Supplementary Appendix 1 and 2**. A 5 mL blood sample was taken from all participants in dry tubes. After clotting, blood was centrifuged 10 min at 2000 g. Supernatant serum was separated and frozen.

### SARS-CoV-2 SPECIFIC IgG AND PSEUDO-NEUTRALIZATION ASSAYS

Development and validation of the methods are described in the methods section of the **Supplementary Appendix 1**. Briefly, the sera to be tested were thawed and distributed into 96 well plates at the Institut Curie. An aliquot of a pool of positive (or negative) sera was distributed into 6 wells of each 96 well plate in a unique combination allowing an unambiguous identification of the plate.

These positive and negative wells were used to control for any drift of the measurements. The plates were then assessed at the Institut Pasteur using an ELISA-based methods for specific IgG assays (**Supplementary Appendix Figure S1-2**) [8-10] and an inhibition assay of neutralization of pseudotype-cell fusion by serum contents (**Supplementary Appendix Figure S3**) [11] described in the supplementary methods section. For the **Figure 3** longitudinal analysis, the *t*_*0*_ and *t*_*1*_ samples were analyzed at the same time from frozen serum samples.

**Figure 1:**
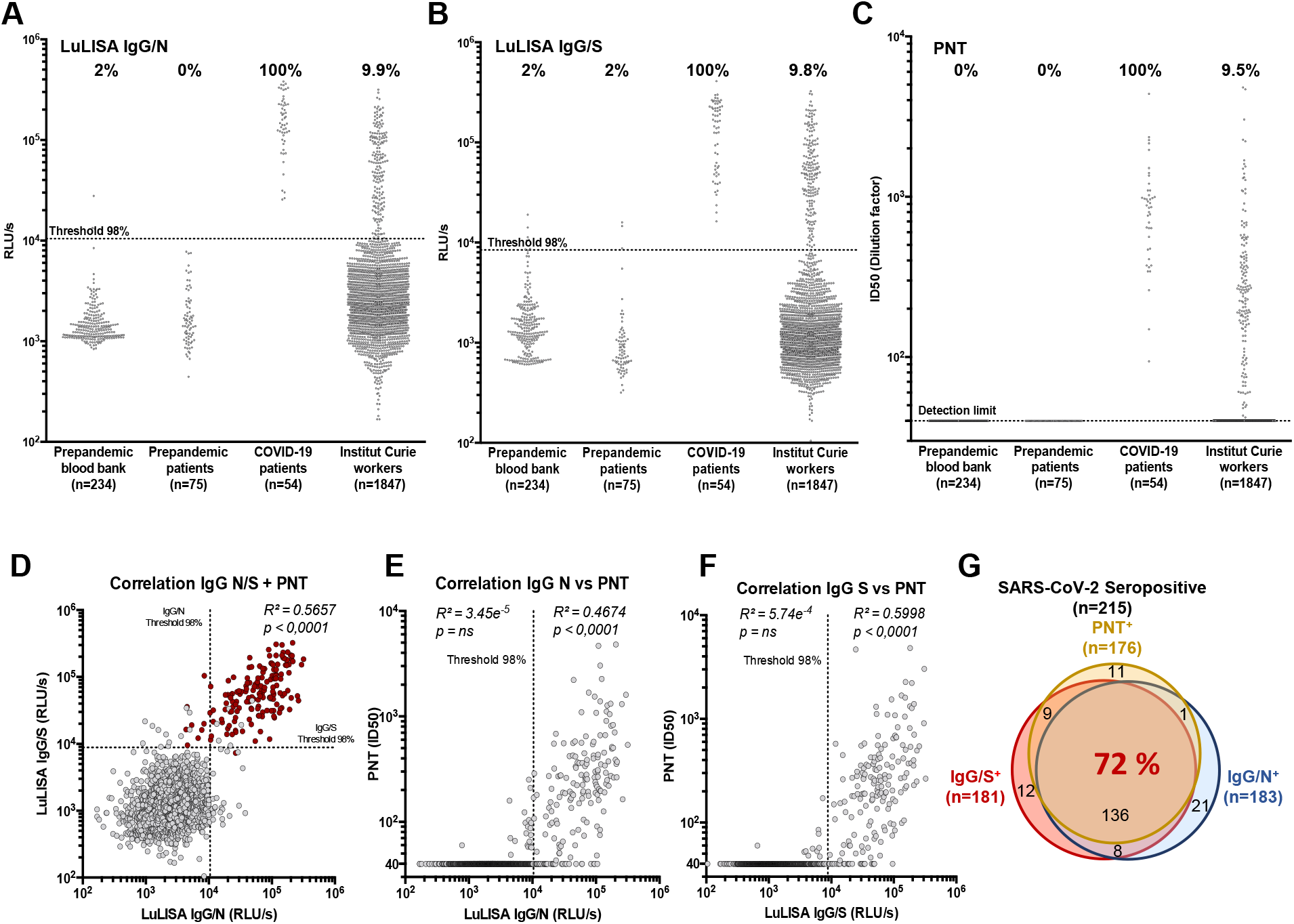
Serological responses to SARS-CoV-2 among Institut Curie workers using LuLISA IgG/N, IgG/S and PNT assays. **(A-C)** Sera from prepandemic samples from a blood bank, prepandemic patients (Breast Cancer), COVID-19 patients (RT-PCR positive) and Institut Curie workers were evaluated in LuLISA IgG/N **(A)** or IgG/S **(B)** and PNT **(C)** assays. For LuLISA, raw values are represented. Sera were considered positive for anti-N or -S IgG if the value was above the 98% threshold (See **Supplementary Appendix 1 Figure S1** for calculation details). For PNT assay, values after ID50 calculation are represented (See **Supplementary Appendix 1 Figure S3** for calculation details and **S7** for raw values). Negative sera are represented with an ID50 below detection limit (40). Percentage of positive are indicated above each series. **(D)** Correlation plot between LuLISA IgG/N, IgG/S and PNT (red dots) or between PNT and LuLISA IgG/N **(E)** or IgG/S **(F)**. Threshold at confidence index of 98% is shown (dotted lines). Correlation coefficient (*R*^*2*^) and associated *p* values from Pearson test (one-tailed) are indicated above each corresponding area. Numerical values of each combination of assays are summarized with a Venn diagram **(G)** in overlapping areas. Proportion (%) of triple-positive individuals is indicated in red.

**Figure 2:**
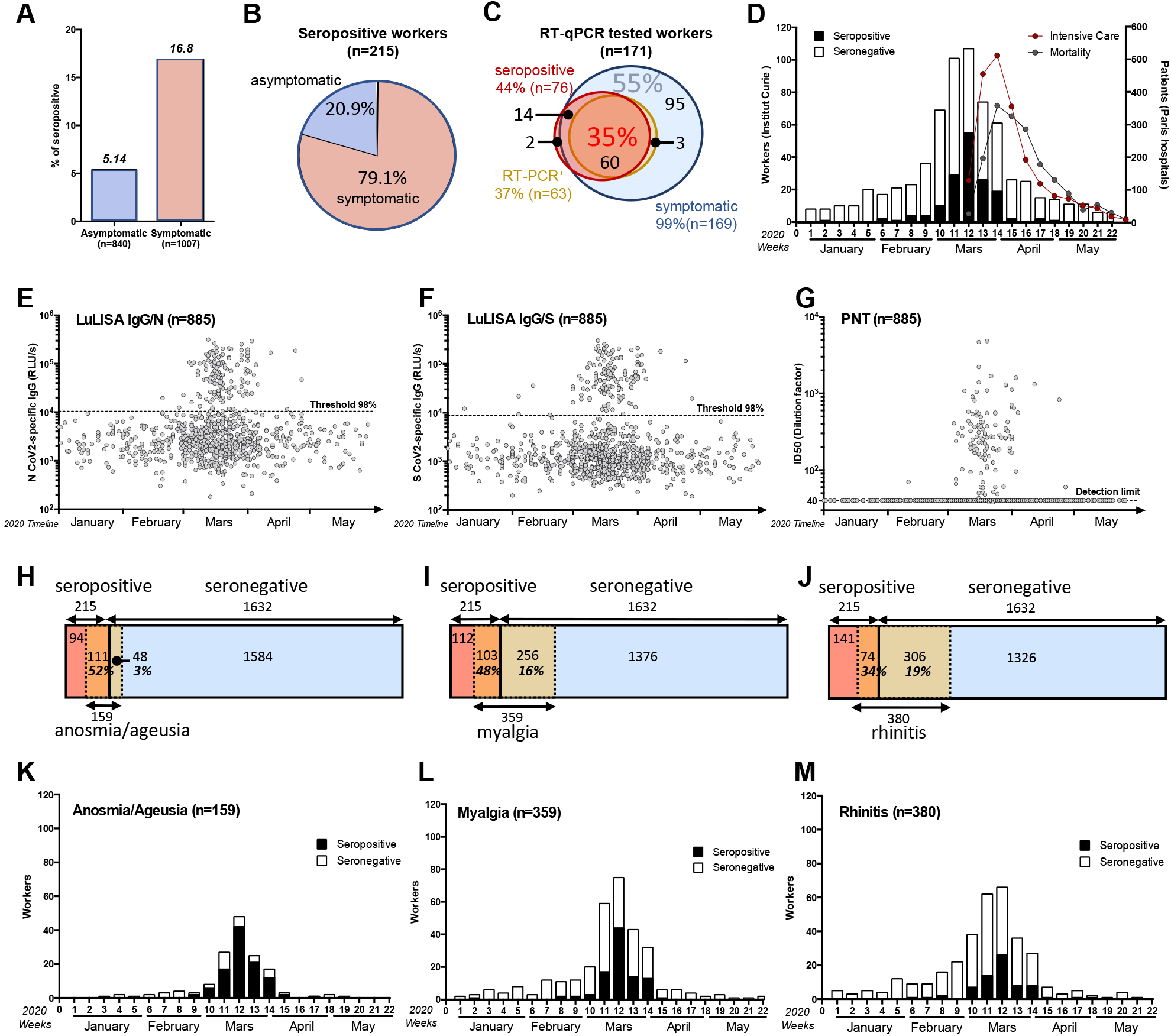
Temporal distribution of symptoms appearance and serology correlates with COVID-19 outbreak in France. **(A)** Seroprevalence among asymptomatic and symptomatic workers. **(B)** Proportion of asymptomatic and symptomatic seropositive workers. **(C)** Correlation between symptom reporting (blue circle), serological profile (red circle) and RT-qPCR result (orange circle) among RT-qPCR tested workers. Proportion (%) of triple-positive individuals is indicated in red and RT-qPCR negative, seronegative among symptomatic workers in blue. **(D-G)** Temporal distribution of serological test results according to first symptom onset. (**D)** Number of workers (left y-axis) reporting at least one symptom but seronegative (white) or seropositive (black). Curves of intensive care admission and mortality in all Paris hospitals are also plotted (right y-axis). (**E-G**): Individual test results according to date of symptom onset. **(E)** LuLISA IgG/N, **(F)** LuLISA IgG/S **(G)** PNT. **(H-J)** Prevalence of symptoms according to serology status. Seropositive workers are represented in red and seronegative in blue. Symptomatic workers are represented in orange/yellow. Number of workers for each area is indicated. Percentage represents the proportion of symptomatic in seropositive (orange area) individuals and symptomatic in seronegative ones (yellow area). **(K-M)** Prevalence of symptom during pandemic outbreak according to serology status. Plots represent the number of workers reporting symptoms (y-axis) per week in 2020 (x-axis). Only the 3 most representative symptoms from **Table 2** are plotted: Anosmia/Ageusia as an example of temporally and clinically correlated to COVID-19 **(H, K)**, Myalgia as clinically only correlated **(I, L)** and rhinitis as poorly correlated **(J, M)**.

**Figure 3:**
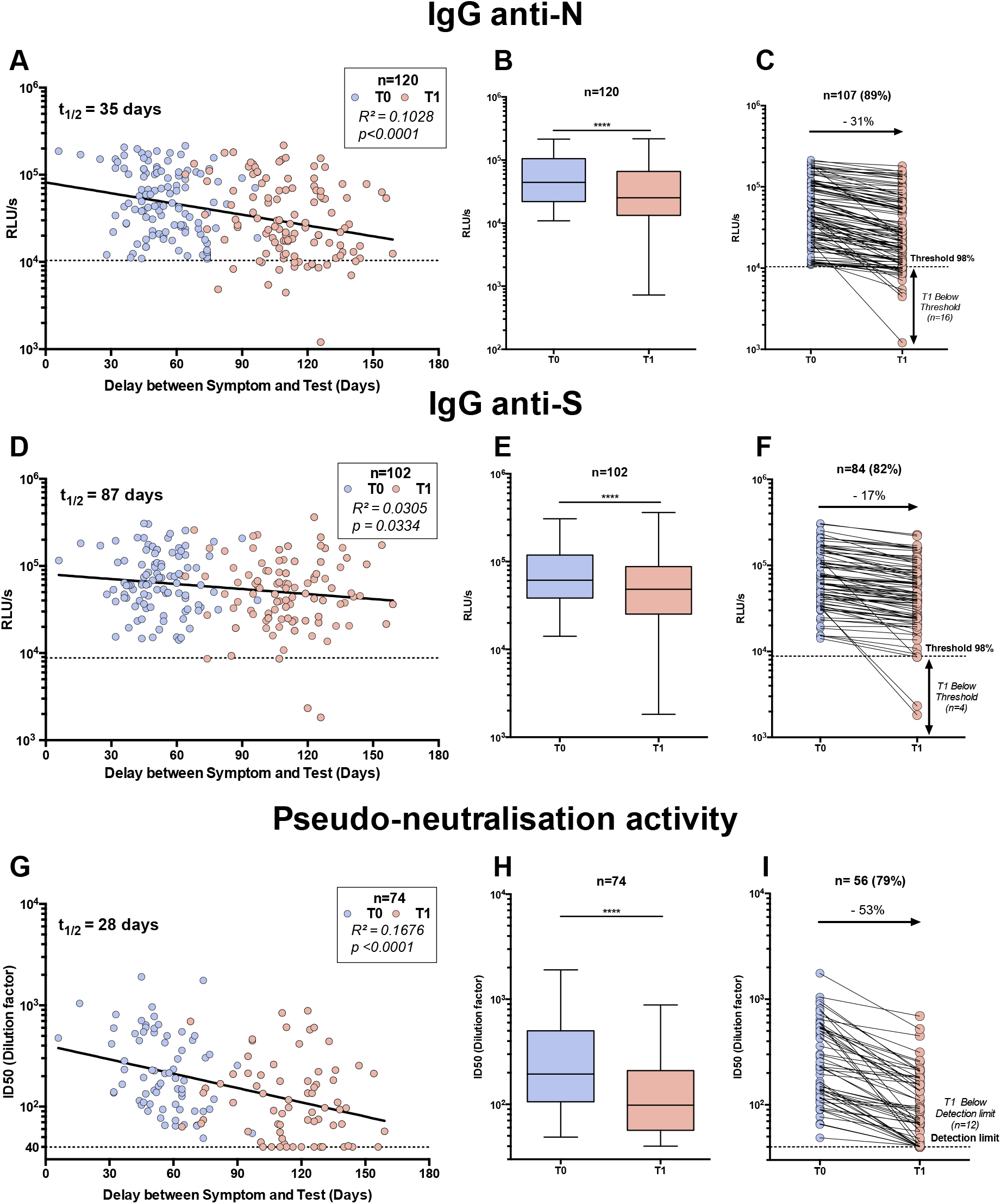
Serological profile follow-up overtime. Workers whose serum was positive for IgG anti-N **(A-B-C)**, anti-S **(D-E-F)** and pseudo-neutralization activity **(G-H-I)** at the first blood sampling (*t*_*0*_) were reassessed together with serum obtained 6-12 weeks later (*t*_*1*_). **A-D-G:** Test values according to delay between date of symptom onset and the two serum analyses: *t*_*0*_ (blue dots) and *t*_*1*_ (red dots). Linear regression is plotted. Coefficient of determination and associated *p* value are indicated. **B-E-H:** Whisker-plots summarizing test value for both tests (*t*_*0*_ and *t*_*1*_). Statistical significance was determined using a Wilcoxon test (****: p<0.0001). **C-F-I:** individual follow-up of seropositive workers with a decreasing value. Variation of mean (*t*_*0*_ */t*_*1*_) is indicated in %.

### STATISTICAL ANALYSIS

The results of the web-based survey were exported into an excel data sheet and analyzed using Prism-graph pad. These results and the raw data of the LuLISA IgG/N and IgG/S and PNT are given in the **Supplementary Appendix 3**.

## RESULTS

### COHORT DESCRIPTION, ASSAY DEVELOPMENT AND VALIDATION

Blood samples were collected from 1847 volunteers at the 3 sites of the Institut Curie: Paris (75), Saint Cloud (92) and Orsay (91) from April 28^th^ until July 31^th^ for the initial time-point. None of the individuals showed clinical signs of COVID-19 or had been subjected to a standard RNA detection of SARS-CoV-2, using RT-qPCR, within 14 days prior to blood sampling. All participants were invited to complete a web-based questionnaire which included demographic variables, symptom occurrences and whether these had led to a sick leave, treatment and/or hospitalization. The participant cohort had a strong (77.4%) female bias (**Table 1**); the mean age was 38 and ranged between 19 and 75 years old. The hospital-working staff represented 72.7% of the volunteers, the rest being researchers and administrative staff.

**Table 1:**
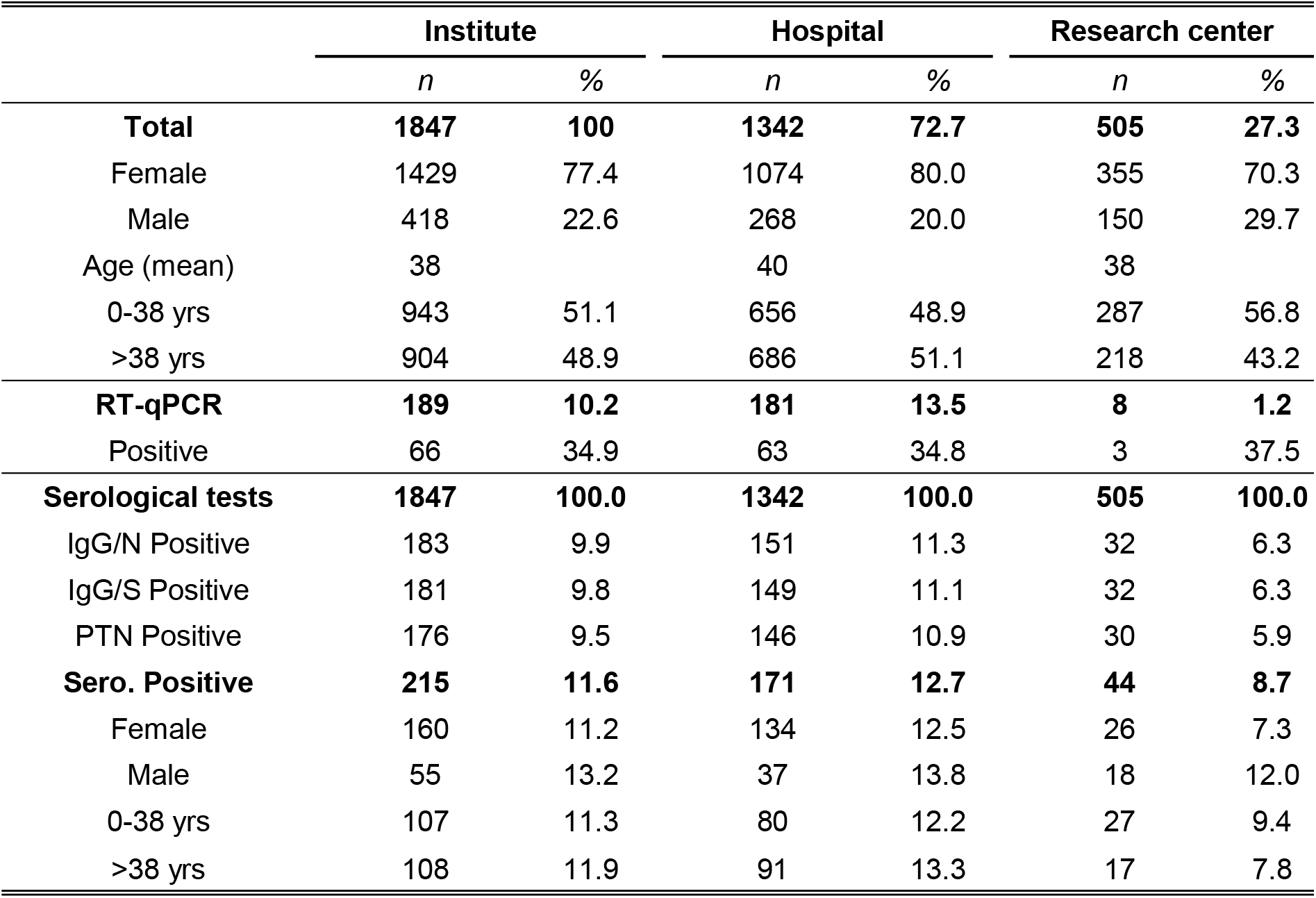
Serological assay results and working groups.

Three serological assays were carried out at the Institut Pasteur in multi-well plates on these 1847 sera samples. Two LuLISA (Luciferase-Linked Immuno-Sorbent Assay) [10], detailed in Supplementary Appendix 1 (**Figures S1** and **S2**), assessed the specific IgG for SARS-CoV-2 Nucleoprotein (N) and Spike (S) proteins. A pseudo-neutralization test (PNT) was also performed [11], to assess the ability of serum components to neutralize the fusion of a SARS-CoV-2 Spike pseudo-typed lentiviral vector encoding a luciferase gene using a permissive human cell line (HEK 293T) constitutively expressing human ACE2 receptors (Supplementary Appendix 1 **Figure S3**). The specificity threshold of the three methods were established by using serum samples from 54 COVID-19 patients (March 2020, Institut Cochin), 234 prepandemic negative healthy donors from a blood bank (2014-2018, EFS/ICAReB) and 75 negative serums from prepandemic breast cancer patients (2012, Institut Curie) (**Figure 1A, B, C**). The positivity thresholds were set to 98% specificity for LuLISA assay allowing the detection of anti-N IgG (10,400 RLU/s) and anti-S IgG (8,400 RLU/s) and to a confidence level of 99% for PNT assay (28,783 RLU/s) from prepandemic negative sera.

The robustness of the specificity thresholds and dynamic ranges were assessed using dilution series of COVID-19 positive sera (**Supplementary Appendix 1 Figures S2 and S3**). The specificity for SARS-CoV-2 anti-N IgG was assessed against purified Nucleoproteins of SARS-CoV-1 as well as seasonal coronaviruses (HCoV) HKU, OC43, NL63, 229E (**Supplementary Appendix 1 Figures S4 and S5**).

### HIGH PREVALENCE OF ANTI-SARS-CoV-2 IgG RESPONSE IN THE STUDY COHORT

For the Institut Curie workers, using a 98% specificity threshold, the seroprevalence of IgG directed against N and S proteins was 9.9% (183/1847, 95% CI: 8.6-11.4) and 9.8% (181/1847, 95% CI: 8.5-11.3), respectively (**Figure 1A-B** and **Table I**). Amongst all the serums tested, 9.5% (176/1847, 95% CI: 8.2-11.0) displayed a pseudo-neutralization activity against the virus (**Figure 1C**). Considering each of these assays independently as a marker of specific immune response leads to a 11.6% (215/1847, 95% CI: 10.2-13.2) positivity of immunization.

The correlative plots (**Figure 1D**) indicates that the responses against the N and S are linked when both are above their respective threshold (*R*^*2*^=0.57). Correlation between PNT and LuLISA is mainly detectable when high levels of both IgG against N and S are detected (red dots in **Figure 1D**). Moreover, above the 98% specificity threshold, a higher correlation is observed between PNT and LuLISA IgG/S (*R*^*2*^=0.60) (**Figure 1F**) than between PNT and LuLISA IgG/N (*R*^*2*^=0.47) (**Figure 1E**). Remarkably, out of the 215 seropositive samples, 72% are positive for the 3 assays (**Figure 1G**), 9.7% are positive only for anti-N IgG (IgG/N), 5.6% for anti-S IgG (IgG/S) and 5.1% for PNT.

### PREVALENCE AND LINETIC OF SYMPTOMS AND SEROLOGICAL RESPONSES

Based on the web-based survey, 54% (1007/1847) participants mentioned at least one symptom (**Supplementary Appendix 3)**. Symptomatic workers were more seropositive (16.8%, 170/1007, CI 95%: 14.6-19.3) than asymptomatic workers (5.3%, 45/840, CI 95%: 3.9-7.1) (**Table 2** and **Figure 2A**). Hence, SARS-CoV-2 infection may have been asymptomatic in at least 20.9% (45/215, 95% CI: 16.5-28.2) of the cases (**Figure 2B**). The amount of anti-N IgG was higher in the symptomatic versus asymptomatic patients while the levels of anti-S or the pseudo-neutralization capacity did not differ (**Supplementary Appendix 1 Figure S6**). This discrepancy suggests that anti-N IgG may be generated in the course of a mild infection.

**Table 2:**
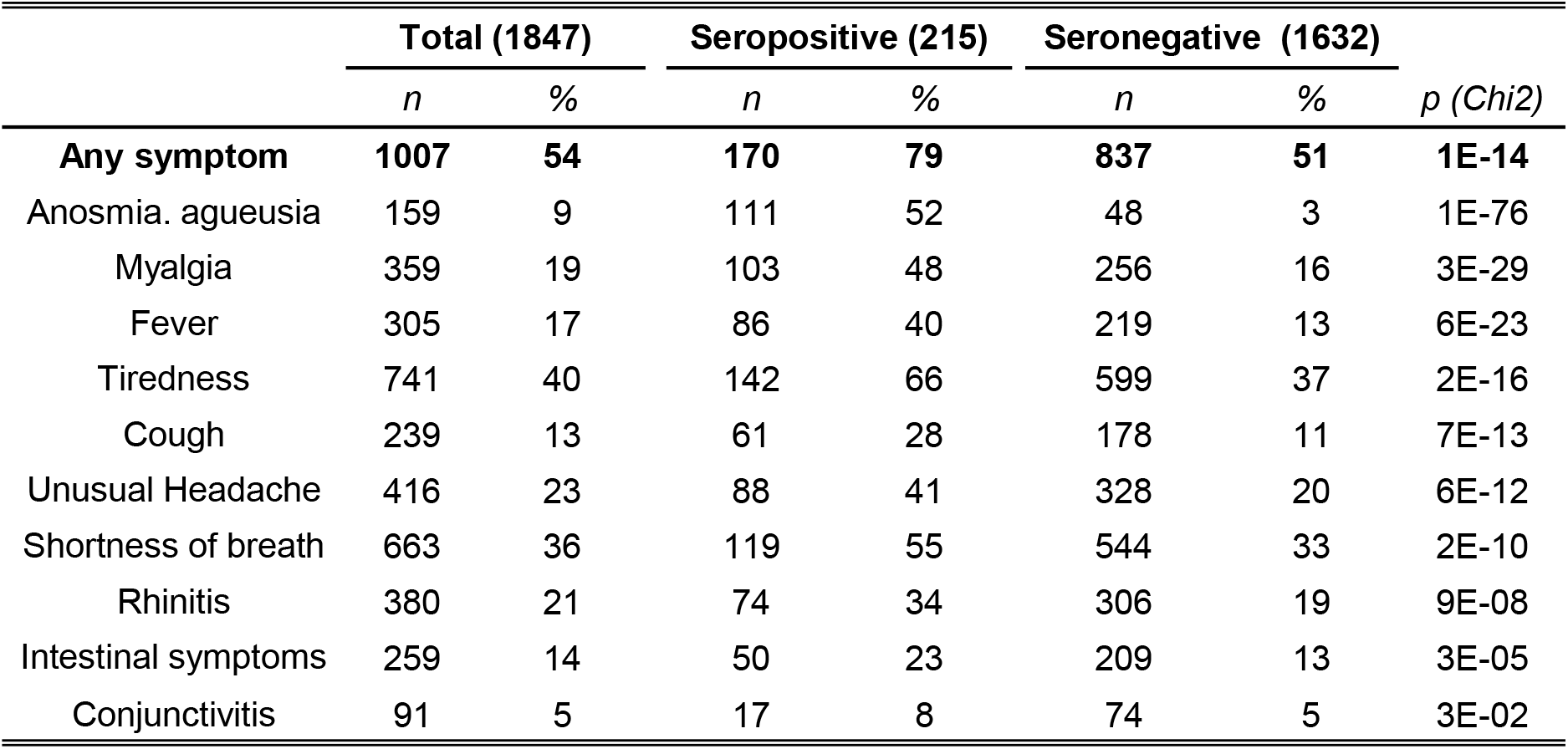
Correlation between serological assays and symptoms.

|  | Total (1847) |  | Seropositive (215) |  | Seronegative (1632) |  | *p* (Chi2) |
| --- | --- | --- | --- | --- | --- | --- | --- |
|  | *n* | % | *n* | % | *n* | % |  |
| **Any symptom** | **1007** | **54** | **170** | **79** | **837** | **51** | **1E-14** |
| Anosmia. agueusia | 159 | 9 | 111 | 52 | 48 | 3 | 1E-76 |
| Myalgia | 359 | 19 | 103 | 48 | 256 | 16 | 3E-29 |
| Fever | 305 | 17 | 86 | 40 | 219 | 13 | 6E-23 |
| Tiredness | 741 | 40 | 142 | 66 | 599 | 37 | 2E-16 |
| Cough | 239 | 13 | 61 | 28 | 178 | 11 | 7E-13 |
| Unusual Headache | 416 | 23 | 88 | 41 | 328 | 20 | 6E-12 |
| Shortness of breath | 663 | 36 | 119 | 55 | 544 | 33 | 2E-10 |
| Rhinitis | 380 | 21 | 74 | 34 | 306 | 19 | 9E-08 |
| Intestinal symptoms | 259 | 14 | 50 | 23 | 209 | 13 | 3E-05 |
| Conjunctivitis | 91 | 5 | 17 | 8 | 74 | 5 | 3E-02 |

A correlation between serological tests, RT-qPCR and symptoms was performed (**Figure 2C**). In the 171 individuals tested by RT-qPCR, 169 (99%) reported symptoms, only 76 (44.4%, CI 95%: 37.9-53.1) were positive in serological assays. Among these 171 RT-qPCR tested workers, 55% were negative, seronegative but symptomatic whereas 35% were positive, seropositive and symptomatic. Moreover, no IgG antibodies were detected in 3 subjects out of 63 with a positive SARS-CoV-2 RT-qPCR indicating that a systemic anti-N or S IgG response may not always be present following a SARS-CoV-2 proven infection. However, low levels of anti-SARS-CoV-2 IgM, were detected using a commercial lateral flow assay, in one of these three subjects (data not shown). Except for one case, all anosmia/ageusia cases without detectable systemic IgG (n=48) were associated with other COVID-19 typical symptoms and occurred in late February, March or April suggesting that they represent true SARS-CoV-2 infections. Indeed, one of them was associated with a positive SARS-CoV-2 RT-qPCR test and, in 7 cases anti-SARS-CoV-2 IgM were detected using a lateral flow assay (data not shown). Thus, in addition to the 215 SARS-CoV-2 immune cases detected by our survey, the cohort may feature an additional 48 infection cases devoid of detectable systemic IgG antibodies. Assuming that the incidence (52%) of the anosmia/ageusia symptom is similar in immune and non-immune individuals the true prevalence of SARS-CoV-2 infection in this population would then be more than 11.6% (215/1847) and as high as 16.6% ((215+(48/0.52)=307); 307/1847).

A date for the symptom onset was mentioned in 885 out of 1007 cases. Symptoms were mostly (61%) reported in March 2020 (**Figure 2D**), consistent with the reported epidemic development as well as the number of Parisian hospital admissions published daily by Santé Publique France [12]. The intensity of immune responses according to the date of symptom occurrence is reported in **Figure 2E-G**. The decrease seen in April (14.8%) probably reflects the efficacy of the population lockdown on the disease spread. The March peak of symptom occurrence represented 82% of the seropositive individuals compared to 56% in people devoid of COVID-19 specific IgG. Although some workers displayed an immune response corresponding to symptoms dated as early as the first week of February 2020, a sharp peak of seropositive individuals corresponded to symptoms declared in March. These results indicate that the virus was circulating in early February in the Paris conurbation and achieved a high prevalence in March.

The frequency of declared symptoms was significantly much higher in seropositive workers (79%) than in those devoid of COVID-19 specific IgG (51%) (**Table 2**). If fever (66%, 142/215) was the most frequent symptoms in the seropositive population, it was also noted in individuals lacking antibodies (37%, 599/1632) suggesting a low correlation with a COVID-19 infection (chi-square scores 2E^-16^) (**Table 2**). To the opposite, anosmia/ageusia and myalgia symptoms were highly prevalent (52%, 111/215 and 48%, 103/215 respectively) in the seropositive group but were rare in the seronegative group 3% (48/1632) and 15.7% (256/1632) respectively (**Figure 2H-J**), resulting in a high correlation with COVID-19 (chi-square scores 5E^-76^ and 3E^-29^) (**Table 2**). Only anosmia/ageusia symptoms were temporally correlated with the epidemic peak in March whereas other symptoms such as myalgia and rhinitis (**Figure 2K-M**) were declared by seronegative workers mainly before but also after this peak suggesting the effects of other circulating diseases.

### DECREASE OF ANTIBODY TITER AND NEUTRALIZATION ACTIVITY WITH TIME

To follow over time the antibody titers and neutralizing activity, a second blood sample (t_1_) was obtained 4-8 weeks after the first one (*t*_*0*_) for more than 1000 individuals. The results, for the 120 samples of individuals previously found positive, are reported in **Figure 3A, D, G** according to the time interval between symptom onset and sampling. A clear decrease in the antibody titers and virus pseudo-neutralization capacity was observed. The half-lives of the antibody titers were 35, 87 and 28 days for anti-N, anti-S IgG and pseudo-neutralization, respectively. A paired analysis showed a systematic decreased response (p<0.0005) (**Figure 3B, E, H**). The titers of antibodies decreased by 31% and 17% for anti-N and anti-S IgG, respectively for a majority of workers (>75%) and this correlated with a major decrease of the ID50 pseudo-neutralization capacity (53%) (**Figure 3C, F, I**). Interestingly, some workers sera became negative in our assays: 15% (16/107) for LuLISA IgG/N (**Figure 3C**), 14% (10/71) for PNT (**Figure 3I**) and 5% (4/84) for LuLISA IgG/S (**Figure 3F**). Thus, after a few months, a serological-based survey of SARS-CoV-2 may run a risk of underestimating the number of formerly infected individuals.

## DISCUSSION

We report here the longitudinal study Curie-O-SA describing the natural immune response against the SARS-CoV-2 in a large population of healthy subjects working in the Paris conurbation following the March 2020 peak. Three bioluminescence-based and sensitive high-throughput assays including a viral pseudo-neutralization allowed repeated measurements on a large number of samples. In contrast with other studies focusing on hospitalized patients or based on the occurrence of symptoms [13-18], this study included more than half of all the employees working at three distant locations of a non-especially COVID-exposed institution with a corresponding web-based questionnaire filled up by more than 96% of the participants.

This survey evidenced a high prevalence (11.6-16.6%) of previous infection by the SARS-CoV-2 in March/April 2020 along with the following points: 1) the levels of IgG/N and IgG/S were highly correlated beyond twice their positive thresholds as wells as with the viral pseudo-neutralization capacity beyond three times their positive thresholds. 2) 21% of infections had been asymptomatic. 3) At least 5% of the RT-qPCR confirmed infections and 30% (92 of 307) of the very probable infection cases according to symptoms did not develop any detectable anti-N or anti-S IgG antibodies nor a serum neutralization capacity. 4) The systemic IgG/N and IgG/S immunity associated with pseudo-neutralization activity decreased rapidly with a half-life between 4 to 12 weeks following infection.

There are some limitations of our study. 1) High throughput methods for assessing IgM or IgA responses were not ready at the time of the study. Assessing these isotypes may be relevant since a commercial lateral flow assay detected anti-SARS-CoV-2 IgM in 7 out of 48 individuals featuring anosmia/ageusia but devoid of IgG response. 2) For regulatory and logistic reasons, the blood sampling started 4-6 weeks after the epidemic peak and were performed over a 2.5-month period. The interval between the infection and the blood draw varied between 2 to 18 weeks. We may thus have missed the antibody response peak in some cases and, again, underestimated the true prevalence of the infection. 3) For logistic reasons, the blood samplings from research center staff were delayed by an average 2 weeks, possibly leading to a slight underestimation of the true prevalence of SARS-CoV-2 infection in the research center. 4) The use of a web-based auto-questionnaire leaves some space for inaccurate or selective memories as well as input errors and missing values. Indeed, despite the high response rate, the symptom onset date was missing in 18% of the cases (187/1007). Symptoms were declared by 79% of the immunized individuals but also by 51% of the individuals without antibodies, suggesting either a still higher proportion of infections without detectable systemic antibodies or the limit of self-reported symptom questionnaire. 5) Although more than half of the employees participated to the study, a selection bias is still possible with recruitment of those with the most symptoms leading to an overestimation of SARS-CoV-2 infection prevalence.

In accordance with recent studies 38/1847 individuals were RT-qPCR positive but negative for serological tests [19] and all subject among the 215/1847 did not display a common scheme of coordinated immune response [20] which included all the parameters studied: the most prevalent response was the anti-N IgG, followed by the anti-S IgG and then the pseudo-neutralizing activity. A few individuals were endowed with neutralizing sera without detectable IgG against S, either because their S-specific IgG had high affinity but low concentration or because other Ig than IgG were responsible for the pseudo-neutralization as recently suggested for S-specific IgA or IgM [21]. The lower prevalence of the PNT activity could be either related to a lower sensitivity of the assay or to a decrease of the immune response as evidenced in **Figure 3**. Indeed, although most of the symptoms occurred from March to early April, the blood samplings were performed between May and July, leading to a variable interval between an eventual infection and the antibody response study.

Our experiments pointed out a clear cross-recognition of IgG for SARS-CoV-1 and -2 Nucleoproteins but none with any of the seasonal HCoV, aside from samples displaying a very high IgG/N content (**Supplementary Appendix 1 Figure S4**). This may be expected in view of the relatively few short peptide patterns common to all the antigens sequences, as shown in their alignment (**Supplementary Appendix 1 Figure S5**). Altogether, as no documented cases of SARS-CoV-1 were observed in France in 2003 during the Asian outbreak and afterward, we do not expect any bias of the seroprevalence for SARS-CoV-2 with SARS-CoV-1 or the seasonal HCoV in Paris from February to June.

Our results are consistent with other large-scale serological studies in conurbations that have been subjected to the SARS-CoV-2 epidemic, mostly with single time-point without symptom reports. Such a high seroprevalence was found in New-York City (USA, n=5129, 22.7%, March) [22] but not in Geneva (Swiss, n= 2766, 8.5%, April) [23]; Wuhan (China, n=17368, 3.8%, March-April) [24] or 9 cities of Rio Grande do Sul (Brazil, n=4500, 0.22%, May) [25]. Seroprevalence in healthcare workers highly exposed to COVID is also contrasted. It was high in London (n=2167, 31.6%, Mai-June 2020) [14] but lower in Essen (Germany, n=316, 1.6%, March) [15]; Madrid (Spain, n=578, 9.3%, April) [13]; Milano (Italia, n=789, 10.8%, March) [17].

Among seropositive individuals, 20% had been asymptomatic in this study which is less than what is mentioned in other studies depending on recording and reporting methods: 40% in Madrid area [13], 50% in Boston area [26] and up to 80% locally in France on September 2020, where it is a likely consequence of reinforced mask wearing policies [27] whom efficacy was observed in Wuhan with 86% of asymptomatic in January 2020 [28], maybe in relation with decreased infectious load [27].

The pattern of symptoms displayed by the immune subjects are consistent with those reported elsewhere [29, 30]. Our results further emphasize the predictive value and specificity of the anosmia/ageusia symptoms. Only 51% of the studied individuals, 77% of the immunized individuals declared symptoms aside from fatigue. The higher prevalence of infection in the hospital workers (12.7%) than in the research center (8.7%) is certainly not related to difference in confinement since most contaminations occurred in March 2020 before or at the time of the lockdown. Because the hospital treated very few COVID patients, it is likely that the observed contaminations in the hospital staff are resulting from public transportation use as well as social encounters rather than work-related. The difference of the RT-qPCR number performed between the hospital and research center is mainly due to the health staff priority access to SARS-CoV-2 RT-qPCR assays in March and April 2020.

In this first report, we studied the immune response of 120 positive individuals 6-12 weeks after the first blood sampling. A majority of these individuals (>75%) displayed a diminishing anti-SARS Cov2 response. Notably, the anti-N IgG decreased faster (31%) than anti-S IgG (17%) suggesting that if anti-N IgG titration is a reliable marker for prevalence follow-up during the early stage of the COVID-19 pandemic, this assay may be less relevant for longer studies and should be seconded with an anti-S IgG assay. This observation emphasizes the difficulty to estimate the real seroprevalence in a large population. Interestingly, the slow drop of anti-S IgG titer did not correlate with the major decrease of pseudo-neutralization activity observed (53%). Since our pseudo-neutralization assay is exclusively associated with anti-S response, the neutralization activity we observed might be explained by the occurrence of other Ig isotypes, such as IgM or IgA, which would disappear much faster than the IgG from subsequent blood samples [21]. This illustrates again the serological complexity of any long-lasting immunity.

From an epidemiological perspective, the 11.6-16.6% seroprevalence results may still underestimate the number of individuals who have been infected by the SARS-CoV-2 because, as discussed earlier, we also observed a lack of systemic IgG response among the RT-qPCR positive individuals along with a gradual loss of the virus-specific IgG titer. In the present epidemic, the rather fast decrease in antibody titers is also hindering any retrospective assessment of its true extent. Since Paris may be considered as representative of the world hard-hit conurbations, such high prevalence of a SARS-CoV-2 previous infection along with a short-lived immune response are raising the issues of possible reinfection and virus persistence in a high-density population and may be important parameters to guide future public health policies.

## Data Availability

Detailed data are provided as spupplementary information

## ACKNOWLEDGEMENTS

Authors thank all the volunteers from Institut Curie for their participation to this study. The help of the personnel in the Departments of diagnostic and theranostic medicine, of early phase clinical trials and of the outpatient clinics is gratefully acknowledged. Authors thank Sylvie Arnaud, Anne Blondel, Anne-Claire Coyne, Aurélie Dos Santos, and Cécile Simondi for their contribution in blood sampling and study logistics at Institut Curie, Pr Frédéric Pène from Institut Cochin for access to COVID-19 patient samples and Dr Yves Jacob from Institut Pasteur for the four HCoV genes. The authors thank Sébastien Brulé, Sylviane Hoos, Dr Bertrand Raynal and Dr Patrick England from the Molecular Biophysics Platform for the quality control of protein targets at the Institut Pasteur. The authors thank Dr Hélène Munier-Lehmann for access to automate and supply management at the Unit of Chemistry and Biocatalysis, Institut Pasteur. The authors thank Pr J. Di Santo and Dr D. Duffy for their comments. This work was made possible thanks to the financial support obtained through the «URGENCE nouveau coronavirus» fundraising campaign of Institut Pasteur and the financial support of the Fondation Total. This study was funded in part by a grant from Fondation de France and by Institut Curie Institutional funding. The luciferin synthesis development has been supported by DARRI (ValoExpress 2016-2018). LuLISA development has been supported by IARP Pasteur-Carnot MI (2019-2020). We thank the technical support of Berthold France and Berthold Technologies Germany.

## AUTHORS

PNT design: FA, PC; PNT processing: FA, PS; LuLISA design: SG; LuLISA processing: SG, TR; Target design: MG, ZC, NE; Target production: FD, ED, OR-LG, SP; Substrate synthesis: YJ; Study logistics and sampling collection: DL, VG, FC-B, AS, MDG, OL; Automate and plate handling: OH; Data analysis: FA, TR, AIL, OL; Contribution to text and figure editing: FA, SG, AIL, YJ, TR; writing manuscript: FA, TR, OL.

## CONFLICT OF INTEREST

YJ, SG and TR have patented the proluciferins (hikarazines) synthesis and uses (EP 3395803 / WO 2018197727, 2018) and applied for a patent which includes claims describing the LuLISA. FA and PC have applied for a patent claiming the PNT.

## SUPPLEMENTARY INFORMATION

**APPENDIX 1 (Supplementary Methods, Data and Figures)**

**APPENDIX 2 (Questionnaire)**

**APPENDIX 3 (Excel file, symptoms, serology)**

## Notes

### Clinical Trial

This study was registered and received ethical approval by the Comite de Protection des Personnes Mediterranee III (2020.04.18 bis 20.04.16.49458, 27/4/2020) registered in the clinical trial database (NCT04369066)

### Author Declarations

This study was registered and received ethical approval by the Comite de Protection des Personnes Mediterranee III (2020.04.18 bis 20.04.16.49458, 27/4/2020) registered in the clinical trial database (NCT04369066). Following informed consent, 18 years of age or older volunteer participant outside of any SARS-CoV-2 acute infectious episode in the last 7 days, working at one of the three Institut Curie locations (Paris, Orsay or Saint Cloud) completed a web-based questionnaire (Ennov Clinical) detailed in Supplementary Appendix 1 and 2.

### Summary of Updates

Figure 1 revised Legend Figure 1 revised Authors and affiliation revised Supplementary Figure S6 revised Supplementary data Appendix 3 modified as requested: 1. Precise ages are replaced with an age range:19-38 or 39-82 in accordance with the median used in the text 2. Exact dates have been removed.

