## Supplementary information: Appendix 1, 2 and 3 for "High seroprevalence but short-lived immune response to SARS-CoV-2 infection in Paris"

**SUPPORTING INFORMATION : APPENDIX 1**
This appendix has been provided by the authors to give readers additional information about their work.
Supplement to: Anna, Goyard, et al.

**High seroprevalence but short-lived immune response to SARS-CoV-2 infection in Paris**

François Anna[1]*, Sophie Goyard[2,3]*, Ana Ines Lalanne[4,5]*, Fabien Nevo[1,6], Marion Gransagne[7], Philippe Souque[6], Delphine Louis[4,5], Véronique Gillon[8], Isabelle Turbiez[8], François-Clement Bidard[5,9,10], Aline Gobillion[11], Alexia Savignoni[11], Maude Guillot-Delost[5,12], François Dejardin[13], Evelyne Dufour[13], Stéphane Petres[13], Odile Richard-Le Goff[14], Zaineb Choucha[7], Olivier Helynck[15], Yves L. Janin[15], Nicolas Escriou[7], Pierre Charneau[1,6], Franck Perez[16], Thierry Rose[2,3]**, Olivier Lantz[4,5,12]**

**Author affiliations**

1. Theravectys, Paris, 75015, France
2. Unit of Lymphocyte Cell Biology, Immunology Department, Institut Pasteur, Paris, 75015, France
3. INSERM 1221, Institut Pasteur, Paris, 75015, France
4. Laboratoire d'Immunologie Clinique, Institut Curie, Paris, 75005, France.
5. Centre d'Investigation Clinique en Biothérapie, Institut Curie (CIC-BT1428), Paris, 75005, France
6. Unit of Molecular Virology and Vaccinology, Virology Department, Institut Pasteur, Paris, 75015, France
7. Innovation Laboratory: Vaccines, Institut Pasteur, Paris, 75015, France
8. Direction of the Clinical Research, Institut Curie, Paris, 75005, France.
9. Medical Oncology Department, Institut Curie, Paris, 75005, France
10. UVSQ, Paris-Saclay University, Saint-Cloud, 92210, France
11. Biometry, Institut Curie, Paris, 75005, France
12. INSERM U932, PSL University, Institut Curie, Paris, 75005, France
13. Production and purification of recombinant proteins technological platform, Paris, 75015, France
14. Unit of Antibody in Therapy and Pathology, Institut Pasteur, Paris, 75015, France
15. Unit of Chemistry and Biocatalysis, Institut Pasteur, UMR 3523 CNRS, Paris, 75015, France
16. Unit of Cell Biology and Cancer, Institut Curie, Paris, 75005, France

**Table of Contents**

**Supplementary Methods**

- Ethical considerations
- Participant enrolment
- Questionnaire
- Luciferase-Linked Immuno Sorbent Assays (LuLISA)
    - Plasmid design and production of Nucleoprotein and Spike from SARS-CoV-2
    - Plasmid design and production of Nucleoprotein from SARS-CoV-1 and HCoV
    - Design and synthesis of plasmid encoding the anti-IgG nanobody-luciferase tandem
    - Expression, purification and validation of anti-IgG nanobody-Luciferase tandem
    - IgG LuLISA assay protocol
- S-Pseudo-typed Neutralization Assay (PNT)
    - Pseudo-virus production and permissive cell line generation
    - Pseudo-neutralization assay protocol
- Statistical analysis

**Supplementary Figures**

- Figure S1. Description of anti-IgG VHH Fc1-KAZ and of SARS-CoV2 viral protein targets Nucleoprotein and Spike
- Figure S2. Luciferase-linked immuno-sorbent assay
- Figure S3. Pseudo-neutralization assay (PNT)
- Figure S4. Specificity of assayed IgG against Nucleoproteins of SARS-CoV2 vs SARS-CoV1 and seasonal virus HCoV
- Figure S5. Sequence comparison of SARS-CoV2, SARS-CoV1 and seasonal virus Nucleoproteins
- Figure S6. Serological profile comparison between asymptomatic and symptomatic workers
- Figure S7: PNT raw data for cohort analysis

**References**

#### SUPPLEMENTARY METHODS

##### Ethical considerations

This study was registered and received ethical approval by the Comité de Protection des Personnes Méditerranée III (2020.04.18 bis_ 20.04.16.49458, 27/4/2020) registered in the clinical trial database (NCT04369066). All participants were informed of the study outcomes. All blood samples have been drawn and anonymized at the Institut Curie and their analysis have been performed at the Institut Pasteur, Paris.

##### Participant enrolment

The study called volunteer participants working at one of the three Institut Curie locations (Paris, Orsay or Saint Cloud) for four longitudinal blood tests at $t_0$ then after 1, 3 and 6 months. Following informed consent, 18 years of age or older participant outside of any SARS-CoV-2 acute infectious episode in the last 7 days, completed a web-based questionnaire (Ennov Clinical). Participants covered hospital and non-hospital related activities, age and sex. A 5 mL blood sample was taken from all participants in dry tubes. After clotting, blood was centrifuged 10 min at 2000 g and frozen. Supernatant serum was separated and frozen.

##### Questionnaire

The questionnaire provided in Supplementary Appendix 2 recorded individual information and details related to SARS-CoV-2 infection, including date of testing, date of symptom onset and a description of symptoms if there were some. Recorded symptoms were fever, fatigue, cough, difficulty breathing, shortness of breath, loss of taste (ageusia) or smell (anosmia), headache, muscle aches, conjunctivitis or colds, digestive disorders (vomiting, diarrhea).

##### Luciferase-Linked Immuno Sorbent Assays (LuLISA)

###### *Plasmid design and production of Nucleoprotein and Spike from SARS-CoV-2*

Production of the Nucleoprotein (N): A codon-optimized gene was synthesized (GeneArt, Thermo Fisher Scientific, Waltham, MA) and subcloned in the pETM11 expression vector[1] allowing the expression of the full-length N protein (UniProtKB ID: P0DTC9) with an amino-end $(His)_6$ tag in the *E. coli* BL21 (DE3) pDIA17 strain. Bacteria were grown in Nzytech auto-inducible-medium (Lisboa, Portugal) for 4 hours at 37°C then 15 hours at 18°C. Cultures are then centrifuged and the pellets were lyzed using a cell disrupter (Constant system Ltd, Daventry Northants, UK) in a phosphate saline lysis buffer (phosphate 50 mM, NaCl 300 mM, pH=8) supplemented with 20 mM imidazole, one complete EDTA-free protease tablet (Merck, Darmstadt, Germany) and 1250 units of benzonase (Merck, Darmstadt, Germany). 2.5mg of RNAse was added (Thermo Fisher Scientific, Waltham, MA) and the lysate was further incubated at room temperature for about 20 min. The lysate was centrifuged at 19000 rpm and the soluble fraction was recovered for affinity purification on Protino Ni-NTA 5 ml column (Macherey Nagel, Dueren, Deutschland). The protein was eluted by a 20 - 250 mM imidazole gradient in lysis buffer. The fractions containing the protein of interest were pooled and injected in a Superdex 200 16/600 size-exclusion chromatography column (Cytiva, Velizy, France) equilibrated in a phosphate 50 mM, NaCl 500 mM pH=8 buffer. The protein was eluted as an homodimer and analyzed in denaturing condition on a stain-free SDS PAGE (4–15% Mini-PROTEAN® TGX Stain-Free™ Protein Gels, Bio-Rad) as shown in Supplementary Appendix 1 Figure S1 and native condition using multi-angle diffusion light scattering (not shown, Wyatt, CA, USA) for quality validation (Molecular Biophysics Facility, Institut Pasteur). Samples were aliquoted (0.35 mg/mL) and stored at 4°C until use.

Production of the Spike protein (S): A codon-optimized nucleotide fragment encoding the ectodomain (amino acid 1 to 1211) of the S protein (UniProtKB ID: P0DTC2) was synthesized (GeneArt, Thermo Fisher), fused to nucleotide fragments encoding the trimerization foldon domain of the bacteriophage T4 fibritin and a Twin-Strep-tag (IBA Life Science, Goettingen, Germany) and subcloned into the pCI vector (Promega, Madison,WI USA). The ectodomain was further stabilized in its uncleaved prefusion state through inactivation of its furine cleavage site (R682G, R683S and R685G) and insertion of the 2 proline mutations (K986P and V987P) described by Kirchdoerfer et al. for SARS-CoV-1 spike protein [2], respectively. The T4 fibritin foldon ($g^{1}$yipeaprdgqA$^{12}$YVRKDGEWVLL$^{23}$stfl) [3] is used here as an artificial trimerization domain (β-hairpin propeller) for mimicking the Spike trimeric assembly at the virion surface. Expi 293F cells (Life Technologies, Courtaboeuf France) were grown in freestyle 293 expression medium and transfected with the expression vector (300 µg DNA/ 30 mL of freestyle 293 medium) and 300 µl of PEI pro (Polyplus transfection, Illkirch, France). Cells were grown 5 days at 37°C and 8% $CO_2$, orbitaly agitated at 110 rpm. The cultures were centrifuged and the supernatant was equilibrated at pH=8.0 with TRIS 1M then concentrated using a 100 kDa MWCO tangential flow filtration cartridge on an Äkta Flux (Cytiva, Velizy France) with a phosphate filtration buffer (phosphate 50 mM, NaCl 300 mM pH=8.0). The retentate was recovered for Twin-Strep-tag-affinity purification on StrepTrap column (Cytiva, Velizy France). The protein was eluted using 2.5 mM desthiobiotine in filtration buffer. The protein was recovered as an homotrimer and analyzed in denaturing condition on a stain-free SDS PAGE (4–15% Mini-PROTEAN® TGX Stain-Free™ Protein Gels, Bio-Rad) as shown in Supplementary Appendix 1 Figure S1 and native condition using analytical size exclusion chromatography and multi-angle diffusion light scattering for quality validation (not shown, Molecular Biophysics Platform, Institut Pasteur). Samples were aliquoted (0.52 mg/mL) and stored at 4°C until use.

Production of Nucleoproteins from SARS-CoV-1 and HCoV: The optimized synthetic gene (GeneArt, Thermo Fisher) of SARS-CoV1, HCoV-HKU, HCoV-OC43, HCoV-NL63, HCoV-229E (UniProtKB ID: P59595, Q5MQC6, P33469, Q6Q1R8, P15130 respectively) with carboxy-end $His_6$-tag and flanking region corresponding to the pET23 sequence (Novagen). pET23 plasmids were amplified with the forward and reverse oligonucleotides using a Q5 DNA polymerase, dNTP mix (New England BioLabs). The PCR products were purified by electrophoresis on agarose gel (1%, Macherey Nagel). The purified pET23 vector and the synthetic genes were assembled using NEBuilder HiFi assembly master mix (New England BioLabs). These plasmid were used to transform *E.coli* BL21 (DE3). Cells were grown at 18°C and induced with IPTG (Sigma-Aldrich). After harvesting the cells by centrifugation (1.5 L), the pellet was resuspended in 50 mM Tris-HCl pH 8.0, 50 mM NaCl with protease inhibitor (Sigma-Aldrich) and lysozyme (0.1 mg/mL, QBIO-gene). Cells were disrupted by freezing-thawing cycle lysis method. DNase (Sigma-Aldrich) was then added to remove DNA from the sample. SARS-CoV-1 and HCoV Nucleoproteins were purified as described for Fc1-nanoKAZ below. The protein quality was analyzed in denaturing condition on a stain-free SDS PAGE (4–15% Mini-PROTEAN® TGX Stain-Free™ Protein Gels, Bio-Rad) as shown in Supplementary Appendix 1 Figure S4 and in native condition by acquisition of UV-spectra (240-300 nm). Concentrations were estimated from the solution absorption at 280 nm and sequence by the method of Gill and von Hippel [4]).

***Design and synthesis of plasmid encoding the anti-IgG nanobody-luciferase tandem***

NanoKAZ (19 kDa) is an optimized sequence of the catalytic domain of the luciferase from the subtropical sea shrimp *Oplophorus gracilirostris* [5, 6]. The gene *kaz* has been optimized and synthetized by Eurofins (Germany) with carboxy-end $His_6$-tag and flanking region corresponding to the pET23 sequence (Novagen). pET23 plasmids were amplified with the forward and reverse oligonucleotides using a Q5 DNA polymerase, dNTP mix (New England BioLabs). The PCR products were purified by electrophoresis on agarose gel (1%, Macherey Nagel). The purified pET23 vector and the synthetic gene were assembled (pET23-*kaz*) using NEBuilder HiFi assembly master mix (New England BioLabs).

The Fc IgG–binding moiety (Kd=2.14 nM) is from an antibody of the camelid species alpaca (*Vicugna pacos*) selected against the Fc domain of human IgG (single-domain antibody, [7]). The gene *fc1* has been synthetized by Eurofins with a flanking region corresponding to the pET23-*kaz* sequence. Synthetic gene *fc1* was amplified with the corresponding forward and reverse oligonucleotides using a Q5 DNA polymerase and dNTP mix. PCR products were purified by electrophoresis on agarose gel. Purified pET23-*kaz* vector and the synthetic gene *fc1* were assembled using the NEBuilder HiFi assembly master mix. The assembled products (5 μL) were used to transform NEB 5-alpha competent *E. coli* and grown overnight on LB/Agar/ampicillin in a Petri dish. Isolated colonies were grown in liquid medium, plasmids were isolated. The full sequence of the anti-IgG nanobody luciferase tandem ([Fc1]-[nanoKAZ]-[$His_6$]) was analyzed to confirm the presence of the *fc1-kaz* insert shown in Supplementary Appendix 1 Figure S1.

***Expression, purification and validation of anti-IgG nanobody-Luciferase tandem***

The pET23-*fc1-nanokaz* was used to transform *E.coli* BL21 (DE3) to achieve high expression in *E.coli.* Cells were grown at 18°C and IPTG (Sigma-Aldrich) was added to induce *fc1-nanoKAZ* production. After harvesting the cells by centrifugation (1.5 L), the pellet was resuspended in 50 mM Tris-HCl pH 8.0, 50 mM NaCl with protease inhibitor (Sigma-Aldrich) and lysozyme (0.1 mg/mL). Cells were disrupted by freezing-thawing cycle lysis method. DNase (Sigma-Aldrich) was then added to remove DNA from the sample.

The crude extract was centrifuged 30 min at 15,000 g. The supernatant was collected and NaCl (500 mM), Imidazole (20 mM, Sigma-Aldrich) and Triton (0.1%, Sigma-Aldrich) were added. The cleared lysate was loaded on a Hi-Trap 5 mL-column (Cytiva, Velizy France) at 4 mL/min using an Äkta pure chromatography system (Cytiva, Velizy France). The column was washed with 20 volumes of column with a running buffer (50 mM Tris-HCl pH 8.0, NaCl 50 mM, 20 mM imidazole) at 5 mL/min. The VHH Fc1-nanoKAZ was eluted with a gradient of imidazole from 20 mM to 200 mM in 50 mM Tris-HCl pH 8.0, 50 mM NaCl at 5 mL/min and fractions of 1 mL were collected in a 96-deepwell plate (Cytiva, Velizy France) . Catalytic activity of fractions was profiled using a luminometer Hydex by diluting $10^7$ fold the fractions in PBS with 27 μM of the imidazo[1,2-*a*]pyrazin-3(7*H*)-one derivative Q-108 resulting from the hydrolysis of hikarazin-108 [8]. The fractions of high activity were pooled, and loaded on a 1 mL HiTrap Q column (Cytiva, Velizy France) equilibrated in 50 mM Tris-HCl pH 8.0, NaCl 50 mM. The protein was eluted in 50 mM MES pH 6.5, 50 mM NaCl at 1 mL/min at 18°C using the Äkta pure chromatography system. The fractions of 500 μL were collected in 96-deepwell plates and their activities were assayed as described above. The fractions with high activity were pooled. The quality of the purified protein was assessed by loading an aliquot (10 μL) on a stain-free SDS gel (4–15% Mini-PROTEAN® TGX Stain-Free™ Protein Gels, Bio-Rad). The gel was activated by UV trans-illumination for 5 min (Bio-Gel Doc XR Imaging System, Bio-Rad). Tryptophan residues undergo an UV-induced reaction with trihalo compounds and produce an imaged fluorescence signal (Supplementary Appendix 1 Figure S1). An UV-spectrum (240-300 nm) was acquired for evaluating the concentration of Fc1-nanoKAZ (MW 34,726 Da) from the solution absorption at 280 nm ($\varepsilon$ = 50,780 $M^{-1}cm^{-1}$ estimated by the method of Gill and von Hippel [4] ). The specific activity is about $4 \cdot 10^{17} \pm 0.5 \cdot 10^{17}$ $RLU \cdot s^{-1} \cdot g^{-1}$ of Fc1-nanoKAZ.

***IgG LuLISA protocol***

The LuLISA (Luciferase-Linked Immuno-Sorbent Assay) is an ELISA-based method (Enzyme-Linked Immuno-Sorbent Assay) for which the enzyme coupled to the antibody fragment is a luciferase [9] and is distinguished by the nature of the measurement, the relative number of photons collected per second, which quantifies an instantaneous rate of catalytic reaction, whereas ELISAs measure an accumulation of a product or the disappearance of a substrate, whether absorbent or fluorescent, and whose reaction can be stopped at any time. If the relative number of photons in the first minutes is proportional to the number of luciferases in solution, the measurement must be carefully calibrated with robust references under the same conditions in the same multi-well plates. Indeed, luciferases catalyze the decarboxylation of a luciferin, here Q108 (made by the deacetylation of from hikarazine-108) [10], into an amide in the presence of molecular oxygen with a reaction speed dependant on temperature, amount of dissolved oxygen and substrate concentration. The luciferase used is a nanoKAZ [6] which exhibits inhibition by excess substrate in excess of 54 μM and stochastic inactivation by one of the transient reaction products. The enzyme at the optimum substrate concentration has a half-life of 60 minutes with Q108 [8, 10]. The antibody fragment is derived from an alpaca heavy single chain (VHH) with an affinity of 0.58, 1.73, 47.8, 0.30 nM (KD) for the Fc domains of human IgG1, IgG2, IgG3 and IgG4 respectively with an average for overall IgG of 2.14 nM [7] . The anti-IgG VHH-nanoKAZ construction is described in Supplementary Appendix 1 Figure S1A.

As shown in the Supplementary Appendix 1 Figure S2, correlation for four replicates of 96 LuLISA IgG/N on a single 384-well plate with fresh sera is greater than 0.97 ($R^2$), roughly pipetting errors aggravated by viscosity and protein aggregation variations. $R^2$ is 0.96 on four different plates, 0.88 to 0.92 between frozen and fresh sera loaded on the same plate and 0.82 to 0.88 when loaded on different plates one month apart while it is 0.88 to 0.96 when loaded with frozen sera at the same time on different plates, 0.92 to 0.96 on the same plates. Both the LuLISA and ELISA are sensitive to the storage quality of the sera, in particular to the thawing conditions which contribute to the denaturation of proteins, in particular albumin, and which cause the formation of aggregates that can pull down immunoglobulins and alter the concentration of soluble specific Ig.

White 384-well plates with flat bottoms (Fluoronunc C384 Maxisorp, Nunc) were coated with either 1 μg/mL of Nucleoprotein or Spike protein in PBS buffer, 50 μL/well for 3 hours at room temperature or overnight at 4°C. Wells were washed using a plate washer (Zoom, Berthold Technologies, Germany) two cycles of three times with 100 μL of PBS/Tween 20 0.1%. Sera were diluted 200 times in PBS, non-fat milk 3% and Tween 20 0.1%. 50μL of serum dilutions were incubated 1 hour at room temperature in their respective wells. Wells were washed two cycles of three times with 100 μL of PBS/Tween 20 0.1%. Purified Fc1-nanoKAZ 1 ng/mL ($400 \cdot 10^6$ $RLU \cdot s^{-1} \cdot mL^{-1}$) in PBS, non-fat milk 3% and Tween 20 0.1% was loaded (50 μL/well) and incubated 30 min at room temperature. Wells were washed two cycles of three times with 100 μL of PBS/Tween 20 0.1%. Plates can be stored at this step in PBS until measurements. Just before reading, each of the wells were emptied and loaded with 50 μL of Q-108 at 13 μM. The plate was orbitaly shaken for 3 seconds and the collected photons were counted during 1 sec per well and measured 3 times in a plate luminometer (Mithras2, Berthold, Wildbad, Germany).

***S-Pseudo-typed neutralization assay***

***Pseudo-virus production and permissive cell line generation***

Pseudo-typed vectors were produced and titrated as previously described [11]. Adaptation of the protocol are : cells were co-transfected with calcium-phosphate precipitation protocol with 10 μg of packaging plasmid encoding for gag-pol-tat-rev proteins (p8.74), 10μg of vector plasmid (pTrip-CMV) expressing luciferase Firefly reporter and 5 μg of envelop plasmid expressing a codon-optimized full-length S SARS-Cov-2 (UniProtKB ID: P0DTC2) sequence amplified by PCR from pMK-RQ_S-2019-nCoV with adaptative primers and introduced by BamHI/XhoI restriction/ligation in a pCMV plasmid. Pseudo-typed vectors were harvested at day 2 post-transfection. Functional titer (TU) was determined by SYBRgreen qPCR after transduction of a stable HEK 293T-hACE2 cell line using two couples of primers to either quantify vector genome (forward CMV: ACT GCC AAA ACC GCA TCA CC reverse CMV: AAT

GAC GGT AAA TGG CCC GC) or cell genome (forward GADPH: TCT CCT CTG ACT TCA ACA GC reverse GADPH: CCC TGC ACT TTT TAA GAG CC). To generate the permissive cell line, HEK 293T cells were transduced at MOI 20 with an integrative lentiviral vector expressing cDNA human ACE2 (UniProtKB ID: Q9BYF1) codon-optimized gene (Eurofins) under the control of UBC promoter. Clones were generated by limiting dilution and selected for their permissivity to SARS-CoV-2 S pseudo-typed lentiviral vector transduction with a Luciferase Assay System (Promega).

***Pseudo-neutralization assay protocol***

First, sera were decomplemented at 56°C during 30 min in a water bath. To determine EC50 serum dilutions (from 1/40 to 1/40960 by successive 4-fold dilutions) are mixed and co-incubated with 300 TU of pseudo-typed vector at room temperature during 30 minutes under agitation. Both, serum and vector are diluted in culture medium DMEM-glutamax (Gibco) + 10% FCS (Gibco) + Pen/Strep (Gibco). Mix is then plated in tissue culture treated black 96-well plate clear bottom (Costar) with 20 000 HEK 293T-hACE2 cells in suspension. To prepare the cell suspension, the cell flask is washed with DPBS twice (Gibco) and cells are individualized with DPBS + 0.1% EDTA (Sigma-Aldrich) to preserve hACE2 protein. After 48h incubation at 37°C 5% $CO_2$, the medium is completely removed by aspiration and bioluminescence is measured using a Luciferase Assay System (Promega) on an EnSpire plate reader (PerkinElmer).

***Statistical analyses***

Seropositivity was defined as the presence of detectable anti-SARS-CoV-2 antibodies against N or S. The proportion of seropositive samples was compared by time between onset of symptoms and collection of blood sample using chi-square test.

LuLISA, and pseudo-neutralization of sera were compared by delay since onset of symptoms using the Kruskall-Wallis non-parametric test. The chi-square test was used to evaluate the association between investigated factors and neutralization levels.

All analyses were performed using GraphPad Prism 8 (GraphPad Software, LLC).

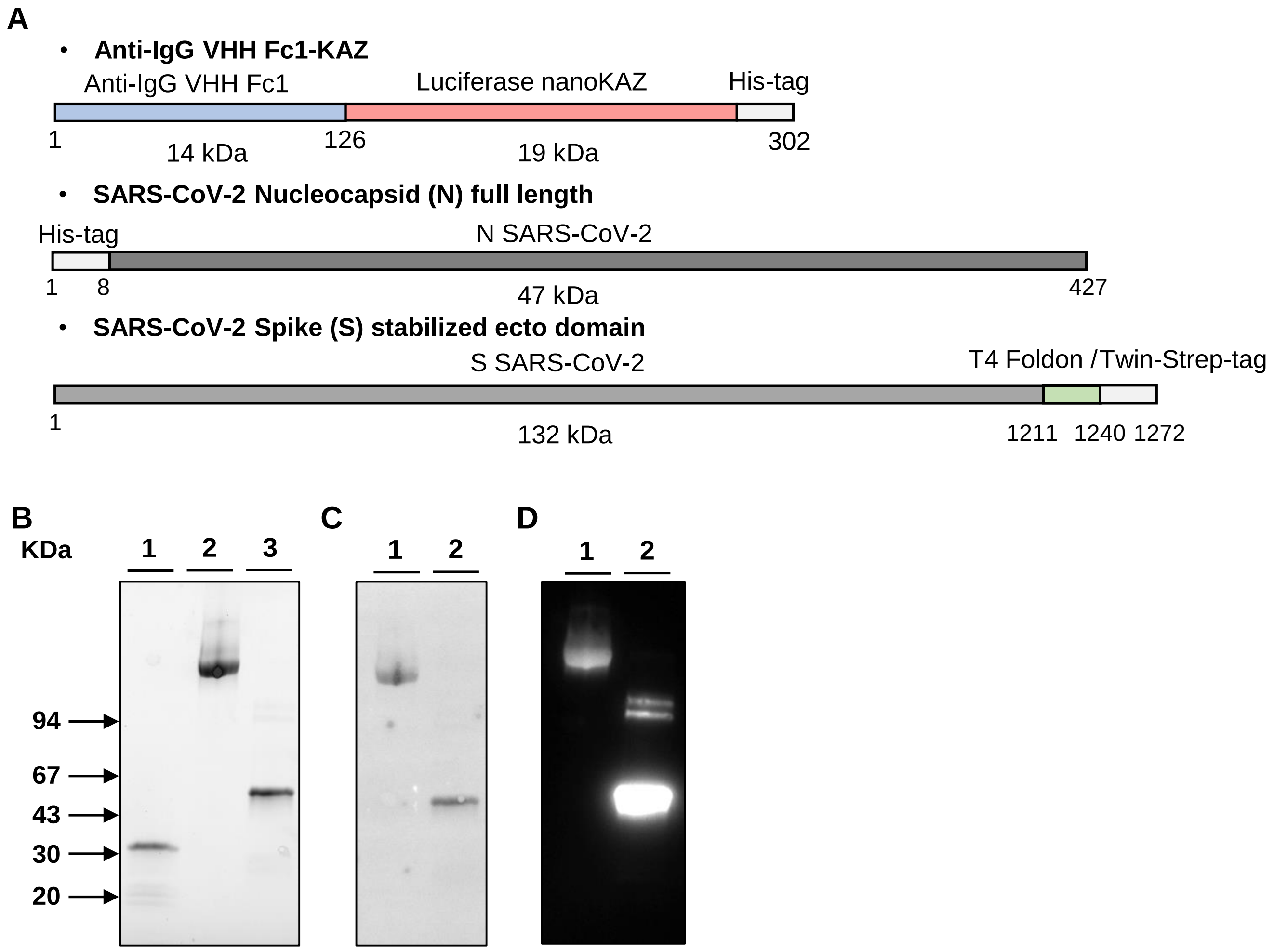


**Figure S1 Description of anti-IgG VHH Fc1-KAZ and of SARS-CoV2 viral protein targets nucleoprotein and spike.**

(**A**) Scheme of protein topology used in the study. Quality control of proteins used as targets on SDS-polyacrylamide gel electrophoresis. Proteins were run on a 4-15% gradient gel and visualized by UV-induced fluorescence of tryptophan-bound trihalo compounds (**B**) or by western blot analysis after transfer onto the PDVF proteins using stain-free system (Bio-Rad) (**C**) or luminescence (**D**) Lane 1: Purified VHH-anti IgG Fc1-nanoKaz (2 μg); Lane 2, purified protein SARS-CoV Spike (6 μg); Lane 3 Nucleoprotein (3 μg). Molecular weights are indicated by arrows.

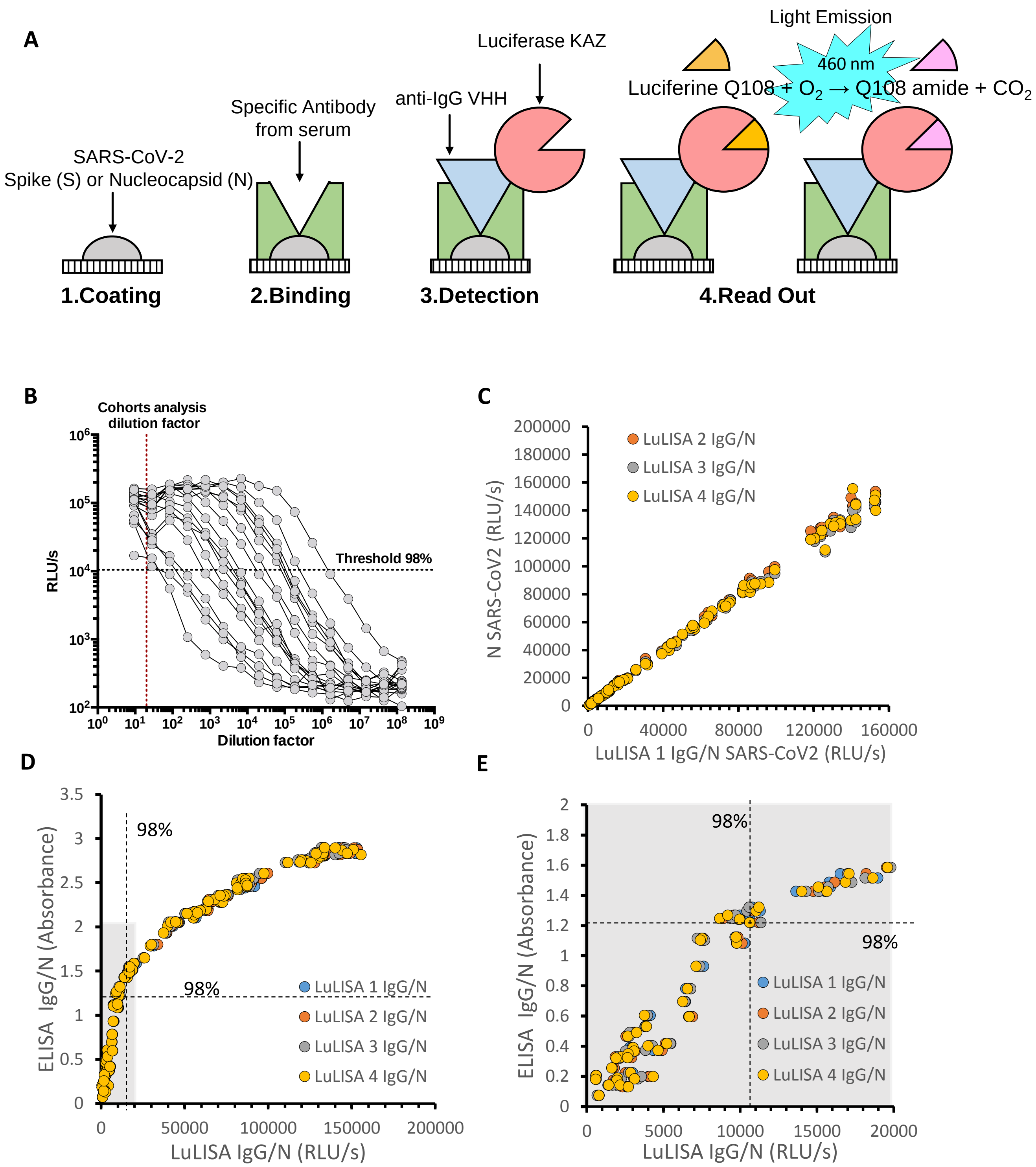


**Figure S2 Luciferase-linked immuno-sorbent assay**

(**A**) Scheme of the LuLISA assay. (**B**) LuLISA of titration of N-specific IgG from patients' sera (Hospital Cochin). Serial dilutions of sera from $10^{-1}$ to $10^{-8}$ in PBS. (**C**) Four repeats of IgG/N LuLISA on the same series of 96 diluted sera in PBS (1/200). (**D**) Comparison of the 4 IgG/N LuLISA vs IgG/N ELISA (HRP-anti-human IgG) showing the large dynamic scale of LuLISA vs ELISA. (**E**) The grey zone in D is zoomed showing the higher sensibility of LuLISA (low dispersion of LuLISA values, high dispersion of ELISA values below DO 1.2 for dilution 1/200) and the loss of linearity beyond absorbance 1.2. Threshold 98%, DO 1.2, 10400 RLU/s are indicated with dashed lines.

Anna et al. Supplementary Appendix 1 Figure S3

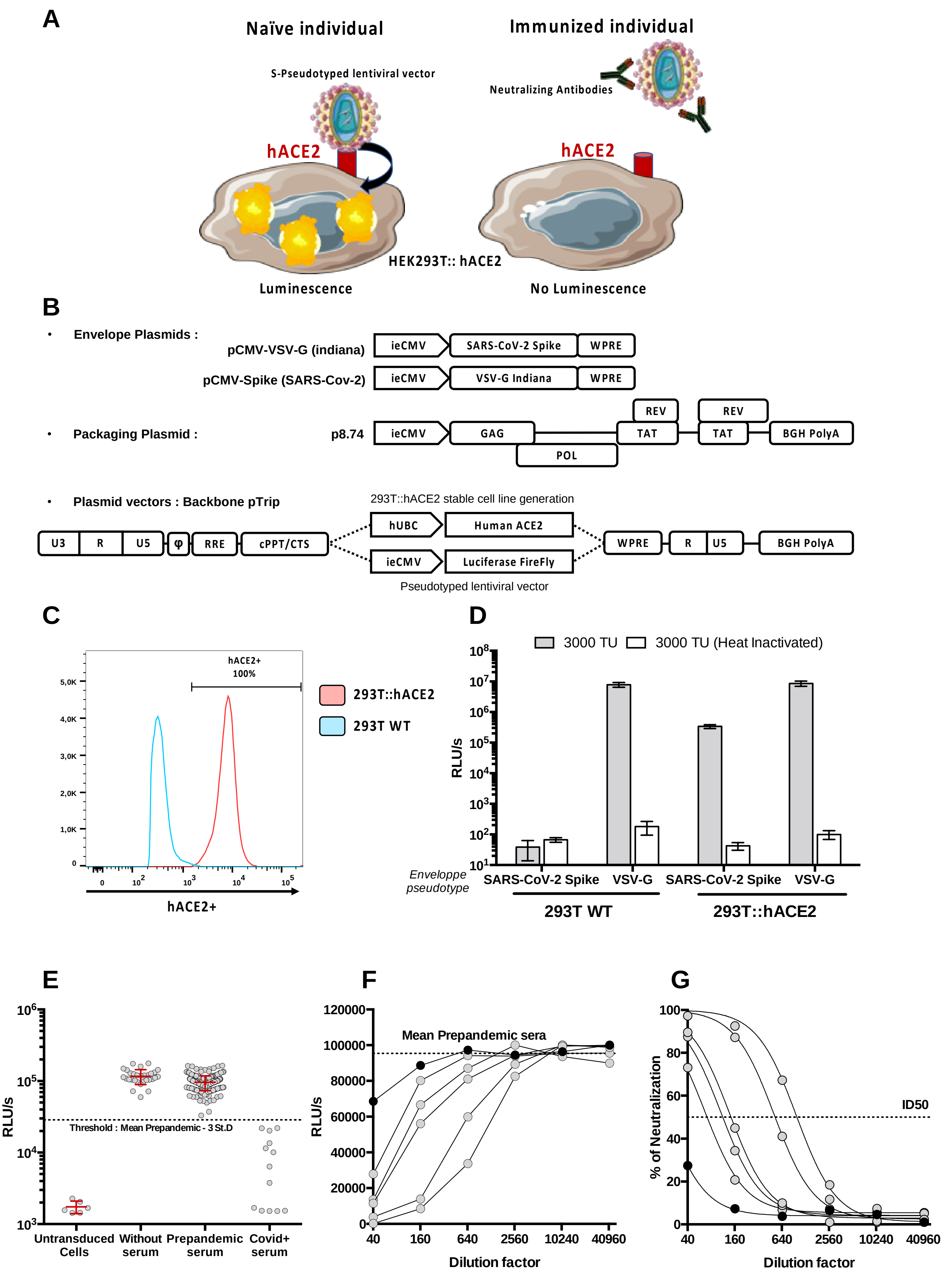


### Figure S3: Pseudo-neutralisation assay (PNT)

**(A) Schematic representation of Pseudo-neutralization assay.**
**(B) Production plasmids used to generate lentiviral vectors.**
ieCMV: human Immediate early Cytomegalovirus promoter. WPRE : Woodchuck Post-translational Regulatory Element. VSV-G Glycoprotein from Vesicular Stomatitis Virus (serotype Indiana). Gag-Pol-Tat-rev proteins from HIV-1 Strain NDK. BGH Poly A : Polyadenylation Site from Bovine Growth Hormone gene. All following fragments come from HIV-1 Strain NL4.3 : U3RU5 : Full length Long Terminal Repeat, φ : Encapsidation sequence, RRE: Rev Response Element, RU5 : Truncated Long Terminal Repeat. hUBC : human Ubiquitine C promoter.
**(C) Control of hACE2 expression in 293T::hACE2 reporter cell line by FACS.** To verify the expression and membrane location of hACE2 protein, parental cell line (293T WT) and hACE2 expressing clone (293T::hACE2) were first labelled polyclonal antibodies anti-hACE2 (AF933, R&D) then a PE-coupled secondary antibody (A32849, Invitrogen). Samples were acquired on a Attune FACS (ThermoFisher).
**(D) Control of 293T::hACE2 permissivity et specificity to S-pseudotyped lentiviral vector transduction.** Parental cell line and hACE2 clone were transduced with S-pseudotyped lentiviral vector (or VSV-G, an amphotropic envelope as control). To assess that signal is specific to vector particles, equivalent heat-inactivated samples (70°C 30min in a water bath) were also tested.
**(E) Threshold determination on different serum collections.** To setup up the experimentation, Min-max values are determined on untransduced cells and prepandemic serum (dilution 1/40) respectively, Covid-19 patients are used as positive controls. To define positiveness threshold with a confidence index >99%, this value is set at Mean (prepandemic) – 3 Standard deviation. During sample analysis, all samples are firstly evaluated for positiveness at dilution 1/40. If the value is below threshold, then ID50 is determined as described below in a second experiment.
**F-G Dilution curves and ID50 determination**. First, raw data (grey points) **(F)** are transformed into percentage of neutralization **(G)**. This percentage is determined according to mean of prepandemic serums (0%) and untransduced cells (100%) values. Second, a non-linear regression is performed (plotted line in graph G) to determine the theoretical dilution that give a 50% inhibition (ID50). A prepandemic sample (Black point) is shown as negative example. Detection limit was set at 40 considering the maximum reached value by prepandemic samples.

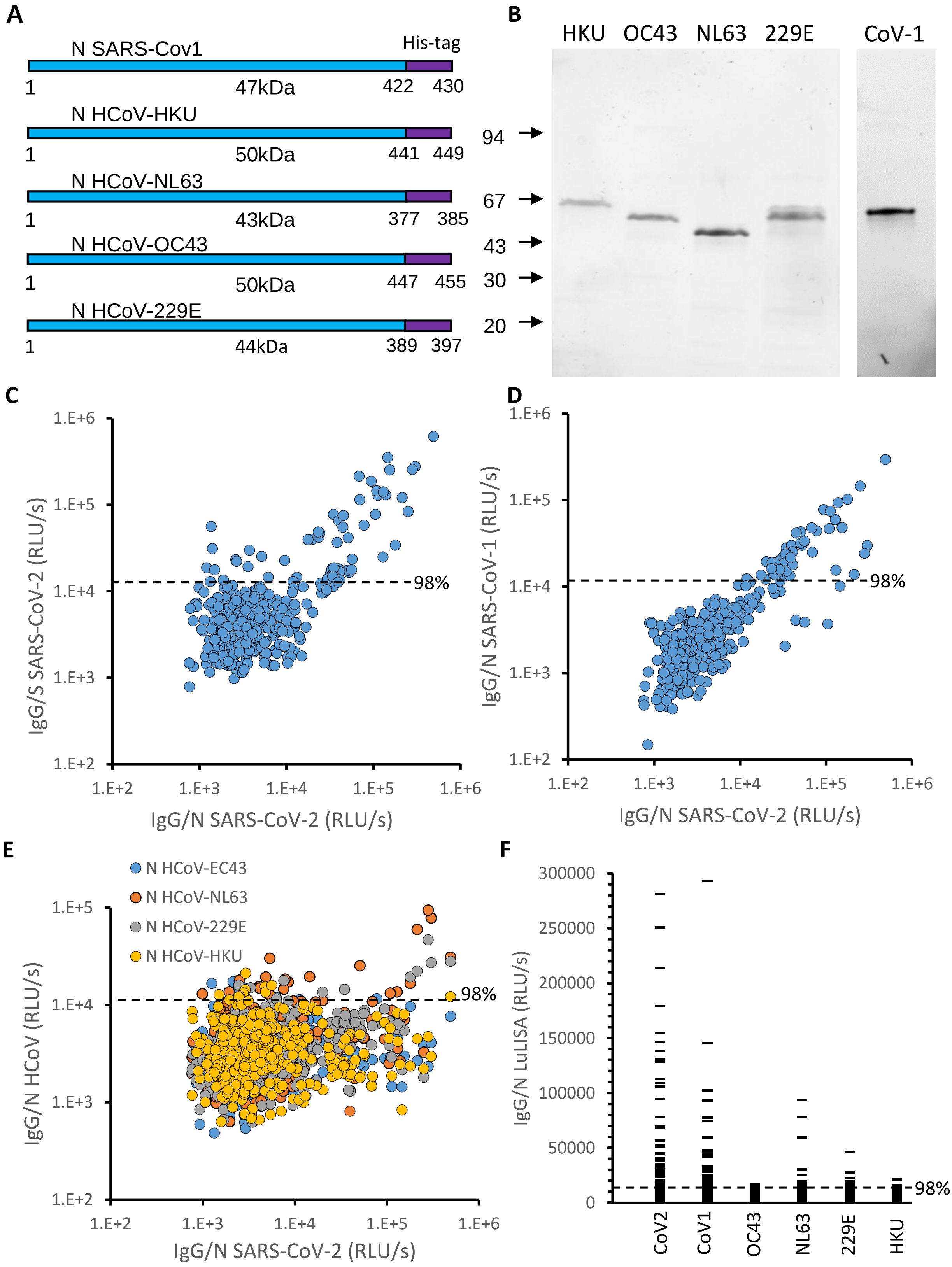


**Figure S4 Specificity of assayed IgG against nucleoprotein of SARS-CoV2 vs SARS-CoV1 and seasonal virus HCoV:** (**A**) Protein constructs with His Tag (LEHHHHHH). (**B**) Control of expressed and purified proteins used as targets using SDS-polyacrylamide gel electrophoresis. The gel is an acrylamide gradient from 4 to 15% . Lane 1: purified protein Nucleocapsid from HCoV-HKU, HCoV-OC43, HCoV-NL63, HCoV-229E and SARS-CoV1 (stain free system, BioRad) . (**C**) IgG/S vs IgG/N SARS-CoV2 from Curie series (n=376) . (**D**) IgG LuLISA against N SARS-CoV1 vs SARS-CoV2 (**E**) IgG LuLISA against N HCoV vs SARS-CoV2 from Curie series (n=376). (F) IgG LuLISA against N SARS-CoV2, SARS-CoV1, HCoV-OC43, HCoV-NL63, HCoV-229E, HCoV-HKU.

**Anna et al. Supplementary Appendix 1 Figure S5**

```
HCoV-229E           ---------------------MATVKWADASEPQR-----GRQ----------------G  18
HCoV-NL63           ---------------------MASVNWADDRAA--------RK----------------K  15
SARS-CoV-2          MSDN-G--------PQNQ-RNAPRITFGGPSDSTGSNQN-GER---SGARSKQRRPQGLP  46
SARS-CoV-1          MSDN-G--------PQSNQRSAPRITFGGPTDSTDNNQN-GGR---NGARPKQRRPQGLP  47
HCOV-OC43           MSFTPGKQSSSRASSGNRSGNG-ILKWADQSDQFRNVQTRGRRAQPKQTATSQQPSGGNV  59
HCoV-HKU1           MSYTPGHYAGSRSSSGNRSGILKKTSWADQSERNYQTFNRGRKTQPKFTVST-QP-QGNT  58
                                             .:..            .:

HCoV-229E           RIPYSLYSPLLVDS-EQPWKVIPRNLVPINKKD-KNKLIGYWNVQ--KRFRTRKGKRVDL  74
HCoV-NL63           FPPPSFYMPLLVSSDKAPYRVIPRNLVPIGKGN-KDEQIGYWNVQ--ERWRMRRGQRVDL  72
Sars-CoV-2          NNTASWFTALTQHGK-EDLKFPRGQGVPINTNSSPDDQIGYYRRA-TRRIRGGDGKMKDL  104
SARS-CoV-1          NNTASWFTALTQHGK-EELRFPRGQGVPINTNSGPDDQIGYYRRA-TRRVRGGDGKMKEL  105
HCOV-OC43           VPYYSWFSGITQFQKGKEFEFVEGQGVPIAPGVPATEAKGYWYRHNRRSFKTADGNQRQL  119
HCoV-HKU1           IPHYSWFSGITQFQKGRDFKFSDGQGVPIAFGVPPSEAKGYWYRHSRRSFKTADGQQKQL  118
                        * :  :         ..   : ***       .  **:     .  :   *:  :*

HCoV-229E           SPKLHFYYLGTGPHKDAKFRERVEGVVWVAVDGAKTEPTG-YGVRRKNSEPEIPH-F--N  130
HCoV-NL63           PPKVHFYYLGTGPHKDLKFRQRSDGVVWVAKEGAKTVNTS-LGNRKRNQKPLEPK-F--S  128
Sars-CoV-2          SPRWYFYYLGTGPEAGLPYGANKDGIIWVATEGALNTPKDHIGTRNPANNAAIVLQLPQG  164
SARS-CoV-1          SPRWYFYYLGTGPEASLPYGANKEGIVWVATEGALNTPKDHIGTRNPNNNAAIVLQLPQG  165
HCOV-OC43           LPRWYFYYLGTGPHAKDQYGTDIDGVYWVASNQADVNTPADIVDRDPSSDEAIPTRFPPG  179
HCoV-HKU1           LPRWYFYYLGTGPYANASYGESLEGVFWVANHQADTSTPSDVSSRDPTTQEAIPTRFPPG  178
                     *: :********     :    :*: *** . *          *    .      :  .

HCoV-229E           QKLPNGVTVVEEPDSRA-----PSRSQSR--SQSRGRGESKPQSRNPSSDRNHNSQDDIM  183
HCoV-NL63           IALPPELSVVEFEDRSNNSSRASSRSSTR--NNSRDSSRSTSRQQSRTRSDSNQSSSDLV  186
Sars-CoV-2          TTLPKGFYAEGSRGG----SQASSRSSSRSRNSSRNS---TPGS-SRGTSPARMAGNG--  214
SARS-CoV-1          TTLPKGFYAEGSRGG----SQASSRSSSRSRNSSRNS---TPGS-SRGNSPARMAGNG--  215
HCOV-OC43           TVLPQGYYIEGSGRS----APN-SRSTSR--TSSRAS---SAGSRSRANSGNRTPTSGVT  229
HCoV-HKU1           TILPQGYYVEGSGRS----ASN-SRPGSR--SQSRGP---NNRSLSRSNSNFRHSDSIVK  228
                      **                   **  :*  ..**     .  . .   .  .   .

HCoV-229E           KAVAAALKSLGFDKPQEKDKKSAKTGTPKPSRNQSPASSQTSAKSLARSQSSETKEQKHE  243
HCoV-NL63           AAVTLALKNLGFDNQSKSPSSS---GTSTPKKPNKPL-----------SQPRADKPSQ--  230
Sars-CoV-2          --GDAALALLLLDRLNQLESKMSGKGQ------QQQ------------GQTVTKKSAAEA  254
SARS-CoV-1          --GETALALLLLDRLNQLESKVSGKGQ------QQQ------------GQTVTKKSAAEA  255
HCOV-OC43           PDMADQIASLVLAKLGKDAT----K-P------QQV------------TKHTAKEVRQKI  266
HCoV-HKU1           PDMADEIANLVLAKLGKDS-----K-P------QQV------------TKQNAKEIRHKI  264
                          :  * : .  :                :.

HCoV-229E           MQKPRWKRQPNDDVTSNVTQCFGPRDLDH---NFGSAGVVANGVKAKGYAQFAELVPSTA  300
HCoV-NL63           LKKPRWKRVPTRE--ENVIQCFGPRDFNH---NMGDSDLVQNGVDAKGFAQLAELIPNQA  285
Sars-CoV-2          SKKPRQKPRTATKA-YNVTQAFGRRGPEQTQGNFGDQELIRQGTDYKHWPQIAQFAPSAS  312
SARS-CoV-1          SKKPRQK-RTATKQ-YNVTQAFGRRGPEQTQGNFGDQDLIRQGTDYKHWPQIAQFAPSAS  313
HCOV-OC43           LNKPRQKRSPNKQ--CTVQQCFGKRGPNQ---NFGGGEMLKLGTSDPQFPILAELAPTAG  321
HCoV-HKU1           LTKPRQKRTPNKH--CNVQQCFGKRGPSQ---NFGNAEMLKLGTNDPQFAILAELAPTPG  319
                      *** **  .     .* *.** *. .:   *:*.  ::  *..   :* :*:: *. .

HCoV-229E           AMLFDSHIVSKESG-----------NTVVLTFTTRVTVPKDHPHLGKF----LEELNAFT  345
HCoV-NL63           ALFFDSEVSTDEVG-----------DNVQITYTYKMLVAKDNKNLPKF----IEQISAFT  330
Sars-CoV-2          AFFGMSRIGMEVTP-----------SGTWLTYTGAIKLDDKDPNFKDQVILLNKHIDAYK  361
SARS-CoV-1          AFFGMSRIGMEVTP-----------SGTWLTYTGAIKLDDKDPQFKDNVILLNKHIDAYK  361
HCOV-OC43           AFFFGSRLELAKVQNLSGNPDEPQKDVYELRYNGAIRFDSTLSGFETIMKVLNENLNAYQ  381
HCoV-HKU1           AFFFGSKLDLVKRD---SEADSPVKDVFELHYSGSIRFDSTLPGFETIMKVLEENLNAYV  376
                    *::  *.:                 .   : :.  : . .    :        :.:.*:

HCoV-229E           REMQQHPLLNP-------SALEFNP------------SQTSPATAE--------PVRDEV  378
HCoV-NL63           KPSSIKEMQSQ-------SSHVAQN----------TVLNASIPESK--------PLADDD  365
Sars-CoV-2          TFPPTEPKKDKKKKADETQALPQRQKKQQT-------VTL--------------LPAADL  400
SARS-CoV-1          TFPPTEPKKDKKKKTDEAQPLPQRQKKQPT-------VTL--------------LPAADM  401
HCOV-OC43           QQDG---MM-------NMSPKPQRQRGHKNGQGENDNISVAVPKSRVQQNKSRELTAEDI  431
HCoV-HKU1           NSNQNTDSD-------SLSSKPQRKRGVKQLPEQFDSLNLSA----GTQHISNDFTPEDH  425
                                      .    .                                  :

HCoV-229E           SIETDIIDEVN---------              389
HCoV-NL63           SAIIEIVNEVLH*-------              377
Sars-CoV-2          DDFSKQLQQSMS--SADSTQA*            419
SARS-CoV-1          DDFSRQLQNSMSGASADSTQA*            422
HCOV-OC43           SLL-KKMDEPYTE---DTSE*-            447
HCoV-HKU1           SLL-ATLDDPYVE---DSVA*-            441
                    .     :::
```

**Figure S5 Sequence comparison of SARS-CoV2, SARS-CoV1 and seasonal virus nucleoproteins**

Multiple sequence alignments of the four human seasonal coronavirus (HCoV) and the SARS-CoV Nucleoproteins. The alignment was performed using Clustal Omega software from [http://www.ebi.ac.uk/Tools/msa/clustalo](http://www.ebi.ac.uk/Tools/msa/clustalo).
Grey background indicates conserved residues in at least 4 sequences. Conserved residues within the two alphacoronavirus and the two beta-coronavirus are highlighted in yellow and blue, respectively with SarS-CoV when applicable. Residues in red denote aromatic residues potentially involved in base stacking interactions when binding to RNA. Red dashed boxes enlight conserved patterns.

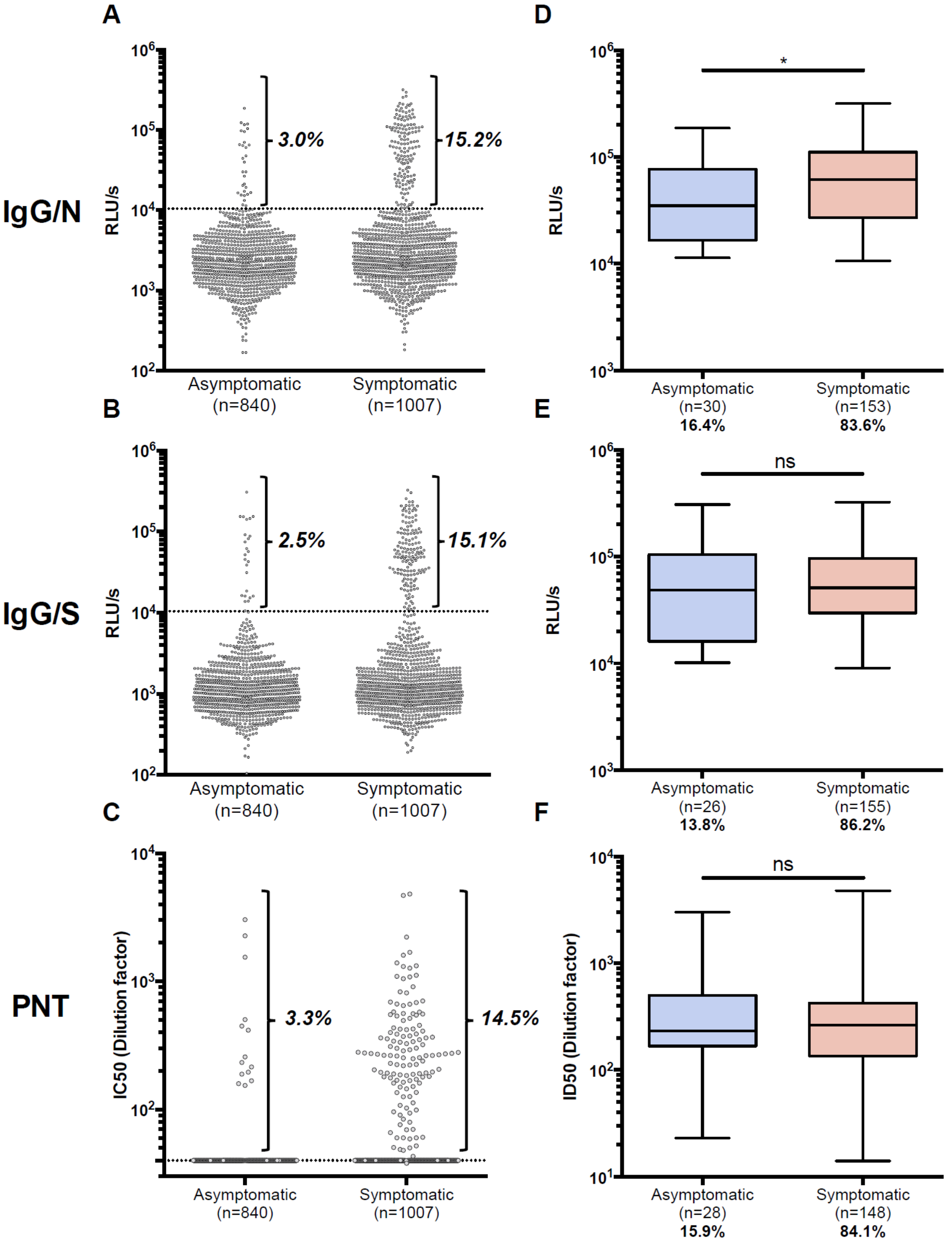


**Figure S6: Serological profile comparison between asymptomatic and symptomatic workers**

Institut Curie workers were separated in asymptomatic and symptomatic groups according to survey answers. Symptomatic workers reported at least one symptom (listed in table II). Global seroprevalence in LuLISA IgG/N **(A),** LuLISA IgG/S **(B),** PNT **(C)** are shown as raw values. Proportion of seropositive workers is indicated in bolt**.** Proportion of asymptomatic and symptomatic among seropositive workers in LuLISA IgG/N **(D),** LuLISA IgG/S **(E),** PNT **(F)** are represented by whisker-plots. Proportion among total seropositive workers is indicated below in bold. Statistical significance was determined using a Mann-Whitney test (*:$p<0.05$; ns: non significant).

Anna et al. Supplementary Information- Appendix 1 - Figure S7

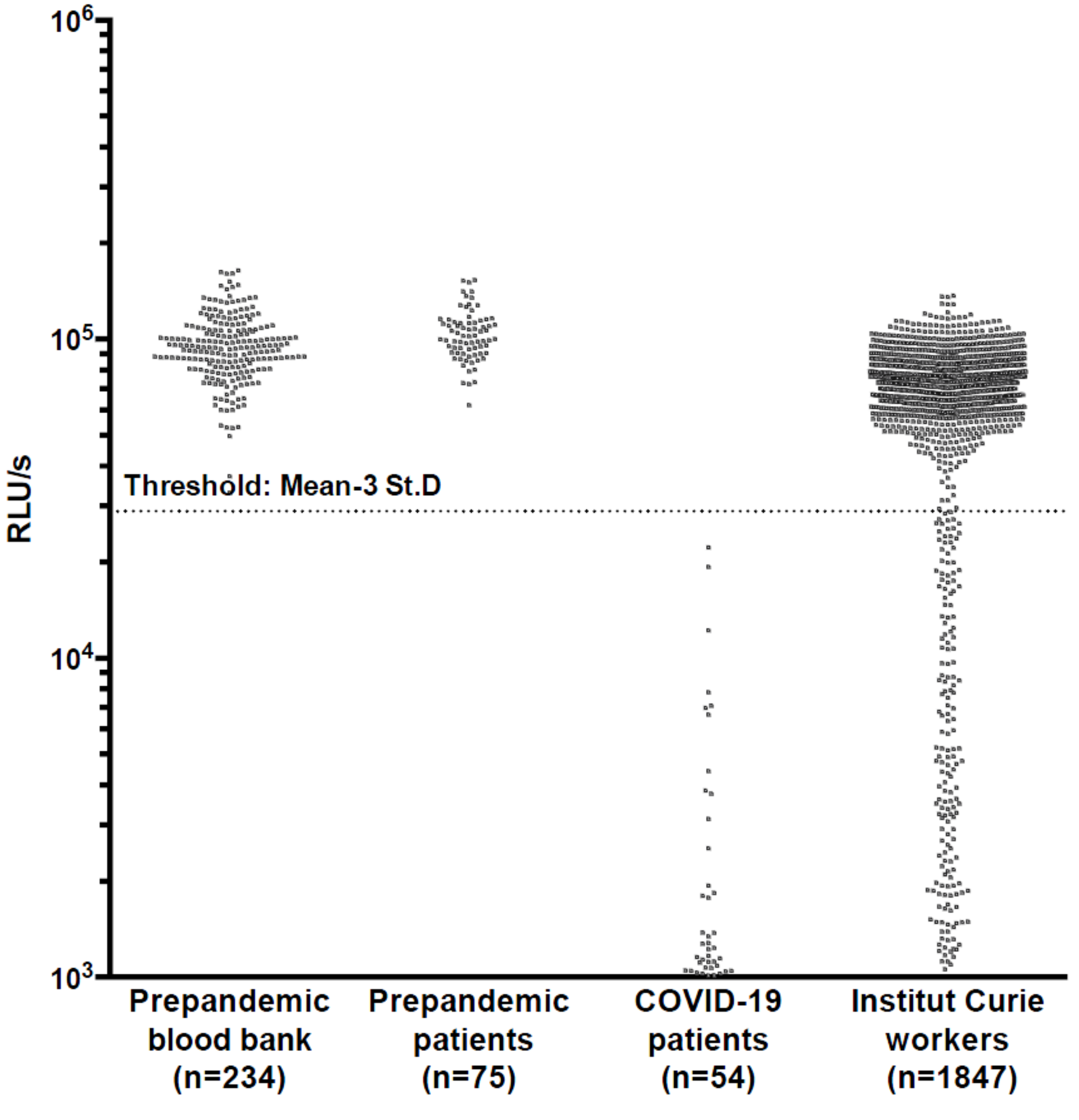


**Figure S7:** **PNT raw data for cohorts analysis**

Serum dilution factor:1/40. Positive samples are below the threshold level. Threshold is calculated on raw values of prepandemic sera from a blood bank (see Figure S3). ID50 was determined on positive samples (see Figure 1 and Figure S3)

**SUPPORTING INFORMATION : APPENDIX 2**
This appendix has been provided by the authors to give readers additional information about their work.
Supplement to: Anna, Goyard, et al.

**High seroprevalence but short-lived immune response to SARS-CoV-2 infection in Paris**

François Anna[1]*, Sophie Goyard[2,3]*, Ana Ines Lalanne[4,5]*, Fabien Nevo[1,6], Marion Gransagne[7], Philippe Souque[6], Delphine Louis[4,5], Véronique Gillon[8], Isabelle Turbiez[8], François-Clement Bidard[5,9,10], Aline Gobillion[11], Alexia Savignoni[11], Maude Guillot-Delost[5,12], François Dejardin[13], Evelyne Dufour[13], Stéphane Petres[13], Odile Richard-Le Goff[14], Zaineb Choucha[7], Olivier Helynck[15], Yves L. Janin[15], Nicolas Escriou[7], Pierre Charneau[1,6], Franck Perez[16], Thierry Rose[2,3]**, Olivier Lantz[4,5,12]**

**Author affiliations**

1. Theravectys, Paris, France
2. Unit of Lymphocyte Cell Biology, Immunology Department, Institut Pasteur, Paris, 75015, France
3. INSERM 1221, Institut Pasteur, Paris, 75015, France
4. Laboratoire d'Immunologie Clinique, Institut Curie, Paris, 75005, France.
5. Centre d'Investigation Clinique en Biothérapie, Institut Curie (CIC-BT1428), Paris, 75005, France
6. Unit of Molecular Virology and Vaccinology, Virology Department, Institut Pasteur, Paris, 75015, France
7. Innovation Laboratory: Vaccines, Institut Pasteur, Paris, 75015, France
8. Direction of the Clinical Research, Institut Curie, Paris, 75005, France.
9. Medical Oncology Department, Institut Curie, Paris, 75005, France
10. UVSQ, Paris-Saclay University, Saint-Cloud, 92210, France
11. Biometry, Institut Curie, Paris, 75005, France
12. INSERM U932, PSL University, Institut Curie, Paris, 75005, France
13. Production and purification of recombinant proteins technological platform, Paris, 75015, France
14. Unit of Antibody in Therapy and Pathology, Institut Pasteur, Paris, 75015, France
15. Unit of Chemistry and Biocatalysis, Institut Pasteur, UMR 3523 CNRS, Paris, 75015, France
16. Unit of Cell Biology and Cancer, Institut Curie, Paris, 75005, France

**Anna et al, Supplementary Appendix 2**

This questionnaire was submitted with the description of the study for ethical approval by the Comité de Protection des Personnes Méditerranée III (2020.04.18 bis_ 20.04.16.49458, 27/4/2020) registered in the clinical trial database (NCT04369066). The items of this questionnaire were recorded in the Curiosa Web-based survey.

**Questionnaire**

- Working Site (Paris Hospital/Saint Cloud Hospital/Orsay site/Administrative Staff/Research Center)
- Age (years)
- Weight (kg)
- Height (cm)
- Confinement (Yes/No)
- Confinement start (date)
- Working mode at the time of the blood withdrawal (Site /Institute and home-office)
- Symptoms (fever, tiredness, cough, shortness of breath, difficulty breathing, muscle aches, conjunctivitis, rhinitis, anosmia/ageusia, unusual headache, digestive disorders (vomiting, diarrhea))
- Symptom onset (date)
- Symptom end (date)
- PCR_assay done (yes/no)
- PCR_assay (date)
- PCR result (Positive/Negative)
- Treatment (hospitalization/systemic treatment)
- Hospitalization (days)
- Intensive care unit (yes/no)

**SUPPORTING INFORMATION : APPENDIX 3**
This appendix has been provided by the authors to give readers additional information about their work.
Supplement to: Anna, Goyard, et al.

**High seroprevalence but short-lived immune response to SARS-CoV-2 infection in Paris**

François Anna[1]*, Sophie Goyard[2,3]*, Ana Ines Lalanne[4,5]*, Fabien Nevo[1,6], Marion Gransagne[7], Philippe Souque[6], Delphine Louis[4,5], Véronique Gillon[8], Isabelle Turbiez[8], François-Clement Bidard[5,9,10], Aline Gobillion[11], Alexia Savignoni[11], Maude Guillot-Delost[5,12], François Dejardin[13], Evelyne Dufour[13], Stéphane Petres[13], Odile Richard-Le Goff[14], Zaineb Choucha[7], Olivier Helynck[15], Yves L. Janin[15], Nicolas Escriou[7], Pierre Charneau[1,6], Franck Perez[16], Thierry Rose[2,3]**, Olivier Lantz[4,5,12]**

**Author affiliations**

1. Theravectys, Paris, France
2. Unit of Lymphocyte Cell Biology, Immunology Department, Institut Pasteur, Paris, 75015, France
3. INSERM 1221, Institut Pasteur, Paris, 75015, France
4. Laboratoire d'Immunologie Clinique, Institut Curie, Paris, 75005, France.
5. Centre d'Investigation Clinique en Biothérapie, Institut Curie (CIC-BT1428), Paris, 75005, France
6. Unit of Molecular Virology and Vaccinology, Virology Department, Institut Pasteur, Paris, 75015, France
7. Innovation Laboratory: Vaccines, Institut Pasteur, Paris, 75015, France
8. Direction of the Clinical Research, Institut Curie, Paris, 75005, France.
9. Medical Oncology Department, Institut Curie, Paris, 75005, France
10. UVSQ, Paris-Saclay University, Saint-Cloud, 92210, France
11. Biometry, Institut Curie, Paris, 75005, France
12. INSERM U932, PSL University, Institut Curie, Paris, 75005, France
13. Production and purification of recombinant proteins technological platform, Paris, 75015, France
14. Unit of Antibody in Therapy and Pathology, Institut Pasteur, Paris, 75015, France
15. Unit of Chemistry and Biocatalysis, Institut Pasteur, UMR 3523 CNRS, Paris, 75015, France
16. Unit of Cell Biology and Cancer, Institut Curie, Paris, 75005, France

**SUPPLEMENTARY APPENDIX 3 to Anna, et al.**
**High seroprevalence but short-lived immune response to SARS-CoV-2 infection in Paris**

| | | | | |
|---|---|---|---|---|
| LuLISA IgG/N | positive if >13400 RLU/s | Threshold 98% specificity | Symptoms | RT-qPCR | RT-qPCR results |
| LuLISA IgG/S | positive if >8400 RLU/s | | negative=0 | no=0 | negative=0 |
| PNT | positive if > 0 % | | positive=1 | yes=1 | positive=1 |

| Individuals | | | | | | | | | | | | Symptoms from questionnaire | | | | | | | | | | | | RT-qPCR | |
|---|---|---|---|---|---|---|---|---|---|---|---|---|---|---|---|---|---|---|---|---|---|---|---|---|---|
| ID | sex | age | Latreal flow test | LuLISA IgG/N M0 | LuLISA IgG/S M0 | PNT M0 | seropositivity M0 | LuLISA IgG/N M1 | LuLISA IgG/S M1 | PNT M1 | seropositivity M1 | any symptom | Fever | Tiredness | Cough | Short breath | ficulty breathi | Muscle ache | Conjonctivitis | Rhinitis | nosmia/ageus | nusual headac | gestive disord | RT-qPCR | T-qPCR results |
| 1 | F | 39-82 | | 1471 | 1384 | 0 | 0 | 5325 | 1123 | 0 | 0 | 1 | 0 | 1 | 0 | 0 | 0 | 1 | 0 | 1 | 0 | 1 | 0 | 0 | |
| 2 | F | 19-38 | | 6518 | 3614 | 0 | 0 | #N/A | #N/A | #N/A | #N/A | 1 | 0 | 1 | 1 | 0 | 0 | 0 | 0 | 1 | 0 | 1 | 1 | 0 | |
| 3 | F | 39-82 | | 3142 | 2333 | 0 | 0 | 1692 | 2391 | 0 | 0 | 1 | 1 | 1 | 1 | 1 | 0 | 1 | 0 | 0 | 0 | 1 | 1 | 1 | 0 |
| 4 | F | 19-38 | | 2978 | 706 | 0 | 0 | 1322 | 1132 | 0 | 0 | 1 | 0 | 1 | 0 | 0 | 0 | 0 | 1 | 1 | 0 | 1 | 1 | 0 | |
| 5 | F | 39-82 | | 3680 | 2039 | 0 | 0 | 2457 | 2349 | 0 | 0 | 1 | 0 | 0 | 0 | 0 | 0 | 0 | 0 | 0 | 0 | 1 | 1 | 0 | |
| 6 | F | 19-38 | | 27456 | 42832 | 67 | 1 | 25056 | 64694 | 85 | 1 | 1 | 1 | 1 | 0 | 1 | 0 | 1 | 0 | 0 | 1 | 1 | 1 | 1 | 1 |
| 7 | F | 19-38 | | 3876 | 1002 | 0 | 0 | 1830 | 531 | 0 | 0 | 0 | 0 | 0 | 0 | 0 | 0 | 0 | 0 | 0 | 0 | 0 | 0 | 0 | |
| 8 | F | 39-82 | | 2230 | 750 | 0 | 0 | #N/A | #N/A | #N/A | #N/A | 1 | 0 | 1 | 1 | 1 | 1 | 1 | 1 | 0 | 0 | 1 | 1 | 1 | 0 |
| 9 | F | 39-82 | | 4825 | 834 | 0 | 0 | 2066 | 728 | 0 | 0 | 1 | 0 | 1 | 0 | 0 | 0 | 0 | 0 | 0 | 0 | 0 | 0 | 0 | |
| 10 | F | 39-82 | | 3386 | 2326 | 0 | 0 | #N/A | #N/A | #N/A | #N/A | 1 | 0 | 1 | 1 | 1 | 1 | 0 | 0 | 0 | 0 | 1 | 1 | 1 | 0 |
| 11 | F | 19-38 | | 1728 | 651 | 0 | 0 | 1064 | 1356 | 0 | 0 | 0 | 0 | 0 | 0 | 0 | 0 | 0 | 0 | 0 | 0 | 0 | 0 | 0 | |
| 12 | F | 19-38 | | 5639 | 2099 | 0 | 0 | 5485 | 1939 | 0 | 0 | 1 | 1 | 1 | 1 | 0 | 0 | 1 | 0 | 0 | 0 | 1 | 0 | 0 | |
| 13 | F | 19-38 | | 4308 | 1222 | 0 | 0 | #N/A | #N/A | #N/A | #N/A | 1 | 1 | 1 | 0 | 0 | 0 | 0 | 0 | 0 | 0 | 1 | 0 | 0 | |
| 14 | F | 19-38 | | 1762 | 1341 | 0 | 0 | 1587 | 834 | 0 | 0 | 1 | 0 | 1 | 0 | 1 | 0 | 1 | 0 | 0 | 0 | 0 | 0 | 0 | |
| 15 | F | 39-82 | | 2340 | 1366 | 0 | 0 | 2265 | 1230 | 0 | 0 | 0 | 0 | 0 | 0 | 0 | 0 | 0 | 0 | 0 | 0 | 0 | 0 | 0 | |
| 16 | F | 19-38 | | 2534 | 768 | 0 | 0 | 2280 | 895 | 0 | 0 | 1 | 0 | 1 | 1 | 0 | 0 | 1 | 0 | 1 | 0 | 1 | 1 | 1 | 0 |
| 17 | F | 19-38 | | 2678 | 1279 | 0 | 0 | #N/A | #N/A | #N/A | #N/A | 1 | 0 | 1 | 1 | 1 | 1 | 1 | 0 | 0 | 0 | 1 | 1 | 1 | 0 |
| 18 | F | 19-38 | | 3124 | 1418 | 0 | 0 | #N/A | #N/A | #N/A | #N/A | 0 | 0 | 0 | 0 | 0 | 0 | 0 | 0 | 0 | 0 | 0 | 0 | 0 | |
| 19 | F | 19-38 | | 19275 | 13925 | 8 | 1 | #N/A | #N/A | #N/A | #N/A | 0 | 0 | 0 | 0 | 0 | 0 | 0 | 0 | 0 | 0 | 0 | 0 | 0 | |
| 20 | M | 39-82 | | 61536 | 26285 | 70 | 1 | 18885 | 15880 | 12 | 1 | 1 | 1 | 1 | 1 | 0 | 0 | 1 | 0 | 0 | 1 | 1 | 0 | 1 | 1 |
| 21 | f | 39-82 | | 3809 | 3197 | 0 | 0 | 2968 | 978 | 0 | 0 | 1 | 0 | 1 | 0 | 0 | 1 | 1 | 1 | 0 | 0 | 1 | 0 | 0 | |
| 22 | F | 39-82 | | 4161 | 787 | 0 | 0 | 3414 | 699 | 0 | 0 | 0 | 0 | 0 | 1 | 0 | 0 | 0 | 0 | 0 | 0 | 0 | 0 | 0 | |
| 23 | F | 39-82 | | 7182 | 2614 | 0 | 0 | 6889 | 2012 | 18 | 1 | 0 | 0 | 0 | 0 | 0 | 0 | 0 | 0 | 0 | 0 | 0 | 0 | 0 | |
| 24 | F | 39-82 | | 6357 | 973 | 0 | 0 | 2585 | 924 | 12 | 1 | 1 | 0 | 1 | 0 | 0 | 0 | 0 | 0 | 1 | 0 | 1 | 0 | 1 | 0 |
| 25 | F | 39-82 | | 5740 | 1430 | 0 | 0 | 2166 | 1557 | 0 | 0 | 1 | 0 | 1 | 0 | 0 | 0 | 0 | 0 | 0 | 0 | 0 | 0 | 0 | |
| 26 | F | 39-82 | | 2065 | 797 | 0 | 0 | 1898 | 1340 | 0 | 0 | 0 | 0 | 0 | 0 | 0 | 0 | 0 | 0 | 0 | 0 | 0 | 0 | 0 | |
| 27 | M | 19-38 | | 2379 | 1198 | 0 | 0 | #N/A | #N/A | #N/A | #N/A | 1 | 0 | 0 | 0 | 0 | 0 | 0 | 0 | 1 | 0 | 0 | 0 | 0 | |
| 28 | F | 19-38 | | 2642 | 2064 | 0 | 0 | 3523 | 3987 | 0 | 0 | 1 | 0 | 0 | 1 | 0 | 1 | 0 | 0 | 0 | 0 | 0 | 0 | 0 | |
| 29 | M | 19-38 | | 1442 | 592 | 0 | 0 | 1081 | 779 | 0 | 0 | 1 | 0 | 1 | 0 | 0 | 0 | 0 | 0 | 0 | 0 | 0 | 0 | 0 | |
| 30 | F | 39-82 | | 2727 | 723 | 0 | 0 | 1662 | 1250 | 0 | 0 | 1 | 0 | 1 | 1 | 1 | 0 | 0 | 0 | 0 | 0 | 1 | 0 | 1 | 0 |
| 31 | F | 39-82 | | 8958 | 1061 | 0 | 0 | 7118 | 818 | 0 | 0 | 1 | 1 | 1 | 1 | 0 | 0 | 0 | 0 | 1 | 0 | 0 | 0 | 0 | |
| 32 | F | 19-38 | | 50372 | 63596 | 89 | 1 | #N/A | #N/A | #N/A | #N/A | 1 | 0 | 1 | 1 | 1 | 0 | 1 | 0 | 1 | 0 | 1 | 0 | 1 | 1 |
| 33 | F | 39-82 | | 170080 | 181385 | 99 | 1 | 134053 | 258571 | 80 | 1 | 1 | 1 | 1 | 1 | 0 | 1 | 1 | 0 | 1 | 1 | 1 | 0 | 1 | 1 |
| 34 | F | 19-38 | | 1184 | 1625 | 0 | 0 | 735 | 1093 | 0 | 0 | 1 | 0 | 0 | 1 | 0 | 0 | 0 | 0 | 1 | 0 | 1 | 0 | 1 | 0 |
| 35 | F | 19-38 | | 1302 | 1679 | 0 | 0 | #N/A | #N/A | #N/A | #N/A | 1 | 0 | 1 | 0 | 1 | 0 | 0 | 0 | 1 | 0 | 1 | 0 | 0 | |
| 36 | F | 39-82 | | 101108 | 104405 | 96 | 1 | 91483 | 88102 | 98 | 1 | 1 | 0 | 1 | 1 | 1 | 1 | 1 | 1 | 1 | 1 | 1 | 0 | 0 | |
| 37 | F | 39-82 | | 1514 | 816 | 0 | 0 | 1119 | 1509 | 0 | 0 | 1 | 0 | 0 | 0 | 0 | 0 | 1 | 0 | 0 | 0 | 0 | 0 | 0 | |
| 38 | F | 19-38 | | 3639 | 1207 | 0 | 0 | 3331 | 537 | 0 | 0 | 1 | 1 | 1 | 1 | 0 | 0 | 1 | 0 | 1 | 0 | 0 | 0 | 1 | 0 |
| 39 | F | 39-82 | | 4293 | 757 | 0 | 0 | 2991 | 1926 | 33 | 1 | 0 | 0 | 0 | 0 | 0 | 0 | 0 | 0 | 0 | 0 | 0 | 0 | 0 | |
| 40 | F | 39-82 | | 2427 | 861 | 0 | 0 | 1043 | 1163 | 0 | 0 | 1 | 0 | 1 | 1 | 1 | 0 | 0 | 1 | 1 | 0 | 0 | 0 | 1 | 0 |
| 41 | M | 19-38 | | 1510 | 2228 | 0 | 0 | #N/A | #N/A | #N/A | #N/A | 0 | 0 | 0 | 0 | 0 | 0 | 0 | 0 | 0 | 0 | 0 | 0 | 0 | |
| 42 | M | 39-82 | | 136689 | 33141 | 15 | 1 | #N/A | #N/A | #N/A | #N/A | 1 | 0 | 1 | 1 | 1 | 1 | 0 | 0 | 0 | 1 | 1 | 1 | 1 | 1 |
| 43 | F | 39-82 | | 4467 | 1388 | 0 | 0 | 2499 | 750 | 0 | 0 | 1 | 1 | 1 | 0 | 1 | 1 | 1 | 0 | 0 | 0 | 1 | 0 | 0 | |
| 44 | M | 39-82 | negative | 2755 | 797 | 0 | 0 | #N/A | #N/A | #N/A | #N/A | 1 | 1 | 0 | 1 | 0 | 0 | 0 | 1 | 1 | 1 | 1 | 1 | 0 | |
| 45 | F | 19-38 | | 2211 | 1245 | 0 | 0 | 750 | 965 | 0 | 0 | 1 | 0 | 1 | 0 | 0 | 0 | 0 | 0 | 1 | 0 | 0 | 0 | 0 | |
| 46 | M | 19-38 | | 4720 | 682 | 0 | 0 | #N/A | #N/A | #N/A | #N/A | 1 | 0 | 1 | 1 | 1 | 1 | 0 | 0 | 1 | 0 | 0 | 0 | 0 | |
| 47 | F | 39-82 | negative | 6642 | 904 | 0 | 0 | 3097 | 621 | 0 | 0 | 1 | 0 | 1 | 0 | 0 | 0 | 1 | 0 | 0 | 1 | 1 | 0 | 0 | |
| 48 | F | 19-38 | | 23523 | 13803 | 63 | 1 | 7702 | 118340 | 30 | 1 | 1 | 1 | 1 | 0 | 1 | 0 | 1 | 1 | 1 | 1 | 1 | 0 | 0 | |
| 49 | F | 39-82 | | 3767 | 1598 | 0 | 0 | 1910 | 1157 | 0 | 0 | 0 | 0 | 0 | 0 | 0 | 0 | 0 | 0 | 0 | 0 | 0 | 0 | 0 | |
| 50 | M | 39-82 | | 317013 | 185033 | 97 | 1 | 217380 | 117537 | 98 | 1 | 1 | 1 | 1 | 1 | 0 | 0 | 0 | 0 | 0 | 0 | 0 | 1 | 0 | |
| 51 | M | 19-38 | | 61199 | 36432 | 60 | 1 | #N/A | #N/A | #N/A | #N/A | 1 | 0 | 1 | 1 | 1 | 1 | 1 | 0 | 1 | 1 | 1 | 1 | 1 | 1 |
| 52 | F | 19-38 | | 31057 | 55494 | 38 | 1 | 6482 | 43401 | 44 | 1 | 1 | 0 | 1 | 1 | 1 | 1 | 0 | 0 | 0 | 1 | 1 | 0 | 1 | 1 |
| 53 | F | 39-82 | | 1501 | 1468 | 0 | 0 | 501 | 380 | 0 | 0 | 0 | 0 | 0 | 0 | 0 | 0 | 0 | 0 | 0 | 0 | 0 | 0 | 0 | |
| 54 | F | 39-82 | | 4240 | 1047 | 0 | 0 | #N/A | #N/A | #N/A | #N/A | 1 | 0 | 0 | 0 | 0 | 0 | 0 | 0 | 0 | 0 | 0 | 1 | 0 | |
| 55 | F | 39-82 | | 2852 | 5273 | 0 | 0 | 1340 | 982 | 0 | 0 | 1 | 0 | 0 | 0 | 0 | 0 | 0 | 0 | 0 | 0 | 1 | 0 | 0 | |
| 56 | F | 19-38 | | 1670 | 705 | 0 | 0 | 1061 | 972 | 0 | 0 | 0 | 0 | 0 | 0 | 0 | 0 | 0 | 0 | 0 | 0 | 0 | 0 | 0 | |
| 57 | F | 39-82 | | 2207 | 509 | 0 | 0 | 1047 | 971 | 0 | 0 | 0 | 0 | 0 | 0 | 0 | 0 | 0 | 0 | 0 | 0 | 0 | 0 | 0 | |
| 58 | M | 19-38 | | 109629 | 24644 | 98 | 1 | 65895 | 39420 | 96 | 1 | 1 | 0 | 1 | 1 | 0 | 0 | 0 | 0 | 0 | 0 | 1 | 0 | 0 | |
| 59 | M | 19-38 | | 5750 | 1215 | 0 | 0 | 4846 | 2345 | 0 | 0 | 1 | 0 | 1 | 0 | 0 | 0 | 1 | 0 | 0 | 0 | 0 | 0 | 0 | |
| 60 | F | 39-82 | | 2944 | 2498 | 0 | 0 | #N/A | #N/A | #N/A | #N/A | 1 | 0 | 1 | 1 | 0 | 0 | 0 | 0 | 0 | 0 | 1 | 0 | 0 | |
| 61 | M | 19-38 | | 1896 | 1111 | 0 | 0 | #N/A | #N/A | #N/A | #N/A | 1 | 0 | 1 | 0 | 0 | 0 | 0 | 0 | 1 | 0 | 1 | 1 | 0 | |
| 62 | F | 39-82 | | 178273 | 62571 | 91 | 1 | 132619 | 47834 | 70 | 1 | 1 | 1 | 1 | 0 | 1 | 0 | 1 | 0 | 1 | 1 | 1 | 1 | 1 | 0 |
| 64 | F | 19-38 | | 1593 | 2863 | 0 | 0 | #N/A | #N/A | #N/A | #N/A | 0 | 0 | 0 | 0 | 0 | 0 | 0 | 0 | 0 | 0 | 0 | 0 | 0 | |
| 65 | F | 19-38 | | 51270 | 46944 | 81 | 1 | 17512 | 60305 | 89 | 1 | 1 | 1 | 1 | 1 | 1 | 1 | 1 | 0 | 0 | 1 | 1 | 0 | 1 | 1 |
| 66 | F | 19-38 | | 295380 | 152571 | 98 | 1 | 178625 | 147803 | 99 | 1 | 1 | 1 | 1 | 1 | 1 | 0 | 1 | 0 | 0 | 0 | 1 | 1 | 1 | 0 |
| 67 | F | 19-38 | | 3803 | 503 | 0 | 0 | #N/A | #N/A | #N/A | #N/A | 1 | 0 | 1 | 1 | 1 | 1 | 1 | 0 | 1 | 0 | 1 | 1 | 1 | |
| 68 | F | 39-82 | | 104926 | 46725 | 97 | 1 | #N/A | #N/A | #N/A | #N/A | 1 | 0 | 1 | 0 | 0 | 0 | 1 | 0 | 0 | 0 | 0 | 0 | 0 | |
| 69 | M | 39-82 | | 3538 | 1835 | 0 | 0 | 1969 | 687 | 0 | 0 | 1 | 1 | 1 | 1 | 0 | 0 | 0 | 0 | 0 | 0 | 1 | 0 | 0 | |

| | | | | | | | | | | | | | | | | | | | | | | | | | |
|---|---|---|---|---|---|---|---|---|---|---|---|---|---|---|---|---|---|---|---|---|---|---|---|---|---|
| 70 | M | 39-82 | | 895 | 10155 | 0 | 1 | #N/A | #N/A | #N/A | #N/A | 0 | 0 | 0 | 0 | 0 | 0 | 0 | 0 | 0 | 0 | 0 | 0 | 0 | |
| 71 | F | 39-82 | | 4351 | 940 | 0 | 0 | 1649 | 1371 | 0 | 0 | 1 | 0 | 0 | 0 | 0 | 0 | 0 | 0 | 1 | 0 | 1 | 0 | 0 | |
| 72 | F | 39-82 | | 145189 | 302202 | 99 | 1 | 107169 | 227738 | 92 | 1 | 1 | 1 | 1 | 1 | 1 | 0 | 1 | 0 | 1 | 1 | 1 | 0 | 0 | |
| 73 | F | 39-82 | | 202705 | 48714 | 87 | 1 | 124246 | 78810 | 70 | 1 | 1 | 0 | 1 | 0 | 0 | 0 | 0 | 1 | 0 | 0 | 1 | 0 | 0 | |
| 74 | F | 39-82 | | 4394 | 1233 | 0 | 0 | #N/A | #N/A | #N/A | #N/A | 0 | 0 | 0 | 0 | 0 | 0 | 0 | 0 | 0 | 0 | 0 | 0 | 0 | |
| 75 | F | 19-38 | | 7517 | 675 | 0 | 0 | 4335 | 2462 | 0 | 0 | 0 | 0 | 0 | 0 | 0 | 0 | 0 | 0 | 0 | 0 | 0 | 0 | 0 | |
| 76 | F | 39-82 | | 8815 | 994 | 0 | 0 | 8184 | 947 | 0 | 0 | 0 | 0 | 0 | 0 | 0 | 0 | 0 | 0 | 0 | 0 | 0 | 0 | 0 | |
| 77 | M | 39-82 | | 1359 | 7414 | 0 | 0 | 1748 | 7767 | 0 | 0 | 0 | 0 | 0 | 0 | 0 | 0 | 0 | 0 | 0 | 0 | 0 | 0 | 0 | |
| 78 | F | 39-82 | | 1712 | 673 | 0 | 0 | 1126 | 884 | 1 | 1 | 0 | 0 | 0 | 0 | 0 | 0 | 0 | 0 | 0 | 0 | 0 | 0 | 0 | |
| 79 | F | 39-82 | | 2220 | 966 | 0 | 0 | 1680 | 2309 | 0 | 0 | 1 | 1 | 1 | 1 | 0 | 0 | 1 | 0 | 0 | 0 | 0 | 1 | 1 | 0 |
| 81 | F | 19-38 | | 1493 | 760 | 0 | 0 | 1120 | 874 | 0 | 0 | 1 | 0 | 1 | 0 | 0 | 0 | 0 | 0 | 0 | 0 | 1 | 0 | 0 | |
| 82 | F | 39-82 | | 4333 | 1341 | 0 | 0 | 1835 | 1674 | 0 | 0 | 1 | 0 | 0 | 0 | 0 | 0 | 0 | 1 | 0 | 0 | 0 | 0 | 0 | |
| 83 | F | 19-38 | | 3464 | 1867 | 0 | 0 | 2937 | 2876 | 0 | 0 | 1 | 0 | 0 | 1 | 0 | 0 | 0 | 0 | 1 | 0 | 0 | 0 | 0 | |
| 84 | F | 39-82 | | 2946 | 1203 | 0 | 0 | 1307 | 1436 | 0 | 0 | 0 | 0 | 0 | 0 | 0 | 0 | 0 | 0 | 0 | 0 | 0 | 0 | 0 | |
| 85 | F | 39-82 | | 5168 | 1264 | 0 | 0 | 1317 | 1058 | 0 | 0 | 0 | 0 | 0 | 0 | 0 | 0 | 0 | 0 | 0 | 0 | 0 | 0 | 0 | |
| 86 | F | 19-38 | | 304 | 3895 | 0 | 0 | 1196 | 3552 | 0 | 0 | 1 | 0 | 1 | 0 | 0 | 0 | 0 | 0 | 1 | 0 | 1 | 0 | 0 | |
| 87 | F | 19-38 | | 3352 | 1319 | 0 | 0 | #N/A | #N/A | #N/A | #N/A | 1 | 0 | 1 | 0 | 0 | 0 | 0 | 0 | 0 | 0 | 1 | 1 | 0 | |
| 88 | F | 19-38 | | 2452 | 640 | 0 | 0 | #N/A | #N/A | #N/A | #N/A | 0 | 0 | 0 | 0 | 0 | 0 | 0 | 0 | 0 | 0 | 0 | 0 | 0 | |
| 89 | F | 19-38 | | 2499 | 590 | 0 | 0 | 2867 | 570 | 0 | 0 | 1 | 0 | 1 | 0 | 0 | 0 | 0 | 0 | 0 | 0 | 1 | 0 | 0 | |
| 90 | F | 39-82 | | 2351 | 865 | 0 | 0 | 1810 | 648 | 0 | 0 | 1 | 0 | 0 | 1 | 0 | 0 | 1 | 0 | 0 | 0 | 1 | 0 | 0 | |
| 91 | F | 19-38 | | 2405 | 888 | 0 | 0 | 1488 | 814 | 0 | 0 | 1 | 0 | 1 | 0 | 0 | 0 | 0 | 0 | 0 | 0 | 1 | 0 | 0 | |
| 92 | F | 19-38 | | 2394 | 1145 | 0 | 0 | #N/A | #N/A | #N/A | #N/A | 1 | 0 | 1 | 0 | 0 | 0 | 0 | 0 | 0 | 0 | 1 | 1 | 0 | |
| 93 | F | 19-38 | | 3596 | 1732 | 0 | 0 | 2987 | 1092 | 0 | 0 | 1 | 0 | 1 | 1 | 0 | 0 | 0 | 0 | 0 | 0 | 1 | 0 | 0 | |
| 94 | F | 39-82 | | 6554 | 1354 | 0 | 0 | 5498 | 1110 | 0 | 0 | 0 | 0 | 0 | 0 | 0 | 0 | 0 | 0 | 0 | 0 | 0 | 0 | 0 | |
| 95 | F | 19-38 | | 76705 | 132747 | 97 | 1 | #N/A | #N/A | #N/A | #N/A | 1 | 0 | 0 | 0 | 0 | 0 | 0 | 0 | 1 | 1 | 1 | 0 | 1 | 1 |
| 96 | F | 19-38 | | 3355 | 2031 | 0 | 0 | 2505 | 4110 | 0 | 0 | 1 | 0 | 1 | 0 | 0 | 0 | 0 | 0 | 0 | 0 | 1 | 1 | 0 | |
| 97 | M | 19-38 | | 4688 | 994 | 0 | 0 | #N/A | #N/A | #N/A | #N/A | 0 | 0 | 0 | 0 | 0 | 0 | 0 | 0 | 0 | 0 | 0 | 0 | 0 | |
| 98 | M | 19-38 | | 2295 | 879 | 0 | 0 | #N/A | #N/A | #N/A | #N/A | 0 | 0 | 0 | 0 | 0 | 0 | 0 | 0 | 0 | 0 | 0 | 0 | 0 | |
| 99 | F | 39-82 | | 4986 | 1852 | 0 | 0 | 2233 | 1721 | 0 | 0 | 1 | 0 | 0 | 1 | 0 | 0 | 0 | 0 | 1 | 0 | 0 | 0 | 0 | |
| 100 | F | 39-82 | | 116195 | 307158 | 98 | 1 | 55293 | 220004 | 98 | 1 | 0 | 0 | 0 | 0 | 0 | 0 | 0 | 0 | 0 | 0 | 0 | 0 | 0 | |
| 101 | F | 19-38 | | 2076 | 4168 | 0 | 0 | 1978 | 2069 | 0 | 0 | 1 | 0 | 1 | 0 | 0 | 0 | 1 | 0 | 0 | 0 | 1 | 0 | 0 | |
| 102 | F | 39-82 | | 4315 | 1102 | 0 | 0 | 2720 | 1310 | 0 | 0 | 0 | 0 | 0 | 0 | 0 | 0 | 0 | 0 | 0 | 0 | 0 | 0 | 0 | |
| 103 | F | 39-82 | | 2572 | 1156 | 0 | 0 | 1460 | 1595 | 0 | 0 | 1 | 0 | 0 | 0 | 0 | 0 | 0 | 0 | 0 | 0 | 0 | 1 | 0 | |
| 104 | M | 39-82 | | 3949 | 717 | 0 | 0 | 2103 | 986 | 0 | 0 | 0 | 0 | 0 | 0 | 0 | 0 | 0 | 0 | 0 | 0 | 0 | 0 | 1 | 0 |
| 105 | F | 39-82 | | 5139 | 725 | 0 | 0 | 2589 | 1364 | 0 | 0 | 0 | 0 | 0 | 0 | 0 | 0 | 0 | 0 | 0 | 0 | 0 | 0 | 0 | |
| 106 | F | 19-38 | | 3200 | 2322 | 0 | 0 | 2218 | 3986 | 0 | 0 | 0 | 0 | 0 | 0 | 0 | 0 | 0 | 0 | 0 | 0 | 0 | 0 | 0 | |
| 107 | M | 19-38 | | 2742 | 970 | 0 | 0 | #N/A | #N/A | #N/A | #N/A | 0 | 0 | 0 | 0 | 0 | 0 | 0 | 0 | 0 | 0 | 0 | 0 | 0 | |
| 108 | M | 39-82 | | 2606 | 1558 | 0 | 0 | 2222 | 3138 | 0 | 0 | 1 | 0 | 0 | 0 | 0 | 0 | 0 | 1 | 1 | 0 | 0 | 0 | 0 | |
| 109 | F | 19-38 | | 2943 | 1210 | 0 | 0 | 1499 | 845 | 0 | 0 | 0 | 0 | 0 | 0 | 0 | 0 | 0 | 0 | 0 | 0 | 0 | 0 | 0 | |
| 110 | F | 39-82 | | 2436 | 643 | 0 | 0 | 847 | 655 | 0 | 0 | 1 | 1 | 0 | 0 | 0 | 0 | 0 | 0 | 1 | 0 | 1 | 0 | 1 | 0 |
| 111 | M | 39-82 | | 6165 | 1652 | 0 | 0 | 1548 | 1290 | 0 | 0 | 1 | 0 | 1 | 0 | 0 | 0 | 0 | 0 | 0 | 0 | 1 | 0 | 0 | |
| 112 | M | 39-82 | | 7503 | 1677 | 0 | 0 | 3868 | 1159 | 0 | 0 | 0 | 0 | 0 | 0 | 0 | 0 | 0 | 0 | 0 | 0 | 0 | 0 | 0 | |
| 113 | F | 39-82 | | 4893 | 1215 | 0 | 0 | 1509 | 698 | 0 | 0 | 0 | 0 | 0 | 0 | 0 | 0 | 0 | 0 | 0 | 0 | 0 | 0 | 0 | |
| 114 | F | 39-82 | | 1617 | 3874 | 0 | 0 | #N/A | #N/A | #N/A | #N/A | 0 | 0 | 0 | 0 | 0 | 0 | 0 | 0 | 0 | 0 | 0 | 0 | 0 | |
| 115 | F | 19-38 | | 1904 | 1947 | 0 | 0 | #N/A | #N/A | #N/A | #N/A | 1 | 0 | 1 | 1 | 1 | 0 | 1 | 0 | 0 | 0 | 1 | 0 | 0 | |
| 116 | M | 19-38 | | 2281 | 1036 | 0 | 0 | #N/A | #N/A | #N/A | #N/A | 0 | 0 | 0 | 0 | 0 | 0 | 0 | 0 | 0 | 0 | 0 | 0 | 0 | |
| 117 | M | 39-82 | | 3081 | 1030 | 0 | 0 | 1193 | 559 | 0 | 0 | 0 | 0 | 0 | 0 | 0 | 0 | 0 | 0 | 0 | 0 | 0 | 0 | 0 | |
| 118 | M | 19-38 | | 34117 | 15364 | 63 | 1 | 10942 | 10845 | 3 | 1 | 1 | 0 | 1 | 0 | 0 | 0 | 1 | 0 | 0 | 1 | 0 | 0 | 1 | 1 |
| 119 | F | 39-82 | | 5514 | 2127 | 0 | 0 | 486 | 318 | 0 | 0 | 1 | 0 | 1 | 0 | 1 | 0 | 1 | 0 | 0 | 0 | 1 | 0 | 0 | |
| 120 | F | 39-82 | | 4806 | 1497 | 0 | 0 | 2335 | 534 | 0 | 0 | 1 | 1 | 1 | 0 | 1 | 0 | 1 | 0 | 1 | 0 | 1 | 1 | 0 | |
| 121 | F | 19-38 | | 6579 | 994 | 0 | 0 | 5567 | 808 | 0 | 0 | 1 | 1 | 1 | 1 | 1 | 1 | 1 | 0 | 1 | 0 | 1 | 1 | 0 | |
| 122 | F | 19-38 | | 972 | 21734 | 0 | 1 | 1056 | 15416 | 0 | 1 | 1 | 0 | 1 | 0 | 0 | 0 | 0 | 0 | 0 | 0 | 0 | 0 | 0 | |
| 123 | F | 39-82 | | 6013 | 1012 | 0 | 0 | 1278 | 763 | 0 | 0 | 1 | 0 | 1 | 1 | 1 | 1 | 1 | 0 | 1 | 0 | 1 | 0 | 0 | |
| 124 | F | 39-82 | | 6216 | 2059 | 0 | 0 | 2621 | 2066 | 0 | 0 | 0 | 0 | 0 | 0 | 0 | 0 | 0 | 0 | 0 | 0 | 0 | 0 | 0 | |
| 125 | F | 19-38 | | 4954 | 16268 | 0 | 1 | 1883 | 916 | 0 | 0 | 1 | 0 | 1 | 0 | 0 | 0 | 0 | 0 | 0 | 0 | 0 | 0 | 0 | |
| 126 | F | 19-38 | | 2491 | 474 | 0 | 0 | 452 | 910 | 0 | 0 | 1 | 0 | 0 | 0 | 1 | 1 | 0 | 0 | 0 | 0 | 1 | 0 | 0 | |
| 127 | M | 39-82 | | 9046 | 1053 | 0 | 0 | 9921 | 713 | 0 | 0 | 0 | 0 | 0 | 0 | 0 | 0 | 0 | 0 | 0 | 0 | 0 | 0 | 0 | |
| 128 | M | 39-82 | | 20965 | 18625 | 0 | 1 | 9351 | 15635 | 0 | 1 | 1 | 1 | 1 | 1 | 0 | 0 | 0 | 0 | 0 | 0 | 0 | 0 | 0 | |
| 129 | F | 19-38 | | 119231 | 39127 | 84 | 1 | 84633 | 27221 | 35 | 1 | 1 | 0 | 1 | 0 | 1 | 1 | 0 | 0 | 1 | 1 | 1 | 0 | 1 | 1 |
| 130 | M | 19-38 | | 11640 | 736 | 0 | 1 | 8854 | 776 | 0 | 0 | 1 | 0 | 1 | 1 | 0 | 0 | 0 | 0 | 0 | 0 | 1 | 0 | 0 | |
| 131 | F | 39-82 | | 3844 | 3280 | 0 | 0 | 822 | 924 | 0 | 0 | 1 | 0 | 0 | 0 | 0 | 0 | 0 | 0 | 1 | 0 | 0 | 1 | 1 | 0 |
| 132 | F | 19-38 | | 4177 | 1763 | 0 | 0 | #N/A | #N/A | #N/A | #N/A | 0 | 0 | 0 | 0 | 0 | 0 | 0 | 0 | 0 | 0 | 0 | 0 | 0 | |
| 133 | M | 39-82 | | 150622 | 194323 | 96 | 1 | 111040 | 159327 | 0 | 1 | 1 | 0 | 1 | 1 | 1 | 0 | 1 | 1 | 0 | 1 | 1 | 0 | 0 | |
| 134 | F | 39-82 | | 186108 | 115400 | 98 | 1 | 23759 | 8616 | 84 | 1 | 1 | 1 | 1 | 0 | 0 | 0 | 0 | 0 | 1 | 1 | 1 | 0 | 0 | |
| 135 | F | 39-82 | negative | 3264 | 811 | 0 | 0 | 2467 | 557 | 0 | 0 | 1 | 0 | 1 | 0 | 0 | 0 | 1 | 0 | 0 | 1 | 1 | 0 | 0 | |
| 136 | F | 39-82 | | 4340 | 1152 | 0 | 0 | 1092 | 1298 | 0 | 0 | 1 | 1 | 1 | 1 | 1 | 1 | 1 | 0 | 1 | 0 | 1 | 0 | 0 | |
| 137 | F | 39-82 | | 4730 | 1480 | 0 | 0 | 2649 | 1533 | 0 | 0 | 1 | 0 | 0 | 0 | 0 | 0 | 0 | 0 | 0 | 0 | 1 | 1 | 1 | 0 |
| 138 | M | 19-38 | | 9310 | 1266 | 0 | 0 | 8562 | 1017 | 0 | 0 | 0 | 0 | 0 | 0 | 0 | 0 | 0 | 0 | 0 | 0 | 0 | 0 | 0 | |
| 139 | F | 39-82 | | 2989 | 1513 | 0 | 0 | 2013 | 1307 | 0 | 0 | 1 | 1 | 0 | 0 | 0 | 0 | 1 | 0 | 0 | 0 | 1 | 0 | 0 | |
| 140 | M | 39-82 | negative | 2298 | 1114 | 0 | 0 | 5691 | 780 | 0 | 0 | 1 | 1 | 0 | 1 | 0 | 0 | 0 | 0 | 1 | 0 | 1 | 0 | 1 | 1 |
| 141 | F | 19-38 | | 2114 | 944 | 0 | 0 | #N/A | #N/A | #N/A | #N/A | 1 | 0 | 0 | 0 | 0 | 0 | 0 | 0 | 1 | 0 | 1 | 0 | 0 | |
| 142 | F | 19-38 | | 2569 | 936 | 0 | 0 | 3086 | 1832 | 0 | 0 | 1 | 0 | 1 | 1 | 1 | 0 | 0 | 0 | 0 | 0 | 1 | 0 | 0 | |
| 143 | F | 39-82 | | 2556 | 1093 | 0 | 0 | 1048 | 1924 | 0 | 0 | 0 | 0 | 0 | 0 | 0 | 0 | 0 | 0 | 0 | 0 | 0 | 0 | 0 | |
| 144 | F | 19-38 | | 3054 | 1756 | 0 | 0 | 2894 | 4108 | 0 | 0 | 1 | 0 | 0 | 0 | 0 | 0 | 0 | 0 | 1 | 0 | 1 | 0 | 0 | |
| 145 | F | 39-82 | | 59246 | 72100 | 96 | 1 | #N/A | #N/A | #N/A | #N/A | 1 | 0 | 1 | 0 | 0 | 0 | 1 | 0 | 1 | 1 | 1 | 0 | 1 | 1 |
| 146 | F | 39-82 | | 23447 | 56210 | 91 | 1 | 8322 | 32717 | 53 | 1 | 1 | 0 | 1 | 0 | 0 | 0 | 0 | 0 | 1 | 1 | 1 | 0 | 1 | 1 |
| 147 | F | 19-38 | negative | 1856 | 681 | 0 | 0 | 1201 | 1369 | 0 | 0 | 1 | 0 | 1 | 1 | 1 | 1 | 1 | 0 | 0 | 1 | 1 | 0 | 0 | |

| | | | | | | | | | | | | | | | | | | | | | | | | | |
|---|---|---|---|---|---|---|---|---|---|---|---|---|---|---|---|---|---|---|---|---|---|---|---|---|---|
| 148 | M | 39-82 | | 8053 | 575 | 0 | 0 | #N/A | #N/A | #N/A | #N/A | 0 | 0 | 0 | 0 | 0 | 0 | 0 | 0 | 0 | 0 | 0 | 0 | 1 | 0 |
| 149 | F | 39-82 | | 1972 | 1128 | 0 | 0 | 2947 | 1253 | 0 | 0 | 0 | 0 | 0 | 0 | 0 | 0 | 0 | 0 | 0 | 0 | 0 | 0 | 0 | |
| 150 | F | 19-38 | | 38295 | 60640 | 74 | 1 | 13061 | 44240 | 5 | 1 | 1 | 0 | 1 | 1 | 1 | 1 | 1 | 0 | 0 | 1 | 1 | 1 | 1 | 1 |
| 151 | F | 19-38 | | 3773 | 1592 | 0 | 0 | #N/A | #N/A | #N/A | #N/A | 1 | 1 | 1 | 0 | 0 | 0 | 1 | 0 | 1 | 0 | 1 | 1 | 0 | |
| 152 | F | 19-38 | | 2212 | 3480 | 0 | 0 | 983 | 2387 | 0 | 0 | 1 | 1 | 1 | 1 | 1 | 1 | 1 | 0 | 1 | 0 | 1 | 1 | 0 | |
| 153 | F | 39-82 | | 5155 | 1381 | 0 | 0 | 599 | 398 | 0 | 0 | 1 | 0 | 1 | 1 | 1 | 1 | 1 | 0 | 1 | 0 | 1 | 0 | 0 | |
| 154 | F | 19-38 | | 2236 | 460 | 0 | 0 | 624 | 240 | 0 | 0 | 1 | 0 | 1 | 0 | 0 | 0 | 1 | 0 | 0 | 0 | 1 | 0 | 0 | |
| 155 | F | 19-38 | | 3091 | 2805 | 0 | 0 | #N/A | #N/A | #N/A | #N/A | 0 | 0 | 0 | 0 | 0 | 0 | 0 | 0 | 0 | 0 | 0 | 0 | 0 | |
| 156 | F | 39-82 | negative | 2931 | 1369 | 0 | 0 | #N/A | #N/A | #N/A | #N/A | 1 | 1 | 0 | 1 | 1 | 1 | 0 | 0 | 0 | 1 | 1 | 1 | 0 | |
| 157 | F | 19-38 | negative | 5056 | 3210 | 0 | 0 | #N/A | #N/A | #N/A | #N/A | 1 | 0 | 1 | 0 | 0 | 0 | 1 | 0 | 1 | 1 | 1 | 0 | 1 | 1 |
| 158 | F | 39-82 | | 1511 | 1626 | 0 | 0 | #N/A | #N/A | #N/A | #N/A | 0 | 0 | 0 | 0 | 0 | 0 | 0 | 0 | 0 | 0 | 0 | 0 | 0 | |
| 159 | F | 39-82 | | 2945 | 1432 | 0 | 0 | #N/A | #N/A | #N/A | #N/A | 1 | 0 | 1 | 1 | 0 | 0 | 0 | 0 | 0 | 0 | 1 | 0 | 1 | 0 |
| 160 | F | 19-38 | | 18151 | 32654 | 20 | 1 | 8812 | 23879 | 17 | 1 | 1 | 1 | 1 | 1 | 1 | 1 | 1 | 0 | 0 | 1 | 1 | 1 | 1 | 1 |
| 161 | M | 19-38 | | 10959 | 13001 | 0 | 1 | 8229 | 136377 | 29 | 1 | 1 | 1 | 1 | 1 | 0 | 0 | 1 | 0 | 0 | 1 | 1 | 1 | 1 | 1 |
| 162 | F | 19-38 | | 22031 | 53264 | 81 | 1 | 7054 | 48740 | 0 | 1 | 1 | 1 | 1 | 1 | 0 | 0 | 0 | 0 | 0 | 1 | 1 | 0 | 0 | |
| 163 | F | 39-82 | | 2339 | 1577 | 0 | 0 | #N/A | #N/A | #N/A | #N/A | 1 | 0 | 1 | 0 | 0 | 0 | 1 | 0 | 0 | 0 | 1 | 0 | 0 | |
| 164 | F | 19-38 | | 67161 | 59137 | 53 | 1 | #N/A | #N/A | #N/A | #N/A | 1 | 0 | 0 | 0 | 0 | 0 | 0 | 0 | 0 | 1 | 0 | 0 | 0 | |
| 165 | F | 19-38 | | 4832 | 1195 | 0 | 0 | #N/A | #N/A | #N/A | #N/A | 1 | 1 | 1 | 0 | 0 | 0 | 1 | 1 | 1 | 0 | 1 | 0 | 0 | |
| 166 | F | 39-82 | | 59038 | 84359 | 72 | 1 | 17424 | 62956 | 78 | 1 | 1 | 1 | 1 | 0 | 1 | 0 | 1 | 0 | 1 | 1 | 0 | 0 | 0 | |
| 167 | F | 19-38 | | 1428 | 1292 | 0 | 0 | #N/A | #N/A | #N/A | #N/A | 1 | 0 | 1 | 0 | 1 | 1 | 0 | 0 | 0 | 0 | 1 | 0 | 0 | |
| 168 | F | 39-82 | | 263693 | 44441 | 29 | 1 | 148210 | 24410 | 0 | 1 | 1 | 1 | 1 | 1 | 0 | 0 | 1 | 0 | 1 | 1 | 1 | 0 | 0 | |
| 169 | F | 39-82 | | 143219 | 34492 | 43 | 1 | #N/A | #N/A | #N/A | #N/A | 1 | 1 | 1 | 1 | 0 | 0 | 1 | 0 | 0 | 1 | 1 | 1 | 0 | |
| 170 | F | 19-38 | | 1431 | 1028 | 0 | 0 | 1396 | 1027 | 0 | 0 | 1 | 0 | 1 | 0 | 0 | 0 | 1 | 0 | 1 | 0 | 1 | 0 | 0 | |
| 171 | F | 19-38 | | 134772 | 57653 | 71 | 1 | #N/A | #N/A | #N/A | #N/A | 1 | 1 | 1 | 1 | 1 | 1 | 1 | 0 | 0 | 1 | 1 | 0 | 1 | 1 |
| 173 | F | 19-38 | | 42325 | 45388 | 48 | 1 | 21224 | 23575 | 39 | 1 | 1 | 0 | 0 | 0 | 1 | 0 | 1 | 0 | 1 | 1 | 1 | 0 | 1 | |
| 174 | F | 39-82 | | 106782 | 56216 | 86 | 1 | 20416 | 54260 | 97 | 1 | 1 | 1 | 1 | 1 | 1 | 1 | 1 | 0 | 1 | 1 | 1 | 1 | 0 | |
| 175 | F | 19-38 | IgM positive | 16282 | 20943 | 37 | 1 | 6282 | 32131 | 0 | 1 | 1 | 1 | 1 | 0 | 1 | 0 | 1 | 0 | 0 | 1 | 1 | 0 | 0 | |
| 176 | F | 39-82 | | 1927 | 1332 | 0 | 0 | 1493 | 1371 | 0 | 0 | 1 | 0 | 1 | 0 | 0 | 0 | 1 | 1 | 0 | 0 | 1 | 1 | 0 | |
| 177 | F | 19-38 | | 3198 | 1021 | 0 | 0 | 2051 | 1137 | 0 | 0 | 0 | 0 | 0 | 0 | 0 | 0 | 0 | 0 | 0 | 0 | 0 | 0 | 0 | |
| 178 | F | 19-38 | | 5380 | 1230 | 0 | 0 | 1015 | 1033 | 0 | 0 | 1 | 1 | 1 | 1 | 0 | 0 | 0 | 0 | 1 | 0 | 1 | 0 | 0 | |
| 179 | F | 19-38 | | 7853 | 989 | 0 | 0 | 3511 | 565 | 0 | 0 | 1 | 0 | 1 | 0 | 0 | 0 | 1 | 0 | 0 | 0 | 1 | 0 | 0 | |
| 180 | F | 39-82 | | 6589 | 4699 | 0 | 0 | 4675 | 3568 | 0 | 0 | 1 | 0 | 1 | 0 | 0 | 0 | 0 | 0 | 0 | 0 | 1 | 0 | 0 | |
| 181 | F | 39-82 | | 9207 | 1518 | 0 | 0 | 3941 | 1679 | 0 | 0 | 1 | 0 | 1 | 0 | 0 | 0 | 1 | 1 | 1 | 0 | 1 | 1 | 0 | |
| 182 | F | 19-38 | | 3841 | 1906 | 0 | 0 | 320 | 872 | 0 | 0 | 1 | 0 | 0 | 0 | 0 | 0 | 1 | 0 | 1 | 0 | 1 | 0 | 0 | |
| 183 | F | 39-82 | | 4177 | 4325 | 0 | 0 | 2057 | 2843 | 0 | 0 | 0 | 0 | 0 | 0 | 0 | 0 | 0 | 0 | 0 | 0 | 0 | 0 | 0 | |
| 184 | F | 19-38 | | 2341 | 1229 | 0 | 0 | 2895 | 3450 | 0 | 0 | 1 | 0 | 1 | 1 | 1 | 0 | 0 | 0 | 0 | 0 | 1 | 0 | 0 | |
| 185 | F | 19-38 | | 3098 | 1352 | 0 | 0 | 1861 | 1412 | 0 | 0 | 0 | 0 | 0 | 0 | 0 | 0 | 0 | 0 | 0 | 0 | 0 | 0 | 0 | |
| 186 | F | 39-82 | | 3751 | 1821 | 0 | 0 | 2341 | 2086 | 0 | 0 | 1 | 0 | 1 | 1 | 1 | 0 | 1 | 1 | 1 | 0 | 1 | 0 | 0 | |
| 187 | F | 39-82 | | 104082 | 152905 | 87 | 1 | 79907 | 107516 | 0 | 1 | 0 | 0 | 0 | 0 | 0 | 0 | 0 | 0 | 0 | 0 | 0 | 0 | 0 | |
| 188 | F | 39-82 | | 4801 | 1390 | 0 | 0 | 1783 | 1055 | 0 | 0 | 0 | 0 | 0 | 0 | 0 | 0 | 0 | 0 | 0 | 0 | 0 | 0 | 0 | |
| 189 | F | 39-82 | | 116668 | 210097 | 95 | 1 | 101509 | 160225 | 90 | 1 | 1 | 1 | 1 | 1 | 0 | 1 | 1 | 1 | 1 | 1 | 0 | 1 | 1 | 1 |
| 190 | F | 39-82 | | 2122 | 591 | 0 | 0 | #N/A | #N/A | #N/A | #N/A | 0 | 0 | 0 | 0 | 0 | 0 | 0 | 0 | 0 | 0 | 0 | 0 | 0 | |
| 191 | F | 19-38 | | 1917 | 1490 | 0 | 0 | 1158 | 1688 | 0 | 0 | 1 | 0 | 0 | 0 | 0 | 0 | 0 | 0 | 0 | 0 | 1 | 0 | 1 | 0 |
| 192 | F | 39-82 | | 1712 | 1050 | 0 | 0 | 2812 | 1304 | 0 | 0 | 0 | 0 | 0 | 0 | 0 | 0 | 0 | 0 | 0 | 0 | 0 | 0 | 0 | |
| 193 | F | 39-82 | | 1920 | 1155 | 0 | 0 | 1083 | 760 | 0 | 0 | 0 | 0 | 0 | 0 | 0 | 0 | 0 | 0 | 0 | 0 | 0 | 0 | 0 | |
| 194 | F | 19-38 | | 71371 | 29653 | 67 | 1 | #N/A | #N/A | #N/A | #N/A | 1 | 0 | 1 | 1 | 0 | 0 | 0 | 0 | 1 | 0 | 1 | 0 | 0 | |
| 195 | M | 19-38 | | 2199 | 2085 | 0 | 0 | #N/A | #N/A | #N/A | #N/A | 1 | 1 | 1 | 1 | 0 | 0 | 1 | 0 | 1 | 0 | 1 | 1 | 0 | |
| 196 | F | 19-38 | | 772 | 1893 | 0 | 0 | #N/A | #N/A | #N/A | #N/A | 0 | 0 | 0 | 0 | 0 | 0 | 0 | 0 | 0 | 0 | 0 | 0 | 0 | |
| 197 | M | 39-82 | | 4577 | 1494 | 0 | 0 | 2273 | 2782 | 0 | 0 | 0 | 0 | 0 | 0 | 0 | 0 | 0 | 0 | 0 | 0 | 0 | 0 | 0 | |
| 198 | F | 19-38 | | 255125 | 51975 | 96 | 1 | 138798 | 30763 | 94 | 1 | 1 | 0 | 1 | 1 | 1 | 0 | 1 | 0 | 0 | 0 | 1 | 0 | 0 | |
| 199 | F | 19-38 | | 2119 | 5315 | 0 | 0 | #N/A | #N/A | #N/A | #N/A | 1 | 1 | 1 | 1 | 1 | 1 | 0 | 0 | 1 | 0 | 1 | 0 | 1 | 0 |
| 200 | M | 19-38 | | 211 | 576 | 0 | 0 | #N/A | #N/A | #N/A | #N/A | 1 | 1 | 1 | 0 | 0 | 0 | 0 | 0 | 1 | 0 | 1 | 1 | 0 | |
| 201 | F | 19-38 | | 137633 | 93690 | 89 | 1 | 29891 | 84517 | 73 | 1 | 1 | 1 | 1 | 1 | 0 | 0 | 1 | 0 | 1 | 1 | 1 | 0 | 0 | |
| 202 | F | 39-82 | | 3138 | 5077 | 0 | 0 | 3060 | 3890 | 0 | 0 | 1 | 0 | 1 | 0 | 0 | 0 | 0 | 0 | 0 | 0 | 0 | 0 | 0 | |
| 203 | F | 19-38 | | 2306 | 789 | 0 | 0 | 1170 | 955 | 0 | 0 | 1 | 0 | 1 | 1 | 1 | 1 | 0 | 0 | 0 | 0 | 1 | 1 | 1 | 0 |
| 204 | F | 19-38 | | 1765 | 1523 | 0 | 0 | #N/A | #N/A | #N/A | #N/A | 1 | 0 | 1 | 0 | 0 | 0 | 1 | 0 | 1 | 0 | 0 | 0 | 0 | |
| 205 | F | 39-82 | | 25474 | 21638 | 85 | 1 | 12465 | 19279 | 82 | 1 | 1 | 1 | 1 | 0 | 1 | 1 | 1 | 0 | 0 | 1 | 1 | 0 | 1 | 1 |
| 206 | F | 39-82 | | 7912 | 1925 | 0 | 0 | 1816 | 1915 | 0 | 0 | 1 | 0 | 1 | 0 | 0 | 0 | 0 | 0 | 0 | 0 | 1 | 0 | 0 | |
| 207 | M | 39-82 | | 5609 | 3546 | 0 | 0 | 5219 | 5580 | 0 | 0 | 1 | 1 | 0 | 1 | 0 | 0 | 0 | 0 | 0 | 0 | 0 | 0 | 0 | |
| 208 | M | 39-82 | | 8048 | 777 | 0 | 0 | 7970 | 1001 | 4 | 1 | 1 | 0 | 1 | 1 | 0 | 0 | 1 | 0 | 1 | 0 | 0 | 0 | 0 | |
| 209 | M | 39-82 | | 6990 | 690 | 0 | 0 | 6319 | 535 | 0 | 0 | 0 | 0 | 0 | 0 | 0 | 0 | 0 | 0 | 0 | 0 | 0 | 0 | 1 | 0 |
| 210 | F | 39-82 | IgM positive | 3883 | 4777 | 0 | 0 | 2624 | 2352 | 0 | 0 | 1 | 0 | 1 | 0 | 1 | 0 | 0 | 0 | 0 | 0 | 1 | 1 | 1 | 1 |
| 211 | F | 39-82 | | 3546 | 902 | 0 | 0 | 1715 | 667 | 0 | 0 | 1 | 1 | 1 | 1 | 0 | 0 | 1 | 1 | 1 | 0 | 1 | 1 | 0 | |
| 212 | F | 19-38 | | 3450 | 672 | 0 | 0 | #N/A | #N/A | #N/A | #N/A | 0 | 0 | 0 | 0 | 0 | 0 | 0 | 0 | 0 | 0 | 0 | 0 | 0 | |
| 213 | F | 19-38 | | 2752 | 2178 | 0 | 0 | 479 | 334 | 0 | 0 | 0 | 0 | 0 | 0 | 0 | 0 | 0 | 0 | 0 | 0 | 0 | 0 | 0 | |
| 214 | F | 39-82 | | 5289 | 1311 | 0 | 0 | 6081 | 1626 | 0 | 0 | 0 | 0 | 0 | 0 | 0 | 0 | 0 | 0 | 0 | 0 | 0 | 0 | 0 | |
| 215 | F | 39-82 | IgG traces | 9404 | 1779 | 0 | 0 | 5974 | 3320 | 0 | 0 | 1 | 1 | 1 | 1 | 1 | 1 | 1 | 0 | 0 | 1 | 0 | 0 | 0 | |
| 216 | F | 19-38 | | 1802 | 651 | 0 | 0 | #N/A | #N/A | #N/A | #N/A | 1 | 1 | 1 | 1 | 1 | 0 | 0 | 0 | 1 | 0 | 0 | 0 | 0 | |
| 218 | F | 19-38 | | 3410 | 1614 | 0 | 0 | 4176 | 2790 | 0 | 0 | 0 | 0 | 0 | 0 | 0 | 0 | 0 | 0 | 0 | 0 | 0 | 0 | 0 | |
| 220 | F | 39-82 | | 4927 | 2494 | 0 | 0 | 4989 | 3518 | 0 | 0 | 1 | 0 | 0 | 0 | 0 | 0 | 0 | 0 | 0 | 0 | 0 | 1 | 0 | |
| 221 | F | 19-38 | | 2823 | 1226 | 0 | 0 | 1001 | 2307 | 0 | 0 | 1 | 0 | 1 | 1 | 1 | 1 | 1 | 1 | 0 | 0 | 0 | 0 | 0 | |
| 222 | F | 19-38 | | 140418 | 109849 | 93 | 1 | 101211 | 75975 | 67 | 1 | 1 | 0 | 1 | 1 | 0 | 1 | 1 | 0 | 0 | 0 | 1 | 0 | 1 | 1 |
| 223 | F | 39-82 | | 6956 | 1271 | 0 | 0 | 8408 | 893 | 0 | 0 | 1 | 0 | 1 | 1 | 1 | 0 | 0 | 1 | 0 | 0 | 1 | 1 | 1 | 0 |
| 224 | F | 19-38 | negative | 4557 | 7407 | 0 | 0 | 2378 | 9080 | 0 | 1 | 1 | 1 | 1 | 1 | 0 | 0 | 1 | 0 | 0 | 1 | 1 | 1 | 0 | |
| 225 | F | 39-82 | | 4454 | 3857 | 0 | 0 | 2088 | 2380 | 0 | 0 | 0 | 0 | 0 | 0 | 0 | 0 | 0 | 0 | 0 | 0 | 0 | 0 | 0 | |
| 226 | F | 39-82 | | 4499 | 871 | 0 | 0 | 2853 | 1047 | 0 | 0 | 1 | 0 | 1 | 1 | 0 | 0 | 0 | 0 | 1 | 0 | 0 | 0 | 0 | |
| 227 | F | 39-82 | | 3447 | 3932 | 0 | 0 | 3102 | 3685 | 2 | 1 | 1 | 0 | 1 | 0 | 0 | 0 | 0 | 0 | 1 | 0 | 1 | 0 | 0 | |

| | | | | | | | | | | | | | | | | | | | | | | | | | |
|---|---|---|---|---|---|---|---|---|---|---|---|---|---|---|---|---|---|---|---|---|---|---|---|---|---|
| 228 | F | 39-82 | | 2155 | 584 | 0 | 0 | 1465 | 1213 | 0 | 0 | 0 | 0 | 0 | 0 | 0 | 0 | 0 | 0 | 0 | 0 | 0 | 0 | 0 | |
| 229 | F | 39-82 | | 83173 | 84697 | 90 | 1 | 31612 | 59584 | 81 | 1 | 1 | 0 | 1 | 1 | 0 | 0 | 1 | 0 | 1 | 1 | 1 | 0 | 1 | 1 |
| 230 | F | 19-38 | | 942 | 845 | 0 | 0 | 1912 | 827 | 0 | 0 | 1 | 1 | 1 | 1 | 0 | 1 | 1 | 0 | 1 | 0 | 1 | 0 | 0 | |
| 231 | F | 39-82 | | 4236 | 983 | 0 | 0 | 4906 | 2130 | 0 | 0 | 0 | 0 | 0 | 0 | 0 | 0 | 0 | 0 | 0 | 0 | 0 | 0 | 0 | |
| 232 | F | 39-82 | | 6936 | 2657 | 0 | 0 | 3968 | 1437 | 0 | 0 | 1 | 0 | 0 | 0 | 0 | 0 | 0 | 0 | 0 | 0 | 1 | 0 | 0 | |
| 233 | M | 39-82 | | 2501 | 4516 | 0 | 0 | 1504 | 5053 | 0 | 0 | 1 | 0 | 0 | 0 | 0 | 0 | 1 | 0 | 0 | 0 | 0 | 0 | 0 | |
| 234 | F | 39-82 | | 9906 | 1252 | 0 | 0 | 6731 | 959 | 0 | 0 | 1 | 0 | 1 | 0 | 0 | 0 | 0 | 0 | 0 | 0 | 0 | 0 | 0 | |
| 235 | F | 39-82 | | 3598 | 826 | 0 | 0 | 2618 | 778 | 0 | 0 | 0 | 0 | 0 | 0 | 0 | 0 | 0 | 0 | 0 | 0 | 0 | 0 | 0 | |
| 236 | M | 19-38 | | 1934 | 6058 | 0 | 0 | 1613 | 999 | 0 | 0 | 0 | 0 | 0 | 0 | 0 | 0 | 0 | 0 | 0 | 0 | 0 | 0 | 0 | |
| 237 | F | 39-82 | | 60298 | 95762 | 28 | 1 | #N/A | #N/A | #N/A | #N/A | 1 | 0 | 0 | 1 | 0 | 0 | 0 | 0 | 0 | 1 | 0 | 0 | 0 | |
| 238 | F | 19-38 | | 3232 | 728 | 0 | 0 | 4199 | 1619 | 0 | 0 | 1 | 1 | 1 | 1 | 1 | 1 | 1 | 0 | 0 | 0 | 1 | 0 | 0 | |
| 239 | F | 39-82 | | 4231 | 3193 | 0 | 0 | 3752 | 1541 | 0 | 0 | 1 | 0 | 1 | 0 | 0 | 0 | 0 | 1 | 1 | 0 | 1 | 1 | 0 | |
| 240 | F | 19-38 | | 2737 | 3107 | 0 | 0 | 1012 | 740 | 0 | 0 | 0 | 0 | 0 | 0 | 0 | 0 | 0 | 0 | 0 | 0 | 0 | 0 | 0 | |
| 241 | F | 39-82 | | 34385 | 4321 | 0 | 1 | 14743 | 1961 | 0 | 1 | 1 | 1 | 0 | 0 | 0 | 0 | 1 | 0 | 0 | 1 | 1 | 0 | 1 | 1 |
| 242 | M | 39-82 | | 11362 | 1057 | 0 | 1 | 10021 | 504 | 0 | 0 | 0 | 0 | 0 | 0 | 0 | 0 | 0 | 0 | 0 | 0 | 0 | 0 | 0 | |
| 243 | F | 19-38 | | 2284 | 741 | 0 | 0 | 2171 | 1813 | 0 | 0 | 1 | 0 | 0 | 0 | 0 | 0 | 0 | 0 | 0 | 0 | 1 | 0 | 0 | |
| 244 | F | 19-38 | | 54690 | 77755 | 85 | 1 | 29863 | 62151 | 92 | 1 | 1 | 1 | 1 | 0 | 1 | 1 | 1 | 1 | 0 | 1 | 1 | 1 | 1 | 1 |
| 245 | F | 39-82 | | 4059 | 1595 | 0 | 0 | 1771 | 637 | 0 | 0 | 0 | 0 | 0 | 0 | 0 | 0 | 0 | 0 | 0 | 0 | 0 | 0 | 0 | |
| 246 | M | 19-38 | | 67594 | 61976 | 77 | 1 | 27465 | 36114 | 50 | 1 | 1 | 1 | 1 | 0 | 0 | 0 | 1 | 0 | 0 | 1 | 1 | 0 | 1 | 1 |
| 247 | F | 19-38 | IgG/IgM positi | 10703 | 72044 | 89 | 1 | 10218 | 55700 | 73 | 1 | 1 | 0 | 1 | 0 | 1 | 1 | 1 | 0 | 0 | 1 | 1 | 0 | 1 | 1 |
| 248 | F | 19-38 | | 2982 | 2996 | 0 | 0 | #N/A | #N/A | #N/A | #N/A | 1 | 0 | 1 | 0 | 1 | 1 | 0 | 0 | 0 | 0 | 1 | 0 | 1 | 0 |
| 249 | F | 19-38 | negative | 2216 | 1686 | 0 | 0 | #N/A | #N/A | #N/A | #N/A | 1 | 0 | 1 | 1 | 0 | 0 | 1 | 0 | 0 | 1 | 1 | 0 | 0 | |
| 250 | F | 39-82 | | 3595 | 863 | 0 | 0 | 2237 | 649 | 0 | 0 | 1 | 0 | 0 | 0 | 0 | 0 | 0 | 0 | 0 | 0 | 1 | 0 | 0 | |
| 251 | F | 19-38 | | 2018 | 1243 | 0 | 0 | 1173 | 1329 | 0 | 0 | 1 | 0 | 0 | 0 | 0 | 0 | 1 | 0 | 1 | 0 | 1 | 0 | 0 | |
| 252 | F | 19-38 | | 2695 | 1205 | 0 | 0 | #N/A | #N/A | #N/A | #N/A | 1 | 0 | 1 | 0 | 0 | 0 | 0 | 0 | 0 | 0 | 1 | 0 | 0 | |
| 253 | F | 39-82 | | 95408 | 91804 | 99 | 1 | 47728 | 87546 | 89 | 1 | 0 | 0 | 0 | 0 | 0 | 0 | 0 | 0 | 0 | 0 | 0 | 0 | 0 | |
| 254 | F | 19-38 | | 2074 | 2418 | 0 | 0 | #N/A | #N/A | #N/A | #N/A | 1 | 1 | 1 | 0 | 0 | 0 | 1 | 0 | 0 | 0 | 1 | 1 | 0 | |
| 255 | F | 39-82 | | 1822 | 1172 | 0 | 0 | 951 | 643 | 0 | 0 | 0 | 0 | 0 | 0 | 0 | 0 | 0 | 0 | 0 | 0 | 0 | 0 | 0 | |
| 256 | F | 39-82 | | 131553 | 29173 | 35 | 1 | #N/A | #N/A | #N/A | #N/A | 1 | 1 | 1 | 1 | 1 | 0 | 0 | 0 | 1 | 0 | 1 | 0 | 1 | 0 |
| 257 | F | 39-82 | | 7968 | 1067 | 0 | 0 | 5185 | 1439 | 7 | 1 | 1 | 0 | 0 | 0 | 0 | 0 | 0 | 0 | 1 | 0 | 0 | 0 | 0 | |
| 258 | F | 19-38 | | 3424 | 723 | 0 | 0 | 2843 | 1875 | 0 | 0 | 1 | 0 | 1 | 0 | 0 | 0 | 0 | 0 | 0 | 0 | 1 | 0 | 0 | |
| 259 | F | 19-38 | | 3559 | 923 | 0 | 0 | #N/A | #N/A | #N/A | #N/A | 1 | 0 | 1 | 1 | 0 | 0 | 1 | 0 | 0 | 0 | 1 | 0 | 0 | |
| 260 | M | 19-38 | | 67158 | 73143 | 93 | 1 | #N/A | #N/A | #N/A | #N/A | 1 | 1 | 1 | 1 | 0 | 1 | 1 | 0 | 0 | 0 | 0 | 0 | 1 | 1 |
| 261 | F | 39-82 | | 5060 | 736 | 0 | 0 | 3981 | 284 | 0 | 0 | 1 | 1 | 1 | 1 | 0 | 1 | 1 | 0 | 0 | 0 | 1 | 0 | 0 | |
| 262 | F | 39-82 | | 847 | 1565 | 0 | 0 | 1102 | 580 | 0 | 0 | 1 | 0 | 0 | 0 | 0 | 0 | 0 | 1 | 0 | 0 | 0 | 0 | 0 | |
| 263 | F | 19-38 | | 1521 | 3429 | 0 | 0 | 755 | 2421 | 0 | 0 | 1 | 0 | 1 | 0 | 1 | 0 | 0 | 0 | 0 | 0 | 0 | 0 | 0 | |
| 264 | F | 39-82 | | 13219 | 1555 | 0 | 1 | #N/A | #N/A | #N/A | #N/A | 0 | 0 | 0 | 0 | 0 | 0 | 0 | 0 | 0 | 0 | 0 | 0 | 0 | |
| 265 | F | 19-38 | | 2230 | 1232 | 0 | 0 | #N/A | #N/A | #N/A | #N/A | 1 | 0 | 1 | 1 | 1 | 1 | 0 | 0 | 0 | 0 | 0 | 0 | 0 | |
| 266 | F | 19-38 | IgM traces | 3616 | 13099 | 0 | 1 | 2237 | 1411 | 0 | 0 | 1 | 1 | 1 | 0 | 0 | 0 | 0 | 0 | 0 | 1 | 1 | 1 | 1 | 1 |
| 267 | F | 39-82 | | 173896 | 179366 | 94 | 1 | #N/A | #N/A | #N/A | #N/A | 1 | 1 | 1 | 0 | 1 | 1 | 1 | 0 | 0 | 1 | 1 | 0 | 1 | 1 |
| 269 | F | 39-82 | | 1641 | 1277 | 0 | 0 | 1853 | 1488 | 0 | 0 | 1 | 0 | 0 | 0 | 0 | 0 | 0 | 0 | 1 | 0 | 0 | 0 | 1 | 0 |
| 270 | F | 19-38 | | 70174 | 57036 | 86 | 1 | #N/A | #N/A | #N/A | #N/A | 0 | 0 | 0 | 0 | 0 | 0 | 0 | 0 | 0 | 0 | 0 | 0 | 0 | |
| 271 | F | 39-82 | | 3177 | 458 | 0 | 0 | 1174 | 618 | 0 | 0 | 0 | 0 | 0 | 0 | 0 | 0 | 0 | 0 | 0 | 0 | 0 | 0 | 0 | |
| 272 | F | 39-82 | | 4616 | 1695 | 0 | 0 | 1372 | 976 | 0 | 0 | 0 | 0 | 0 | 0 | 0 | 0 | 0 | 0 | 0 | 0 | 0 | 0 | 0 | |
| 273 | F | 19-38 | | 2748 | 1527 | 0 | 0 | 1509 | 2193 | 0 | 0 | 0 | 0 | 0 | 0 | 0 | 0 | 0 | 0 | 0 | 0 | 0 | 0 | 0 | |
| 274 | F | 39-82 | | 1147 | 1571 | 0 | 0 | 1484 | 2350 | 0 | 0 | 1 | 0 | 1 | 0 | 1 | 1 | 1 | 0 | 0 | 0 | 0 | 1 | 0 | |
| 275 | F | 19-38 | | 1521 | 777 | 0 | 0 | 933 | 623 | 0 | 0 | 0 | 0 | 0 | 0 | 0 | 0 | 0 | 0 | 0 | 0 | 0 | 0 | 0 | |
| 276 | F | 39-82 | | 286 | 1537 | 0 | 0 | 536 | 832 | 0 | 0 | 0 | 0 | 0 | 0 | 0 | 0 | 0 | 0 | 0 | 0 | 0 | 0 | 0 | |
| 277 | F | 39-82 | | 7001 | 2937 | 0 | 0 | 2542 | 4609 | 11 | 1 | 0 | 0 | 0 | 0 | 0 | 0 | 0 | 0 | 0 | 0 | 0 | 0 | 0 | |
| 278 | F | 39-82 | | 11577 | 1505 | 0 | 1 | 12023 | 129422 | 0 | 1 | 1 | 0 | 1 | 0 | 0 | 0 | 0 | 0 | 0 | 0 | 1 | 0 | 0 | |
| 279 | M | 19-38 | | 9321 | 3988 | 0 | 0 | #N/A | #N/A | #N/A | #N/A | 1 | 1 | 1 | 0 | 0 | 0 | 1 | 0 | 1 | 0 | 0 | 0 | 0 | |
| 280 | F | 39-82 | | 2353 | 1760 | 0 | 0 | 2525 | 1067 | 0 | 0 | 1 | 0 | 0 | 0 | 1 | 0 | 0 | 0 | 1 | 0 | 0 | 0 | 0 | |
| 281 | F | 19-38 | | 1985 | 1317 | 0 | 0 | 1957 | 985 | 0 | 0 | 1 | 0 | 1 | 0 | 1 | 0 | 1 | 0 | 0 | 0 | 0 | 0 | 1 | 0 |
| 282 | F | 39-82 | | 1408 | 987 | 0 | 0 | 1002 | 648 | 0 | 0 | 1 | 0 | 1 | 1 | 1 | 1 | 1 | 0 | 0 | 0 | 0 | 0 | 1 | 0 |
| 283 | F | 19-38 | | 44465 | 194351 | 98 | 1 | 23809 | 115087 | 97 | 1 | 1 | 1 | 1 | 1 | 1 | 1 | 1 | 1 | 0 | 1 | 1 | 1 | 1 | 1 |
| 284 | F | 19-38 | | 7538 | 1267 | 0 | 0 | 7470 | 782 | 0 | 0 | 1 | 0 | 1 | 0 | 0 | 0 | 0 | 0 | 1 | 0 | 0 | 0 | 0 | |
| 285 | M | 39-82 | | 3495 | 2866 | 0 | 0 | 2475 | 1599 | 0 | 0 | 0 | 0 | 0 | 0 | 0 | 0 | 0 | 0 | 0 | 0 | 0 | 0 | 0 | |
| 286 | F | 19-38 | | 3134 | 3139 | 0 | 0 | #N/A | #N/A | #N/A | #N/A | 0 | 0 | 0 | 0 | 0 | 0 | 0 | 0 | 0 | 0 | 0 | 0 | 0 | |
| 287 | F | 39-82 | | 2295 | 545 | 0 | 0 | #N/A | #N/A | #N/A | #N/A | 1 | 1 | 1 | 1 | 1 | 1 | 1 | 0 | 0 | 0 | 1 | 1 | 0 | |
| 288 | F | 39-82 | | 1859 | 400 | 0 | 0 | 681 | 443 | 0 | 0 | 1 | 0 | 1 | 1 | 1 | 1 | 0 | 0 | 0 | 0 | 1 | 0 | 0 | |
| 289 | F | 19-38 | | 6727 | 581 | 0 | 0 | #N/A | #N/A | #N/A | #N/A | 1 | 0 | 1 | 0 | 0 | 0 | 1 | 0 | 1 | 0 | 1 | 0 | 0 | |
| 290 | F | 39-82 | | 14309 | 11179 | 0 | 1 | #N/A | #N/A | #N/A | #N/A | 1 | 0 | 1 | 1 | 0 | 0 | 1 | 0 | 1 | 0 | 0 | 0 | 0 | |
| 291 | F | 39-82 | | 1597 | 327 | 0 | 0 | 765 | 684 | 0 | 0 | 0 | 0 | 0 | 0 | 0 | 0 | 0 | 0 | 0 | 0 | 0 | 0 | 0 | |
| 292 | M | 39-82 | | 2528 | 318 | 0 | 0 | 1532 | 673 | 0 | 0 | 0 | 0 | 0 | 1 | 0 | 0 | 0 | 0 | 0 | 0 | 0 | 0 | 1 | 0 |
| 293 | F | 19-38 | | 2323 | 761 | 0 | 0 | 1762 | 1842 | 0 | 0 | 1 | 0 | 0 | 1 | 0 | 0 | 0 | 0 | 1 | 0 | 1 | 0 | 0 | |
| 294 | F | 39-82 | | 3637 | 636 | 0 | 0 | 3802 | 457 | 0 | 0 | 1 | 0 | 0 | 0 | 0 | 0 | 0 | 0 | 0 | 0 | 1 | 0 | 0 | |
| 295 | F | 39-82 | | 2910 | 276 | 0 | 0 | 1639 | 626 | 0 | 0 | 0 | 0 | 0 | 0 | 0 | 0 | 0 | 0 | 0 | 0 | 0 | 0 | 0 | |
| 296 | M | 19-38 | | 26593 | 7303 | 9 | 1 | #N/A | #N/A | #N/A | #N/A | 1 | 1 | 1 | 1 | 0 | 0 | 1 | 0 | 1 | 1 | 1 | 0 | 0 | |
| 297 | F | 39-82 | | 122976 | 154027 | 95 | 1 | 73920 | 74464 | 88 | 1 | 0 | 0 | 0 | 0 | 0 | 0 | 0 | 0 | 0 | 0 | 0 | 0 | 0 | |
| 299 | F | 39-82 | | 47639 | 32992 | 84 | 1 | #N/A | #N/A | #N/A | #N/A | 1 | 0 | 1 | 1 | 0 | 0 | 1 | 0 | 0 | 0 | 1 | 0 | 0 | |
| 300 | F | 39-82 | | 4311 | 931 | 0 | 0 | 1582 | 533 | 0 | 0 | 1 | 0 | 0 | 0 | 0 | 0 | 0 | 0 | 0 | 0 | 1 | 0 | 0 | |
| 301 | F | 19-38 | | 4000 | 1047 | 0 | 0 | 1541 | 1043 | 0 | 0 | 1 | 0 | 0 | 1 | 0 | 0 | 0 | 0 | 1 | 0 | 1 | 1 | 0 | |
| 302 | F | 19-38 | | 4477 | 801 | 0 | 0 | 4485 | 1007 | 0 | 0 | 0 | 0 | 0 | 0 | 0 | 0 | 0 | 0 | 0 | 0 | 0 | 0 | 0 | |
| 303 | F | 19-38 | | 4179 | 1103 | 0 | 0 | 4069 | 3630 | 0 | 0 | 1 | 0 | 1 | 0 | 0 | 0 | 0 | 0 | 0 | 0 | 0 | 0 | 0 | |
| 304 | F | 39-82 | | 3307 | 1030 | 0 | 0 | #N/A | #N/A | #N/A | #N/A | 1 | 0 | 1 | 0 | 1 | 0 | 1 | 0 | 1 | 0 | 1 | 1 | 1 | 0 |
| 305 | F | 39-82 | | 5599 | 538 | 0 | 0 | 1613 | 1374 | 0 | 0 | 0 | 0 | 0 | 0 | 0 | 0 | 0 | 0 | 0 | 0 | 0 | 0 | 0 | |
| 306 | F | 39-82 | | 2384 | 368 | 0 | 0 | 1181 | 555 | 18 | 1 | 1 | 0 | 1 | 1 | 1 | 0 | 1 | 0 | 1 | 0 | 1 | 1 | 0 | |

| | | | | | | | | | | | | | | | | | | | | | | | | | |
|---|---|---|---|---|---|---|---|---|---|---|---|---|---|---|---|---|---|---|---|---|---|---|---|---|---|
| 307 | F | 39-82 | | 44320 | 81752 | 48 | 1 | 15237 | 45152 | 0 | 1 | 0 | 0 | 0 | 0 | 0 | 0 | 0 | 0 | 0 | 0 | 0 | 0 | 0 | |
| 308 | F | 39-82 | | 2526 | 811 | 0 | 0 | 2833 | 2607 | 0 | 0 | 1 | 0 | 0 | 1 | 0 | 0 | 0 | 0 | 1 | 0 | 0 | 0 | 0 | |
| 309 | F | 19-38 | | 4118 | 3120 | 0 | 0 | #N/A | #N/A | #N/A | #N/A | 0 | 0 | 0 | 0 | 0 | 0 | 0 | 0 | 0 | 0 | 0 | 0 | 0 | |
| 310 | F | 39-82 | negative | 1003 | 389 | 0 | 0 | 1012 | 1027 | 0 | 0 | 1 | 0 | 1 | 0 | 0 | 0 | 0 | 0 | 0 | 1 | 0 | 0 | 0 | |
| 311 | F | 39-82 | | 1523 | 2413 | 0 | 0 | 1238 | 5684 | 0 | 0 | 1 | 1 | 1 | 0 | 0 | 0 | 0 | 0 | 0 | 0 | 0 | 1 | 0 | |
| 312 | F | 39-82 | | 919 | 422 | 0 | 0 | #N/A | #N/A | #N/A | #N/A | 0 | 0 | 0 | 1 | 0 | 0 | 0 | 0 | 0 | 0 | 0 | 0 | 0 | |
| 313 | F | 39-82 | | 186846 | 153959 | 97 | 1 | 122613 | 71249 | 3 | 1 | 0 | 0 | 0 | 0 | 0 | 0 | 0 | 0 | 0 | 0 | 0 | 0 | 0 | |
| 314 | M | 39-82 | | 3263 | 410 | 0 | 0 | 3324 | 1285 | 0 | 0 | 0 | 0 | 0 | 0 | 0 | 0 | 0 | 0 | 0 | 0 | 0 | 0 | 0 | |
| 315 | F | 19-38 | | 8782 | 1844 | 0 | 0 | 3134 | 1630 | 0 | 0 | 1 | 1 | 1 | 0 | 0 | 0 | 1 | 0 | 0 | 0 | 1 | 1 | 0 | |
| 316 | F | 19-38 | | 2092 | 13802 | 0 | 1 | 1831 | 11601 | 0 | 1 | 0 | 0 | 0 | 0 | 0 | 0 | 0 | 0 | 0 | 0 | 0 | 0 | 0 | |
| 317 | F | 39-82 | | 3882 | 461 | 0 | 0 | 1604 | 656 | 0 | 0 | 0 | 0 | 0 | 0 | 0 | 0 | 0 | 0 | 0 | 0 | 0 | 0 | 0 | |
| 318 | F | 39-82 | | 3058 | 656 | 0 | 0 | 3256 | 1083 | 0 | 0 | 1 | 0 | 0 | 0 | 0 | 0 | 0 | 0 | 1 | 0 | 0 | 0 | 0 | |
| 319 | F | 39-82 | | 3375 | 523 | 0 | 0 | 3867 | 937 | 0 | 0 | 0 | 0 | 0 | 0 | 0 | 0 | 0 | 0 | 0 | 0 | 0 | 0 | 0 | |
| 320 | M | 39-82 | | 7717 | 483 | 0 | 0 | 3244 | 702 | 0 | 0 | 1 | 0 | 0 | 0 | 0 | 0 | 0 | 0 | 1 | 0 | 0 | 0 | 0 | |
| 321 | F | 39-82 | | 2198 | 367 | 0 | 0 | #N/A | #N/A | #N/A | #N/A | 0 | 0 | 0 | 0 | 0 | 0 | 0 | 0 | 0 | 0 | 0 | 0 | 0 | |
| 322 | F | 19-38 | | 4921 | 1323 | 0 | 0 | 2240 | 2703 | 0 | 0 | 0 | 0 | 0 | 0 | 0 | 0 | 0 | 0 | 0 | 0 | 0 | 0 | 0 | |
| 323 | M | 39-82 | | 2117 | 1321 | 0 | 0 | #N/A | #N/A | #N/A | #N/A | 0 | 0 | 0 | 0 | 0 | 0 | 0 | 0 | 0 | 0 | 0 | 0 | 0 | |
| 324 | F | 19-38 | | 920 | 1904 | 0 | 0 | 1901 | 1209 | 0 | 0 | 0 | 0 | 0 | 0 | 0 | 0 | 0 | 0 | 0 | 0 | 0 | 0 | 0 | |
| 325 | F | 19-38 | | 4989 | 373 | 0 | 0 | 14529 | 963 | 0 | 1 | 0 | 0 | 0 | 0 | 0 | 0 | 0 | 0 | 0 | 0 | 0 | 0 | 0 | |
| 326 | M | 39-82 | | 3009 | 321 | 0 | 0 | 6289 | 505 | 0 | 0 | 1 | 0 | 1 | 1 | 0 | 0 | 0 | 0 | 0 | 0 | 0 | 0 | 0 | |
| 327 | F | 19-38 | | 3716 | 587 | 0 | 0 | #N/A | #N/A | #N/A | #N/A | 1 | 0 | 0 | 0 | 0 | 0 | 0 | 0 | 0 | 0 | 0 | 0 | 0 | |
| 328 | F | 39-82 | | 2595 | 594 | 0 | 0 | 2979 | 1249 | 0 | 0 | 0 | 0 | 0 | 0 | 0 | 0 | 0 | 0 | 0 | 0 | 0 | 0 | 0 | |
| 329 | F | 19-38 | | 1914 | 2307 | 0 | 0 | 2623 | 2232 | 0 | 0 | 0 | 0 | 0 | 0 | 0 | 0 | 0 | 0 | 0 | 0 | 0 | 0 | 0 | |
| 330 | F | 39-82 | | 1453 | 1249 | 0 | 0 | 1765 | 974 | 0 | 0 | 1 | 1 | 0 | 1 | 0 | 0 | 0 | 0 | 0 | 0 | 0 | 0 | 0 | |
| 331 | F | 19-38 | | 2049 | 755 | 0 | 0 | 2259 | 1213 | 0 | 0 | 1 | 0 | 1 | 0 | 0 | 0 | 0 | 0 | 0 | 0 | 0 | 1 | 0 | |
| 332 | F | 19-38 | | 695 | 510 | 0 | 0 | 1537 | 1305 | 0 | 0 | 0 | 0 | 0 | 0 | 0 | 0 | 0 | 0 | 0 | 0 | 0 | 0 | 0 | |
| 333 | F | 19-38 | | 1780 | 741 | 0 | 0 | 1341 | 1459 | 0 | 0 | 0 | 0 | 0 | 0 | 0 | 0 | 0 | 0 | 0 | 0 | 0 | 0 | 0 | |
| 334 | M | 39-82 | | 2058 | 1144 | 0 | 0 | 3700 | 1025 | 0 | 0 | 1 | 0 | 0 | 1 | 0 | 0 | 0 | 0 | 1 | 0 | 0 | 0 | 1 | 0 |
| 335 | M | 19-38 | | 5353 | 1502 | 0 | 0 | 6730 | 1089 | 0 | 0 | 0 | 0 | 0 | 0 | 0 | 0 | 0 | 0 | 0 | 0 | 0 | 0 | 0 | |
| 336 | M | 19-38 | | 2930 | 2391 | 0 | 0 | 2891 | 5193 | 0 | 0 | 1 | 0 | 1 | 1 | 1 | 0 | 1 | 0 | 1 | 0 | 1 | 1 | 0 | |
| 337 | F | 39-82 | | 1351 | 1100 | 0 | 0 | 847 | 2011 | 0 | 0 | 0 | 0 | 0 | 0 | 0 | 0 | 0 | 0 | 0 | 0 | 0 | 0 | 0 | |
| 339 | F | 39-82 | | 947 | 325 | 0 | 0 | 6417 | 610 | 0 | 0 | 0 | 0 | 0 | 0 | 0 | 0 | 0 | 0 | 0 | 0 | 0 | 0 | 0 | |
| 341 | F | 19-38 | | 1586 | 441 | 0 | 0 | 1067 | 1738 | 0 | 0 | 1 | 0 | 1 | 0 | 0 | 0 | 1 | 0 | 0 | 0 | 1 | 0 | 0 | |
| 342 | F | 39-82 | | 43237 | 42915 | 85 | 1 | 23395 | 25275 | 0 | 1 | 1 | 1 | 1 | 1 | 1 | 1 | 1 | 0 | 0 | 0 | 1 | 0 | 1 | 1 |
| 343 | F | 39-82 | | 116644 | 108723 | 97 | 1 | 48353 | 70011 | 85 | 1 | 1 | 0 | 1 | 0 | 0 | 0 | 1 | 0 | 1 | 1 | 1 | 0 | 0 | |
| 345 | F | 19-38 | | 2314 | 603 | 0 | 0 | 3117 | 2048 | 0 | 0 | 0 | 0 | 0 | 0 | 0 | 0 | 0 | 0 | 0 | 0 | 0 | 0 | 0 | |
| 346 | F | 39-82 | | 1217 | 354 | 0 | 0 | #N/A | #N/A | #N/A | #N/A | 0 | 0 | 0 | 0 | 0 | 0 | 0 | 0 | 0 | 0 | 0 | 0 | 0 | |
| 348 | F | 19-38 | | 4847 | 1117 | 0 | 0 | #N/A | #N/A | #N/A | #N/A | 1 | 0 | 1 | 0 | 1 | 0 | 1 | 0 | 0 | 0 | 1 | 0 | 0 | |
| 349 | M | 19-38 | | 2804 | 1422 | 0 | 0 | #N/A | #N/A | #N/A | #N/A | 1 | 0 | 0 | 0 | 0 | 0 | 0 | 0 | 0 | 0 | 0 | 1 | 1 | 0 |
| 350 | F | 39-82 | | 105204 | 53600 | 11 | 1 | 70204 | 43775 | 0 | 1 | 1 | 0 | 1 | 1 | 1 | 0 | 1 | 0 | 0 | 1 | 1 | 0 | 0 | |
| 351 | F | 19-38 | | 3042 | 820 | 0 | 0 | #N/A | #N/A | #N/A | #N/A | 1 | 0 | 1 | 0 | 0 | 0 | 0 | 0 | 0 | 0 | 0 | 0 | 0 | |
| 352 | F | 39-82 | negative | 5920 | 3689 | 0 | 0 | 4848 | 3026 | 0 | 0 | 1 | 1 | 1 | 1 | 1 | 1 | 1 | 0 | 0 | 1 | 1 | 0 | 1 | 0 |
| 353 | F | 39-82 | | 1633 | 848 | 0 | 0 | 458 | 781 | 14 | 1 | 0 | 0 | 0 | 0 | 0 | 0 | 0 | 0 | 0 | 0 | 0 | 0 | 0 | |
| 354 | F | 19-38 | | 4760 | 477 | 0 | 0 | #N/A | #N/A | #N/A | #N/A | 0 | 0 | 0 | 0 | 0 | 0 | 0 | 0 | 0 | 0 | 0 | 0 | 0 | |
| 355 | F | 39-82 | | 9638 | 2397 | 0 | 0 | 478 | 439 | 0 | 0 | 0 | 0 | 0 | 0 | 0 | 0 | 0 | 0 | 0 | 0 | 0 | 0 | 0 | |
| 356 | F | 19-38 | | 2035 | 624 | 0 | 0 | 6856 | 638 | 0 | 0 | 1 | 0 | 1 | 0 | 0 | 0 | 0 | 0 | 0 | 0 | 1 | 0 | 0 | |
| 357 | F | 19-38 | | 2966 | 357 | 0 | 0 | 1573 | 1238 | 0 | 0 | 0 | 0 | 0 | 0 | 0 | 0 | 0 | 0 | 0 | 0 | 0 | 0 | 0 | |
| 358 | M | 19-38 | | 1506 | 812 | 0 | 0 | #N/A | #N/A | #N/A | #N/A | 1 | 0 | 0 | 0 | 0 | 0 | 1 | 0 | 0 | 0 | 0 | 0 | 0 | |
| 359 | M | 39-82 | | 4492 | 561 | 0 | 0 | 2372 | 2828 | 46 | 1 | 1 | 0 | 0 | 0 | 0 | 0 | 0 | 0 | 0 | 0 | 0 | 1 | 0 | |
| 360 | F | 19-38 | | 9255 | 626 | 0 | 0 | 4839 | 2108 | 0 | 0 | 0 | 0 | 0 | 0 | 0 | 0 | 0 | 0 | 0 | 0 | 0 | 0 | 0 | |
| 361 | M | 39-82 | | 2057 | 484 | 0 | 0 | #N/A | #N/A | #N/A | #N/A | 0 | 0 | 0 | 0 | 0 | 0 | 0 | 0 | 0 | 0 | 0 | 0 | 0 | |
| 362 | F | 39-82 | | 4963 | 1419 | 0 | 0 | 25463 | 31500 | 0 | 1 | 1 | 1 | 1 | 0 | 1 | 0 | 1 | 0 | 1 | 1 | 1 | 1 | 0 | |
| 363 | F | 19-38 | | 2407 | 691 | 0 | 0 | 5233 | 844 | 0 | 0 | 0 | 0 | 0 | 0 | 0 | 0 | 0 | 0 | 0 | 0 | 0 | 0 | 0 | |
| 364 | F | 39-82 | | 9867 | 1296 | 0 | 0 | 2341 | 2568 | 0 | 0 | 1 | 0 | 1 | 0 | 0 | 0 | 0 | 0 | 0 | 0 | 1 | 0 | 0 | |
| 365 | F | 39-82 | | 1636 | 212 | 0 | 0 | 722 | 591 | 0 | 0 | 0 | 0 | 0 | 0 | 0 | 0 | 0 | 0 | 0 | 0 | 0 | 0 | 0 | |
| 366 | F | 39-82 | | 59135 | 79266 | 90 | 1 | 26588 | 38804 | 95 | 1 | 1 | 0 | 1 | 0 | 0 | 0 | 0 | 0 | 1 | 0 | 1 | 0 | 0 | |
| 367 | F | 19-38 | | 3762 | 1265 | 0 | 0 | 2327 | 2306 | 0 | 0 | 1 | 0 | 1 | 0 | 0 | 0 | 0 | 0 | 0 | 0 | 1 | 0 | 0 | |
| 368 | F | 19-38 | | 2539 | 324 | 0 | 0 | 2120 | 819 | 0 | 0 | 1 | 0 | 1 | 0 | 0 | 0 | 0 | 0 | 0 | 0 | 1 | 0 | 0 | |
| 369 | F | 39-82 | | 4079 | 1162 | 0 | 0 | #N/A | #N/A | #N/A | #N/A | 1 | 0 | 0 | 1 | 1 | 0 | 0 | 0 | 1 | 0 | 1 | 0 | 0 | |
| 370 | F | 19-38 | | 91928 | 11761 | 79 | 1 | 44315 | 23610 | 0 | 1 | 1 | 0 | 1 | 1 | 0 | 1 | 1 | 1 | 1 | 0 | 1 | 0 | 0 | |
| 371 | M | 39-82 | | 4794 | 764 | 0 | 0 | #N/A | #N/A | #N/A | #N/A | 1 | 0 | 1 | 0 | 0 | 0 | 0 | 0 | 1 | 0 | 1 | 0 | 0 | |
| 372 | F | 19-38 | | 2085 | 520 | 0 | 0 | 722 | 530 | 0 | 0 | 0 | 0 | 0 | 0 | 0 | 0 | 0 | 0 | 0 | 0 | 0 | 0 | 0 | |
| 373 | M | 39-82 | | 1699 | 270 | 0 | 0 | #N/A | #N/A | #N/A | #N/A | 1 | 1 | 0 | 0 | 0 | 0 | 0 | 0 | 0 | 0 | 1 | 0 | 0 | |
| 374 | F | 39-82 | | 4868 | 682 | 0 | 0 | 2166 | 1326 | 0 | 0 | 1 | 0 | 1 | 0 | 0 | 0 | 1 | 1 | 0 | 0 | 1 | 0 | 0 | |
| 375 | F | 39-82 | | 2250 | 559 | 0 | 0 | #N/A | #N/A | #N/A | #N/A | 1 | 1 | 0 | 1 | 0 | 0 | 0 | 0 | 1 | 0 | 0 | 0 | 0 | |
| 376 | F | 19-38 | | 3283 | 474 | 0 | 0 | 683 | 485 | 0 | 0 | 0 | 0 | 0 | 0 | 0 | 0 | 0 | 0 | 0 | 0 | 0 | 0 | 0 | |
| 377 | M | 39-82 | | 1271 | 485 | 0 | 0 | #N/A | #N/A | #N/A | #N/A | 0 | 0 | 0 | 0 | 0 | 0 | 0 | 0 | 0 | 0 | 0 | 0 | 0 | |
| 378 | M | 39-82 | | 4419 | 538 | 0 | 0 | #N/A | #N/A | #N/A | #N/A | 1 | 0 | 1 | 1 | 0 | 0 | 0 | 0 | 1 | 0 | 0 | 1 | 1 | 0 |
| 379 | F | 19-38 | | 1878 | 217 | 0 | 0 | #N/A | #N/A | #N/A | #N/A | 1 | 0 | 0 | 0 | 0 | 0 | 0 | 1 | 0 | 0 | 0 | 0 | 0 | |
| 380 | F | 39-82 | | 1221 | 430 | 0 | 0 | 2000 | 518 | 0 | 0 | 0 | 0 | 0 | 0 | 0 | 0 | 0 | 0 | 0 | 0 | 0 | 0 | 0 | |
| 381 | F | 39-82 | | 2533 | 545 | 0 | 0 | 1041 | 682 | 0 | 0 | 1 | 0 | 1 | 0 | 1 | 0 | 0 | 0 | 0 | 0 | 1 | 1 | 0 | |
| 382 | F | 19-38 | | 1962 | 641 | 0 | 0 | 2631 | 933 | 0 | 0 | 0 | 0 | 0 | 0 | 0 | 0 | 0 | 0 | 0 | 0 | 0 | 0 | 0 | |
| 383 | M | 39-82 | | 2219 | 328 | 0 | 0 | 734 | 1095 | 0 | 0 | 0 | 0 | 0 | 0 | 0 | 0 | 0 | 0 | 0 | 0 | 0 | 0 | 0 | |
| 384 | F | 39-82 | | 3480 | 496 | 0 | 0 | 3909 | 539 | 0 | 0 | 0 | 0 | 0 | 1 | 0 | 0 | 0 | 0 | 0 | 0 | 0 | 0 | 0 | |
| 385 | F | 19-38 | | 2225 | 916 | 0 | 0 | #N/A | #N/A | #N/A | #N/A | 0 | 0 | 0 | 0 | 0 | 0 | 0 | 0 | 0 | 0 | 0 | 0 | 0 | |
| 386 | F | 39-82 | | 3682 | 1642 | 0 | 0 | 4186 | 4861 | 0 | 0 | 0 | 0 | 0 | 0 | 0 | 0 | 0 | 0 | 0 | 0 | 0 | 0 | 0 | |
| 387 | F | 39-82 | | 8616 | 661 | 0 | 0 | 8757 | 349 | 0 | 0 | 1 | 0 | 1 | 0 | 0 | 0 | 0 | 0 | 0 | 0 | 1 | 0 | 0 | |

| | | | | | | | | | | | | | | | | | | | | | | | | | |
|---|---|---|---|---|---|---|---|---|---|---|---|---|---|---|---|---|---|---|---|---|---|---|---|---|---|
| 388 | F | 19-38 | | 2715 | 918 | 0 | 0 | #N/A | #N/A | #N/A | #N/A | 0 | 0 | 0 | 0 | 0 | 0 | 0 | 0 | 0 | 0 | 0 | 0 | 0 | |
| 389 | F | 39-82 | | 3449 | 519 | 0 | 0 | 858 | 509 | 0 | 0 | 1 | 0 | 1 | 0 | 0 | 0 | 0 | 0 | 0 | 0 | 1 | 0 | 0 | |
| 390 | F | 39-82 | | 5687 | 508 | 0 | 0 | 2878 | 1496 | 0 | 0 | 1 | 0 | 1 | 0 | 0 | 0 | 1 | 0 | 0 | 0 | 0 | 0 | 0 | |
| 391 | F | 39-82 | | 2385 | 455 | 0 | 0 | 1325 | 585 | 0 | 0 | 1 | 1 | 1 | 0 | 0 | 0 | 0 | 0 | 0 | 0 | 1 | 0 | 0 | |
| 392 | F | 39-82 | | 2852 | 789 | 0 | 0 | 2317 | 1961 | 0 | 0 | 0 | 0 | 0 | 0 | 0 | 0 | 0 | 0 | 0 | 0 | 0 | 0 | 0 | |
| 393 | F | 19-38 | | 5210 | 1168 | 0 | 0 | 1486 | 2103 | 0 | 0 | 1 | 0 | 0 | 1 | 0 | 0 | 0 | 1 | 0 | 0 | 0 | 0 | 0 | |
| 394 | F | 39-82 | | 3287 | 825 | 0 | 0 | 2230 | 1360 | 0 | 0 | 0 | 0 | 0 | 0 | 0 | 0 | 0 | 0 | 0 | 0 | 0 | 0 | 0 | |
| 395 | F | 19-38 | | 2650 | 1879 | 0 | 0 | 3313 | 1696 | 0 | 0 | 1 | 0 | 0 | 0 | 0 | 0 | 0 | 0 | 0 | 0 | 1 | 0 | 0 | |
| 396 | F | 19-38 | | 2003 | 681 | 0 | 0 | 3608 | 1318 | 2 | 1 | 0 | 0 | 0 | 0 | 0 | 0 | 0 | 0 | 0 | 0 | 0 | 0 | 0 | |
| 397 | F | 39-82 | | 3364 | 749 | 0 | 0 | 4550 | 504 | 0 | 0 | 1 | 0 | 1 | 0 | 0 | 0 | 0 | 0 | 0 | 0 | 0 | 0 | 0 | |
| 398 | F | 19-38 | | 1096 | 370 | 0 | 0 | #N/A | #N/A | #N/A | #N/A | 1 | 1 | 1 | 1 | 0 | 0 | 1 | 0 | 0 | 0 | 1 | 0 | 0 | |
| 399 | F | 19-38 | | 5588 | 740 | 0 | 0 | #N/A | #N/A | #N/A | #N/A | 0 | 0 | 0 | 0 | 0 | 0 | 0 | 0 | 0 | 0 | 0 | 0 | 0 | |
| 400 | F | 19-38 | | 3947 | 644 | 0 | 0 | 3692 | 2145 | 0 | 0 | 1 | 0 | 1 | 0 | 0 | 1 | 0 | 0 | 0 | 0 | 1 | 1 | 0 | |
| 401 | F | 19-38 | | 1080 | 1071 | 0 | 0 | #N/A | #N/A | #N/A | #N/A | 1 | 0 | 1 | 1 | 1 | 1 | 0 | 0 | 0 | 0 | 0 | 0 | 1 | 0 |
| 402 | F | 19-38 | | 1409 | 708 | 0 | 0 | 2062 | 1326 | 0 | 0 | 1 | 0 | 1 | 0 | 0 | 0 | 0 | 0 | 0 | 0 | 1 | 0 | 0 | |
| 403 | F | 39-82 | | 9100 | 2285 | 0 | 0 | 6960 | 1139 | 0 | 0 | 0 | 0 | 0 | 0 | 0 | 0 | 0 | 0 | 0 | 0 | 0 | 0 | 0 | |
| 404 | F | 19-38 | | 7190 | 1894 | 0 | 0 | 1999 | 4323 | 0 | 0 | 0 | 0 | 0 | 0 | 0 | 0 | 0 | 0 | 0 | 0 | 0 | 0 | 0 | |
| 405 | F | 39-82 | | 1853 | 1508 | 0 | 0 | 212 | 233 | 0 | 0 | 0 | 0 | 0 | 0 | 0 | 0 | 0 | 0 | 0 | 0 | 0 | 0 | 0 | |
| 406 | F | 39-82 | | 9996 | 1445 | 0 | 0 | 9417 | 2622 | 0 | 0 | 0 | 0 | 0 | 0 | 0 | 0 | 0 | 0 | 0 | 0 | 0 | 0 | 0 | |
| 407 | F | 19-38 | | 2380 | 574 | 0 | 0 | 1754 | 874 | 0 | 0 | 0 | 0 | 0 | 0 | 0 | 0 | 0 | 0 | 0 | 0 | 0 | 0 | 0 | |
| 408 | F | 39-82 | | 1155 | 1228 | 0 | 0 | 1024 | 1995 | 10 | 1 | 1 | 1 | 1 | 1 | 1 | 1 | 1 | 0 | 1 | 0 | 1 | 0 | 0 | |
| 409 | M | 19-38 | | 5095 | 1080 | 0 | 0 | #N/A | #N/A | #N/A | #N/A | 1 | 1 | 1 | 1 | 0 | 0 | 1 | 0 | 1 | 0 | 1 | 0 | 1 | |
| 410 | M | 39-82 | IgM traces | 2987 | 1308 | 0 | 0 | #N/A | #N/A | #N/A | #N/A | 1 | 1 | 1 | 0 | 0 | 0 | 1 | 0 | 1 | 1 | 1 | 0 | 1 | 0 |
| 411 | F | 39-82 | | 1564 | 797 | 0 | 0 | 1471 | 3877 | 0 | 0 | 1 | 0 | 1 | 1 | 1 | 0 | 1 | 1 | 1 | 0 | 1 | 0 | 0 | |
| 412 | F | 39-82 | | 3300 | 3510 | 0 | 0 | #N/A | #N/A | #N/A | #N/A | 0 | 0 | 0 | 0 | 0 | 0 | 0 | 0 | 0 | 0 | 0 | 0 | 0 | |
| 413 | F | 19-38 | negative | 1513 | 1153 | 0 | 0 | 1260 | 4622 | 0 | 0 | 1 | 1 | 1 | 1 | 0 | 0 | 1 | 0 | 0 | 1 | 0 | 0 | 0 | |
| 414 | M | 39-82 | | 968 | 394 | 0 | 0 | 1955 | 473 | 0 | 0 | 1 | 1 | 1 | 1 | 0 | 0 | 0 | 0 | 1 | 0 | 0 | 0 | 0 | |
| 415 | F | 19-38 | | 1766 | 494 | 0 | 0 | #N/A | #N/A | #N/A | #N/A | 1 | 0 | 1 | 1 | 0 | 0 | 1 | 1 | 1 | 0 | 1 | 1 | 0 | |
| 416 | F | 19-38 | | 3791 | 765 | 0 | 0 | #N/A | #N/A | #N/A | #N/A | 1 | 1 | 1 | 1 | 0 | 0 | 1 | 0 | 1 | 0 | 1 | 1 | 1 | 0 |
| 417 | F | 19-38 | | 2495 | 866 | 0 | 0 | #N/A | #N/A | #N/A | #N/A | 1 | 0 | 1 | 0 | 1 | 0 | 1 | 0 | 0 | 0 | 1 | 0 | 0 | |
| 418 | M | 39-82 | | 1897 | 515 | 0 | 0 | 1663 | 1338 | 0 | 0 | 0 | 0 | 0 | 0 | 0 | 0 | 0 | 0 | 0 | 0 | 0 | 0 | 0 | |
| 420 | M | 19-38 | | 3992 | 831 | 0 | 0 | 2113 | 1324 | 0 | 0 | 1 | 1 | 1 | 1 | 0 | 0 | 1 | 0 | 0 | 0 | 1 | 0 | 0 | |
| 421 | F | 19-38 | | 2699 | 990 | 0 | 0 | 1317 | 1669 | 0 | 0 | 0 | 0 | 0 | 0 | 0 | 0 | 0 | 0 | 0 | 0 | 0 | 0 | 0 | |
| 422 | F | 39-82 | negative | 2115 | 1058 | 0 | 0 | 951 | 1776 | 0 | 0 | 1 | 1 | 1 | 1 | 1 | 1 | 1 | 0 | 0 | 1 | 1 | 0 | 1 | 0 |
| 423 | M | 19-38 | | 3623 | 988 | 0 | 0 | #N/A | #N/A | #N/A | #N/A | 0 | 0 | 0 | 0 | 0 | 0 | 0 | 0 | 0 | 0 | 0 | 0 | 0 | |
| 424 | F | 19-38 | | 2397 | 768 | 0 | 0 | 2186 | 1838 | 0 | 0 | 1 | 0 | 1 | 0 | 1 | 0 | 0 | 0 | 0 | 0 | 1 | 1 | 0 | |
| 425 | F | 39-82 | | 190967 | 32660 | 90 | 1 | 122201 | 62768 | 0 | 1 | 1 | 1 | 0 | 0 | 0 | 0 | 0 | 0 | 0 | 0 | 0 | 1 | 1 | 1 |
| 426 | F | 39-82 | | 3343 | 2811 | 0 | 0 | 4460 | 3144 | 0 | 0 | 0 | 0 | 0 | 0 | 0 | 0 | 0 | 0 | 0 | 0 | 0 | 0 | 0 | |
| 427 | F | 19-38 | | 7989 | 1324 | 0 | 0 | 4124 | 5884 | 0 | 0 | 1 | 0 | 1 | 0 | 0 | 0 | 0 | 0 | 0 | 0 | 0 | 0 | 0 | |
| 428 | M | 19-38 | | 2407 | 1550 | 0 | 0 | #N/A | #N/A | #N/A | #N/A | 1 | 1 | 1 | 0 | 0 | 0 | 0 | 0 | 1 | 0 | 1 | 1 | 1 | 0 |
| 429 | F | 19-38 | | 15939 | 23150 | 2 | 1 | 10334 | 64251 | 89 | 1 | 1 | 0 | 1 | 0 | 0 | 0 | 0 | 0 | 1 | 1 | 0 | 0 | 1 | 0 |
| 430 | F | 19-38 | | 6359 | 846 | 0 | 0 | #N/A | #N/A | #N/A | #N/A | 0 | 0 | 0 | 0 | 0 | 0 | 0 | 0 | 0 | 0 | 0 | 0 | 0 | |
| 431 | F | 39-82 | | 4034 | 821 | 0 | 0 | #N/A | #N/A | #N/A | #N/A | 0 | 0 | 0 | 0 | 0 | 0 | 0 | 0 | 0 | 0 | 0 | 0 | 0 | |
| 432 | F | 19-38 | | 7428 | 562 | 0 | 0 | 3507 | 1082 | 0 | 0 | 1 | 0 | 1 | 0 | 1 | 0 | 1 | 0 | 1 | 0 | 1 | 0 | 0 | |
| 433 | F | 19-38 | | 2732 | 884 | 0 | 0 | 732 | 1056 | 0 | 0 | 1 | 0 | 1 | 0 | 1 | 1 | 1 | 0 | 0 | 0 | 1 | 0 | 0 | |
| 434 | F | 19-38 | | 3133 | 1058 | 0 | 0 | 2422 | 2741 | 0 | 0 | 1 | 1 | 1 | 1 | 0 | 0 | 0 | 0 | 1 | 0 | 1 | 0 | 0 | |
| 435 | F | 19-38 | | 37875 | 923 | 0 | 1 | 47894 | 1021 | 0 | 1 | 1 | 0 | 1 | 0 | 0 | 0 | 0 | 0 | 0 | 1 | 0 | 0 | 1 | 1 |
| 436 | M | 19-38 | | 3811 | 1055 | 0 | 0 | #N/A | #N/A | #N/A | #N/A | 0 | 0 | 0 | 0 | 0 | 0 | 0 | 0 | 0 | 0 | 0 | 0 | 0 | |
| 437 | F | 39-82 | | 1463 | 341 | 0 | 0 | 535 | 380 | 0 | 0 | 1 | 0 | 1 | 0 | 0 | 0 | 1 | 1 | 0 | 0 | 0 | 0 | 0 | |
| 438 | M | 19-38 | | 3138 | 1862 | 0 | 0 | #N/A | #N/A | #N/A | #N/A | 1 | 1 | 0 | 1 | 0 | 0 | 0 | 0 | 1 | 0 | 0 | 0 | 1 | 0 |
| 439 | F | 19-38 | | 3524 | 814 | 0 | 0 | #N/A | #N/A | #N/A | #N/A | 1 | 0 | 0 | 0 | 0 | 0 | 0 | 0 | 0 | 0 | 1 | 1 | 0 | |
| 440 | F | 19-38 | | 3742 | 1283 | 0 | 0 | 1597 | 1285 | 0 | 0 | 1 | 0 | 1 | 0 | 0 | 0 | 0 | 0 | 0 | 0 | 1 | 0 | 0 | |
| 441 | F | 39-82 | | 2621 | 399 | 0 | 0 | 733 | 701 | 0 | 0 | 1 | 0 | 0 | 0 | 0 | 0 | 0 | 0 | 0 | 0 | 1 | 0 | 0 | |
| 442 | F | 39-82 | | 4392 | 1083 | 0 | 0 | #N/A | #N/A | #N/A | #N/A | 0 | 0 | 0 | 0 | 0 | 0 | 0 | 0 | 0 | 0 | 0 | 0 | 0 | |
| 443 | F | 39-82 | | 2019 | 623 | 0 | 0 | #N/A | #N/A | #N/A | #N/A | 1 | 1 | 1 | 0 | 0 | 0 | 0 | 0 | 0 | 0 | 1 | 0 | 0 | |
| 444 | M | 19-38 | | 1643 | 593 | 0 | 0 | #N/A | #N/A | #N/A | #N/A | 1 | 0 | 1 | 0 | 0 | 0 | 0 | 0 | 0 | 0 | 1 | 1 | 0 | |
| 445 | F | 39-82 | | 1726 | 561 | 0 | 0 | 830 | 1177 | 0 | 0 | 0 | 0 | 0 | 0 | 0 | 0 | 0 | 0 | 0 | 0 | 0 | 0 | 0 | |
| 446 | F | 39-82 | | 1504 | 899 | 0 | 0 | 712 | 2275 | 0 | 0 | 1 | 1 | 1 | 0 | 0 | 0 | 0 | 0 | 0 | 0 | 0 | 0 | 0 | |
| 447 | F | 39-82 | | 59688 | 33687 | 5 | 1 | 27002 | 20562 | 3 | 1 | 1 | 0 | 1 | 0 | 0 | 0 | 1 | 0 | 0 | 0 | 0 | 1 | 0 | |
| 448 | F | 39-82 | | 70690 | 25703 | 85 | 1 | #N/A | #N/A | #N/A | #N/A | 1 | 0 | 1 | 0 | 0 | 0 | 0 | 0 | 1 | 1 | 0 | 0 | 0 | |
| 449 | M | 39-82 | | 9532 | 2928 | 0 | 0 | #N/A | #N/A | #N/A | #N/A | 1 | 0 | 1 | 1 | 0 | 0 | 0 | 0 | 1 | 0 | 0 | 0 | 0 | |
| 450 | F | 39-82 | | 1150 | 322 | 0 | 0 | 1261 | 584 | 0 | 0 | 1 | 1 | 0 | 1 | 0 | 0 | 0 | 0 | 1 | 0 | 0 | 1 | 1 | 0 |
| 451 | M | 19-38 | | 2513 | 1505 | 0 | 0 | #N/A | #N/A | #N/A | #N/A | 1 | 0 | 1 | 1 | 0 | 0 | 0 | 0 | 0 | 0 | 0 | 0 | 0 | |
| 452 | F | 39-82 | | 1184 | 439 | 0 | 0 | 936 | 702 | 0 | 0 | 1 | 0 | 0 | 0 | 0 | 0 | 1 | 0 | 0 | 0 | 1 | 0 | 0 | |
| 453 | M | 39-82 | | 1125 | 770 | 0 | 0 | 1324 | 480 | 0 | 0 | 0 | 0 | 0 | 0 | 0 | 0 | 0 | 0 | 0 | 0 | 0 | 0 | 0 | |
| 455 | M | 19-38 | | 1372 | 472 | 0 | 0 | #N/A | #N/A | #N/A | #N/A | 0 | 0 | 0 | 0 | 0 | 0 | 0 | 0 | 0 | 0 | 0 | 0 | 0 | |
| 456 | M | 39-82 | | 2625 | 2059 | 0 | 0 | #N/A | #N/A | #N/A | #N/A | 1 | 0 | 1 | 1 | 0 | 1 | 0 | 1 | 1 | 0 | 1 | 0 | 0 | |
| 457 | M | 39-82 | | 2130 | 699 | 0 | 0 | 954 | 899 | 0 | 0 | 1 | 0 | 1 | 0 | 0 | 0 | 1 | 0 | 0 | 0 | 1 | 1 | 0 | |
| 458 | F | 39-82 | | 1934 | 723 | 0 | 0 | 2302 | 862 | 0 | 0 | 0 | 0 | 0 | 0 | 0 | 0 | 0 | 0 | 0 | 0 | 0 | 0 | 0 | |
| 459 | F | 39-82 | | 2594 | 489 | 0 | 0 | 1908 | 1800 | 0 | 0 | 0 | 0 | 0 | 0 | 0 | 0 | 0 | 0 | 0 | 0 | 0 | 0 | 0 | |
| 460 | F | 39-82 | trace igM unic | 1282 | 402 | 0 | 0 | 1978 | 1166 | 0 | 0 | 1 | 1 | 1 | 1 | 0 | 0 | 0 | 0 | 1 | 1 | 1 | 0 | 1 | 0 |
| 461 | F | 39-82 | | 1832 | 603 | 0 | 0 | #N/A | #N/A | #N/A | #N/A | 0 | 0 | 0 | 0 | 0 | 0 | 0 | 0 | 0 | 0 | 0 | 0 | 0 | |
| 462 | M | 19-38 | | 3330 | 862 | 0 | 0 | #N/A | #N/A | #N/A | #N/A | 0 | 0 | 0 | 0 | 0 | 0 | 0 | 0 | 0 | 0 | 0 | 0 | 0 | |
| 463 | F | 19-38 | | 5777 | 1442 | 0 | 0 | 3849 | 2822 | 0 | 0 | 0 | 0 | 0 | 0 | 0 | 0 | 0 | 0 | 0 | 0 | 0 | 0 | 0 | |
| 464 | F | 39-82 | | 1795 | 488 | 0 | 0 | #N/A | #N/A | #N/A | #N/A | 1 | 0 | 1 | 0 | 0 | 0 | 0 | 0 | 0 | 0 | 0 | 1 | 0 | |
| 465 | F | 39-82 | | 1172 | 698 | 0 | 0 | #N/A | #N/A | #N/A | #N/A | 0 | 0 | 0 | 0 | 0 | 0 | 0 | 0 | 0 | 0 | 0 | 0 | 0 | |
| 466 | F | 39-82 | | 3254 | 496 | 0 | 0 | 2074 | 1944 | 0 | 0 | 1 | 0 | 1 | 0 | 0 | 0 | 0 | 0 | 0 | 0 | 1 | 0 | 0 | |

| | | | | | | | | | | | | | | | | | | | | | | | | | |
|---|---|---|---|---|---|---|---|---|---|---|---|---|---|---|---|---|---|---|---|---|---|---|---|---|---|
| 467 | F | 19-38 | | 2465 | 623 | 0 | 0 | 1505 | 1094 | 0 | 0 | 0 | 0 | 0 | 0 | 0 | 0 | 0 | 0 | 0 | 0 | 0 | 0 | 0 | |
| 468 | F | 39-82 | | 3424 | 1262 | 0 | 0 | 2264 | 1256 | 0 | 0 | 1 | 0 | 0 | 0 | 1 | 1 | 0 | 0 | 0 | 0 | 1 | 0 | 0 | |
| 469 | M | 39-82 | | 1844 | 883 | 0 | 0 | 832 | 473 | 0 | 0 | 0 | 0 | 0 | 0 | 0 | 0 | 0 | 0 | 0 | 0 | 0 | 0 | 0 | |
| 470 | M | 19-38 | | 1540 | 960 | 0 | 0 | 1920 | 1244 | 0 | 0 | 1 | 1 | 1 | 1 | 0 | 0 | 0 | 0 | 0 | 0 | 1 | 0 | 0 | |
| 471 | F | 19-38 | | 3060 | 819 | 0 | 0 | #N/A | #N/A | #N/A | #N/A | 1 | 0 | 1 | 0 | 0 | 0 | 0 | 0 | 0 | 0 | 1 | 0 | 0 | |
| 472 | F | 39-82 | | 1874 | 1260 | 0 | 0 | 2289 | 1508 | 0 | 0 | 1 | 0 | 1 | 1 | 1 | 1 | 0 | 0 | 0 | 0 | 0 | 0 | 0 | |
| 473 | F | 39-82 | | 1062 | 247 | 0 | 0 | 1484 | 1118 | 0 | 0 | 1 | 0 | 1 | 1 | 0 | 0 | 1 | 0 | 1 | 0 | 1 | 0 | 0 | |
| 474 | F | 19-38 | | 1651 | 1403 | 0 | 0 | 1461 | 1887 | 0 | 0 | 0 | 0 | 0 | 0 | 0 | 0 | 0 | 0 | 0 | 0 | 0 | 0 | 0 | |
| 475 | F | 39-82 | | 6229 | 819 | 0 | 0 | 6344 | 783 | 0 | 0 | 1 | 0 | 1 | 1 | 0 | 0 | 0 | 1 | 1 | 0 | 1 | 1 | 0 | |
| 476 | F | 19-38 | | 3072 | 906 | 0 | 0 | 1687 | 977 | 0 | 0 | 1 | 0 | 1 | 0 | 0 | 0 | 0 | 0 | 0 | 0 | 0 | 0 | 0 | |
| 477 | F | 19-38 | IgM positive | 3771 | 1008 | 0 | 0 | 4900 | 1522 | 0 | 0 | 1 | 1 | 1 | 1 | 1 | 0 | 0 | 0 | 1 | 1 | 1 | 1 | 0 | |
| 478 | F | 19-38 | | 1765 | 1870 | 0 | 0 | 1981 | 2951 | 0 | 0 | 0 | 0 | 0 | 0 | 0 | 0 | 0 | 0 | 0 | 0 | 0 | 0 | 0 | |
| 479 | F | 19-38 | | 1352 | 2052 | 0 | 0 | #N/A | #N/A | #N/A | #N/A | 0 | 0 | 0 | 0 | 0 | 0 | 0 | 0 | 0 | 0 | 0 | 0 | 0 | |
| 480 | F | 19-38 | | 1359 | 723 | 0 | 0 | #N/A | #N/A | #N/A | #N/A | 0 | 0 | 0 | 0 | 0 | 0 | 0 | 0 | 0 | 0 | 0 | 0 | 0 | |
| 481 | F | 19-38 | | 2167 | 1486 | 0 | 0 | #N/A | #N/A | #N/A | #N/A | 1 | 0 | 1 | 1 | 0 | 0 | 0 | 0 | 1 | 0 | 0 | 0 | 0 | |
| 482 | F | 39-82 | | 1165 | 432 | 0 | 0 | 1554 | 841 | 0 | 0 | 1 | 0 | 1 | 0 | 0 | 0 | 0 | 0 | 0 | 0 | 0 | 0 | 0 | |
| 483 | F | 19-38 | | 841 | 390 | 0 | 0 | 1313 | 807 | 0 | 0 | 1 | 1 | 1 | 1 | 0 | 1 | 0 | 0 | 1 | 0 | 1 | 0 | 0 | |
| 484 | F | 19-38 | | 2011 | 1192 | 0 | 0 | #N/A | #N/A | #N/A | #N/A | 0 | 0 | 0 | 0 | 0 | 0 | 0 | 0 | 0 | 0 | 0 | 0 | 0 | |
| 485 | F | 39-82 | | 2060 | 779 | 0 | 0 | #N/A | #N/A | #N/A | #N/A | 1 | 0 | 0 | 0 | 0 | 0 | 0 | 0 | 1 | 0 | 0 | 1 | 0 | |
| 486 | F | 39-82 | | 2011 | 661 | 0 | 0 | 3462 | 669 | 0 | 0 | 1 | 0 | 0 | 0 | 0 | 0 | 0 | 0 | 0 | 0 | 1 | 0 | 0 | |
| 487 | F | 19-38 | | 1722 | 1325 | 0 | 0 | #N/A | #N/A | #N/A | #N/A | 1 | 0 | 1 | 0 | 0 | 0 | 0 | 0 | 0 | 0 | 1 | 0 | 0 | |
| 488 | F | 19-38 | | 1913 | 537 | 0 | 0 | 1588 | 523 | 0 | 0 | 1 | 0 | 1 | 0 | 0 | 0 | 1 | 0 | 0 | 0 | 1 | 0 | 1 | 0 |
| 489 | F | 39-82 | | 6084 | 3360 | 0 | 0 | 4059 | 2109 | 53 | 1 | 1 | 0 | 0 | 0 | 0 | 0 | 0 | 0 | 1 | 0 | 0 | 0 | 0 | |
| 490 | M | 39-82 | | 3380 | 650 | 0 | 0 | 4753 | 456 | 0 | 0 | 1 | 0 | 1 | 0 | 0 | 0 | 0 | 0 | 0 | 0 | 0 | 0 | 0 | |
| 491 | F | 39-82 | | 2604 | 542 | 0 | 0 | 3450 | 819 | 0 | 0 | 0 | 0 | 0 | 0 | 0 | 0 | 0 | 0 | 0 | 0 | 0 | 0 | 0 | |
| 492 | F | 39-82 | | 16358 | 752 | 0 | 1 | 13879 | 234 | 0 | 1 | 1 | 0 | 1 | 0 | 0 | 0 | 0 | 0 | 0 | 1 | 0 | 0 | 0 | |
| 493 | F | 39-82 | | 2512 | 498 | 0 | 0 | 1497 | 236 | 0 | 0 | 0 | 0 | 0 | 0 | 0 | 0 | 0 | 0 | 0 | 0 | 0 | 0 | 0 | |
| 494 | F | 39-82 | | 4545 | 537 | 0 | 0 | #N/A | #N/A | #N/A | #N/A | 1 | 0 | 1 | 1 | 0 | 0 | 0 | 0 | 0 | 0 | 1 | 0 | 0 | |
| 495 | M | 19-38 | negative | 2417 | 717 | 0 | 0 | #N/A | #N/A | #N/A | #N/A | 1 | 1 | 1 | 1 | 0 | 0 | 1 | 0 | 1 | 1 | 1 | 0 | 0 | |
| 497 | F | 19-38 | | 2034 | 599 | 0 | 0 | #N/A | #N/A | #N/A | #N/A | 1 | 0 | 1 | 1 | 0 | 0 | 0 | 1 | 1 | 0 | 1 | 1 | 0 | |
| 498 | M | 39-82 | | 1863 | 698 | 0 | 0 | 1983 | 1188 | 0 | 0 | 0 | 0 | 0 | 0 | 0 | 0 | 0 | 0 | 0 | 0 | 0 | 0 | 0 | |
| 499 | F | 19-38 | | 5267 | 788 | 0 | 0 | 7737 | 1840 | 0 | 0 | 1 | 0 | 1 | 0 | 0 | 0 | 1 | 0 | 0 | 0 | 1 | 1 | 0 | |
| 500 | F | 19-38 | IgG/IgM positi | 11698 | 21457 | 34 | 1 | #N/A | #N/A | #N/A | #N/A | 1 | 0 | 1 | 1 | 1 | 1 | 1 | 0 | 1 | 1 | 1 | 0 | 1 | 1 |
| 501 | M | 19-38 | | 4209 | 1919 | 0 | 0 | #N/A | #N/A | #N/A | #N/A | 0 | 0 | 0 | 0 | 0 | 0 | 0 | 0 | 0 | 0 | 0 | 0 | 0 | |
| 502 | F | 19-38 | | 2013 | 718 | 0 | 0 | #N/A | #N/A | #N/A | #N/A | 0 | 0 | 0 | 0 | 0 | 0 | 0 | 0 | 0 | 0 | 0 | 0 | 0 | |
| 503 | M | 19-38 | | 7909 | 3017 | 0 | 0 | #N/A | #N/A | #N/A | #N/A | 1 | 0 | 0 | 0 | 0 | 0 | 0 | 0 | 0 | 0 | 0 | 1 | 0 | |
| 504 | F | 19-38 | | 4333 | 952 | 0 | 0 | #N/A | #N/A | #N/A | #N/A | 0 | 0 | 0 | 0 | 0 | 0 | 0 | 0 | 0 | 0 | 0 | 0 | 0 | |
| 505 | F | 39-82 | | 7586 | 1027 | 0 | 0 | 8998 | 1749 | 0 | 0 | 1 | 1 | 1 | 0 | 0 | 0 | 1 | 0 | 1 | 0 | 0 | 0 | 1 | 0 |
| 506 | M | 39-82 | | 2522 | 512 | 0 | 0 | 1247 | 2050 | 0 | 0 | 1 | 0 | 1 | 0 | 0 | 0 | 0 | 0 | 1 | 0 | 1 | 0 | 0 | |
| 507 | F | 39-82 | | 3502 | 1157 | 0 | 0 | 1849 | 1200 | 0 | 0 | 1 | 1 | 0 | 0 | 0 | 0 | 0 | 0 | 0 | 0 | 1 | 1 | 0 | |
| 508 | F | 39-82 | | 1610 | 394 | 0 | 0 | 1345 | 776 | 0 | 0 | 0 | 0 | 0 | 0 | 0 | 0 | 0 | 0 | 0 | 0 | 0 | 0 | 0 | |
| 509 | F | 39-82 | | 7037 | 1101 | 0 | 0 | 4045 | 1479 | 0 | 0 | 0 | 0 | 0 | 0 | 0 | 0 | 0 | 0 | 0 | 0 | 0 | 0 | 0 | |
| 510 | M | 19-38 | | 4477 | 757 | 0 | 0 | #N/A | #N/A | #N/A | #N/A | 1 | 1 | 1 | 1 | 0 | 0 | 0 | 0 | 0 | 0 | 1 | 0 | 0 | |
| 511 | F | 39-82 | | 3790 | 1054 | 0 | 0 | 1123 | 1247 | 0 | 0 | 1 | 0 | 0 | 0 | 0 | 0 | 0 | 0 | 1 | 0 | 1 | 0 | 0 | |
| 512 | F | 19-38 | | 3007 | 1147 | 0 | 0 | 2766 | 3341 | 0 | 0 | 0 | 0 | 0 | 0 | 0 | 0 | 0 | 0 | 0 | 0 | 0 | 0 | 0 | |
| 513 | F | 19-38 | | 2129 | 359 | 0 | 0 | #N/A | #N/A | #N/A | #N/A | 0 | 0 | 0 | 0 | 0 | 0 | 0 | 0 | 0 | 0 | 0 | 0 | 0 | |
| 514 | F | 39-82 | | 2365 | 467 | 0 | 0 | 1325 | 1698 | 0 | 0 | 1 | 0 | 1 | 0 | 1 | 1 | 1 | 0 | 0 | 0 | 1 | 1 | 0 | |
| 515 | F | 39-82 | | 1784 | 618 | 0 | 0 | 3132 | 733 | 2 | 1 | 1 | 0 | 1 | 1 | 0 | 0 | 0 | 1 | 1 | 0 | 1 | 1 | 0 | |
| 516 | F | 39-82 | | 2435 | 554 | 0 | 0 | #N/A | #N/A | #N/A | #N/A | 0 | 0 | 0 | 0 | 0 | 0 | 0 | 0 | 0 | 0 | 0 | 0 | 0 | |
| 517 | M | 39-82 | | 2941 | 313 | 0 | 0 | #N/A | #N/A | #N/A | #N/A | 0 | 0 | 0 | 0 | 0 | 0 | 0 | 0 | 0 | 0 | 0 | 0 | 0 | |
| 518 | M | 39-82 | | 3265 | 433 | 0 | 0 | 1919 | 1880 | 0 | 0 | 0 | 0 | 0 | 0 | 0 | 0 | 0 | 0 | 0 | 0 | 0 | 0 | 0 | |
| 519 | F | 19-38 | | 8499 | 1774 | 0 | 0 | #N/A | #N/A | #N/A | #N/A | 1 | 0 | 1 | 0 | 0 | 0 | 1 | 0 | 0 | 0 | 1 | 0 | 0 | |
| 520 | F | 39-82 | | 2753 | 680 | 0 | 0 | 1617 | 1048 | 0 | 0 | 0 | 0 | 0 | 0 | 0 | 0 | 0 | 0 | 0 | 0 | 0 | 0 | 0 | |
| 521 | F | 39-82 | | 4968 | 1896 | 0 | 0 | 1063 | 1317 | 0 | 0 | 0 | 0 | 0 | 0 | 0 | 0 | 0 | 0 | 0 | 0 | 0 | 0 | 1 | 0 |
| 522 | F | 19-38 | | 5696 | 2655 | 0 | 0 | 3507 | 2197 | 0 | 0 | 1 | 1 | 1 | 1 | 1 | 1 | 1 | 0 | 0 | 0 | 1 | 1 | 0 | |
| 523 | F | 39-82 | | 4909 | 1216 | 0 | 0 | 3632 | 1612 | 0 | 0 | 0 | 0 | 0 | 0 | 0 | 0 | 0 | 0 | 0 | 0 | 0 | 0 | 0 | |
| 524 | F | 19-38 | | 2901 | 802 | 0 | 0 | 2184 | 1433 | 0 | 0 | 1 | 0 | 1 | 0 | 1 | 0 | 0 | 0 | 1 | 0 | 1 | 1 | 0 | |
| 525 | M | 39-82 | | 4449 | 5410 | 0 | 0 | #N/A | #N/A | #N/A | #N/A | 1 | 0 | 0 | 1 | 0 | 0 | 1 | 1 | 1 | 0 | 1 | 0 | 0 | |
| 526 | M | 19-38 | IgG/IgM positi | 27885 | 59700 | 91 | 1 | 34846 | 75982 | 0 | 1 | 1 | 1 | 1 | 0 | 0 | 1 | 1 | 1 | 0 | 1 | 0 | 0 | 1 | 1 |
| 527 | M | 19-38 | | 1503 | 688 | 0 | 0 | #N/A | #N/A | #N/A | #N/A | 0 | 0 | 0 | 0 | 0 | 0 | 0 | 0 | 0 | 0 | 0 | 0 | 0 | |
| 528 | F | 19-38 | | 3126 | 880 | 0 | 0 | #N/A | #N/A | #N/A | #N/A | 0 | 0 | 0 | 0 | 0 | 0 | 0 | 0 | 0 | 0 | 0 | 0 | 0 | |
| 529 | F | 39-82 | | 5380 | 553 | 0 | 0 | 4643 | 1091 | 0 | 0 | 1 | 1 | 0 | 0 | 0 | 0 | 0 | 0 | 0 | 0 | 0 | 0 | 0 | |
| 530 | F | 39-82 | | 2968 | 2132 | 0 | 0 | 2960 | 2850 | 0 | 0 | 0 | 0 | 0 | 0 | 0 | 0 | 0 | 0 | 0 | 0 | 0 | 0 | 0 | |
| 531 | F | 19-38 | | 3431 | 2892 | 0 | 0 | 6041 | 5762 | 0 | 0 | 0 | 0 | 0 | 0 | 0 | 0 | 0 | 0 | 0 | 0 | 0 | 0 | 0 | |
| 532 | F | 19-38 | | 3508 | 654 | 0 | 0 | 2059 | 964 | 0 | 0 | 1 | 0 | 0 | 0 | 0 | 0 | 0 | 0 | 0 | 0 | 1 | 0 | 0 | |
| 534 | M | 19-38 | | 2835 | 231 | 0 | 0 | 2440 | 1021 | 0 | 0 | 0 | 0 | 0 | 0 | 0 | 0 | 0 | 0 | 0 | 0 | 0 | 0 | 0 | |
| 535 | M | 19-38 | | 1485 | 474 | 0 | 0 | 1963 | 845 | 0 | 0 | 1 | 0 | 1 | 0 | 1 | 0 | 0 | 0 | 0 | 0 | 0 | 0 | 0 | |
| 536 | F | 39-82 | | 146998 | 52781 | 97 | 1 | 181153 | 95232 | 92 | 1 | 1 | 0 | 1 | 1 | 0 | 0 | 0 | 0 | 1 | 0 | 0 | 0 | 0 | |
| 537 | F | 19-38 | | 3853 | 638 | 0 | 0 | #N/A | #N/A | #N/A | #N/A | 1 | 0 | 1 | 0 | 0 | 0 | 0 | 0 | 1 | 0 | 0 | 0 | 0 | |
| 538 | F | 19-38 | | 2035 | 739 | 0 | 0 | 2145 | 1604 | 0 | 0 | 0 | 0 | 0 | 0 | 0 | 0 | 0 | 0 | 0 | 0 | 0 | 0 | 0 | |
| 539 | F | 39-82 | | 4154 | 501 | 0 | 0 | 1069 | 881 | 0 | 0 | 0 | 0 | 0 | 0 | 0 | 0 | 0 | 0 | 0 | 0 | 0 | 0 | 0 | |
| 540 | F | 19-38 | | 1057 | 392 | 0 | 0 | #N/A | #N/A | #N/A | #N/A | 1 | 0 | 1 | 0 | 1 | 0 | 1 | 0 | 1 | 0 | 1 | 0 | 1 | 0 |
| 541 | F | 19-38 | | 1106 | 443 | 0 | 0 | 603 | 620 | 0 | 0 | 0 | 0 | 0 | 0 | 0 | 0 | 0 | 0 | 0 | 0 | 0 | 0 | 0 | |
| 542 | F | 19-38 | negative | 6263 | 1680 | 0 | 0 | #N/A | #N/A | #N/A | #N/A | 1 | 0 | 1 | 0 | 0 | 0 | 1 | 0 | 1 | 1 | 1 | 0 | 1 | 0 |
| 543 | F | 19-38 | IgG/IgM positi | 46743 | 33924 | 89 | 1 | 34755 | 56378 | 77 | 1 | 1 | 1 | 1 | 1 | 1 | 1 | 0 | 1 | 0 | 1 | 1 | 1 | 1 | 1 |
| 544 | F | 19-38 | | 1043 | 1276 | 0 | 0 | #N/A | #N/A | #N/A | #N/A | 0 | 0 | 0 | 0 | 0 | 0 | 0 | 0 | 0 | 0 | 0 | 0 | 0 | |
| 546 | F | 39-82 | | 1819 | 413 | 0 | 0 | 1452 | 1147 | 0 | 0 | 0 | 0 | 0 | 0 | 0 | 0 | 0 | 0 | 0 | 0 | 0 | 0 | 0 | |

| | | | | | | | | | | | | | | | | | | | | | | | | | |
|---|---|---|---|---|---|---|---|---|---|---|---|---|---|---|---|---|---|---|---|---|---|---|---|---|---|
| 547 | F | 19-38 | | 62242 | 26170 | 40 | 1 | #N/A | #N/A | #N/A | #N/A | 1 | 0 | 0 | 1 | 0 | 0 | 1 | 0 | 1 | 1 | 1 | 0 | 1 | 0 |
| 548 | F | 19-38 | | 1697 | 677 | 0 | 0 | 1184 | 1912 | 0 | 0 | 0 | 0 | 0 | 1 | 0 | 0 | 0 | 0 | 0 | 0 | 0 | 0 | 1 | 0 |
| 549 | F | 19-38 | | 5135 | 3463 | 0 | 0 | 6647 | 5514 | 0 | 0 | 1 | 0 | 1 | 0 | 0 | 0 | 0 | 0 | 0 | 0 | 1 | 1 | 0 | |
| 550 | M | 19-38 | | 21993 | 9451 | 0 | 1 | 14531 | 22031 | 0 | 1 | 1 | 0 | 0 | 0 | 0 | 0 | 0 | 0 | 1 | 0 | 1 | 0 | 0 | |
| 551 | F | 39-82 | | 4350 | 796 | 0 | 0 | 1834 | 1019 | 0 | 0 | 0 | 0 | 0 | 0 | 0 | 0 | 0 | 0 | 0 | 0 | 0 | 0 | 0 | |
| 552 | M | 19-38 | | 2118 | 624 | 0 | 0 | 1121 | 690 | 0 | 0 | 1 | 0 | 1 | 0 | 0 | 0 | 1 | 0 | 0 | 0 | 0 | 0 | 0 | |
| 553 | M | 39-82 | | 989 | 1031 | 0 | 0 | 1006 | 1165 | 0 | 0 | 1 | 1 | 0 | 1 | 0 | 0 | 0 | 0 | 0 | 0 | 0 | 0 | 0 | |
| 554 | F | 19-38 | | 1847 | 1057 | 0 | 0 | 2451 | 1340 | 0 | 0 | 1 | 0 | 0 | 0 | 0 | 0 | 1 | 0 | 0 | 0 | 1 | 0 | 0 | |
| 555 | F | 39-82 | IgG/IgM positi | 95001 | 47615 | 85 | 1 | 54128 | 25084 | 77 | 1 | 1 | 0 | 1 | 0 | 1 | 1 | 1 | 0 | 0 | 1 | 1 | 0 | 1 | 1 |
| 556 | f | 19-38 | | 4573 | 2083 | 0 | 0 | #N/A | #N/A | #N/A | #N/A | 1 | 0 | 1 | 0 | 0 | 0 | 0 | 0 | 0 | 0 | 1 | 0 | 0 | |
| 557 | F | 19-38 | | 2262 | 1978 | 0 | 0 | #N/A | #N/A | #N/A | #N/A | 0 | 0 | 0 | 0 | 0 | 0 | 0 | 0 | 0 | 0 | 0 | 0 | 0 | |
| 558 | F | 39-82 | | 97111 | 47767 | 55 | 1 | #N/A | #N/A | #N/A | #N/A | 1 | 0 | 1 | 0 | 0 | 0 | 0 | 0 | 1 | 1 | 0 | 0 | 0 | |
| 559 | F | 19-38 | | 1433 | 2001 | 0 | 0 | #N/A | #N/A | #N/A | #N/A | 1 | 0 | 1 | 0 | 0 | 0 | 0 | 0 | 0 | 0 | 0 | 0 | 0 | |
| 560 | F | 19-38 | | 2034 | 798 | 0 | 0 | 4450 | 571 | 0 | 0 | 1 | 0 | 1 | 0 | 0 | 0 | 0 | 0 | 0 | 0 | 0 | 0 | 0 | |
| 561 | F | 39-82 | | 19965 | 54292 | 88 | 1 | #N/A | #N/A | #N/A | #N/A | 1 | 1 | 1 | 1 | 1 | 0 | 1 | 0 | 1 | 1 | 1 | 1 | 0 | |
| 562 | F | 19-38 | | 3231 | 383 | 0 | 0 | 1270 | 926 | 0 | 0 | 1 | 0 | 0 | 0 | 0 | 0 | 0 | 0 | 1 | 0 | 1 | 1 | 1 | 0 |
| 563 | F | 19-38 | | 2000 | 743 | 0 | 0 | 3299 | 1429 | 0 | 0 | 0 | 0 | 0 | 0 | 0 | 0 | 0 | 0 | 0 | 0 | 0 | 0 | 0 | |
| 564 | F | 39-82 | IgG/IgM positi | 19953 | 16924 | 45 | 1 | 9296 | 13619 | 0 | 1 | 1 | 0 | 1 | 0 | 0 | 0 | 0 | 1 | 1 | 1 | 0 | 0 | 0 | |
| 565 | F | 19-38 | | 1399 | 716 | 0 | 0 | 999 | 286 | 0 | 0 | 0 | 0 | 0 | 0 | 0 | 0 | 0 | 0 | 0 | 0 | 0 | 0 | 0 | |
| 566 | M | 19-38 | | 2005 | 599 | 0 | 0 | 753 | 697 | 0 | 0 | 0 | 0 | 0 | 0 | 0 | 0 | 0 | 0 | 0 | 0 | 0 | 0 | 0 | |
| 567 | F | 19-38 | | 1002 | 966 | 0 | 0 | 834 | 1088 | 0 | 0 | 1 | 1 | 1 | 1 | 1 | 0 | 0 | 0 | 0 | 0 | 1 | 1 | 0 | |
| 568 | F | 39-82 | | 928 | 964 | 0 | 0 | 1932 | 2904 | 0 | 0 | 0 | 0 | 0 | 0 | 0 | 0 | 0 | 0 | 0 | 0 | 0 | 0 | 0 | |
| 569 | F | 19-38 | | 807 | 476 | 0 | 0 | #N/A | #N/A | #N/A | #N/A | 1 | 1 | 1 | 0 | 0 | 0 | 0 | 0 | 0 | 0 | 1 | 0 | 0 | |
| 570 | F | 39-82 | | 1585 | 679 | 0 | 0 | 1344 | 1151 | 0 | 0 | 0 | 0 | 0 | 0 | 0 | 0 | 0 | 0 | 0 | 0 | 0 | 0 | 0 | |
| 571 | F | 39-82 | | 32716 | 60426 | 78 | 1 | 18017 | 47705 | 85 | 1 | 1 | 0 | 0 | 0 | 0 | 0 | 0 | 0 | 1 | 1 | 0 | 0 | 0 | |
| 572 | F | 19-38 | | 26371 | 15796 | 2 | 1 | 36987 | 27909 | 13 | 1 | 1 | 0 | 1 | 0 | 0 | 0 | 1 | 0 | 0 | 1 | 1 | 1 | 1 | 1 |
| 573 | F | 39-82 | | 1766 | 389 | 0 | 0 | #N/A | #N/A | #N/A | #N/A | 0 | 0 | 0 | 0 | 0 | 0 | 0 | 0 | 0 | 0 | 0 | 0 | 0 | |
| 574 | F | 19-38 | | 3581 | 1294 | 0 | 0 | #N/A | #N/A | #N/A | #N/A | 1 | 0 | 1 | 1 | 1 | 1 | 1 | 0 | 1 | 0 | 1 | 0 | 0 | |
| 575 | F | 19-38 | | 3873 | 2019 | 0 | 0 | #N/A | #N/A | #N/A | #N/A | 1 | 0 | 1 | 1 | 0 | 0 | 0 | 0 | 0 | 0 | 0 | 0 | 0 | |
| 576 | F | 39-82 | negative | 1610 | 641 | 0 | 0 | #N/A | #N/A | #N/A | #N/A | 1 | 0 | 1 | 0 | 0 | 0 | 1 | 0 | 1 | 1 | 1 | 1 | 0 | |
| 577 | F | 39-82 | | 4142 | 6382 | 0 | 0 | #N/A | #N/A | #N/A | #N/A | 0 | 0 | 0 | 0 | 0 | 0 | 0 | 0 | 0 | 0 | 0 | 0 | 0 | |
| 578 | F | 19-38 | | 6359 | 1173 | 0 | 0 | 2621 | 1442 | 0 | 0 | 1 | 0 | 1 | 1 | 0 | 1 | 0 | 0 | 1 | 0 | 0 | 0 | 0 | |
| 579 | F | 19-38 | | 4681 | 1702 | 0 | 0 | 3269 | 2079 | 0 | 0 | 1 | 0 | 1 | 0 | 0 | 0 | 0 | 0 | 0 | 0 | 1 | 0 | 0 | |
| 580 | F | 39-82 | | 110055 | 105382 | 85 | 1 | 66866 | 82251 | 37 | 1 | 1 | 1 | 1 | 0 | 1 | 1 | 1 | 0 | 0 | 1 | 1 | 0 | 1 | 1 |
| 581 | F | 19-38 | | 4931 | 994 | 0 | 0 | 1318 | 1656 | 0 | 0 | 0 | 0 | 0 | 0 | 0 | 0 | 0 | 0 | 0 | 0 | 0 | 0 | 0 | |
| 582 | F | 39-82 | | 9310 | 3359 | 0 | 0 | 2044 | 1787 | 0 | 0 | 1 | 0 | 1 | 1 | 0 | 0 | 0 | 0 | 0 | 0 | 1 | 0 | 1 | 0 |
| 583 | F | 39-82 | negative | 5135 | 1405 | 0 | 0 | 1242 | 725 | 0 | 0 | 0 | 0 | 0 | 0 | 0 | 0 | 0 | 0 | 0 | 0 | 0 | 0 | 0 | |
| 584 | M | 19-38 | negative | 3218 | 829 | 30 | 1 | 406 | 312 | 0 | 0 | 0 | 0 | 0 | 1 | 0 | 0 | 0 | 0 | 0 | 0 | 0 | 0 | 0 | |
| 585 | M | 39-82 | | 697 | 436 | 0 | 0 | 1522 | 973 | 0 | 0 | 0 | 0 | 0 | 0 | 0 | 0 | 0 | 0 | 0 | 0 | 0 | 0 | 0 | |
| 586 | F | 39-82 | negative | 2998 | 955 | 0 | 0 | 1797 | 1330 | 0 | 0 | 1 | 0 | 1 | 1 | 1 | 1 | 0 | 0 | 1 | 1 | 1 | 0 | 1 | 0 |
| 587 | F | 19-38 | | 5880 | 884 | 0 | 0 | 2811 | 1354 | 0 | 0 | 1 | 0 | 1 | 0 | 0 | 0 | 0 | 0 | 0 | 0 | 1 | 1 | 0 | |
| 588 | M | 39-82 | | 6263 | 1840 | 0 | 0 | #N/A | #N/A | #N/A | #N/A | 0 | 0 | 0 | 0 | 0 | 0 | 0 | 0 | 0 | 0 | 0 | 0 | 0 | |
| 589 | M | 19-38 | | 2759 | 868 | 0 | 0 | #N/A | #N/A | #N/A | #N/A | 1 | 1 | 1 | 0 | 0 | 0 | 0 | 0 | 0 | 0 | 1 | 0 | 0 | |
| 590 | F | 39-82 | | 3187 | 2017 | 0 | 0 | 2533 | 1468 | 0 | 0 | 1 | 0 | 0 | 0 | 0 | 0 | 0 | 0 | 1 | 0 | 1 | 0 | 1 | 0 |
| 591 | M | 19-38 | | 571 | 550 | 0 | 0 | #N/A | #N/A | #N/A | #N/A | 1 | 0 | 0 | 0 | 0 | 1 | 0 | 0 | 0 | 0 | 0 | 1 | 1 | 0 |
| 592 | F | 19-38 | | 4025 | 1165 | 0 | 0 | 2979 | 805 | 0 | 0 | 1 | 1 | 1 | 1 | 0 | 0 | 1 | 0 | 1 | 0 | 1 | 0 | 0 | |
| 593 | F | 39-82 | | 6497 | 804 | 0 | 0 | 7002 | 1658 | 0 | 0 | 0 | 0 | 0 | 0 | 0 | 0 | 0 | 0 | 0 | 0 | 0 | 0 | 0 | |
| 594 | F | 19-38 | | 808 | 662 | 0 | 0 | 448 | 787 | 0 | 0 | 1 | 1 | 1 | 1 | 1 | 1 | 1 | 0 | 1 | 0 | 1 | 1 | 0 | |
| 595 | M | 19-38 | | 4268 | 665 | 0 | 0 | 1206 | 576 | 0 | 0 | 1 | 0 | 1 | 1 | 0 | 0 | 0 | 0 | 1 | 0 | 1 | 1 | 0 | |
| 596 | M | 39-82 | | 5302 | 920 | 0 | 0 | #N/A | #N/A | #N/A | #N/A | 1 | 0 | 1 | 1 | 1 | 0 | 1 | 0 | 0 | 0 | 1 | 0 | 0 | |
| 597 | F | 19-38 | | 3597 | 4041 | 0 | 0 | #N/A | #N/A | #N/A | #N/A | 1 | 0 | 1 | 0 | 0 | 0 | 0 | 0 | 0 | 0 | 1 | 1 | 0 | |
| 598 | F | 19-38 | | 1656 | 663 | 0 | 0 | 2108 | 742 | 0 | 0 | 1 | 0 | 1 | 1 | 0 | 0 | 1 | 1 | 1 | 0 | 1 | 0 | 0 | |
| 599 | F | 39-82 | | 2489 | 1518 | 0 | 0 | 763 | 975 | 0 | 0 | 0 | 0 | 0 | 0 | 0 | 0 | 0 | 0 | 0 | 0 | 0 | 0 | 0 | |
| 600 | F | 39-82 | | 5009 | 1877 | 0 | 0 | 4083 | 1612 | 0 | 0 | 0 | 0 | 0 | 0 | 0 | 0 | 0 | 0 | 0 | 0 | 0 | 0 | 0 | |
| 601 | F | 39-82 | | 7994 | 2217 | 0 | 0 | #N/A | #N/A | #N/A | #N/A | 1 | 0 | 1 | 0 | 0 | 0 | 0 | 0 | 1 | 0 | 1 | 0 | 1 | 0 |
| 602 | F | 39-82 | | 9486 | 1660 | 0 | 0 | 6720 | 1712 | 0 | 0 | 1 | 0 | 1 | 0 | 0 | 0 | 1 | 0 | 0 | 0 | 1 | 0 | 0 | |
| 603 | F | 39-82 | | 4565 | 688 | 0 | 0 | #N/A | #N/A | #N/A | #N/A | 1 | 0 | 1 | 1 | 0 | 0 | 0 | 0 | 0 | 0 | 1 | 1 | 0 | |
| 604 | F | 39-82 | | 8733 | 952 | 0 | 0 | 4934 | 641 | 0 | 0 | 1 | 0 | 1 | 0 | 0 | 0 | 0 | 0 | 0 | 0 | 1 | 0 | 0 | |
| 605 | F | 19-38 | | 2473 | 445 | 0 | 0 | 1819 | 531 | 0 | 0 | 0 | 0 | 0 | 0 | 0 | 0 | 0 | 0 | 0 | 0 | 0 | 0 | 0 | |
| 606 | F | 19-38 | | 38893 | 73484 | 79 | 1 | 17273 | 61972 | 70 | 1 | 1 | 0 | 0 | 1 | 0 | 0 | 1 | 0 | 0 | 1 | 0 | 0 | 1 | 1 |
| 607 | F | 39-82 | | 75217 | 63306 | 93 | 1 | 24423 | 37738 | 7 | 1 | 1 | 1 | 1 | 0 | 0 | 0 | 1 | 0 | 0 | 0 | 1 | 1 | 0 | |
| 608 | M | 19-38 | | 804 | 728 | 0 | 0 | #N/A | #N/A | #N/A | #N/A | 0 | 0 | 0 | 0 | 0 | 0 | 0 | 0 | 0 | 0 | 0 | 0 | 0 | |
| 609 | F | 19-38 | | 30065 | 1189 | 0 | 1 | #N/A | #N/A | #N/A | #N/A | 0 | 0 | 0 | 0 | 0 | 0 | 0 | 0 | 0 | 0 | 0 | 0 | 0 | |
| 610 | F | 19-38 | | 2421 | 635 | 0 | 0 | 1012 | 651 | 0 | 0 | 1 | 0 | 1 | 0 | 1 | 0 | 1 | 0 | 0 | 0 | 1 | 1 | 1 | 0 |
| 611 | F | 19-38 | | 6512 | 4655 | 0 | 0 | 6789 | 4388 | 0 | 0 | 1 | 0 | 1 | 1 | 0 | 0 | 0 | 1 | 1 | 0 | 1 | 0 | 0 | |
| 612 | F | 19-38 | | 5845 | 749 | 0 | 0 | 2719 | 1209 | 0 | 0 | 0 | 0 | 0 | 0 | 0 | 0 | 0 | 0 | 0 | 0 | 0 | 0 | 0 | |
| 613 | F | 19-38 | | 2873 | 1565 | 0 | 0 | 2193 | 918 | 0 | 0 | 1 | 0 | 1 | 1 | 0 | 0 | 0 | 0 | 1 | 0 | 1 | 0 | 0 | |
| 614 | M | 39-82 | | 4820 | 4948 | 0 | 0 | #N/A | #N/A | #N/A | #N/A | 0 | 0 | 0 | 0 | 0 | 0 | 0 | 0 | 0 | 0 | 0 | 0 | 0 | |
| 615 | F | 39-82 | | 1572 | 1143 | 0 | 0 | 1424 | 2194 | 0 | 0 | 0 | 0 | 0 | 0 | 0 | 0 | 0 | 0 | 0 | 0 | 0 | 0 | 0 | |
| 616 | M | 39-82 | | 1686 | 487 | 0 | 0 | 976 | 1224 | 0 | 0 | 0 | 0 | 0 | 0 | 0 | 0 | 0 | 0 | 0 | 0 | 0 | 0 | 0 | |
| 617 | F | 19-38 | | 144548 | 209350 | 98 | 1 | 78465 | 161609 | 18 | 1 | 1 | 1 | 1 | 0 | 0 | 0 | 1 | 0 | 0 | 1 | 0 | 0 | 1 | 1 |
| 618 | F | 19-38 | | 2368 | 1584 | 0 | 0 | #N/A | #N/A | #N/A | #N/A | 1 | 0 | 1 | 1 | 1 | 1 | 0 | 0 | 0 | 0 | 1 | 1 | 0 | |
| 619 | F | 19-38 | | 11640 | 3595 | 0 | 1 | 10999 | 2921 | 3 | 1 | 0 | 0 | 0 | 0 | 0 | 0 | 0 | 0 | 0 | 0 | 0 | 0 | 0 | |
| 620 | F | 19-38 | | 10621 | 11447 | 20 | 1 | 13188 | 18242 | 27 | 1 | 1 | 0 | 1 | 0 | 0 | 0 | 0 | 0 | 0 | 1 | 0 | 1 | 1 | 1 |
| 621 | F | 19-38 | | 5050 | 1159 | 0 | 0 | #N/A | #N/A | #N/A | #N/A | 0 | 0 | 0 | 0 | 0 | 0 | 0 | 0 | 0 | 0 | 0 | 0 | 0 | |
| 622 | F | 19-38 | | 4652 | 974 | 0 | 0 | 2011 | 913 | 0 | 0 | 0 | 0 | 0 | 0 | 0 | 0 | 0 | 0 | 0 | 0 | 0 | 0 | 0 | |
| 623 | F | 19-38 | | 3984 | 2959 | 0 | 0 | 5533 | 1612 | 0 | 0 | 0 | 0 | 0 | 0 | 0 | 0 | 0 | 0 | 0 | 0 | 0 | 0 | 0 | |

| | | | | | | | | | | | | | | | | | | | | | | | | | |
|---|---|---|---|---|---|---|---|---|---|---|---|---|---|---|---|---|---|---|---|---|---|---|---|---|---|
| 624 | F | 19-38 | | 2439 | 1775 | 0 | 0 | 2231 | 1328 | 0 | 0 | 1 | 0 | 1 | 0 | 0 | 0 | 0 | 0 | 0 | 0 | 1 | 0 | 0 | |
| 625 | M | 19-38 | | 6245 | 1149 | 0 | 0 | #N/A | #N/A | #N/A | #N/A | 0 | 0 | 0 | 0 | 0 | 0 | 0 | 0 | 0 | 0 | 0 | 0 | 0 | |
| 626 | F | 39-82 | | 1686 | 2461 | 0 | 0 | 2219 | 1278 | 0 | 0 | 0 | 0 | 0 | 0 | 0 | 0 | 0 | 0 | 0 | 0 | 0 | 0 | 0 | |
| 627 | F | 19-38 | | 1595 | 1871 | 0 | 0 | 3670 | 1866 | 0 | 0 | 0 | 0 | 0 | 0 | 0 | 0 | 0 | 0 | 0 | 0 | 0 | 0 | 0 | |
| 628 | M | 39-82 | | 2691 | 448 | 0 | 0 | 1046 | 692 | 0 | 0 | 1 | 0 | 0 | 0 | 0 | 0 | 0 | 0 | 1 | 0 | 0 | 0 | 0 | |
| 629 | M | 39-82 | | 1386 | 2662 | 0 | 0 | #N/A | #N/A | #N/A | #N/A | 1 | 1 | 1 | 1 | 0 | 0 | 0 | 0 | 1 | 0 | 0 | 1 | 0 | |
| 630 | M | 39-82 | | 3994 | 3302 | 0 | 0 | 4729 | 6182 | 0 | 0 | 0 | 0 | 0 | 0 | 0 | 0 | 0 | 0 | 0 | 0 | 0 | 0 | 0 | |
| 631 | M | 39-82 | | 43894 | 23211 | 93 | 1 | 11076 | 20487 | 38 | 1 | 1 | 1 | 0 | 0 | 0 | 0 | 1 | 0 | 0 | 0 | 0 | 0 | 1 | 1 |
| 632 | F | 39-82 | | 1270 | 894 | 0 | 0 | #N/A | #N/A | #N/A | #N/A | 1 | 0 | 1 | 0 | 1 | 0 | 0 | 0 | 1 | 0 | 0 | 0 | 0 | |
| 633 | F | 19-38 | | 47560 | 141683 | 95 | 1 | 21984 | 96397 | 14 | 1 | 0 | 0 | 0 | 0 | 0 | 0 | 0 | 0 | 0 | 0 | 0 | 0 | 0 | |
| 634 | F | 19-38 | | 3349 | 590 | 0 | 0 | #N/A | #N/A | #N/A | #N/A | 0 | 0 | 0 | 0 | 0 | 0 | 0 | 0 | 0 | 0 | 0 | 0 | 0 | |
| 635 | F | 39-82 | | 1744 | 557 | 0 | 0 | 4638 | 557 | 0 | 0 | 0 | 0 | 0 | 0 | 0 | 0 | 0 | 0 | 0 | 0 | 0 | 0 | 0 | |
| 636 | F | 19-38 | | 23758 | 42708 | 46 | 1 | 15570 | 36421 | 2 | 1 | 1 | 1 | 1 | 1 | 0 | 1 | 0 | 0 | 0 | 1 | 1 | 1 | 1 | 1 |
| 637 | F | 19-38 | | 1800 | 719 | 0 | 0 | #N/A | #N/A | #N/A | #N/A | 1 | 0 | 1 | 0 | 0 | 0 | 0 | 0 | 1 | 0 | 1 | 0 | 0 | |
| 638 | F | 19-38 | negative | 1908 | 679 | 0 | 0 | 1193 | 807 | 0 | 0 | 1 | 0 | 1 | 1 | 0 | 0 | 0 | 1 | 0 | 1 | 0 | 0 | 1 | 0 |
| 639 | F | 19-38 | | 4839 | 510 | 0 | 0 | 2042 | 629 | 0 | 0 | 0 | 0 | 0 | 0 | 0 | 0 | 0 | 0 | 0 | 0 | 0 | 0 | 0 | |
| 640 | F | 39-82 | | 1907 | 917 | 0 | 0 | 1809 | 1316 | 0 | 0 | 1 | 0 | 1 | 0 | 1 | 1 | 0 | 0 | 0 | 0 | 1 | 1 | 1 | 0 |
| 641 | F | 19-38 | | 2575 | 1453 | 0 | 0 | 1219 | 488 | 0 | 0 | 1 | 0 | 0 | 0 | 0 | 0 | 0 | 0 | 0 | 0 | 1 | 0 | 0 | |
| 642 | F | 19-38 | | 4288 | 885 | 0 | 0 | 2305 | 1401 | 0 | 0 | 1 | 0 | 0 | 0 | 0 | 0 | 0 | 0 | 0 | 0 | 0 | 1 | 0 | |
| 643 | F | 39-82 | | 3610 | 1014 | 0 | 0 | 760 | 413 | 0 | 0 | 1 | 0 | 0 | 0 | 0 | 0 | 1 | 0 | 0 | 0 | 1 | 0 | 0 | |
| 644 | F | 19-38 | | 2850 | 2025 | 0 | 0 | 1091 | 597 | 0 | 0 | 1 | 1 | 1 | 0 | 0 | 0 | 1 | 0 | 0 | 0 | 1 | 1 | 0 | |
| 645 | F | 19-38 | | 1107 | 1065 | 0 | 0 | 1011 | 1368 | 0 | 0 | 1 | 0 | 1 | 1 | 0 | 0 | 0 | 0 | 0 | 0 | 1 | 1 | 0 | |
| 646 | F | 19-38 | negative | 4302 | 820 | 0 | 0 | 1801 | 899 | 0 | 0 | 1 | 0 | 0 | 1 | 0 | 0 | 0 | 0 | 1 | 1 | 1 | 0 | 0 | |
| 647 | F | 19-38 | | 5034 | 3695 | 0 | 0 | 1546 | 2836 | 0 | 0 | 0 | 0 | 0 | 0 | 0 | 0 | 0 | 0 | 0 | 0 | 0 | 0 | 0 | |
| 648 | M | 19-38 | | 22061 | 48793 | 85 | 1 | 20540 | 55355 | 86 | 1 | 1 | 1 | 1 | 0 | 0 | 0 | 1 | 0 | 1 | 0 | 1 | 0 | 0 | |
| 649 | F | 19-38 | | 4190 | 1213 | 0 | 0 | 764 | 530 | 0 | 0 | 1 | 0 | 0 | 0 | 0 | 0 | 0 | 0 | 0 | 0 | 1 | 0 | 0 | |
| 650 | F | 19-38 | | 2864 | 890 | 0 | 0 | #N/A | #N/A | #N/A | #N/A | 1 | 0 | 0 | 0 | 0 | 0 | 0 | 0 | 1 | 0 | 1 | 1 | 0 | |
| 651 | F | 39-82 | | 1528 | 4951 | 0 | 0 | 710 | 1971 | 0 | 0 | 1 | 0 | 1 | 0 | 0 | 0 | 0 | 1 | 0 | 0 | 1 | 0 | 0 | |
| 652 | F | 19-38 | | 3837 | 575 | 0 | 0 | 1412 | 1934 | 0 | 0 | 1 | 0 | 1 | 0 | 0 | 0 | 0 | 0 | 0 | 0 | 1 | 0 | 0 | |
| 653 | M | 19-38 | | 2127 | 404 | 0 | 0 | 839 | 197 | 0 | 0 | 1 | 0 | 0 | 0 | 0 | 0 | 0 | 0 | 0 | 0 | 1 | 0 | 0 | |
| 654 | F | 39-82 | | 7549 | 542 | 0 | 0 | 2714 | 693 | 0 | 0 | 0 | 0 | 0 | 0 | 0 | 0 | 0 | 0 | 0 | 0 | 0 | 0 | 0 | |
| 655 | F | 39-82 | | 6660 | 1169 | 0 | 0 | 7378 | 1547 | 0 | 0 | 1 | 0 | 0 | 0 | 1 | 1 | 0 | 1 | 0 | 0 | 1 | 1 | 0 | |
| 656 | F | 19-38 | | 2874 | 1385 | 0 | 0 | 2337 | 1188 | 0 | 0 | 0 | 0 | 0 | 0 | 0 | 0 | 0 | 0 | 0 | 0 | 0 | 0 | 0 | |
| 657 | F | 39-82 | | 1136 | 455 | 0 | 0 | #N/A | #N/A | #N/A | #N/A | 0 | 0 | 0 | 0 | 0 | 0 | 0 | 0 | 0 | 0 | 0 | 0 | 0 | |
| 658 | F | 39-82 | | 1722 | 639 | 0 | 0 | #N/A | #N/A | #N/A | #N/A | 0 | 0 | 0 | 0 | 0 | 0 | 0 | 0 | 0 | 0 | 0 | 0 | 0 | |
| 659 | F | 19-38 | | 1369 | 806 | 0 | 0 | 733 | 482 | 0 | 0 | 0 | 0 | 0 | 0 | 0 | 0 | 0 | 0 | 0 | 0 | 0 | 0 | 0 | |
| 660 | F | 19-38 | | 3030 | 3406 | 0 | 0 | 4156 | 2058 | 0 | 0 | 1 | 0 | 1 | 0 | 0 | 1 | 1 | 0 | 0 | 0 | 1 | 0 | 0 | |
| 661 | F | 19-38 | | 844 | 578 | 0 | 0 | 1094 | 589 | 0 | 0 | 1 | 1 | 1 | 0 | 1 | 1 | 1 | 0 | 0 | 0 | 1 | 1 | 1 | 0 |
| 662 | F | 39-82 | | 744 | 776 | 0 | 0 | 2359 | 1040 | 0 | 0 | 0 | 0 | 0 | 0 | 0 | 0 | 0 | 0 | 0 | 0 | 0 | 0 | 0 | |
| 663 | M | 19-38 | | 1182 | 963 | 0 | 0 | #N/A | #N/A | #N/A | #N/A | 0 | 0 | 0 | 0 | 0 | 0 | 0 | 0 | 0 | 0 | 0 | 0 | 0 | |
| 664 | F | 39-82 | | 5474 | 1317 | 0 | 0 | 3212 | 2522 | 0 | 0 | 0 | 0 | 0 | 0 | 0 | 0 | 0 | 0 | 0 | 0 | 0 | 0 | 0 | |
| 665 | F | 39-82 | negative | 3335 | 4270 | 0 | 0 | #N/A | #N/A | #N/A | #N/A | 1 | 1 | 1 | 0 | 0 | 1 | 1 | 1 | 0 | 1 | 1 | 0 | 0 | |
| 666 | F | 39-82 | | 2238 | 476 | 0 | 0 | 4813 | 735 | 2 | 1 | 1 | 0 | 0 | 0 | 0 | 0 | 0 | 0 | 0 | 0 | 1 | 0 | 0 | |
| 667 | F | 19-38 | | 2259 | 1006 | 0 | 0 | #N/A | #N/A | #N/A | #N/A | 1 | 0 | 1 | 0 | 0 | 0 | 0 | 0 | 1 | 0 | 1 | 0 | 0 | |
| 668 | F | 19-38 | | 3744 | 4135 | 0 | 0 | 2222 | 1381 | 0 | 0 | 0 | 0 | 0 | 0 | 0 | 0 | 0 | 0 | 0 | 0 | 0 | 0 | 0 | |
| 669 | M | 39-82 | | 491 | 1682 | 0 | 0 | 224 | 278 | 0 | 0 | 0 | 0 | 0 | 0 | 0 | 0 | 0 | 0 | 0 | 0 | 0 | 0 | 0 | |
| 670 | F | 39-82 | | 1952 | 410 | 0 | 0 | 1461 | 546 | 0 | 0 | 0 | 0 | 0 | 0 | 0 | 0 | 0 | 0 | 0 | 0 | 0 | 0 | 0 | |
| 671 | M | 19-38 | | 3665 | 3978 | 0 | 0 | 7754 | 2285 | 0 | 0 | 0 | 0 | 0 | 0 | 0 | 0 | 0 | 0 | 0 | 0 | 0 | 0 | 0 | |
| 672 | F | 39-82 | | 2356 | 2567 | 0 | 0 | 1895 | 1888 | 0 | 0 | 0 | 0 | 0 | 0 | 0 | 0 | 0 | 0 | 0 | 0 | 0 | 0 | 0 | |
| 673 | F | 39-82 | | 4104 | 2343 | 0 | 0 | 2696 | 1354 | 0 | 0 | 0 | 0 | 0 | 0 | 0 | 0 | 0 | 0 | 0 | 0 | 0 | 0 | 1 | |
| 674 | F | 19-38 | | 18251 | 53897 | 82 | 1 | 12418 | 40410 | 32 | 1 | 1 | 0 | 1 | 1 | 0 | 0 | 0 | 0 | 1 | 1 | 1 | 0 | 0 | |
| 675 | F | 19-38 | | 2676 | 2109 | 0 | 0 | 4116 | 1686 | 0 | 0 | 0 | 0 | 0 | 0 | 0 | 0 | 0 | 0 | 0 | 0 | 0 | 0 | 0 | |
| 676 | F | 19-38 | | 1570 | 1636 | 0 | 0 | 1579 | 1614 | 0 | 0 | 0 | 0 | 0 | 0 | 0 | 0 | 0 | 0 | 0 | 0 | 0 | 0 | 0 | |
| 677 | F | 39-82 | | 3027 | 406 | 0 | 0 | 3182 | 915 | 0 | 0 | 0 | 0 | 0 | 0 | 0 | 0 | 0 | 0 | 0 | 0 | 0 | 0 | 0 | |
| 678 | F | 19-38 | | 2301 | 2225 | 0 | 0 | 2273 | 1197 | 0 | 0 | 0 | 0 | 0 | 0 | 0 | 0 | 0 | 0 | 0 | 0 | 0 | 0 | 0 | |
| 679 | F | 19-38 | | 6584 | 5913 | 0 | 0 | 2968 | 3376 | 0 | 0 | 0 | 0 | 0 | 0 | 0 | 0 | 0 | 0 | 0 | 0 | 0 | 0 | 0 | |
| 680 | F | 39-82 | | 2351 | 837 | 0 | 0 | #N/A | #N/A | #N/A | #N/A | 0 | 0 | 0 | 0 | 0 | 0 | 0 | 0 | 0 | 0 | 0 | 0 | 0 | |
| 681 | F | 39-82 | | 2752 | 4318 | 0 | 0 | 1146 | 1192 | 0 | 0 | 1 | 0 | 1 | 0 | 1 | 0 | 0 | 0 | 0 | 0 | 1 | 1 | 0 | |
| 682 | M | 39-82 | | 2575 | 1070 | 0 | 0 | #N/A | #N/A | #N/A | #N/A | 0 | 0 | 0 | 0 | 0 | 0 | 0 | 0 | 0 | 0 | 0 | 0 | 0 | |
| 683 | M | 39-82 | | 1447 | 451 | 0 | 0 | 1247 | 460 | 0 | 0 | 1 | 0 | 0 | 0 | 0 | 0 | 1 | 0 | 0 | 0 | 0 | 1 | 0 | |
| 684 | F | 39-82 | | 531 | 479 | 0 | 0 | 604 | 705 | 0 | 0 | 0 | 0 | 0 | 0 | 0 | 0 | 0 | 0 | 0 | 0 | 0 | 0 | 0 | |
| 685 | F | 19-38 | | 43606 | 139083 | 99 | 1 | 29197 | 128926 | 93 | 1 | 1 | 1 | 1 | 0 | 0 | 0 | 1 | 0 | 1 | 1 | 1 | 1 | 0 | |
| 686 | F | 39-82 | | 177989 | 234405 | 99 | 1 | 78196 | 104388 | 0 | 1 | 1 | 1 | 1 | 1 | 1 | 0 | 1 | 0 | 0 | 0 | 1 | 0 | 1 | 1 |
| 687 | F | 19-38 | | 2316 | 601 | 0 | 0 | 1909 | 876 | 0 | 0 | 1 | 0 | 1 | 0 | 0 | 0 | 1 | 0 | 1 | 0 | 1 | 0 | 0 | |
| 688 | F | 39-82 | | 1731 | 831 | 0 | 0 | 1331 | 642 | 0 | 0 | 0 | 0 | 0 | 0 | 0 | 0 | 0 | 0 | 0 | 0 | 0 | 0 | 0 | |
| 689 | M | 39-82 | | 2131 | 1075 | 0 | 0 | 1476 | 1545 | 0 | 0 | 0 | 0 | 0 | 0 | 0 | 0 | 0 | 0 | 0 | 0 | 0 | 0 | 0 | |
| 690 | F | 39-82 | | 9179 | 2207 | 0 | 0 | 3028 | 1339 | 0 | 0 | 0 | 0 | 0 | 0 | 0 | 0 | 0 | 0 | 0 | 0 | 0 | 0 | 0 | |
| 691 | F | 19-38 | | 28929 | 7501 | 0 | 1 | #N/A | #N/A | #N/A | #N/A | 1 | 0 | 1 | 0 | 0 | 0 | 0 | 0 | 0 | 0 | 0 | 0 | 0 | |
| 692 | F | 19-38 | | 6183 | 1452 | 0 | 0 | #N/A | #N/A | #N/A | #N/A | 0 | 0 | 0 | 1 | 0 | 0 | 0 | 0 | 0 | 0 | 0 | 0 | 0 | |
| 693 | F | 19-38 | | 2208 | 858 | 0 | 0 | 907 | 979 | 0 | 0 | 0 | 0 | 0 | 1 | 0 | 0 | 0 | 0 | 0 | 0 | 0 | 0 | 1 | 0 |
| 694 | F | 19-38 | | 5354 | 1761 | 0 | 0 | 2625 | 2503 | 0 | 0 | 1 | 0 | 0 | 1 | 0 | 0 | 0 | 0 | 0 | 0 | 1 | 0 | 0 | |
| 695 | F | 19-38 | | 7518 | 528 | 0 | 0 | 6097 | 478 | 0 | 0 | 0 | 0 | 0 | 0 | 0 | 0 | 0 | 0 | 0 | 0 | 0 | 0 | 0 | |
| 696 | M | 19-38 | | 4079 | 4123 | 0 | 0 | 4925 | 6109 | 0 | 0 | 1 | 1 | 1 | 1 | 1 | 1 | 0 | 0 | 0 | 0 | 1 | 0 | 1 | 0 |
| 697 | F | 39-82 | | 4992 | 2397 | 0 | 0 | 1644 | 1835 | 6 | 1 | 0 | 0 | 0 | 0 | 0 | 0 | 0 | 0 | 0 | 0 | 0 | 0 | 0 | |
| 698 | F | 19-38 | | 6353 | 1887 | 0 | 0 | 2754 | 2236 | 0 | 0 | 0 | 0 | 0 | 0 | 0 | 0 | 0 | 0 | 0 | 0 | 0 | 0 | 0 | |
| 699 | F | 19-38 | | 1444 | 1032 | 0 | 0 | 5566 | 938 | 0 | 0 | 1 | 0 | 1 | 0 | 0 | 0 | 0 | 0 | 0 | 0 | 1 | 0 | 0 | |
| 700 | F | 39-82 | | 3212 | 543 | 0 | 0 | 609 | 309 | 0 | 0 | 0 | 0 | 0 | 0 | 0 | 0 | 0 | 0 | 0 | 0 | 0 | 0 | 0 | |

| | | | | | | | | | | | | | | | | | | | | | | | | | |
|---|---|---|---|---|---|---|---|---|---|---|---|---|---|---|---|---|---|---|---|---|---|---|---|---|---|
| 701 | F | 19-38 | | 3925 | 3429 | 0 | 0 | 1902 | 1123 | 0 | 0 | 0 | 0 | 0 | 0 | 0 | 0 | 0 | 0 | 0 | 0 | 0 | 0 | 0 | |
| 702 | F | 39-82 | | 91579 | 49253 | 98 | 1 | 53777 | 40084 | 12 | 1 | 1 | 1 | 1 | 1 | 0 | 0 | 1 | 0 | 0 | 1 | 1 | 0 | 0 | |
| 703 | F | 39-82 | | 3438 | 1268 | 0 | 0 | 2180 | 1638 | 0 | 0 | 0 | 0 | 0 | 0 | 0 | 0 | 0 | 0 | 0 | 0 | 0 | 0 | 0 | |
| 704 | F | 39-82 | | 197859 | 211881 | 99 | 1 | 141473 | 153822 | 21 | 1 | 1 | 1 | 1 | 1 | 1 | 1 | 1 | 0 | 0 | 1 | 1 | 1 | 1 | 1 |
| 705 | F | 39-82 | | 3693 | 571 | 0 | 0 | 4156 | 613 | 0 | 0 | 1 | 0 | 1 | 0 | 0 | 0 | 1 | 1 | 1 | 0 | 1 | 1 | 0 | |
| 706 | F | 19-38 | | 3176 | 901 | 0 | 0 | 2760 | 1116 | 0 | 0 | 1 | 0 | 1 | 0 | 0 | 0 | 0 | 0 | 1 | 0 | 1 | 0 | 1 | 0 |
| 707 | F | 19-38 | | 2015 | 1881 | 0 | 0 | 2035 | 1483 | 0 | 0 | 1 | 0 | 0 | 0 | 0 | 0 | 0 | 1 | 0 | 0 | 1 | 1 | 0 | |
| 708 | F | 19-38 | | 1913 | 943 | 0 | 0 | 1741 | 1129 | 0 | 0 | 1 | 0 | 1 | 1 | 1 | 0 | 0 | 0 | 1 | 0 | 1 | 0 | 0 | |
| 709 | F | 39-82 | | 4478 | 2614 | 0 | 0 | 1549 | 2881 | 0 | 0 | 1 | 0 | 1 | 1 | 0 | 0 | 0 | 0 | 0 | 0 | 1 | 0 | 0 | |
| 710 | M | 39-82 | | 4059 | 921 | 0 | 0 | 4271 | 808 | 0 | 0 | 0 | 0 | 0 | 0 | 0 | 0 | 0 | 0 | 0 | 0 | 0 | 0 | 0 | |
| 711 | F | 39-82 | | 7485 | 3931 | 0 | 0 | #N/A | #N/A | #N/A | #N/A | 1 | 0 | 1 | 0 | 0 | 0 | 0 | 0 | 1 | 0 | 1 | 1 | 0 | |
| 712 | F | 39-82 | | 564 | 509 | 0 | 0 | 1070 | 631 | 0 | 0 | 1 | 0 | 1 | 0 | 0 | 0 | 0 | 0 | 0 | 0 | 0 | 0 | 0 | |
| 713 | M | 39-82 | | 1904 | 814 | 0 | 0 | 1224 | 836 | 0 | 0 | 0 | 0 | 0 | 0 | 0 | 0 | 0 | 0 | 0 | 0 | 0 | 0 | 0 | |
| 714 | F | 19-38 | | 2640 | 1165 | 0 | 0 | 1916 | 996 | 0 | 0 | 1 | 0 | 0 | 0 | 0 | 0 | 0 | 0 | 0 | 0 | 1 | 1 | 0 | |
| 715 | F | 39-82 | | 7686 | 1171 | 0 | 0 | 3188 | 1317 | 0 | 0 | 0 | 0 | 0 | 0 | 0 | 0 | 0 | 0 | 0 | 0 | 0 | 0 | 0 | |
| 716 | F | 39-82 | | 2128 | 2085 | 0 | 0 | #N/A | #N/A | #N/A | #N/A | 1 | 1 | 1 | 1 | 0 | 0 | 1 | 0 | 1 | 0 | 0 | 0 | 0 | |
| 717 | F | 39-82 | | 4789 | 1760 | 0 | 0 | 4727 | 1715 | 0 | 0 | 0 | 0 | 0 | 0 | 0 | 0 | 0 | 0 | 0 | 0 | 0 | 0 | 1 | 0 |
| 718 | F | 39-82 | | 2525 | 1111 | 0 | 0 | 395 | 414 | 0 | 0 | 1 | 0 | 1 | 0 | 0 | 0 | 0 | 0 | 0 | 0 | 0 | 0 | 0 | |
| 719 | F | 39-82 | | 5002 | 1017 | 0 | 0 | 2640 | 1543 | 0 | 0 | 0 | 0 | 0 | 0 | 0 | 0 | 0 | 0 | 0 | 0 | 0 | 0 | 0 | |
| 720 | F | 19-38 | | 168 | 1046 | 0 | 0 | #N/A | #N/A | #N/A | #N/A | 0 | 0 | 0 | 0 | 0 | 0 | 0 | 0 | 0 | 0 | 0 | 0 | 0 | |
| 721 | F | 19-38 | negative | 301 | 450 | 0 | 0 | #N/A | #N/A | #N/A | #N/A | 1 | 1 | 1 | 1 | 0 | 0 | 1 | 0 | 1 | 1 | 1 | 1 | 0 | |
| 722 | F | 39-82 | | 65709 | 15270 | 98 | 1 | 46826 | 9813 | 0 | 1 | 0 | 0 | 0 | 0 | 0 | 0 | 0 | 0 | 0 | 0 | 0 | 0 | 0 | |
| 723 | F | 39-82 | | 4565 | 370 | 0 | 0 | 2224 | 741 | 0 | 0 | 0 | 0 | 0 | 0 | 0 | 0 | 0 | 0 | 0 | 0 | 0 | 0 | 0 | |
| 724 | F | 19-38 | | 1049 | 1214 | 0 | 0 | 2343 | 3567 | 0 | 0 | 0 | 0 | 0 | 0 | 0 | 0 | 0 | 0 | 0 | 0 | 0 | 0 | 0 | |
| 725 | F | 39-82 | | 6676 | 716 | 0 | 0 | #N/A | #N/A | #N/A | #N/A | 1 | 1 | 1 | 0 | 0 | 0 | 0 | 0 | 0 | 0 | 0 | 0 | 0 | |
| 726 | F | 39-82 | | 1594 | 1333 | 0 | 0 | 2089 | 2463 | 0 | 0 | 1 | 0 | 1 | 1 | 0 | 0 | 0 | 0 | 0 | 0 | 0 | 0 | 0 | |
| 727 | F | 19-38 | | 4790 | 1253 | 0 | 0 | #N/A | #N/A | #N/A | #N/A | 1 | 0 | 0 | 0 | 0 | 0 | 0 | 0 | 0 | 0 | 0 | 1 | 0 | |
| 728 | F | 19-38 | | 5870 | 1321 | 0 | 0 | #N/A | #N/A | #N/A | #N/A | 1 | 0 | 1 | 0 | 0 | 0 | 0 | 0 | 0 | 0 | 0 | 0 | 0 | |
| 729 | F | 19-38 | | 98007 | 31234 | 80 | 1 | #N/A | #N/A | #N/A | #N/A | 0 | 0 | 0 | 0 | 0 | 0 | 0 | 0 | 0 | 0 | 0 | 0 | 0 | |
| 730 | F | 19-38 | | 6621 | 1460 | 0 | 0 | 1509 | 1445 | 0 | 0 | 0 | 0 | 0 | 0 | 0 | 0 | 0 | 0 | 0 | 0 | 0 | 0 | 0 | |
| 731 | F | 39-82 | | 1840 | 1013 | 0 | 0 | 355 | 146 | 0 | 0 | 0 | 0 | 0 | 0 | 0 | 0 | 0 | 0 | 0 | 0 | 0 | 0 | 0 | |
| 732 | F | 19-38 | | 5442 | 1716 | 0 | 0 | #N/A | #N/A | #N/A | #N/A | 0 | 0 | 0 | 0 | 0 | 0 | 0 | 0 | 0 | 0 | 0 | 0 | 0 | |
| 733 | F | 39-82 | | 2292 | 2087 | 0 | 0 | 205 | 122 | 0 | 0 | 0 | 0 | 0 | 0 | 0 | 0 | 0 | 0 | 0 | 0 | 0 | 0 | 0 | |
| 734 | F | 39-82 | | 909 | 816 | 0 | 0 | 1487 | 1033 | 0 | 0 | 0 | 0 | 0 | 0 | 0 | 0 | 0 | 0 | 0 | 0 | 0 | 0 | 0 | |
| 735 | F | 19-38 | | 2862 | 1037 | 0 | 0 | 736 | 1011 | 0 | 0 | 1 | 1 | 1 | 1 | 1 | 0 | 0 | 1 | 0 | 0 | 1 | 0 | 0 | |
| 736 | M | 39-82 | | 3805 | 1332 | 0 | 0 | #N/A | #N/A | #N/A | #N/A | 0 | 0 | 0 | 0 | 0 | 0 | 0 | 0 | 0 | 0 | 0 | 0 | 0 | |
| 737 | M | 19-38 | | 3482 | 826 | 0 | 0 | 1226 | 607 | 0 | 0 | 0 | 0 | 0 | 0 | 0 | 0 | 0 | 0 | 0 | 0 | 0 | 0 | 0 | |
| 738 | M | 39-82 | | 690 | 497 | 0 | 0 | #N/A | #N/A | #N/A | #N/A | 0 | 0 | 0 | 0 | 0 | 0 | 0 | 0 | 0 | 0 | 0 | 0 | 0 | |
| 739 | M | 19-38 | | 4804 | 1295 | 0 | 0 | 1403 | 1412 | 0 | 0 | 0 | 0 | 0 | 0 | 0 | 0 | 0 | 0 | 0 | 0 | 0 | 0 | 0 | |
| 740 | F | 39-82 | | 1185 | 1200 | 0 | 0 | 881 | 1811 | 0 | 0 | 1 | 0 | 1 | 0 | 0 | 0 | 0 | 0 | 1 | 0 | 1 | 0 | 0 | |
| 741 | F | 39-82 | | 2220 | 1090 | 0 | 0 | 1982 | 492 | 0 | 0 | 1 | 1 | 0 | 1 | 0 | 0 | 0 | 0 | 1 | 0 | 1 | 0 | 0 | |
| 742 | F | 39-82 | | 1173 | 632 | 0 | 0 | 290 | 456 | 0 | 0 | 1 | 0 | 1 | 0 | 0 | 0 | 0 | 0 | 0 | 0 | 1 | 0 | 0 | |
| 743 | M | 39-82 | | 1593 | 1940 | 0 | 0 | 758 | 2602 | 0 | 0 | 1 | 0 | 0 | 0 | 0 | 1 | 0 | 0 | 0 | 0 | 0 | 0 | 1 | 0 |
| 744 | F | 39-82 | | 2935 | 679 | 0 | 0 | 1989 | 858 | 0 | 0 | 1 | 1 | 1 | 1 | 0 | 0 | 0 | 0 | 1 | 0 | 1 | 0 | 0 | |
| 745 | M | 39-82 | | 17174 | 1302 | 0 | 1 | #N/A | #N/A | #N/A | #N/A | 0 | 0 | 0 | 0 | 0 | 0 | 0 | 0 | 0 | 0 | 0 | 0 | 0 | |
| 746 | F | 39-82 | | 709 | 2884 | 0 | 0 | #N/A | #N/A | #N/A | #N/A | 1 | 0 | 0 | 0 | 0 | 0 | 0 | 0 | 0 | 0 | 1 | 0 | 0 | |
| 747 | M | 19-38 | | 5896 | 4134 | 0 | 0 | 5371 | 1944 | 0 | 0 | 1 | 0 | 1 | 1 | 0 | 0 | 0 | 0 | 0 | 0 | 1 | 0 | 1 | 0 |
| 748 | F | 39-82 | | 378 | 648 | 0 | 0 | 1461 | 803 | 0 | 0 | 1 | 0 | 1 | 0 | 0 | 0 | 0 | 0 | 0 | 0 | 1 | 0 | 0 | |
| 749 | F | 19-38 | | 2547 | 788 | 0 | 0 | 8840 | 4414 | 0 | 0 | 1 | 0 | 1 | 1 | 0 | 1 | 1 | 0 | 0 | 0 | 1 | 0 | 1 | 0 |
| 750 | M | 39-82 | | 1611 | 1651 | 0 | 0 | 3430 | 2758 | 0 | 0 | 1 | 0 | 1 | 0 | 0 | 0 | 1 | 0 | 1 | 0 | 0 | 1 | 0 | |
| 751 | M | 39-82 | | 8841 | 712 | 0 | 0 | 7172 | 1100 | 0 | 0 | 0 | 0 | 0 | 0 | 0 | 0 | 0 | 0 | 0 | 0 | 0 | 0 | 0 | |
| 752 | F | 19-38 | | 518 | 929 | 0 | 0 | 832 | 1230 | 0 | 0 | 1 | 0 | 0 | 0 | 0 | 0 | 1 | 0 | 0 | 0 | 1 | 0 | 0 | |
| 753 | M | 39-82 | | 1517 | 577 | 0 | 0 | 2431 | 1467 | 0 | 0 | 0 | 0 | 0 | 0 | 0 | 0 | 0 | 0 | 0 | 0 | 0 | 0 | 0 | |
| 754 | F | 19-38 | | 5174 | 2747 | 0 | 0 | 1515 | 2175 | 0 | 0 | 1 | 0 | 1 | 0 | 0 | 0 | 0 | 0 | 0 | 0 | 0 | 0 | 0 | |
| 755 | M | 19-38 | | 5006 | 706 | 0 | 0 | 4091 | 656 | 0 | 0 | 0 | 0 | 0 | 0 | 0 | 0 | 0 | 0 | 0 | 0 | 0 | 0 | 0 | |
| 756 | M | 19-38 | | 8367 | 2790 | 0 | 0 | 4164 | 3631 | 0 | 0 | 1 | 0 | 1 | 1 | 0 | 0 | 0 | 0 | 0 | 0 | 0 | 0 | 0 | |
| 757 | F | 39-82 | | 2621 | 2767 | 0 | 0 | 1108 | 3125 | 0 | 0 | 0 | 0 | 0 | 0 | 0 | 0 | 0 | 0 | 0 | 0 | 0 | 0 | 0 | |
| 758 | F | 19-38 | | 1299 | 1205 | 0 | 0 | #N/A | #N/A | #N/A | #N/A | 0 | 0 | 0 | 0 | 0 | 0 | 0 | 0 | 0 | 0 | 0 | 0 | 0 | |
| 759 | F | 19-38 | | 5255 | 1445 | 0 | 0 | 3360 | 2931 | 0 | 0 | 1 | 0 | 0 | 0 | 0 | 0 | 0 | 0 | 0 | 0 | 1 | 0 | 0 | |
| 760 | F | 39-82 | | 2805 | 662 | 0 | 0 | 1406 | 717 | 0 | 0 | 1 | 0 | 0 | 0 | 0 | 1 | 1 | 0 | 0 | 0 | 0 | 0 | 0 | |
| 761 | M | 19-38 | | 3298 | 1088 | 0 | 0 | 1463 | 511 | 0 | 0 | 0 | 0 | 0 | 0 | 0 | 0 | 0 | 0 | 0 | 0 | 0 | 0 | 0 | |
| 762 | F | 19-38 | | 1818 | 782 | 0 | 0 | 1039 | 890 | 0 | 0 | 1 | 1 | 1 | 0 | 0 | 0 | 1 | 0 | 1 | 0 | 1 | 1 | 0 | |
| 763 | M | 39-82 | | 4172 | 869 | 0 | 0 | 1530 | 961 | 0 | 0 | 1 | 1 | 0 | 1 | 0 | 0 | 1 | 0 | 1 | 0 | 0 | 0 | 0 | |
| 764 | M | 19-38 | | 3150 | 892 | 0 | 0 | 2493 | 660 | 0 | 0 | 0 | 0 | 0 | 0 | 0 | 0 | 0 | 0 | 0 | 0 | 0 | 0 | 0 | |
| 765 | F | 19-38 | | 3242 | 699 | 0 | 0 | #N/A | #N/A | #N/A | #N/A | 1 | 0 | 0 | 0 | 0 | 0 | 1 | 0 | 0 | 0 | 1 | 0 | 0 | |
| 766 | F | 39-82 | | 2117 | 2056 | 0 | 0 | 873 | 721 | 0 | 0 | 0 | 0 | 0 | 0 | 0 | 0 | 0 | 0 | 0 | 0 | 0 | 0 | 0 | |
| 767 | M | 19-38 | | 1064 | 575 | 0 | 0 | 1675 | 790 | 0 | 0 | 1 | 0 | 0 | 0 | 0 | 1 | 0 | 0 | 0 | 0 | 1 | 0 | 0 | |
| 768 | F | 19-38 | | 5355 | 4786 | 0 | 0 | 735 | 1318 | 0 | 0 | 1 | 0 | 1 | 0 | 0 | 0 | 0 | 0 | 0 | 0 | 1 | 0 | 0 | |
| 769 | F | 19-38 | | 4632 | 1737 | 0 | 0 | 5412 | 2819 | 0 | 0 | 0 | 0 | 0 | 1 | 0 | 0 | 0 | 0 | 0 | 0 | 0 | 0 | 0 | |
| 770 | F | 39-82 | | 524 | 321 | 0 | 0 | 1273 | 1001 | 0 | 0 | 0 | 0 | 0 | 0 | 0 | 0 | 0 | 0 | 0 | 0 | 0 | 0 | 0 | |
| 771 | F | 39-82 | | 4481 | 453 | 0 | 0 | 2244 | 712 | 0 | 0 | 0 | 0 | 0 | 0 | 0 | 0 | 0 | 0 | 0 | 0 | 0 | 0 | 0 | |
| 772 | M | 39-82 | | 106434 | 31310 | 82 | 1 | #N/A | #N/A | #N/A | #N/A | 1 | 1 | 0 | 0 | 0 | 0 | 0 | 0 | 0 | 0 | 0 | 0 | 0 | |
| 773 | M | 39-82 | | 2813 | 557 | 0 | 0 | #N/A | #N/A | #N/A | #N/A | 1 | 0 | 1 | 0 | 0 | 0 | 0 | 0 | 1 | 0 | 1 | 0 | 0 | |
| 774 | M | 39-82 | | 1710 | 1427 | 0 | 0 | 891 | 517 | 0 | 0 | 0 | 0 | 0 | 0 | 0 | 0 | 0 | 0 | 0 | 0 | 0 | 0 | 0 | |
| 775 | M | 19-38 | | 4276 | 1438 | 0 | 0 | 2610 | 1263 | 0 | 0 | 0 | 0 | 0 | 0 | 0 | 0 | 0 | 0 | 0 | 0 | 0 | 0 | 0 | |
| 776 | F | 19-38 | | 2634 | 570 | 0 | 0 | #N/A | #N/A | #N/A | #N/A | 1 | 0 | 1 | 0 | 0 | 0 | 0 | 0 | 1 | 0 | 1 | 0 | 0 | |
| 777 | M | 19-38 | | 1585 | 831 | 0 | 0 | 1807 | 2262 | 0 | 0 | 0 | 0 | 0 | 0 | 0 | 0 | 0 | 0 | 0 | 0 | 0 | 0 | 0 | |

| | | | | | | | | | | | | | | | | | | | | | | | | |
|---|---|---|---|---|---|---|---|---|---|---|---|---|---|---|---|---|---|---|---|---|---|---|---|---|
| 778 | F | 39-82 | 1245 | 1355 | 0 | 0 | 2580 | 1397 | 0 | 0 | 1 | 1 | 1 | 0 | 0 | 0 | 0 | 0 | 1 | 0 | 1 | 0 | 0 | |
| 779 | F | 19-38 | 6421 | 5251 | 0 | 0 | #N/A | #N/A | #N/A | #N/A | 0 | 0 | 0 | 0 | 0 | 0 | 0 | 0 | 0 | 0 | 0 | 0 | 0 | |
| 780 | F | 19-38 | 1283 | 2236 | 0 | 0 | #N/A | #N/A | #N/A | #N/A | 0 | 0 | 0 | 0 | 0 | 0 | 0 | 0 | 0 | 0 | 0 | 0 | 0 | |
| 781 | F | 19-38 | 5318 | 559 | 0 | 0 | 2130 | 956 | 0 | 0 | 0 | 0 | 0 | 0 | 0 | 0 | 0 | 0 | 0 | 0 | 0 | 0 | 0 | |
| 782 | M | 19-38 | 7139 | 989 | 0 | 0 | 2847 | 957 | 0 | 0 | 0 | 0 | 0 | 0 | 0 | 0 | 0 | 0 | 0 | 0 | 0 | 0 | 0 | |
| 783 | F | 19-38 | 2034 | 509 | 0 | 0 | #N/A | #N/A | #N/A | #N/A | 1 | 0 | 1 | 0 | 0 | 0 | 0 | 0 | 0 | 0 | 1 | 1 | 0 | |
| 784 | F | 39-82 | 5614 | 1001 | 0 | 0 | #N/A | #N/A | #N/A | #N/A | 1 | 1 | 1 | 0 | 0 | 0 | 0 | 0 | 1 | 0 | 1 | 1 | 0 | |
| 785 | F | 39-82 | 1342 | 3084 | 0 | 0 | 1155 | 3131 | 0 | 0 | 0 | 0 | 0 | 0 | 0 | 0 | 0 | 0 | 0 | 0 | 0 | 0 | 0 | |
| 786 | F | 19-38 | 841 | 422 | 0 | 0 | 849 | 547 | 0 | 0 | 1 | 0 | 1 | 1 | 0 | 0 | 0 | 0 | 0 | 0 | 1 | 0 | 0 | |
| 787 | F | 19-38 | 3337 | 3654 | 0 | 0 | 736 | 1159 | 0 | 0 | 1 | 0 | 1 | 0 | 0 | 0 | 0 | 0 | 0 | 0 | 1 | 1 | 0 | |
| 788 | F | 39-82 | 2420 | 425 | 0 | 0 | #N/A | #N/A | #N/A | #N/A | 1 | 1 | 1 | 1 | 1 | 1 | 0 | 0 | 0 | 0 | 1 | 1 | 0 | |
| 789 | F | 39-82 | 104219 | 69779 | 81 | 1 | 16760 | 47551 | 65 | 1 | 1 | 1 | 0 | 1 | 0 | 0 | 0 | 0 | 0 | 1 | 0 | 0 | 0 | |
| 790 | M | 39-82 | 2592 | 985 | 0 | 0 | 2401 | 1534 | 0 | 0 | 0 | 0 | 0 | 0 | 0 | 0 | 0 | 0 | 0 | 0 | 0 | 0 | 0 | |
| 791 | F | 39-82 | 9055 | 3250 | 0 | 0 | 4250 | 4938 | 0 | 0 | 0 | 0 | 0 | 0 | 0 | 0 | 0 | 0 | 0 | 0 | 0 | 0 | 0 | |
| 792 | M | 39-82 | 15477 | 8309 | 0 | 1 | 14376 | 8602 | 0 | 1 | 0 | 0 | 0 | 0 | 0 | 0 | 0 | 0 | 0 | 0 | 0 | 0 | 0 | |
| 793 | F | 19-38 | 1568 | 3743 | 0 | 0 | #N/A | #N/A | #N/A | #N/A | 1 | 0 | 0 | 0 | 0 | 0 | 0 | 0 | 0 | 0 | 1 | 1 | 0 | |
| 794 | F | 39-82 | 2016 | 1083 | 0 | 0 | #N/A | #N/A | #N/A | #N/A | 1 | 0 | 1 | 0 | 0 | 0 | 1 | 0 | 1 | 0 | 1 | 0 | 0 | |
| 795 | M | 39-82 | 1022 | 7764 | 0 | 0 | 833 | 5697 | 0 | 0 | 0 | 0 | 0 | 0 | 0 | 0 | 0 | 0 | 0 | 0 | 0 | 0 | 0 | |
| 796 | F | 19-38 | 2319 | 3771 | 0 | 0 | #N/A | #N/A | #N/A | #N/A | 0 | 0 | 0 | 0 | 0 | 0 | 0 | 0 | 0 | 0 | 0 | 0 | 0 | |
| 797 | M | 39-82 | 5086 | 618 | 0 | 0 | 1300 | 330 | 0 | 0 | 0 | 0 | 0 | 0 | 0 | 0 | 0 | 0 | 0 | 0 | 0 | 0 | 0 | |
| 798 | F | 39-82 | 3233 | 2157 | 0 | 0 | 2265 | 1908 | 0 | 0 | 0 | 0 | 0 | 0 | 0 | 0 | 0 | 0 | 0 | 0 | 0 | 0 | 0 | |
| 799 | F | 39-82 | 2011 | 2082 | 0 | 0 | 2750 | 1967 | 18 | 1 | 1 | 0 | 1 | 1 | 0 | 0 | 1 | 0 | 1 | 0 | 1 | 0 | 0 | |
| 800 | M | 19-38 | 2360 | 2854 | 0 | 0 | #N/A | #N/A | #N/A | #N/A | 0 | 0 | 0 | 0 | 0 | 0 | 0 | 0 | 0 | 0 | 0 | 0 | 0 | |
| 801 | M | 39-82 | 6623 | 6226 | 0 | 0 | 1764 | 1426 | 0 | 0 | 0 | 0 | 0 | 0 | 0 | 0 | 0 | 0 | 0 | 0 | 0 | 0 | 0 | |
| 802 | F | 39-82 | 445 | 688 | 0 | 0 | 671 | 736 | 0 | 0 | 0 | 0 | 0 | 0 | 0 | 0 | 0 | 0 | 0 | 0 | 0 | 0 | 0 | |
| 803 | F | 39-82 | 1540 | 605 | 0 | 0 | 1180 | 567 | 0 | 0 | 0 | 0 | 0 | 0 | 0 | 0 | 0 | 0 | 0 | 0 | 0 | 0 | 0 | |
| 804 | M | 19-38 | 3568 | 862 | 0 | 0 | #N/A | #N/A | #N/A | #N/A | 0 | 0 | 0 | 0 | 0 | 0 | 0 | 0 | 0 | 0 | 0 | 0 | 0 | |
| 805 | F | 19-38 | 43029 | 48682 | 70 | 1 | 21182 | 112738 | 97 | 1 | 1 | 1 | 1 | 1 | 1 | 0 | 1 | 0 | 0 | 0 | 0 | 0 | 0 | |
| 806 | M | 39-82 | 1598 | 3238 | 0 | 0 | 1491 | 3310 | 0 | 0 | 1 | 1 | 1 | 1 | 0 | 0 | 1 | 0 | 1 | 0 | 1 | 0 | 0 | |
| 807 | M | 39-82 | 1599 | 50 | 0 | 0 | 1079 | 679 | 0 | 0 | 0 | 0 | 0 | 0 | 0 | 0 | 0 | 0 | 0 | 0 | 0 | 0 | 0 | |
| 808 | F | 39-82 | 5565 | 67 | 0 | 0 | #N/A | #N/A | #N/A | #N/A | 1 | 0 | 1 | 0 | 1 | 0 | 0 | 0 | 1 | 0 | 0 | 0 | 0 | |
| 809 | M | 19-38 | 2422 | 757 | 0 | 0 | #N/A | #N/A | #N/A | #N/A | 1 | 1 | 1 | 0 | 1 | 1 | 1 | 0 | 0 | 0 | 1 | 1 | 0 | |
| 810 | M | 19-38 | 72380 | 117519 | 97 | 1 | 18810 | 66318 | 97 | 1 | 1 | 1 | 1 | 1 | 0 | 0 | 1 | 0 | 1 | 1 | 1 | 1 | 0 | |
| 811 | F | 39-82 | 993 | 38 | 0 | 0 | 1824 | 2523 | 0 | 0 | 1 | 1 | 1 | 0 | 0 | 0 | 1 | 1 | 1 | 0 | 1 | 1 | 0 | |
| 812 | F | 39-82 | 1390 | 82 | 0 | 0 | 1470 | 519 | 0 | 0 | 1 | 0 | 1 | 0 | 0 | 0 | 0 | 0 | 0 | 0 | 1 | 1 | 0 | |
| 813 | F | 19-38 | 790 | 23 | 0 | 0 | #N/A | #N/A | #N/A | #N/A | 1 | 0 | 1 | 0 | 0 | 0 | 1 | 0 | 1 | 0 | 1 | 0 | 0 | |
| 814 | F | 19-38 | 1705 | 97 | 0 | 0 | 1092 | 1121 | 0 | 0 | 1 | 1 | 1 | 1 | 0 | 1 | 1 | 0 | 0 | 0 | 1 | 0 | 0 | |
| 815 | F | 19-38 | 7183 | 799 | 0 | 0 | 8604 | 937 | 0 | 0 | 1 | 0 | 1 | 0 | 0 | 0 | 0 | 1 | 1 | 0 | 1 | 0 | 0 | |
| 816 | F | 39-82 | 12479 | 1105 | 0 | 1 | 14397 | 13885 | 0 | 1 | 1 | 0 | 1 | 0 | 0 | 0 | 1 | 0 | 0 | 0 | 1 | 0 | 0 | |
| 817 | F | 39-82 | 4031 | 1553 | 0 | 0 | 1830 | 953 | 0 | 0 | 1 | 1 | 1 | 0 | 0 | 0 | 1 | 0 | 0 | 0 | 1 | 0 | 0 | |
| 818 | F | 39-82 | 1816 | 664 | 0 | 0 | 310 | 511 | 0 | 0 | 1 | 0 | 1 | 1 | 0 | 1 | 1 | 0 | 1 | 0 | 1 | 0 | 0 | |
| 819 | F | 39-82 | 3666 | 480 | 0 | 0 | 2396 | 567 | 0 | 0 | 1 | 0 | 1 | 0 | 0 | 0 | 0 | 0 | 0 | 0 | 1 | 0 | 0 | |
| 820 | F | 19-38 | 1059 | 2794 | 0 | 0 | #N/A | #N/A | #N/A | #N/A | 1 | 0 | 1 | 1 | 0 | 1 | 0 | 0 | 1 | 0 | 0 | 0 | 0 | |
| 821 | F | 39-82 | 743 | 1414 | 0 | 0 | 999 | 1008 | 0 | 0 | 0 | 0 | 0 | 0 | 0 | 0 | 0 | 0 | 0 | 0 | 0 | 0 | 0 | |
| 822 | F | 19-38 | 2489 | 1178 | 0 | 0 | 1757 | 742 | 0 | 0 | 1 | 0 | 1 | 1 | 0 | 0 | 0 | 1 | 0 | 0 | 0 | 0 | 0 | |
| 823 | F | 39-82 | 9837 | 1190 | 0 | 0 | 5862 | 1368 | 0 | 0 | 0 | 0 | 0 | 0 | 0 | 0 | 0 | 0 | 0 | 0 | 0 | 0 | 0 | |
| 824 | F | 19-38 | 1195 | 494 | 0 | 0 | #N/A | #N/A | #N/A | #N/A | 1 | 1 | 1 | 1 | 0 | 1 | 1 | 1 | 0 | 0 | 1 | 0 | 0 | |
| 825 | F | 19-38 | 1111 | 825 | 0 | 0 | 1478 | 1089 | 0 | 0 | 0 | 0 | 0 | 0 | 0 | 0 | 0 | 0 | 0 | 0 | 0 | 0 | 0 | |
| 826 | F | 19-38 | 2766 | 1196 | 0 | 0 | 1181 | 527 | 0 | 0 | 0 | 0 | 0 | 0 | 0 | 0 | 0 | 0 | 0 | 0 | 0 | 0 | 0 | |
| 827 | F | 39-82 | 4480 | 1205 | 0 | 0 | 3156 | 743 | 0 | 0 | 0 | 0 | 0 | 0 | 0 | 0 | 0 | 0 | 0 | 0 | 0 | 0 | 0 | |
| 828 | F | 19-38 | 648 | 2261 | 0 | 0 | 2281 | 4345 | 0 | 0 | 1 | 1 | 1 | 1 | 0 | 0 | 1 | 0 | 1 | 0 | 1 | 0 | 0 | |
| 829 | F | 19-38 | 4415 | 988 | 0 | 0 | 4897 | 1295 | 0 | 0 | 1 | 0 | 1 | 1 | 1 | 1 | 1 | 0 | 1 | 1 | 0 | 0 | 0 | |
| 830 | F | 39-82 | 1146 | 454 | 0 | 0 | 1054 | 437 | 0 | 0 | 1 | 0 | 0 | 0 | 0 | 1 | 1 | 0 | 0 | 0 | 0 | 0 | 1 | 0 |
| 831 | F | 19-38 | 1640 | 1864 | 0 | 0 | 4729 | 5121 | 0 | 0 | 0 | 0 | 0 | 0 | 0 | 0 | 0 | 0 | 0 | 0 | 0 | 0 | 0 | |
| 832 | M | 19-38 | 107478 | 25830 | 84 | 1 | #N/A | #N/A | #N/A | #N/A | 1 | 0 | 1 | 1 | 0 | 0 | 0 | 0 | 1 | 1 | 1 | 0 | 0 | |
| 833 | M | 19-38 | 3267 | 858 | 0 | 0 | #N/A | #N/A | #N/A | #N/A | 1 | 0 | 0 | 0 | 0 | 0 | 0 | 1 | 0 | 0 | 1 | 1 | 0 | |
| 834 | F | 19-38 | 2629 | 1139 | 0 | 0 | 3519 | 1467 | 0 | 0 | 1 | 1 | 1 | 1 | 1 | 1 | 1 | 0 | 1 | 1 | 1 | 0 | 0 | |
| 835 | F | 19-38 | 5182 | 3823 | 0 | 0 | #N/A | #N/A | #N/A | #N/A | 0 | 0 | 0 | 0 | 0 | 0 | 0 | 0 | 0 | 0 | 0 | 0 | 0 | |
| 836 | M | 39-82 | 5647 | 5193 | 0 | 0 | #N/A | #N/A | #N/A | #N/A | 0 | 0 | 0 | 0 | 0 | 0 | 0 | 0 | 0 | 0 | 0 | 0 | 0 | |
| 837 | F | 19-38 | 7014 | 3621 | 0 | 0 | #N/A | #N/A | #N/A | #N/A | 0 | 0 | 0 | 0 | 0 | 0 | 0 | 0 | 0 | 0 | 0 | 0 | 0 | |
| 838 | M | 19-38 | 205635 | 182327 | 99 | 1 | #N/A | #N/A | #N/A | #N/A | 1 | 0 | 1 | 0 | 0 | 0 | 1 | 0 | 0 | 1 | 0 | 0 | 1 | 1 |
| 839 | M | 39-82 | 3845 | 996 | 0 | 0 | 2221 | 2446 | 0 | 0 | 0 | 0 | 0 | 0 | 0 | 0 | 0 | 0 | 0 | 0 | 0 | 0 | 0 | |
| 840 | F | 39-82 | 2432 | 1688 | 0 | 0 | 2113 | 1992 | 0 | 0 | 1 | 0 | 1 | 1 | 0 | 0 | 1 | 0 | 1 | 0 | 0 | 0 | 0 | |
| 841 | F | 19-38 | 3725 | 1049 | 0 | 0 | 2315 | 725 | 0 | 0 | 1 | 0 | 1 | 0 | 0 | 0 | 0 | 0 | 0 | 0 | 1 | 0 | 0 | |
| 842 | F | 19-38 | 10206 | 801 | 0 | 0 | 4334 | 2873 | 0 | 0 | 0 | 0 | 0 | 0 | 0 | 0 | 0 | 0 | 0 | 0 | 0 | 0 | 0 | |
| 843 | M | 39-82 | 158428 | 56132 | 89 | 1 | 21856 | 33847 | 60 | 1 | 1 | 1 | 1 | 0 | 0 | 0 | 0 | 0 | 0 | 0 | 1 | 0 | 0 | |
| 844 | F | 39-82 | 1386 | 1017 | 0 | 0 | 891 | 867 | 4 | 1 | 1 | 0 | 1 | 1 | 0 | 0 | 0 | 0 | 0 | 0 | 0 | 0 | 0 | |
| 845 | F | 39-82 | 2272 | 689 | 0 | 0 | 1518 | 925 | 0 | 0 | 0 | 0 | 0 | 0 | 0 | 0 | 0 | 0 | 0 | 0 | 0 | 0 | 0 | |
| 846 | M | 39-82 | 4406 | 6985 | 0 | 0 | 2705 | 6308 | 7 | 1 | 0 | 0 | 0 | 0 | 0 | 0 | 0 | 0 | 0 | 0 | 0 | 0 | 0 | |
| 847 | F | 39-82 | 1873 | 794 | 0 | 0 | 1075 | 800 | 0 | 0 | 1 | 0 | 1 | 0 | 0 | 0 | 0 | 0 | 0 | 0 | 0 | 1 | 0 | |
| 848 | F | 19-38 | 30250 | 34324 | 77 | 1 | #N/A | #N/A | #N/A | #N/A | 1 | 0 | 1 | 0 | 1 | 1 | 1 | 0 | 1 | 1 | 0 | 0 | 0 | |
| 849 | F | 19-38 | 6576 | 3138 | 0 | 0 | 870 | 490 | 0 | 0 | 0 | 0 | 0 | 0 | 0 | 0 | 0 | 0 | 0 | 0 | 0 | 0 | 0 | |
| 850 | F | 19-38 | 3658 | 1969 | 0 | 0 | 2862 | 1442 | 0 | 0 | 1 | 0 | 0 | 0 | 0 | 0 | 0 | 0 | 0 | 0 | 0 | 1 | 0 | |
| 851 | M | 19-38 | 9516 | 2476 | 0 | 0 | 7796 | 2345 | 0 | 0 | 0 | 0 | 0 | 0 | 0 | 0 | 0 | 0 | 0 | 0 | 0 | 0 | 0 | |
| 852 | F | 39-82 | 2857 | 462 | 0 | 0 | #N/A | #N/A | #N/A | #N/A | 1 | 0 | 0 | 1 | 0 | 0 | 1 | 0 | 1 | 0 | 1 | 0 | 0 | |
| 853 | M | 39-82 | 1490 | 684 | 0 | 0 | #N/A | #N/A | #N/A | #N/A | 1 | 0 | 1 | 1 | 1 | 1 | 0 | 1 | 0 | 0 | 1 | 0 | 0 | |
| 854 | M | 19-38 | 1655 | 1980 | 0 | 0 | 420 | 384 | 0 | 0 | 0 | 0 | 0 | 1 | 0 | 0 | 0 | 0 | 0 | 0 | 0 | 0 | 0 | |

| | | | | | | | | | | |
|---|---|---|---|---|---|---|---|---|---|---|
| 855 | M | 39-82 | 8062 | 965 | 0 | 0 | 9336 | 614 | 0 | 0 |
| 856 | F | 19-38 | 1749 | 1551 | 0 | 0 | 1557 | 1969 | 10 | 1 |
| 857 | F | 39-82 | 4523 | 1245 | 0 | 0 | #N/A | #N/A | #N/A | #N/A |
| 858 | F | 19-38 | 1380 | 1822 | 0 | 0 | 1725 | 2517 | 0 | 0 |
| 859 | F | 39-82 | 596 | 426 | 0 | 0 | 1031 | 314 | 0 | 0 |
| 860 | F | 39-82 | 512 | 444 | 0 | 0 | 401 | 280 | 0 | 0 |
| 861 | F | 19-38 | 3211 | 379 | 0 | 0 | 1565 | 150 | 0 | 0 |
| 862 | M | 19-38 | 5959 | 3017 | 0 | 0 | 1756 | 688 | 0 | 0 |
| 863 | F | 19-38 | 8418 | 997 | 0 | 0 | #N/A | #N/A | #N/A | #N/A |
| 864 | F | 39-82 | 4567 | 9535 | 67 | 1 | 2869 | 4688 | 33 | 1 |
| 865 | M | 39-82 | 2521 | 1786 | 0 | 0 | 2343 | 1374 | 0 | 0 |
| 866 | F | 19-38 | 111889 | 95446 | 98 | 1 | 66126 | 81127 | 88 | 1 |
| 867 | M | 19-38 | 2792 | 3768 | 0 | 0 | #N/A | #N/A | #N/A | #N/A |
| 868 | F | 19-38 | 4528 | 1072 | 0 | 0 | 1651 | 1363 | 0 | 0 |
| 869 | M | 19-38 | 6789 | 1426 | 0 | 0 | #N/A | #N/A | #N/A | #N/A |
| 870 | M | 19-38 | 3638 | 1148 | 0 | 0 | #N/A | #N/A | #N/A | #N/A |
| 871 | F | 19-38 | 4658 | 1405 | 0 | 0 | 1984 | 1083 | 0 | 0 |
| 872 | F | 39-82 | 483 | 710 | 0 | 0 | #N/A | #N/A | #N/A | #N/A |
| 873 | F | 19-38 | 1451 | 543 | 0 | 0 | 1777 | 1537 | 0 | 0 |
| 874 | F | 19-38 | -673 | 844 | 0 | 0 | 639 | 385 | 0 | 0 |
| 875 | F | 39-82 | 2254 | 736 | 0 | 0 | 1733 | 1018 | 0 | 0 |
| 876 | F | 19-38 | 2121 | 1110 | 0 | 0 | #N/A | #N/A | #N/A | #N/A |
| 877 | F | 19-38 | 4550 | 1901 | 0 | 0 | #N/A | #N/A | #N/A | #N/A |
| 878 | F | 19-38 | 8725 | 1328 | 0 | 0 | 4276 | 1227 | 0 | 0 |
| 879 | M | 19-38 | 2780 | 897 | 0 | 0 | #N/A | #N/A | #N/A | #N/A |
| 880 | F | 39-82 | 26356 | 24619 | 74 | 1 | 18400 | 22934 | 42 | 1 |
| 881 | F | 19-38 | 1934 | 1444 | 0 | 0 | #N/A | #N/A | #N/A | #N/A |
| 882 | F | 39-82 | 481 | 2055 | 0 | 0 | 3073 | 1768 | 0 | 0 |
| 883 | F | 39-82 | 2464 | 732 | 0 | 0 | 1149 | 490 | 0 | 0 |
| 884 | F | 19-38 | 3194 | 951 | 0 | 0 | #N/A | #N/A | #N/A | #N/A |
| 885 | F | 39-82 | 7835 | 1134 | 0 | 0 | 7410 | 324 | 0 | 0 |
| 886 | F | 19-38 | 91 | 568 | 0 | 0 | 621 | 604 | 0 | 0 |
| 887 | F | 19-38 | 5227 | 2606 | 0 | 0 | 2489 | 1825 | 0 | 0 |
| 888 | F | 19-38 | 4153 | 2567 | 0 | 0 | #N/A | #N/A | #N/A | #N/A |
| 889 | F | 19-38 | 688 | 1061 | 0 | 0 | 1044 | 1519 | 0 | 0 |
| 890 | F | 39-82 | 1641 | 58 | 0 | 0 | #N/A | #N/A | #N/A | #N/A |
| 891 | F | 19-38 | 2434 | 172 | 0 | 0 | #N/A | #N/A | #N/A | #N/A |
| 892 | F | 39-82 | 4568 | 48 | 0 | 0 | 2608 | 823 | 0 | 0 |
| 893 | F | 19-38 | 4738 | 83 | 0 | 0 | #N/A | #N/A | #N/A | #N/A |
| 894 | F | 39-82 | 2826 | 1178 | 0 | 0 | 3020 | 999 | 0 | 0 |
| 895 | F | 19-38 | 762 | 34 | 0 | 0 | #N/A | #N/A | #N/A | #N/A |
| 896 | F | 19-38 | 1461 | 44 | 0 | 0 | 986 | 976 | 0 | 0 |
| 897 | F | 19-38 | 1490 | 1232 | 0 | 0 | #N/A | #N/A | #N/A | #N/A |
| 898 | F | 39-82 | 1864 | 878 | 0 | 0 | 1613 | 987 | 0 | 0 |
| 899 | F | 39-82 | 4544 | 3992 | 0 | 0 | #N/A | #N/A | #N/A | #N/A |
| 900 | F | 39-82 | 2180 | 1131 | 0 | 0 | 1109 | 1284 | 0 | 0 |
| 901 | F | 19-38 | 10215 | 679 | 0 | 0 | 8349 | 1103 | 0 | 0 |
| 902 | F | 39-82 | 1276 | 18535 | 0 | 1 | 817 | 15985 | 5 | 1 |
| 903 | F | 39-82 | 5235 | 897 | 0 | 0 | 2494 | 624 | 0 | 0 |
| 904 | F | 39-82 | 517 | 1045 | 0 | 0 | 733 | 1094 | 0 | 0 |
| 905 | F | 39-82 | 3529 | 2624 | 0 | 0 | 1177 | 487 | 0 | 0 |
| 906 | F | 19-38 | 5451 | 898 | 0 | 0 | 2387 | 445 | 0 | 0 |
| 907 | F | 19-38 | 4901 | 823 | 0 | 0 | 5293 | 867 | 0 | 0 |
| 908 | F | 19-38 | 2102 | 1520 | 0 | 0 | 1484 | 601 | 0 | 0 |
| 909 | F | 19-38 | 168 | 814 | 0 | 0 | 1040 | 624 | 0 | 0 |
| 910 | F | 19-38 | 1427 | 2278 | 0 | 0 | 1288 | 1179 | 0 | 0 |
| 911 | F | 39-82 | 4652 | 1826 | 0 | 0 | 526 | 927 | 0 | 0 |
| 912 | F | 19-38 | 2323 | 671 | 0 | 0 | #N/A | #N/A | #N/A | #N/A |
| 913 | F | 19-38 | 3770 | 6519 | 0 | 0 | 1750 | 4567 | 0 | 0 |
| 914 | F | 39-82 | 2194 | 871 | 0 | 0 | 1069 | 553 | 0 | 0 |
| 915 | F | 19-38 | 3092 | 1676 | 0 | 0 | #N/A | #N/A | #N/A | #N/A |
| 916 | F | 19-38 | 8128 | 884 | 0 | 0 | 3487 | 1214 | 0 | 0 |
| 917 | F | 39-82 | 5729 | 717 | 0 | 0 | 6536 | 971 | 0 | 0 |
| 918 | F | 19-38 | 5135 | 955 | 0 | 0 | 3157 | 1294 | 0 | 0 |
| 919 | H | 19-38 | 2648 | 1493 | 0 | 0 | 2658 | 837 | 0 | 0 |
| 920 | F | 39-82 | 1100 | 1157 | 0 | 0 | 937 | 2727 | 0 | 0 |
| 921 | H | 19-38 | 8085 | 602 | 0 | 0 | #N/A | #N/A | #N/A | #N/A |
| 922 | F | 19-38 | 7816 | 1730 | 0 | 0 | 3192 | 2041 | 0 | 0 |
| 923 | H | 19-38 | 4423 | 693 | 0 | 0 | #N/A | #N/A | #N/A | #N/A |
| 924 | F | 39-82 | 3318 | 946 | 0 | 0 | #N/A | #N/A | #N/A | #N/A |
| 925 | F | 39-82 | 4695 | 1558 | 0 | 0 | #N/A | #N/A | #N/A | #N/A |
| 926 | F | 19-38 | 5323 | 621 | 0 | 0 | 6128 | 1219 | 0 | 0 |
| 927 | F | 19-38 | 40869 | 81352 | 90 | 1 | 24789 | 48205 | 48 | 1 |
| 928 | F | 19-38 | 2136 | 770 | 0 | 0 | 1260 | 693 | 0 | 0 |
| 929 | F | 19-38 | 4026 | 3689 | 0 | 0 | 5088 | 2556 | 0 | 0 |
| 930 | H | 19-38 | 3289 | 580 | 0 | 0 | 1459 | 755 | 0 | 0 |
| 931 | F | 39-82 | 4935 | 1280 | 0 | 0 | 1711 | 1457 | 0 | 0 |

| | | | | | | | | | | | | | |
|---|---|---|---|---|---|---|---|---|---|---|---|---|---|---|
| 855 | 1 | 1 | 0 | 1 | 0 | 0 | 0 | 1 | 1 | 0 | 1 | 0 | 1 | 0 |
| 856 | 1 | 0 | 0 | 0 | 1 | 0 | 0 | 0 | 0 | 0 | 1 | 0 | 0 | |
| 857 | 1 | 1 | 1 | 1 | 0 | 1 | 1 | 0 | 1 | 0 | 1 | 0 | 0 | |
| 858 | 0 | 0 | 0 | 0 | 0 | 0 | 0 | 0 | 0 | 0 | 0 | 0 | 0 | |
| 859 | 1 | 0 | 1 | 0 | 0 | 0 | 0 | 1 | 0 | 0 | 0 | 0 | 0 | |
| 860 | 1 | 0 | 1 | 0 | 1 | 1 | 0 | 0 | 0 | 0 | 1 | 1 | 0 | |
| 861 | 1 | 0 | 0 | 0 | 0 | 1 | 0 | 0 | 0 | 0 | 0 | 1 | 0 | |
| 862 | 1 | 1 | 1 | 1 | 0 | 0 | 1 | 0 | 1 | 0 | 1 | 1 | 0 | |
| 863 | 1 | 1 | 1 | 1 | 1 | 1 | 1 | 0 | 1 | 0 | 1 | 1 | 0 | |
| 864 | 1 | 1 | 1 | 0 | 1 | 1 | 1 | 0 | 1 | 1 | 1 | 1 | 1 | 1 |
| 865 | 0 | 0 | 0 | 0 | 0 | 0 | 0 | 0 | 0 | 0 | 0 | 0 | 0 | |
| 866 | 1 | 0 | 1 | 0 | 0 | 0 | 0 | 0 | 0 | 0 | 1 | 0 | 0 | |
| 867 | 0 | 0 | 0 | 0 | 0 | 0 | 0 | 0 | 0 | 0 | 0 | 0 | 0 | |
| 868 | 0 | 0 | 0 | 0 | 0 | 0 | 0 | 0 | 0 | 0 | 0 | 0 | 0 | |
| 869 | 0 | 0 | 0 | 0 | 0 | 0 | 0 | 0 | 0 | 0 | 0 | 0 | 0 | |
| 870 | 0 | 0 | 0 | 0 | 0 | 0 | 0 | 0 | 0 | 0 | 0 | 0 | 0 | |
| 871 | 0 | 0 | 0 | 0 | 0 | 0 | 0 | 0 | 0 | 0 | 0 | 0 | 0 | |
| 872 | 1 | 0 | 0 | 0 | 0 | 0 | 0 | 0 | 1 | 0 | 0 | 0 | 0 | |
| 873 | 0 | 0 | 0 | 0 | 0 | 0 | 0 | 0 | 0 | 0 | 0 | 0 | 0 | |
| 874 | 1 | 1 | 1 | 1 | 0 | 0 | 0 | 0 | 0 | 0 | 0 | 0 | 1 | 0 |
| 875 | 0 | 0 | 0 | 0 | 0 | 0 | 0 | 0 | 0 | 0 | 0 | 0 | 0 | |
| 876 | 1 | 1 | 1 | 1 | 1 | 0 | 0 | 0 | 0 | 0 | 0 | 0 | 0 | |
| 877 | 1 | 0 | 0 | 1 | 0 | 0 | 0 | 0 | 1 | 0 | 1 | 0 | 0 | |
| 878 | 0 | 0 | 0 | 0 | 0 | 0 | 0 | 0 | 0 | 0 | 0 | 0 | 0 | |
| 879 | 0 | 0 | 0 | 0 | 0 | 0 | 0 | 0 | 0 | 0 | 0 | 0 | 0 | |
| 880 | 1 | 0 | 0 | 1 | 0 | 0 | 0 | 0 | 1 | 1 | 1 | 1 | 1 | 1 |
| 881 | 1 | 0 | 1 | 0 | 0 | 0 | 0 | 0 | 0 | 0 | 0 | 0 | 0 | |
| 882 | 0 | 0 | 0 | 0 | 0 | 0 | 0 | 0 | 0 | 0 | 0 | 0 | 0 | |
| 883 | 0 | 0 | 0 | 0 | 0 | 0 | 0 | 0 | 0 | 0 | 0 | 0 | 0 | |
| 884 | 0 | 0 | 0 | 0 | 0 | 0 | 0 | 0 | 0 | 0 | 0 | 0 | 0 | |
| 885 | 0 | 0 | 0 | 0 | 0 | 0 | 0 | 0 | 0 | 0 | 0 | 0 | 0 | |
| 886 | 1 | 0 | 1 | 0 | 0 | 0 | 0 | 0 | 0 | 0 | 0 | 0 | 0 | |
| 887 | 1 | 0 | 1 | 0 | 0 | 0 | 0 | 0 | 0 | 0 | 1 | 1 | 0 | |
| 888 | 1 | 1 | 1 | 1 | 0 | 0 | 0 | 0 | 0 | 0 | 1 | 1 | 0 | |
| 889 | 1 | 0 | 1 | 0 | 1 | 1 | 1 | 0 | 0 | 0 | 1 | 1 | 0 | |
| 890 | 0 | 0 | 0 | 0 | 0 | 0 | 0 | 0 | 0 | 0 | 0 | 0 | 0 | |
| 891 | 0 | 0 | 0 | 0 | 0 | 0 | 0 | 0 | 0 | 0 | 0 | 0 | 0 | |
| 892 | 0 | 0 | 0 | 0 | 0 | 0 | 0 | 0 | 0 | 0 | 0 | 0 | 0 | |
| 893 | 1 | 1 | 1 | 0 | 0 | 0 | 1 | 0 | 0 | 0 | 0 | 1 | 0 | |
| 894 | 1 | 0 | 1 | 0 | 0 | 0 | 0 | 0 | 1 | 0 | 1 | 1 | 0 | |
| 895 | 1 | 0 | 1 | 1 | 0 | 0 | 0 | 0 | 0 | 0 | 0 | 1 | 0 | |
| 896 | 0 | 0 | 0 | 0 | 0 | 0 | 0 | 0 | 0 | 0 | 0 | 0 | 0 | |
| 897 | 1 | 1 | 1 | 1 | 1 | 1 | 1 | 0 | 0 | 0 | 1 | 1 | 1 | 0 |
| 898 | 0 | 0 | 0 | 0 | 0 | 0 | 0 | 0 | 0 | 0 | 0 | 0 | 0 | |
| 899 | 0 | 0 | 0 | 0 | 0 | 0 | 0 | 0 | 0 | 0 | 0 | 0 | 0 | |
| 900 | 1 | 0 | 0 | 1 | 1 | 0 | 0 | 1 | 0 | 0 | 1 | 1 | 0 | |
| 901 | 0 | 0 | 0 | 0 | 0 | 0 | 0 | 0 | 0 | 0 | 0 | 0 | 0 | |
| 902 | 0 | 0 | 0 | 0 | 0 | 0 | 0 | 0 | 0 | 0 | 0 | 0 | 0 | |
| 903 | 1 | 1 | 1 | 0 | 0 | 0 | 1 | 0 | 0 | 0 | 1 | 0 | 0 | |
| 904 | 0 | 0 | 0 | 0 | 0 | 0 | 0 | 0 | 0 | 0 | 0 | 0 | 0 | |
| 905 | 0 | 0 | 0 | 0 | 0 | 0 | 0 | 0 | 0 | 0 | 0 | 0 | 0 | |
| 906 | 1 | 1 | 1 | 0 | 0 | 0 | 0 | 0 | 1 | 0 | 1 | 0 | 0 | |
| 907 | 1 | 0 | 1 | 0 | 0 | 0 | 0 | 0 | 0 | 0 | 0 | 0 | 0 | |
| 908 | 1 | 0 | 1 | 1 | 0 | 0 | 0 | 0 | 0 | 0 | 0 | 0 | 0 | |
| 909 | 0 | 0 | 0 | 0 | 0 | 0 | 0 | 0 | 0 | 0 | 0 | 0 | 0 | |
| 910 | 0 | 0 | 0 | 0 | 0 | 0 | 0 | 0 | 0 | 0 | 0 | 0 | 0 | |
| 911 | 0 | 0 | 0 | 0 | 0 | 0 | 0 | 0 | 0 | 0 | 0 | 0 | 0 | |
| 912 | 1 | 0 | 1 | 0 | 0 | 0 | 0 | 0 | 0 | 0 | 1 | 1 | 0 | |
| 913 | 1 | 0 | 0 | 0 | 0 | 0 | 0 | 0 | 1 | 0 | 1 | 0 | 0 | |
| 914 | 0 | 0 | 0 | 0 | 0 | 0 | 0 | 0 | 0 | 0 | 0 | 0 | 0 | |
| 915 | 0 | 0 | 0 | 1 | 0 | 0 | 0 | 0 | 0 | 0 | 0 | 0 | 0 | |
| 916 | 0 | 0 | 0 | 0 | 0 | 0 | 0 | 0 | 0 | 0 | 0 | 0 | 0 | |
| 917 | 1 | 0 | 1 | 0 | 0 | 0 | 0 | 0 | 0 | 0 | 1 | 0 | 0 | |
| 918 | 1 | 0 | 1 | 0 | 0 | 0 | 0 | 0 | 1 | 0 | 1 | 0 | 0 | |
| 919 | 1 | 1 | 1 | 0 | 0 | 0 | 1 | 0 | 0 | 0 | 1 | 0 | 0 | |
| 920 | 1 | 0 | 1 | 0 | 1 | 0 | 0 | 0 | 0 | 0 | 1 | 0 | 0 | |
| 921 | 0 | 0 | 0 | 0 | 0 | 0 | 0 | 0 | 0 | 0 | 0 | 0 | 0 | |
| 922 | 1 | 1 | 1 | 0 | 0 | 0 | 0 | 1 | 0 | 0 | 1 | 0 | 0 | |
| 923 | 1 | 0 | 0 | 0 | 0 | 0 | 0 | 0 | 0 | 0 | 1 | 0 | 0 | |
| 924 | 0 | 0 | 0 | 0 | 0 | 0 | 0 | 0 | 0 | 0 | 0 | 0 | 0 | |
| 925 | 1 | 0 | 0 | 0 | 0 | 0 | 0 | 0 | 0 | 0 | 1 | 0 | 0 | |
| 926 | 0 | 0 | 0 | 0 | 0 | 0 | 0 | 0 | 0 | 0 | 0 | 0 | 0 | |
| 927 | 1 | 0 | 1 | 1 | 1 | 1 | 1 | 0 | 1 | 1 | 1 | 0 | 0 | |
| 928 | 0 | 0 | 0 | 0 | 0 | 0 | 0 | 0 | 0 | 0 | 0 | 0 | 0 | |
| 929 | 1 | 1 | 1 | 1 | 0 | 0 | 0 | 0 | 0 | 0 | 1 | 0 | 0 | |
| 930 | 1 | 1 | 0 | 1 | 0 | 0 | 0 | 0 | 1 | 0 | 1 | 0 | 0 | |
| 931 | 1 | 0 | 0 | 0 | 0 | 0 | 1 | 0 | 0 | 0 | 1 | 1 | 0 | |

| | | | | | | | | | | | | | | | | | | | | | | | | | |
|---|---|---|---|---|---|---|---|---|---|---|---|---|---|---|---|---|---|---|---|---|---|---|---|---|---|
| 932 | F | 39-82 | | 4226 | 314 | 0 | 0 | 778 | 219 | 0 | 0 | 1 | 0 | 0 | 1 | 0 | 0 | 0 | 0 | 1 | 0 | 0 | 0 | 0 | |
| 933 | F | 19-38 | | 5516 | 1298 | 0 | 0 | #N/A | #N/A | #N/A | #N/A | 0 | 0 | 0 | 0 | 0 | 0 | 0 | 0 | 0 | 0 | 0 | 0 | 0 | |
| 934 | F | 39-82 | | 3058 | 1001 | 0 | 0 | 1082 | 972 | 0 | 0 | 0 | 0 | 0 | 0 | 0 | 0 | 0 | 0 | 0 | 0 | 0 | 0 | 0 | |
| 935 | F | 39-82 | | 12552 | 1139 | 0 | 1 | #N/A | #N/A | #N/A | #N/A | 1 | 0 | 1 | 0 | 0 | 0 | 0 | 0 | 0 | 0 | 1 | 1 | 0 | |
| 936 | F | 19-38 | | 3787 | 985 | 0 | 0 | 2958 | 990 | 0 | 0 | 0 | 0 | 0 | 0 | 0 | 0 | 0 | 0 | 0 | 0 | 0 | 0 | 0 | |
| 937 | F | 39-82 | | 2852 | 2586 | 0 | 0 | 3376 | 1733 | 0 | 0 | 0 | 0 | 0 | 0 | 0 | 0 | 0 | 0 | 0 | 0 | 0 | 0 | 0 | |
| 938 | F | 19-38 | | 5089 | 3596 | 0 | 0 | 2762 | 3279 | 0 | 0 | 1 | 0 | 1 | 0 | 0 | 0 | 0 | 0 | 0 | 0 | 1 | 0 | 0 | |
| 939 | F | 19-38 | | 97299 | 160819 | 96 | 1 | 54270 | 123514 | 5 | 1 | 1 | 1 | 1 | 1 | 1 | 1 | 1 | 0 | 1 | 1 | 1 | 1 | 1 | 1 |
| 940 | M | 39-82 | | 107697 | 95229 | 98 | 1 | 108664 | 67442 | 99 | 1 | 1 | 1 | 1 | 1 | 1 | 1 | 1 | 0 | 1 | 1 | 1 | 0 | 1 | 1 |
| 941 | F | 39-82 | | 5501 | 1378 | 0 | 0 | 4317 | 2421 | 0 | 0 | 0 | 0 | 0 | 0 | 0 | 0 | 0 | 0 | 0 | 0 | 0 | 0 | 0 | |
| 942 | F | 39-82 | | 1924 | 910 | 0 | 0 | 1600 | 1506 | 0 | 0 | 0 | 0 | 0 | 0 | 0 | 0 | 0 | 0 | 0 | 0 | 0 | 0 | 0 | |
| 943 | M | 19-38 | | 3224 | 655 | 0 | 0 | #N/A | #N/A | #N/A | #N/A | 1 | 0 | 0 | 0 | 1 | 1 | 0 | 0 | 0 | 0 | 1 | 0 | 0 | |
| 944 | M | 19-38 | | 2086 | 1007 | 0 | 0 | 1411 | 1605 | 0 | 0 | 1 | 0 | 0 | 0 | 0 | 0 | 0 | 0 | 0 | 0 | 0 | 1 | 0 | |
| 945 | M | 19-38 | | 5802 | 6921 | 0 | 0 | 1993 | 1684 | 0 | 0 | 0 | 0 | 0 | 0 | 0 | 0 | 0 | 0 | 0 | 0 | 0 | 0 | 0 | |
| 946 | F | 19-38 | | 6503 | 1860 | 0 | 0 | 724 | 866 | 0 | 0 | 1 | 0 | 1 | 0 | 0 | 0 | 0 | 0 | 1 | 0 | 1 | 0 | 0 | |
| 947 | F | 19-38 | | 4616 | 3695 | 0 | 0 | 3345 | 4181 | 0 | 0 | 0 | 0 | 0 | 0 | 0 | 0 | 0 | 0 | 0 | 0 | 0 | 0 | 0 | |
| 948 | F | 19-38 | | 2201 | 851 | 0 | 0 | 1306 | 686 | 0 | 0 | 0 | 0 | 0 | 0 | 0 | 0 | 0 | 0 | 0 | 0 | 0 | 0 | 0 | |
| 949 | F | 19-38 | | 1527 | 2007 | 0 | 0 | #N/A | #N/A | #N/A | #N/A | 0 | 0 | 0 | 0 | 0 | 0 | 0 | 0 | 0 | 0 | 0 | 0 | 0 | |
| 950 | M | 19-38 | | 5019 | 643 | 0 | 0 | #N/A | #N/A | #N/A | #N/A | 1 | 0 | 1 | 1 | 0 | 0 | 0 | 0 | 0 | 0 | 0 | 0 | 0 | |
| 951 | F | 19-38 | | 761 | 1024 | 0 | 0 | 694 | 640 | 0 | 0 | 1 | 0 | 1 | 0 | 0 | 0 | 0 | 0 | 0 | 0 | 0 | 0 | 0 | |
| 952 | F | 39-82 | | 1488 | 422 | 0 | 0 | 1061 | 487 | 0 | 0 | 1 | 0 | 1 | 0 | 0 | 0 | 1 | 0 | 1 | 0 | 1 | 1 | 0 | |
| 953 | F | 19-38 | | 1979 | 945 | 0 | 0 | 1450 | 1119 | 0 | 0 | 1 | 0 | 1 | 1 | 1 | 1 | 0 | 0 | 1 | 0 | 0 | 0 | 0 | |
| 954 | F | 19-38 | | 54369 | 10988 | 33 | 1 | 6504 | 64159 | 0 | 1 | 1 | 1 | 1 | 1 | 0 | 0 | 1 | 1 | 0 | 1 | 1 | 0 | 0 | |
| 955 | F | 19-38 | | 3871 | 979 | 0 | 0 | #N/A | #N/A | #N/A | #N/A | 1 | 0 | 1 | 0 | 0 | 0 | 0 | 0 | 0 | 0 | 1 | 0 | 0 | |
| 956 | F | 19-38 | | 5010 | 1409 | 0 | 0 | 620 | 855 | 0 | 0 | 1 | 0 | 0 | 0 | 0 | 0 | 0 | 0 | 0 | 0 | 1 | 0 | 0 | |
| 957 | H | 19-38 | | 774 | 910 | 0 | 0 | #N/A | #N/A | #N/A | #N/A | 1 | 0 | 1 | 0 | 0 | 0 | 1 | 0 | 0 | 0 | 1 | 0 | 0 | |
| 958 | H | 19-38 | | 3281 | 4630 | 0 | 0 | #N/A | #N/A | #N/A | #N/A | 0 | 0 | 0 | 0 | 0 | 0 | 0 | 0 | 0 | 0 | 0 | 0 | 0 | |
| 959 | F | 39-82 | | 1540 | 750 | 0 | 0 | 715 | 724 | 0 | 0 | 1 | 1 | 1 | 1 | 0 | 0 | 0 | 1 | 0 | 0 | 1 | 1 | 0 | |
| 960 | F | 39-82 | | 1440 | 1238 | 0 | 0 | 1688 | 1708 | 0 | 0 | 0 | 0 | 0 | 0 | 0 | 0 | 0 | 0 | 0 | 0 | 0 | 0 | 0 | |
| 961 | H | 19-38 | | 4608 | 844 | 0 | 0 | 2419 | 5259 | 0 | 0 | 1 | 0 | 0 | 0 | 0 | 0 | 0 | 0 | 0 | 0 | 1 | 0 | 0 | |
| 962 | F | 39-82 | | 6826 | 2805 | 0 | 0 | 6989 | 2354 | 0 | 0 | 0 | 0 | 0 | 0 | 0 | 0 | 0 | 0 | 0 | 0 | 0 | 0 | 0 | |
| 963 | F | 39-82 | | 845 | 2753 | 0 | 0 | #N/A | #N/A | #N/A | #N/A | 0 | 0 | 0 | 0 | 0 | 0 | 0 | 0 | 0 | 0 | 0 | 0 | 0 | |
| 964 | F | 39-82 | | 714 | 788 | 0 | 0 | 613 | 363 | 0 | 0 | 1 | 1 | 0 | 0 | 0 | 0 | 0 | 0 | 0 | 0 | 0 | 0 | 0 | |
| 965 | H | 19-38 | | 1731 | 9055 | 0 | 1 | 2205 | 6828 | 0 | 0 | 1 | 1 | 1 | 0 | 0 | 0 | 1 | 0 | 1 | 0 | 1 | 0 | 0 | |
| 966 | F | 19-38 | | 118590 | 74752 | 94 | 1 | #N/A | #N/A | #N/A | #N/A | 0 | 0 | 0 | 0 | 0 | 0 | 0 | 0 | 0 | 0 | 0 | 0 | 0 | |
| 967 | F | 39-82 | | 915 | 876 | 0 | 0 | 945 | 789 | 0 | 0 | 1 | 1 | 1 | 1 | 1 | 0 | 1 | 0 | 0 | 0 | 1 | 0 | 0 | |
| 968 | F | 19-38 | | 3671 | 1148 | 0 | 0 | 943 | 350 | 0 | 0 | 0 | 0 | 0 | 0 | 0 | 0 | 0 | 0 | 0 | 0 | 0 | 0 | 0 | |
| 969 | F | 39-82 | | 2076 | 961 | 0 | 0 | 1932 | 656 | 0 | 0 | 1 | 1 | 1 | 1 | 1 | 0 | 0 | 0 | 1 | 0 | 0 | 0 | 0 | |
| 970 | F | 39-82 | | 37 | 334 | 0 | 0 | #N/A | #N/A | #N/A | #N/A | 1 | 0 | 0 | 0 | 0 | 0 | 0 | 0 | 0 | 0 | 1 | 0 | 1 | 0 |
| 971 | F | 19-38 | | 1177 | 1816 | 0 | 0 | 1647 | 1010 | 0 | 0 | 0 | 0 | 0 | 0 | 0 | 0 | 0 | 0 | 0 | 0 | 0 | 0 | 0 | |
| 972 | F | 39-82 | | 948 | 972 | 0 | 0 | #N/A | #N/A | #N/A | #N/A | 1 | 1 | 1 | 1 | 1 | 1 | 1 | 0 | 0 | 0 | 1 | 0 | 1 | 0 |
| 973 | H | 19-38 | | 5995 | 1279 | 0 | 0 | 2543 | 959 | 0 | 0 | 1 | 0 | 0 | 0 | 0 | 0 | 0 | 1 | 0 | 0 | 0 | 0 | 0 | |
| 974 | F | 39-82 | | 720 | 796 | 0 | 0 | 3211 | 978 | 0 | 0 | 0 | 0 | 0 | 0 | 0 | 0 | 0 | 0 | 0 | 0 | 0 | 0 | 0 | |
| 975 | F | 39-82 | | 402 | 1387 | 0 | 0 | #N/A | #N/A | #N/A | #N/A | 0 | 0 | 0 | 0 | 0 | 0 | 0 | 0 | 0 | 0 | 0 | 0 | 0 | |
| 976 | F | 39-82 | | 88851 | 255794 | 98 | 1 | 53064 | 159393 | 54 | 1 | 1 | 1 | 1 | 0 | 1 | 1 | 1 | 0 | 0 | 0 | 0 | 0 | 1 | 1 |
| 977 | F | 39-82 | | 9226 | 5798 | 0 | 0 | 4872 | 647 | 0 | 0 | 1 | 0 | 0 | 1 | 0 | 1 | 0 | 0 | 1 | 0 | 1 | 0 | 0 | |
| 978 | H | 19-38 | | 164581 | 150164 | 99 | 1 | 24858 | 121270 | 8 | 1 | 1 | 1 | 1 | 1 | 0 | 0 | 1 | 0 | 0 | 1 | 1 | 1 | 0 | |
| 979 | F | 19-38 | | 4144 | 2802 | 0 | 0 | 809 | 652 | 0 | 0 | 0 | 0 | 0 | 0 | 0 | 0 | 0 | 0 | 0 | 0 | 0 | 0 | 0 | |
| 980 | F | 39-82 | neg | 8090 | 2350 | 0 | 0 | 1374 | 536 | 0 | 0 | 1 | 1 | 1 | 1 | 0 | 0 | 0 | 0 | 0 | 1 | 0 | 1 | 0 | |
| 981 | H | 19-38 | | 6188 | 2639 | 0 | 0 | 567 | 287 | 0 | 0 | 0 | 0 | 0 | 0 | 0 | 0 | 0 | 0 | 0 | 0 | 0 | 0 | 0 | |
| 982 | H | 39-82 | | 5161 | 576 | 0 | 0 | 4880 | 334 | 0 | 0 | 0 | 0 | 0 | 0 | 0 | 0 | 0 | 0 | 0 | 0 | 0 | 0 | 0 | |
| 983 | H | 19-38 | | 336 | 1064 | 0 | 0 | #N/A | #N/A | #N/A | #N/A | 1 | 0 | 0 | 1 | 0 | 0 | 0 | 0 | 1 | 0 | 0 | 0 | 0 | |
| 984 | F | 39-82 | | 9656 | 2019 | 0 | 0 | 4616 | 798 | 0 | 0 | 1 | 1 | 1 | 1 | 1 | 1 | 0 | 0 | 0 | 0 | 1 | 0 | 0 | |
| 985 | F | 39-82 | | 5154 | 2921 | 0 | 0 | #N/A | #N/A | #N/A | #N/A | 0 | 0 | 0 | 0 | 0 | 0 | 0 | 0 | 0 | 0 | 0 | 0 | 0 | |
| 986 | H | 19-38 | | 5350 | 2198 | 0 | 0 | #N/A | #N/A | #N/A | #N/A | 0 | 0 | 0 | 0 | 0 | 0 | 0 | 0 | 0 | 0 | 0 | 0 | 0 | |
| 987 | F | 39-82 | | 1520 | 2109 | 0 | 0 | 642 | 966 | 0 | 0 | 1 | 1 | 1 | 1 | 0 | 0 | 0 | 0 | 0 | 0 | 0 | 0 | 0 | |
| 988 | F | 39-82 | | 2337 | 1271 | 0 | 0 | 1661 | 541 | 0 | 0 | 1 | 0 | 1 | 1 | 0 | 0 | 1 | 0 | 1 | 0 | 0 | 0 | 0 | |
| 989 | F | 19-38 | | 390 | 2612 | 0 | 0 | #N/A | #N/A | #N/A | #N/A | 0 | 0 | 0 | 0 | 0 | 0 | 0 | 0 | 0 | 0 | 0 | 0 | 0 | |
| 990 | H | 19-38 | | 3391 | 1710 | 0 | 0 | #N/A | #N/A | #N/A | #N/A | 1 | 0 | 1 | 0 | 0 | 0 | 0 | 0 | 0 | 0 | 0 | 1 | 0 | |
| 991 | M | 19-38 | | 2759 | 2045 | 0 | 0 | #N/A | #N/A | #N/A | #N/A | 1 | 0 | 1 | 0 | 0 | 0 | 1 | 0 | 0 | 0 | 1 | 0 | 0 | |
| 992 | M | 39-82 | | 9793 | 1475 | 0 | 0 | #N/A | #N/A | #N/A | #N/A | 1 | 1 | 1 | 0 | 1 | 1 | 1 | 0 | 1 | 0 | 1 | 1 | 0 | |
| 993 | M | 39-82 | | 8792 | 1902 | 0 | 0 | 7774 | 5712 | 0 | 0 | 0 | 0 | 0 | 1 | 0 | 0 | 0 | 0 | 0 | 0 | 0 | 0 | 0 | |
| 994 | F | 39-82 | | 2328 | 754 | 0 | 0 | #N/A | #N/A | #N/A | #N/A | 1 | 0 | 0 | 1 | 1 | 0 | 0 | 1 | 0 | 0 | 0 | 1 | 0 | |
| 995 | F | 39-82 | | 9311 | 1475 | 0 | 0 | #N/A | #N/A | #N/A | #N/A | 1 | 1 | 1 | 1 | 0 | 0 | 1 | 0 | 1 | 0 | 0 | 0 | 1 | 0 |
| 996 | F | 39-82 | | 2529 | 1281 | 0 | 0 | 1712 | 1860 | 0 | 0 | 1 | 0 | 0 | 0 | 0 | 0 | 0 | 0 | 0 | 0 | 1 | 1 | 0 | |
| 997 | F | 19-38 | | 2016 | 867 | 0 | 0 | #N/A | #N/A | #N/A | #N/A | 0 | 0 | 0 | 0 | 0 | 0 | 0 | 0 | 0 | 0 | 0 | 0 | 0 | |
| 998 | F | 39-82 | | 756 | 2128 | 0 | 0 | 1705 | 1920 | 0 | 0 | 0 | 0 | 0 | 0 | 0 | 0 | 0 | 0 | 0 | 0 | 0 | 0 | 0 | |
| 999 | M | 39-82 | | 5815 | 2733 | 0 | 0 | #N/A | #N/A | #N/A | #N/A | 1 | 0 | 0 | 1 | 0 | 0 | 0 | 1 | 0 | 0 | 1 | 0 | 0 | |
| 1000 | M | 19-38 | | 2403 | 1742 | 0 | 0 | #N/A | #N/A | #N/A | #N/A | 1 | 0 | 0 | 1 | 0 | 1 | 0 | 0 | 1 | 0 | 0 | 0 | 1 | 0 |
| 1001 | M | 39-82 | | 1339 | 2140 | 0 | 0 | 1259 | 1762 | 0 | 0 | 1 | 0 | 1 | 0 | 0 | 0 | 0 | 0 | 1 | 0 | 0 | 0 | 0 | |
| 1002 | M | 39-82 | | 1981 | 970 | 0 | 0 | #N/A | #N/A | #N/A | #N/A | 0 | 0 | 0 | 0 | 0 | 0 | 0 | 0 | 0 | 0 | 0 | 0 | 0 | |
| 1003 | M | 19-38 | | 8593 | 1357 | 0 | 0 | 6432 | 871 | 0 | 0 | 1 | 0 | 1 | 0 | 0 | 1 | 1 | 1 | 0 | 0 | 1 | 0 | 0 | |
| 1004 | F | 19-38 | | 1348 | 1180 | 0 | 0 | 1363 | 1449 | 0 | 0 | 0 | 0 | 0 | 0 | 0 | 0 | 0 | 0 | 0 | 0 | 0 | 0 | 0 | |
| 1005 | F | 19-38 | | 26654 | 14957 | 41 | 1 | 20233 | 22369 | 5 | 1 | 1 | 1 | 1 | 0 | 1 | 1 | 1 | 0 | 0 | 1 | 1 | 1 | 0 | |
| 1006 | M | 19-38 | | 1912 | 394 | 0 | 0 | #N/A | #N/A | #N/A | #N/A | 0 | 0 | 0 | 0 | 0 | 0 | 0 | 0 | 0 | 0 | 0 | 0 | 0 | |
| 1007 | F | 19-38 | | 2717 | 979 | 0 | 0 | 2849 | 876 | 0 | 0 | 0 | 0 | 0 | 0 | 0 | 0 | 0 | 0 | 0 | 0 | 0 | 0 | 0 | |
| 1008 | M | 39-82 | | 4049 | 1486 | 0 | 0 | 2223 | 2740 | 0 | 0 | 0 | 0 | 0 | 0 | 0 | 0 | 0 | 0 | 0 | 0 | 0 | 0 | 0 | |

| | | | | | | | | | | | | | | | | | | | | | | | | | |
|---|---|---|---|---|---|---|---|---|---|---|---|---|---|---|---|---|---|---|---|---|---|---|---|---|---|
| 1009 | F | 39-82 | | 4743 | 718 | 0 | 0 | 2558 | 924 | 0 | 0 | 0 | 0 | 0 | 0 | 0 | 0 | 0 | 0 | 0 | 0 | 0 | 0 | 0 | |
| 1010 | F | 39-82 | | 3200 | 1221 | 0 | 0 | 2749 | 1159 | 0 | 0 | 0 | 0 | 0 | 0 | 0 | 0 | 0 | 0 | 0 | 0 | 0 | 0 | 0 | |
| 1011 | M | 19-38 | | 2655 | 983 | 0 | 0 | 998 | 383 | 0 | 0 | 0 | 0 | 0 | 0 | 0 | 0 | 0 | 0 | 0 | 0 | 0 | 0 | 0 | |
| 1012 | F | 39-82 | IgM positive | 7059 | 190 | 0 | 0 | 339 | 337 | 0 | 0 | 1 | 0 | 1 | 1 | 0 | 1 | 1 | 1 | 0 | 1 | 1 | 1 | 1 | 0 |
| 1013 | F | 19-38 | | 2611 | 684 | 0 | 0 | 2248 | 674 | 0 | 0 | 0 | 0 | 0 | 0 | 0 | 0 | 0 | 0 | 0 | 0 | 0 | 0 | 0 | |
| 1014 | F | 19-38 | | 964 | 687 | 0 | 0 | 1095 | 1560 | 0 | 0 | 1 | 0 | 1 | 0 | 0 | 0 | 0 | 0 | 0 | 0 | 0 | 0 | 0 | |
| 1015 | F | 39-82 | | 2826 | 166 | 0 | 0 | #N/A | #N/A | #N/A | #N/A | 0 | 0 | 0 | 0 | 0 | 0 | 0 | 0 | 0 | 0 | 0 | 0 | 0 | |
| 1016 | M | 19-38 | | 7605 | 846 | 0 | 0 | 7237 | 472 | 0 | 0 | 0 | 0 | 0 | 0 | 0 | 0 | 0 | 0 | 0 | 0 | 0 | 0 | 0 | |
| 1017 | F | 39-82 | | 8429 | 104422 | 97 | 1 | 4709 | 1471 | 76 | 1 | 1 | 0 | 1 | 0 | 0 | 0 | 0 | 0 | 0 | 1 | 1 | 0 | 0 | |
| 1018 | F | 19-38 | | 8348 | 105 | 0 | 0 | #N/A | #N/A | #N/A | #N/A | 0 | 0 | 0 | 0 | 0 | 0 | 0 | 0 | 0 | 0 | 0 | 0 | 0 | |
| 1019 | F | 19-38 | | 5197 | 1888 | 0 | 0 | 169 | 128 | 0 | 0 | 1 | 1 | 1 | 1 | 1 | 1 | 1 | 0 | 1 | 0 | 1 | 1 | 1 | 0 |
| 1020 | M | 39-82 | | 1091 | 486 | 0 | 0 | 468 | 726 | 0 | 0 | 0 | 0 | 0 | 0 | 0 | 0 | 0 | 0 | 0 | 0 | 0 | 0 | 0 | |
| 1021 | M | 39-82 | | 215250 | 170977 | 94 | 1 | 154353 | 111074 | 81 | 1 | 1 | 1 | 1 | 1 | 0 | 0 | 1 | 0 | 1 | 1 | 1 | 1 | 1 | 1 |
| 1022 | F | 19-38 | | 9213 | 1384 | 0 | 0 | 4522 | 1135 | 0 | 0 | 0 | 0 | 0 | 0 | 0 | 0 | 0 | 0 | 0 | 0 | 0 | 0 | 0 | |
| 1023 | F | 19-38 | | 6749 | 2387 | 0 | 0 | 2938 | 2771 | 0 | 0 | 1 | 0 | 1 | 0 | 0 | 0 | 0 | 0 | 0 | 0 | 1 | 0 | 0 | |
| 1024 | M | 19-38 | | 76525 | 69513 | 88 | 1 | 34178 | 72015 | 23 | 1 | 1 | 0 | 1 | 1 | 1 | 1 | 1 | 0 | 1 | 1 | 1 | 0 | 0 | |
| 1025 | H | 39-82 | | 6302 | 1764 | 0 | 0 | #N/A | #N/A | #N/A | #N/A | 1 | 0 | 0 | 0 | 0 | 0 | 0 | 0 | 0 | 0 | 1 | 0 | 0 | |
| 1026 | F | 19-38 | | 3679 | 422 | 0 | 0 | 845 | 498 | 0 | 0 | 0 | 0 | 0 | 0 | 0 | 0 | 0 | 0 | 0 | 0 | 0 | 0 | 0 | |
| 1027 | M | 39-82 | | 3970 | 3112 | 0 | 0 | #N/A | #N/A | #N/A | #N/A | 0 | 0 | 0 | 0 | 0 | 0 | 0 | 0 | 0 | 0 | 0 | 0 | 0 | |
| 1028 | M | 39-82 | | 1858 | 478 | 0 | 0 | 2229 | 703 | 0 | 0 | 0 | 0 | 0 | 0 | 0 | 0 | 0 | 0 | 0 | 0 | 0 | 0 | 0 | |
| 1029 | F | 39-82 | | 4094 | 820 | 0 | 0 | 1996 | 1006 | 0 | 0 | 0 | 0 | 0 | 0 | 0 | 0 | 0 | 0 | 0 | 0 | 0 | 0 | 0 | |
| 1030 | F | 39-82 | | 6357 | 2116 | 0 | 0 | 2201 | 984 | 0 | 0 | 1 | 0 | 0 | 0 | 0 | 0 | 0 | 0 | 1 | 0 | 0 | 0 | 0 | |
| 1031 | F | 19-38 | | 5687 | 1254 | 0 | 0 | 4478 | 618 | 0 | 0 | 1 | 0 | 0 | 0 | 1 | 0 | 0 | 0 | 0 | 0 | 1 | 0 | 0 | |
| 1032 | F | 39-82 | | 7382 | 2343 | 0 | 0 | 4548 | 3708 | 0 | 0 | 1 | 1 | 1 | 0 | 1 | 0 | 1 | 0 | 0 | 0 | 1 | 0 | 0 | |
| 1033 | F | 39-82 | | 237 | 928 | 0 | 0 | 543 | 580 | 0 | 0 | 0 | 0 | 0 | 0 | 0 | 0 | 0 | 0 | 0 | 0 | 0 | 0 | 0 | |
| 1034 | M | 39-82 | | 3445 | 3447 | 0 | 0 | #N/A | #N/A | #N/A | #N/A | 0 | 0 | 0 | 0 | 0 | 0 | 0 | 0 | 0 | 0 | 0 | 0 | 0 | |
| 1035 | F | 19-38 | | 4561 | 2171 | 0 | 0 | #N/A | #N/A | #N/A | #N/A | 1 | 0 | 1 | 0 | 0 | 0 | 0 | 0 | 0 | 0 | 1 | 0 | 0 | |
| 1036 | F | 39-82 | | 5416 | 752 | 0 | 0 | 2872 | 612 | 0 | 0 | 0 | 0 | 0 | 0 | 0 | 0 | 0 | 0 | 0 | 0 | 0 | 0 | 0 | |
| 1037 | F | 19-38 | | 1844 | 626 | 0 | 0 | 1763 | 911 | 0 | 0 | 0 | 0 | 0 | 0 | 0 | 0 | 0 | 0 | 0 | 0 | 0 | 0 | 0 | |
| 1038 | F | 19-38 | | 9083 | 1290 | 0 | 0 | 630 | 276 | 0 | 0 | 0 | 0 | 0 | 0 | 0 | 0 | 0 | 0 | 0 | 0 | 0 | 0 | 0 | |
| 1039 | F | 19-38 | | 1508 | 2410 | 0 | 0 | #N/A | #N/A | #N/A | #N/A | 0 | 0 | 0 | 0 | 0 | 0 | 0 | 0 | 0 | 0 | 0 | 0 | 0 | |
| 1040 | F | 19-38 | | 1349 | 751 | 0 | 0 | #N/A | #N/A | #N/A | #N/A | 0 | 0 | 0 | 0 | 0 | 0 | 0 | 0 | 0 | 0 | 0 | 0 | 0 | |
| 1041 | F | 19-38 | | 7743 | 672 | 0 | 0 | 5572 | 2115 | 0 | 0 | 1 | 0 | 1 | 1 | 0 | 0 | 0 | 0 | 1 | 0 | 1 | 0 | 0 | |
| 1042 | F | 39-82 | | 2982 | 436 | 0 | 0 | 1356 | 1744 | 0 | 0 | 0 | 0 | 0 | 0 | 0 | 0 | 0 | 0 | 0 | 0 | 0 | 0 | 0 | |
| 1043 | F | 39-82 | | 3437 | 1480 | 0 | 0 | 1746 | 615 | 0 | 0 | 0 | 0 | 0 | 0 | 0 | 0 | 0 | 0 | 0 | 0 | 0 | 0 | 1 | |
| 1044 | F | 19-38 | | 4512 | 1975 | 0 | 0 | 1423 | 1677 | 0 | 0 | 0 | 0 | 0 | 0 | 0 | 0 | 0 | 0 | 0 | 0 | 0 | 0 | 0 | |
| 1045 | F | 19-38 | | 1021 | 1195 | 0 | 0 | #N/A | #N/A | #N/A | #N/A | 1 | 0 | 1 | 0 | 0 | 0 | 0 | 0 | 0 | 0 | 1 | 0 | 0 | |
| 1046 | F | 19-38 | | 5863 | 2765 | 0 | 0 | 3218 | 1698 | 0 | 0 | 1 | 0 | 0 | 0 | 0 | 0 | 0 | 1 | 1 | 0 | 0 | 0 | 0 | |
| 1047 | F | 39-82 | | 7864 | 2959 | 0 | 0 | #N/A | #N/A | #N/A | #N/A | 1 | 0 | 1 | 0 | 0 | 0 | 1 | 0 | 0 | 0 | 0 | 0 | 1 | |
| 1048 | F | 19-38 | | 4667 | 6572 | 0 | 0 | #N/A | #N/A | #N/A | #N/A | 0 | 0 | 0 | 0 | 0 | 0 | 0 | 0 | 0 | 0 | 0 | 0 | 0 | |
| 1049 | F | 19-38 | | 7932 | 5027 | 0 | 0 | 3009 | 610 | 0 | 0 | 0 | 0 | 0 | 0 | 0 | 0 | 0 | 0 | 0 | 0 | 0 | 0 | 0 | |
| 1050 | F | 19-38 | | 181 | 737 | 0 | 0 | 771 | 669 | 0 | 0 | 1 | 0 | 1 | 1 | 0 | 0 | 0 | 0 | 0 | 0 | 1 | 1 | 0 | |
| 1051 | F | 39-82 | | 1235 | 1550 | 0 | 0 | 1463 | 7028 | 0 | 0 | 0 | 0 | 0 | 0 | 0 | 0 | 0 | 0 | 0 | 0 | 0 | 0 | 0 | |
| 1052 | F | 39-82 | | 4506 | 35828 | 58 | 1 | 2651 | 27251 | 0 | 1 | 1 | 1 | 1 | 0 | 0 | 1 | 0 | 0 | 1 | 0 | 1 | 0 | 0 | |
| 1053 | F | 19-38 | | 1256 | 4768 | 0 | 0 | 1617 | 6178 | 0 | 0 | 0 | 0 | 0 | 0 | 0 | 0 | 0 | 0 | 0 | 0 | 0 | 0 | 0 | |
| 1054 | M | 39-82 | | 2291 | 2906 | 0 | 0 | #N/A | #N/A | #N/A | #N/A | 0 | 0 | 0 | 0 | 0 | 0 | 0 | 0 | 0 | 0 | 0 | 0 | 0 | |
| 1055 | F | 19-38 | | 107230 | 129251 | 95 | 1 | 56768 | 213306 | 54 | 1 | 1 | 1 | 1 | 0 | 1 | 1 | 1 | 0 | 1 | 0 | 1 | 0 | 1 | 1 |
| 1056 | F | 39-82 | | 727 | 562 | 0 | 0 | 709 | 899 | 0 | 0 | 0 | 0 | 0 | 0 | 0 | 0 | 0 | 0 | 0 | 0 | 0 | 0 | 0 | |
| 1057 | M | 39-82 | | 1675 | 868 | 0 | 0 | 1500 | 383 | 0 | 0 | 0 | 0 | 0 | 0 | 0 | 0 | 0 | 0 | 0 | 0 | 0 | 0 | 0 | |
| 1058 | F | 19-38 | | 945 | 1128 | 0 | 0 | 506 | 488 | 0 | 0 | 1 | 0 | 0 | 0 | 0 | 0 | 0 | 0 | 1 | 0 | 0 | 0 | 0 | |
| 1059 | F | 39-82 | | 4998 | 2228 | 0 | 0 | 4039 | 1390 | 0 | 0 | 1 | 0 | 0 | 0 | 0 | 0 | 0 | 0 | 0 | 0 | 0 | 1 | 0 | |
| 1060 | F | 39-82 | | 1763 | 1109 | 0 | 0 | 1376 | 1199 | 0 | 0 | 1 | 0 | 1 | 1 | 0 | 0 | 0 | 0 | 1 | 0 | 0 | 0 | 0 | |
| 1061 | F | 19-38 | | 1753 | 1994 | 0 | 0 | 1382 | 834 | 0 | 0 | 1 | 0 | 1 | 0 | 0 | 0 | 0 | 0 | 0 | 0 | 1 | 0 | 0 | |
| 1062 | M | 39-82 | | 7085 | 1070 | 0 | 0 | 9317 | 620 | 0 | 0 | 0 | 0 | 0 | 0 | 0 | 0 | 0 | 0 | 0 | 0 | 0 | 0 | 0 | |
| 1063 | F | 19-38 | | 616 | 1273 | 0 | 0 | 716 | 1161 | 0 | 0 | 0 | 0 | 0 | 0 | 0 | 0 | 0 | 0 | 0 | 0 | 0 | 0 | 0 | |
| 1064 | F | 19-38 | | 239 | 930 | 0 | 0 | 598 | 520 | 0 | 0 | 0 | 0 | 0 | 0 | 0 | 0 | 0 | 0 | 0 | 0 | 0 | 0 | 0 | |
| 1065 | F | 19-38 | | 1176 | 1862 | 0 | 0 | 879 | 1381 | 0 | 0 | 0 | 0 | 0 | 0 | 0 | 0 | 0 | 0 | 0 | 0 | 0 | 0 | 0 | |
| 1066 | F | 39-82 | | 263 | 1556 | 0 | 0 | 904 | 642 | 0 | 0 | 0 | 0 | 0 | 0 | 0 | 0 | 0 | 0 | 0 | 0 | 0 | 0 | 0 | |
| 1067 | F | 39-82 | | 5560 | 983 | 0 | 0 | 4490 | 1325 | 0 | 0 | 1 | 1 | 1 | 0 | 1 | 0 | 0 | 0 | 0 | 0 | 1 | 0 | 0 | |
| 1068 | F | 19-38 | | 3892 | 2098 | 0 | 0 | 1601 | 591 | 0 | 0 | 0 | 0 | 0 | 0 | 0 | 0 | 0 | 0 | 0 | 0 | 0 | 0 | 0 | |
| 1069 | M | 19-38 | | 1201 | 1893 | 0 | 0 | 1343 | 735 | 0 | 0 | 0 | 0 | 0 | 0 | 0 | 0 | 0 | 0 | 0 | 0 | 0 | 0 | 0 | |
| 1070 | F | 19-38 | | 2811 | 2855 | 0 | 0 | 2172 | 1687 | 0 | 0 | 0 | 0 | 0 | 0 | 0 | 0 | 0 | 0 | 0 | 0 | 0 | 0 | 0 | |
| 1071 | F | 19-38 | | 4945 | 973 | 0 | 0 | #N/A | #N/A | #N/A | #N/A | 1 | 1 | 1 | 1 | 1 | 0 | 1 | 0 | 1 | 0 | 1 | 0 | 0 | |
| 1072 | F | 39-82 | | 993 | 1443 | 0 | 0 | 986 | 2804 | 0 | 0 | 0 | 0 | 0 | 0 | 0 | 0 | 0 | 0 | 0 | 0 | 0 | 0 | 0 | |
| 1073 | F | 39-82 | | 1132 | 1283 | 0 | 0 | 789 | 375 | 0 | 0 | 0 | 0 | 0 | 0 | 0 | 0 | 0 | 0 | 0 | 0 | 0 | 0 | 0 | |
| 1074 | F | 39-82 | | 2385 | 1478 | 0 | 0 | 3099 | 1197 | 0 | 0 | 0 | 0 | 0 | 0 | 0 | 0 | 0 | 0 | 0 | 0 | 0 | 0 | 0 | |
| 1075 | F | 39-82 | | 6313 | 1594 | 0 | 0 | 1019 | 1624 | 0 | 0 | 0 | 0 | 0 | 0 | 0 | 0 | 0 | 0 | 0 | 0 | 0 | 0 | 0 | |
| 1076 | F | 19-38 | | 1722 | 842 | 0 | 0 | 1139 | 1391 | 0 | 0 | 0 | 0 | 0 | 0 | 0 | 0 | 0 | 0 | 0 | 0 | 0 | 0 | 0 | |
| 1077 | F | 19-38 | | 5696 | 1084 | 0 | 0 | 2906 | 2217 | 0 | 0 | 1 | 0 | 0 | 0 | 0 | 0 | 0 | 0 | 1 | 0 | 1 | 1 | 0 | |
| 1078 | F | 39-82 | | 2062 | 1395 | 0 | 0 | #N/A | #N/A | #N/A | #N/A | 0 | 0 | 0 | 0 | 0 | 0 | 0 | 0 | 0 | 0 | 0 | 0 | 0 | |
| 1079 | F | 39-82 | | 3851 | 901 | 0 | 0 | 1506 | 1412 | 0 | 0 | 0 | 0 | 0 | 0 | 0 | 0 | 0 | 0 | 0 | 0 | 0 | 0 | 0 | |
| 1080 | M | 39-82 | | 2799 | 826 | 0 | 0 | #N/A | #N/A | #N/A | #N/A | 1 | 1 | 1 | 1 | 0 | 1 | 0 | 0 | 1 | 0 | 1 | 0 | 0 | |
| 1081 | F | 39-82 | | 3420 | 598 | 0 | 0 | 2792 | 781 | 0 | 0 | 0 | 0 | 0 | 0 | 0 | 0 | 0 | 0 | 0 | 0 | 0 | 0 | 0 | |
| 1082 | F | 39-82 | | 2571 | 901 | 0 | 0 | 813 | 450 | 0 | 0 | 1 | 1 | 1 | 0 | 0 | 0 | 0 | 0 | 0 | 0 | 1 | 0 | 0 | |
| 1083 | F | 39-82 | | 5790 | 1129 | 0 | 0 | 2028 | 399 | 0 | 0 | 1 | 0 | 1 | 0 | 0 | 0 | 0 | 0 | 1 | 0 | 1 | 1 | 0 | |
| 1084 | F | 39-82 | | 3081 | 1080 | 0 | 0 | 1387 | 974 | 0 | 0 | 0 | 0 | 0 | 0 | 0 | 0 | 0 | 0 | 0 | 0 | 0 | 0 | 0 | |
| 1085 | F | 19-38 | | 2603 | 1047 | 0 | 0 | #N/A | #N/A | #N/A | #N/A | 0 | 0 | 0 | 0 | 0 | 0 | 0 | 0 | 0 | 0 | 0 | 0 | 0 | |

| | | | | | | | | | | | | | | | | | | | | | | | | | |
|---|---|---|---|---|---|---|---|---|---|---|---|---|---|---|---|---|---|---|---|---|---|---|---|---|---|
| 1086 | M | 19-38 | | 6442 | 1206 | 0 | 0 | 2913 | 671 | 0 | 0 | 1 | 0 | 0 | 0 | 0 | 0 | 0 | 0 | 0 | 0 | 1 | 0 | 0 | |
| 1087 | F | 19-38 | | 2366 | 1709 | 0 | 0 | 399 | 788 | 0 | 0 | 1 | 0 | 0 | 0 | 0 | 0 | 0 | 0 | 0 | 0 | 1 | 0 | 0 | |
| 1088 | M | 19-38 | | 1970 | 1191 | 0 | 0 | #N/A | #N/A | #N/A | #N/A | 1 | 0 | 1 | 1 | 0 | 0 | 0 | 0 | 0 | 0 | 1 | 0 | 1 | 0 |
| 1089 | M | 19-38 | | 4947 | 29294 | 59 | 1 | 4637 | 59063 | 0 | 1 | 1 | 0 | 0 | 0 | 0 | 0 | 0 | 0 | 0 | 1 | 0 | 0 | 0 | |
| 1090 | F | 39-82 | | 1963 | 1254 | 0 | 0 | 1356 | 1032 | 0 | 0 | 1 | 0 | 1 | 0 | 1 | 1 | 1 | 0 | 0 | 0 | 1 | 0 | 1 | |
| 1091 | F | 39-82 | | 69476 | 97921 | 97 | 1 | 90442 | 168451 | 77 | 1 | 1 | 1 | 1 | 1 | 0 | 0 | 1 | 0 | 1 | 0 | 1 | 0 | 1 | 1 |
| 1092 | F | 19-38 | | 1352 | 1024 | 0 | 0 | 450 | 394 | 0 | 0 | 0 | 0 | 0 | 0 | 0 | 0 | 0 | 0 | 0 | 0 | 0 | 0 | 0 | |
| 1093 | F | 19-38 | | 2201 | 1475 | 0 | 0 | 1227 | 560 | 0 | 0 | 1 | 0 | 0 | 0 | 0 | 0 | 0 | 0 | 1 | 0 | 0 | 0 | 0 | |
| 1094 | H | 19-38 | | 4229 | 388 | 0 | 0 | 448 | 425 | 0 | 0 | 1 | 0 | 0 | 1 | 0 | 0 | 0 | 0 | 1 | 0 | 0 | 0 | 0 | |
| 1095 | F | 19-38 | | 8997 | 641 | 0 | 0 | 1177 | 977 | 0 | 0 | 1 | 1 | 1 | 1 | 1 | 0 | 0 | 0 | 0 | 0 | 1 | 1 | 1 | 0 |
| 1096 | F | 19-38 | | 2560 | 815 | 0 | 0 | 2289 | 1078 | 0 | 0 | 0 | 0 | 0 | 0 | 0 | 0 | 0 | 0 | 0 | 0 | 0 | 0 | 0 | |
| 1097 | F | 19-38 | | 8613 | 891 | 0 | 0 | 2966 | 664 | 0 | 0 | 0 | 0 | 0 | 0 | 0 | 0 | 0 | 0 | 0 | 0 | 0 | 0 | 0 | |
| 1098 | F | 19-38 | | 6826 | 571 | 0 | 0 | 8893 | 979 | 0 | 0 | 1 | 0 | 1 | 0 | 0 | 0 | 0 | 0 | 0 | 0 | 1 | 1 | 1 | 0 |
| 1099 | H | 39-82 | | 2241 | 2893 | 0 | 0 | 702 | 2214 | 0 | 0 | 0 | 0 | 0 | 0 | 0 | 0 | 0 | 0 | 0 | 0 | 0 | 0 | 0 | |
| 1100 | H | 19-38 | | 1412 | 734 | 0 | 0 | #N/A | #N/A | #N/A | #N/A | 0 | 0 | 0 | 0 | 0 | 0 | 0 | 0 | 0 | 0 | 0 | 0 | 0 | |
| 1101 | F | 19-38 | | 4881 | 2109 | 0 | 0 | 1303 | 893 | 0 | 0 | 0 | 0 | 0 | 0 | 0 | 0 | 0 | 0 | 0 | 0 | 0 | 0 | 0 | |
| 1102 | F | 39-82 | | 6302 | 2226 | 0 | 0 | #N/A | #N/A | #N/A | #N/A | 0 | 0 | 0 | 0 | 0 | 0 | 0 | 0 | 0 | 0 | 0 | 0 | 0 | |
| 1103 | F | 39-82 | | 3652 | 2176 | 0 | 0 | #N/A | #N/A | #N/A | #N/A | 0 | 0 | 0 | 0 | 0 | 0 | 0 | 0 | 0 | 0 | 0 | 0 | 0 | |
| 1104 | F | 39-82 | | 874 | 931 | 0 | 0 | 772 | 575 | 0 | 0 | 0 | 0 | 0 | 0 | 0 | 0 | 0 | 0 | 0 | 0 | 0 | 0 | 0 | |
| 1105 | F | 39-82 | | 596 | 1962 | 0 | 0 | 904 | 1464 | 0 | 0 | 1 | 0 | 0 | 0 | 0 | 0 | 0 | 0 | 0 | 0 | 1 | 0 | 0 | |
| 1106 | F | 19-38 | | 4786 | 872 | 0 | 0 | 5036 | 1239 | 0 | 0 | 1 | 0 | 1 | 1 | 0 | 1 | 0 | 0 | 1 | 0 | 1 | 0 | 0 | |
| 1107 | F | 39-82 | negative | 4173 | 360 | 0 | 0 | 253 | 195 | 0 | 0 | 1 | 1 | 1 | 1 | 1 | 1 | 1 | 0 | 1 | 1 | 1 | 0 | 1 | 0 |
| 1108 | F | 39-82 | | 1115 | 820 | 0 | 0 | #N/A | #N/A | #N/A | #N/A | 0 | 0 | 0 | 0 | 0 | 0 | 0 | 0 | 0 | 0 | 0 | 0 | 0 | |
| 1109 | M | 39-82 | IgM positive | 1182 | 968 | 0 | 0 | #N/A | #N/A | #N/A | #N/A | 1 | 1 | 1 | 1 | 1 | 1 | 0 | 0 | 0 | 1 | 0 | 0 | 1 | 0 |
| 1110 | F | 19-38 | | 1095 | 734 | 0 | 0 | 710 | 509 | 0 | 0 | 0 | 0 | 0 | 0 | 0 | 0 | 0 | 0 | 0 | 0 | 0 | 0 | 0 | |
| 1111 | F | 39-82 | | 2252 | 848 | 0 | 0 | 1549 | 286 | 0 | 0 | 1 | 1 | 1 | 0 | 1 | 1 | 1 | 0 | 0 | 0 | 1 | 1 | 0 | |
| 1112 | F | 39-82 | | 803 | 1176 | 0 | 0 | #N/A | #N/A | #N/A | #N/A | 1 | 0 | 0 | 0 | 0 | 0 | 0 | 0 | 0 | 0 | 0 | 1 | 0 | |
| 1113 | h | 19-38 | | 15802 | 1103 | 0 | 1 | 16603 | 1713 | 0 | 1 | 0 | 0 | 0 | 0 | 0 | 0 | 0 | 0 | 0 | 0 | 0 | 0 | 0 | |
| 1114 | H | 39-82 | | 1423 | 769 | 0 | 0 | #N/A | #N/A | #N/A | #N/A | 0 | 0 | 0 | 0 | 0 | 0 | 0 | 0 | 0 | 0 | 0 | 0 | 0 | |
| 1115 | F | 19-38 | | 1207 | 1154 | 0 | 0 | 1637 | 1071 | 0 | 0 | 0 | 0 | 0 | 0 | 0 | 0 | 0 | 0 | 0 | 0 | 0 | 0 | 0 | |
| 1116 | F | 19-38 | | 1188 | 1025 | 0 | 0 | #N/A | #N/A | #N/A | #N/A | 0 | 0 | 0 | 0 | 0 | 0 | 0 | 0 | 0 | 0 | 0 | 0 | 0 | |
| 1117 | F | 19-38 | | 2910 | 856 | 0 | 0 | 1002 | 817 | 0 | 0 | 0 | 0 | 0 | 0 | 0 | 0 | 0 | 0 | 0 | 0 | 0 | 0 | 0 | |
| 1118 | F | 19-38 | | 2067 | 1125 | 0 | 0 | 3207 | 789 | 0 | 0 | 0 | 0 | 0 | 0 | 0 | 0 | 0 | 0 | 0 | 0 | 0 | 0 | 0 | |
| 1119 | H | 39-82 | | 1124 | 1480 | 0 | 0 | #N/A | #N/A | #N/A | #N/A | 0 | 0 | 0 | 0 | 0 | 0 | 0 | 0 | 0 | 0 | 0 | 0 | 0 | |
| 1120 | F | 39-82 | | 1302 | 947 | 0 | 0 | 492 | 260 | 0 | 0 | 1 | 0 | 0 | 0 | 0 | 0 | 0 | 1 | 0 | 0 | 1 | 0 | 0 | |
| 1121 | F | 39-82 | | 93460 | 48277 | 7 | 1 | #N/A | #N/A | #N/A | #N/A | 1 | 1 | 1 | 1 | 1 | 0 | 1 | 1 | 1 | 1 | 1 | 0 | 0 | |
| 1122 | F | 39-82 | | 62893 | 55987 | 79 | 1 | 33821 | 67334 | 97 | 1 | 1 | 0 | 0 | 0 | 0 | 0 | 1 | 0 | 1 | 1 | 1 | 0 | 0 | |
| 1123 | F | 39-82 | | 976 | 545 | 0 | 0 | 837 | 892 | 0 | 0 | 1 | 0 | 0 | 1 | 0 | 0 | 1 | 0 | 0 | 0 | 0 | 0 | 0 | |
| 1124 | F | 39-82 | | 2392 | 625 | 0 | 0 | #N/A | #N/A | #N/A | #N/A | 1 | 0 | 1 | 0 | 0 | 0 | 0 | 0 | 0 | 0 | 0 | 0 | 0 | |
| 1125 | H | 39-82 | | 1403 | 432 | 0 | 0 | #N/A | #N/A | #N/A | #N/A | 0 | 0 | 0 | 0 | 0 | 0 | 0 | 0 | 0 | 0 | 0 | 0 | 0 | |
| 1126 | F | 39-82 | | 2859 | 548 | 0 | 0 | 2631 | 424 | 0 | 0 | 0 | 0 | 0 | 0 | 0 | 0 | 0 | 0 | 0 | 0 | 0 | 0 | 0 | |
| 1127 | F | 19-38 | | 71662 | 133877 | 83 | 1 | 30149 | 64832 | 52 | 1 | 1 | 0 | 1 | 1 | 1 | 1 | 1 | 0 | 0 | 1 | 1 | 0 | 1 | 1 |
| 1128 | H | 39-82 | | 618 | 386 | 0 | 0 | #N/A | #N/A | #N/A | #N/A | 0 | 0 | 0 | 0 | 0 | 0 | 0 | 0 | 0 | 0 | 0 | 0 | 0 | |
| 1129 | F | 19-38 | | 878 | 1312 | 0 | 0 | 803 | 915 | 0 | 0 | 1 | 1 | 1 | 1 | 1 | 1 | 1 | 0 | 0 | 0 | 1 | 1 | 1 | 0 |
| 1130 | F | 39-82 | | 764 | 1963 | 0 | 0 | 888 | 1246 | 0 | 0 | 1 | 0 | 0 | 0 | 0 | 0 | 0 | 0 | 0 | 0 | 0 | 1 | 0 | |
| 1132 | F | 19-38 | | 778 | 752 | 0 | 0 | 1042 | 731 | 0 | 0 | 1 | 0 | 1 | 1 | 0 | 0 | 0 | 0 | 1 | 0 | 1 | 0 | 0 | |
| 1133 | F | 39-82 | | 5516 | 1271 | 0 | 0 | 6674 | 1865 | 0 | 0 | 0 | 0 | 0 | 0 | 0 | 0 | 0 | 0 | 0 | 0 | 0 | 0 | 0 | |
| 1134 | F | 39-82 | | 1570 | 1026 | 0 | 0 | #N/A | #N/A | #N/A | #N/A | 0 | 0 | 0 | 0 | 0 | 0 | 0 | 0 | 0 | 0 | 0 | 0 | 0 | |
| 1135 | F | 19-38 | | 117143 | 129367 | 89 | 1 | 63169 | 173366 | 16 | 1 | 1 | 1 | 1 | 1 | 1 | 1 | 1 | 0 | 1 | 1 | 1 | 1 | 1 | 1 |
| 1136 | F | 39-82 | | 1043 | 706 | 0 | 0 | #N/A | #N/A | #N/A | #N/A | 1 | 1 | 1 | 0 | 0 | 0 | 0 | 0 | 0 | 0 | 1 | 0 | 0 | |
| 1137 | F | 19-38 | | 685 | 489 | 0 | 0 | 1306 | 836 | 0 | 0 | 0 | 0 | 0 | 0 | 0 | 0 | 0 | 0 | 0 | 0 | 0 | 0 | 0 | |
| 1138 | M | 19-38 | | 21830 | 30085 | 92 | 1 | 11668 | 18074 | 24 | 1 | 1 | 1 | 1 | 1 | 0 | 1 | 0 | 0 | 0 | 1 | 0 | 0 | 0 | |
| 1139 | M | 39-82 | | 5887 | 978 | 0 | 0 | 2645 | 743 | 0 | 0 | 1 | 0 | 0 | 0 | 0 | 0 | 1 | 0 | 0 | 0 | 1 | 0 | 0 | |
| 1140 | F | 19-38 | | 755 | 447 | 0 | 0 | 1587 | 674 | 0 | 0 | 0 | 0 | 0 | 0 | 0 | 0 | 0 | 0 | 0 | 0 | 0 | 0 | 0 | |
| 1141 | F | 39-82 | | 1232 | 1478 | 0 | 0 | 1533 | 997 | 0 | 0 | 0 | 0 | 0 | 0 | 0 | 0 | 0 | 0 | 0 | 0 | 0 | 0 | 0 | |
| 1142 | M | 19-38 | | 1241 | 846 | 0 | 0 | #N/A | #N/A | #N/A | #N/A | 1 | 0 | 1 | 0 | 0 | 0 | 0 | 0 | 0 | 0 | 0 | 0 | 0 | |
| 1143 | M | 19-38 | | 713 | 1502 | 0 | 0 | 711 | 1079 | 0 | 0 | 0 | 0 | 0 | 0 | 0 | 0 | 0 | 0 | 0 | 0 | 0 | 0 | 0 | |
| 1144 | M | 39-82 | | 431 | 869 | 0 | 0 | 510 | 782 | 0 | 0 | 0 | 0 | 0 | 0 | 0 | 0 | 0 | 0 | 0 | 0 | 0 | 0 | 0 | |
| 1145 | M | 39-82 | | 819 | 1080 | 0 | 0 | #N/A | #N/A | #N/A | #N/A | 1 | 1 | 1 | 0 | 0 | 0 | 1 | 1 | 0 | 0 | 1 | 0 | 0 | |
| 1146 | M | 39-82 | | 1689 | 1389 | 0 | 0 | 3073 | 1444 | 0 | 0 | 1 | 0 | 0 | 0 | 0 | 1 | 0 | 1 | 1 | 0 | 0 | 0 | 0 | |
| 1147 | F | 39-82 | | 3062 | 692 | 0 | 0 | 913 | 791 | 0 | 0 | 0 | 0 | 0 | 0 | 0 | 0 | 0 | 0 | 0 | 0 | 0 | 0 | 0 | |
| 1148 | F | 19-38 | | 984 | 528 | 0 | 0 | 2128 | 1014 | 0 | 0 | 1 | 0 | 0 | 1 | 0 | 0 | 0 | 0 | 1 | 0 | 1 | 0 | 0 | |
| 1149 | M | 39-82 | | 1795 | 3465 | 0 | 0 | 2924 | 2868 | 0 | 0 | 1 | 1 | 1 | 1 | 1 | 1 | 0 | 1 | 1 | 0 | 1 | 0 | 0 | |
| 1150 | M | 19-38 | | 841 | 746 | 0 | 0 | 1601 | 1029 | 0 | 0 | 0 | 0 | 0 | 0 | 0 | 0 | 0 | 0 | 0 | 0 | 0 | 0 | 0 | |
| 1151 | F | 19-38 | | 2275 | 728 | 0 | 0 | 2257 | 926 | 0 | 0 | 1 | 0 | 1 | 0 | 0 | 0 | 0 | 0 | 0 | 0 | 0 | 0 | 0 | |
| 1152 | F | 19-38 | | 439 | 828 | 0 | 0 | 1847 | 1876 | 0 | 0 | 0 | 0 | 0 | 0 | 0 | 0 | 0 | 0 | 0 | 0 | 0 | 0 | 0 | |
| 1153 | M | 39-82 | | 1619 | 789 | 0 | 0 | 1682 | 1451 | 0 | 0 | 1 | 0 | 1 | 0 | 0 | 0 | 0 | 0 | 0 | 0 | 0 | 0 | 0 | |
| 1154 | F | 39-82 | | 2574 | 590 | 0 | 0 | 5544 | 1057 | 0 | 0 | 0 | 0 | 0 | 0 | 0 | 0 | 0 | 0 | 0 | 0 | 0 | 0 | 0 | |
| 1155 | F | 19-38 | | 1463 | 617 | 0 | 0 | 2565 | 879 | 0 | 0 | 1 | 1 | 1 | 0 | 1 | 1 | 0 | 0 | 1 | 0 | 1 | 0 | 0 | |
| 1156 | F | 39-82 | | 512 | 766 | 0 | 0 | 299 | 252 | 0 | 0 | 0 | 0 | 0 | 0 | 0 | 0 | 0 | 0 | 0 | 0 | 0 | 0 | 0 | |
| 1157 | F | 19-38 | | 2071 | 2446 | 0 | 0 | 2589 | 2472 | 0 | 0 | 0 | 0 | 0 | 0 | 0 | 0 | 0 | 0 | 0 | 0 | 0 | 0 | 0 | |
| 1158 | F | 19-38 | | 1963 | 1814 | 0 | 0 | 2738 | 1344 | 0 | 0 | 0 | 0 | 0 | 0 | 0 | 0 | 0 | 0 | 0 | 0 | 0 | 0 | 0 | |
| 1159 | F | 39-82 | | 6264 | 949 | 0 | 0 | 4687 | 571 | 0 | 0 | 0 | 0 | 0 | 0 | 0 | 0 | 0 | 0 | 0 | 0 | 0 | 0 | 0 | |
| 1160 | F | 39-82 | | 1462 | 2329 | 0 | 0 | 3576 | 657 | 0 | 0 | 0 | 0 | 0 | 0 | 0 | 0 | 0 | 0 | 0 | 0 | 0 | 0 | 0 | |
| 1161 | F | 19-38 | IgM positive | 3823 | 2203 | 0 | 0 | 4381 | 872 | 0 | 0 | 1 | 1 | 0 | 1 | 0 | 1 | 0 | 0 | 1 | 1 | 1 | 0 | 0 | |
| 1162 | F | 39-82 | | 8293 | 1037 | 0 | 0 | 8613 | 1088 | 0 | 0 | 0 | 0 | 0 | 0 | 0 | 0 | 0 | 0 | 0 | 0 | 0 | 0 | 0 | |
| 1163 | M | 39-82 | | 978 | 594 | 0 | 0 | 1046 | 1471 | 0 | 0 | 0 | 0 | 0 | 0 | 0 | 0 | 0 | 0 | 0 | 0 | 0 | 0 | 0 | |

| | | | | | | | | | | | | | | | | | | | | | | | | | |
|---|---|---|---|---|---|---|---|---|---|---|---|---|---|---|---|---|---|---|---|---|---|---|---|---|---|
| 1164 | F | 19-38 | | 505 | 616 | 0 | 0 | 2079 | 882 | 0 | 0 | 1 | 0 | 0 | 0 | 0 | 0 | 0 | 0 | 1 | 0 | 0 | 1 | 0 | |
| 1165 | F | 39-82 | | 1021 | 1246 | 0 | 0 | 1518 | 900 | 0 | 0 | 0 | 0 | 0 | 0 | 0 | 0 | 0 | 0 | 0 | 0 | 0 | 0 | 0 | |
| 1166 | F | 19-38 | | 411 | 913 | 0 | 0 | 1213 | 1745 | 0 | 0 | 0 | 0 | 0 | 0 | 0 | 0 | 0 | 0 | 0 | 0 | 0 | 0 | 0 | |
| 1167 | F | 19-38 | | 892 | 430 | 0 | 0 | #N/A | #N/A | #N/A | #N/A | 0 | 0 | 0 | 0 | 0 | 0 | 0 | 0 | 0 | 0 | 0 | 0 | 0 | |
| 1168 | F | 39-82 | | 504 | 723 | 0 | 0 | 387 | 1079 | 0 | 0 | 0 | 0 | 0 | 0 | 0 | 0 | 0 | 0 | 0 | 0 | 0 | 0 | 0 | |
| 1169 | F | 19-38 | | 1204 | 1218 | 0 | 0 | 2000 | 1475 | 0 | 0 | 1 | 0 | 1 | 0 | 0 | 0 | 0 | 0 | 1 | 0 | 0 | 0 | 0 | |
| 1170 | F | 39-82 | | 5557 | 1075 | 0 | 0 | 2018 | 977 | 0 | 0 | 0 | 0 | 0 | 0 | 0 | 0 | 0 | 0 | 0 | 0 | 0 | 0 | 0 | |
| 1171 | F | 39-82 | | 879 | 1695 | 0 | 0 | 908 | 1605 | 0 | 0 | 1 | 1 | 1 | 1 | 1 | 0 | 1 | 0 | 1 | 0 | 0 | 0 | 0 | |
| 1172 | M | 39-82 | | 679 | 471 | 0 | 0 | 1581 | 500 | 0 | 0 | 0 | 0 | 0 | 0 | 0 | 0 | 0 | 0 | 0 | 0 | 0 | 0 | 0 | |
| 1173 | M | 19-38 | | 1144 | 573 | 0 | 0 | 2046 | 949 | 0 | 0 | 0 | 0 | 0 | 0 | 0 | 0 | 0 | 0 | 0 | 0 | 0 | 0 | 0 | |
| 1174 | F | 19-38 | | 3579 | 1669 | 0 | 0 | #N/A | #N/A | #N/A | #N/A | 1 | 0 | 1 | 0 | 0 | 0 | 0 | 0 | 0 | 0 | 0 | 1 | 0 | |
| 1175 | M | 39-82 | | 6583 | 925 | 0 | 0 | #N/A | #N/A | #N/A | #N/A | 1 | 0 | 0 | 0 | 0 | 0 | 0 | 0 | 0 | 0 | 1 | 0 | 0 | |
| 1176 | F | 19-38 | | 3499 | 2234 | 0 | 0 | 3007 | 1045 | 0 | 0 | 1 | 0 | 1 | 0 | 1 | 1 | 1 | 0 | 1 | 0 | 1 | 1 | 1 | 0 |
| 1177 | F | 39-82 | | 11742 | 31845 | 29 | 1 | 724 | 2328 | 0 | 0 | 1 | 0 | 0 | 0 | 0 | 0 | 0 | 0 | 0 | 1 | 1 | 0 | 0 | |
| 1178 | M | 39-82 | | 24572 | 99093 | 76 | 1 | #N/A | #N/A | #N/A | #N/A | 1 | 1 | 1 | 1 | 1 | 1 | 1 | 0 | 1 | 1 | 1 | 1 | 0 | |
| 1179 | M | 39-82 | | 20820 | 33471 | 14 | 1 | 1207 | 1823 | 0 | 0 | 1 | 1 | 1 | 1 | 1 | 1 | 1 | 0 | 1 | 1 | 1 | 1 | 0 | |
| 1180 | M | 39-82 | | 3851 | 570 | 0 | 0 | #N/A | #N/A | #N/A | #N/A | 1 | 0 | 1 | 0 | 0 | 0 | 1 | 0 | 0 | 0 | 1 | 0 | 0 | |
| 1181 | F | 19-38 | | 865 | 750 | 0 | 0 | 875 | 562 | 0 | 0 | 1 | 0 | 0 | 0 | 0 | 0 | 0 | 0 | 0 | 0 | 0 | 1 | 0 | |
| 1182 | F | 19-38 | | 3822 | 1051 | 0 | 0 | 5136 | 1164 | 0 | 0 | 1 | 0 | 1 | 0 | 0 | 0 | 0 | 0 | 0 | 0 | 0 | 0 | 0 | |
| 1183 | F | 39-82 | | 1552 | 2276 | 0 | 0 | 1316 | 3283 | 0 | 0 | 1 | 0 | 1 | 0 | 0 | 0 | 0 | 0 | 0 | 0 | 0 | 1 | 0 | |
| 1184 | F | 19-38 | | 1565 | 427 | 0 | 0 | #N/A | #N/A | #N/A | #N/A | 0 | 0 | 0 | 0 | 0 | 0 | 0 | 0 | 0 | 0 | 0 | 0 | 0 | |
| 1185 | F | 39-82 | | 1190 | 798 | 0 | 0 | #N/A | #N/A | #N/A | #N/A | 1 | 0 | 0 | 0 | 0 | 0 | 0 | 0 | 0 | 0 | 1 | 0 | 0 | |
| 1186 | F | 19-38 | | 1349 | 1671 | 0 | 0 | 3439 | 4486 | 0 | 0 | 1 | 0 | 0 | 1 | 0 | 0 | 0 | 0 | 1 | 0 | 0 | 0 | 0 | |
| 1187 | F | 19-38 | | 1294 | 1429 | 0 | 0 | 23857 | 587 | 0 | 1 | 0 | 0 | 0 | 0 | 0 | 0 | 0 | 0 | 0 | 0 | 0 | 0 | 0 | |
| 1188 | F | 19-38 | | 1964 | 1851 | 0 | 0 | #N/A | #N/A | #N/A | #N/A | 1 | 1 | 1 | 0 | 0 | 0 | 0 | 0 | 1 | 0 | 0 | 1 | 0 | |
| 1189 | F | 39-82 | | 619 | 1170 | 0 | 0 | 689 | 1064 | 0 | 0 | 0 | 0 | 0 | 0 | 0 | 0 | 0 | 0 | 0 | 0 | 0 | 0 | 0 | |
| 1190 | F | 19-38 | | 809 | 845 | 0 | 0 | #N/A | #N/A | #N/A | #N/A | 1 | 0 | 0 | 1 | 0 | 0 | 0 | 0 | 0 | 0 | 1 | 1 | 0 | |
| 1191 | M | 19-38 | | 72513 | 49416 | 80 | 1 | 52368 | 38921 | 0 | 1 | 1 | 0 | 0 | 0 | 0 | 0 | 0 | 0 | 0 | 1 | 0 | 0 | 0 | |
| 1192 | F | 39-82 | IgM positive | 4410 | 34429 | 0 | 1 | 3336 | 52615 | 0 | 1 | 1 | 0 | 0 | 1 | 0 | 0 | 0 | 0 | 0 | 1 | 0 | 1 | 0 | |
| 1194 | M | 19-38 | | 3706 | 1961 | 0 | 0 | 1681 | 2346 | 0 | 0 | 0 | 0 | 0 | 0 | 0 | 0 | 0 | 0 | 0 | 0 | 0 | 0 | 0 | |
| 1195 | F | 39-82 | | 3368 | 4266 | 0 | 0 | 3546 | 3534 | 0 | 0 | 0 | 0 | 0 | 0 | 0 | 0 | 0 | 0 | 0 | 0 | 0 | 0 | 0 | |
| 1196 | M | 19-38 | | 1635 | 3319 | 0 | 0 | #N/A | #N/A | #N/A | #N/A | 1 | 0 | 0 | 1 | 1 | 1 | 0 | 0 | 1 | 0 | 1 | 0 | 0 | |
| 1197 | M | 39-82 | | 7897 | 479 | 0 | 0 | 7318 | 817 | 0 | 0 | 0 | 0 | 0 | 0 | 0 | 0 | 0 | 0 | 0 | 0 | 0 | 0 | 0 | |
| 1198 | F | 19-38 | | 1290 | 487 | 0 | 0 | 2029 | 975 | 0 | 0 | 1 | 0 | 0 | 0 | 0 | 0 | 0 | 0 | 1 | 0 | 0 | 0 | 0 | |
| 1199 | F | 39-82 | | 1985 | 676 | 0 | 0 | 2042 | 866 | 0 | 0 | 1 | 0 | 1 | 1 | 0 | 0 | 0 | 0 | 1 | 0 | 0 | 0 | 0 | |
| 1200 | F | 19-38 | | 1102 | 605 | 0 | 0 | 898 | 468 | 0 | 0 | 0 | 0 | 0 | 0 | 0 | 0 | 0 | 0 | 0 | 0 | 0 | 0 | 0 | |
| 1201 | F | 19-38 | | 1704 | 950 | 0 | 0 | 907 | 746 | 0 | 0 | 1 | 0 | 1 | 1 | 0 | 1 | 0 | 0 | 1 | 0 | 0 | 0 | 0 | |
| 1202 | F | 19-38 | | 1049 | 991 | 0 | 0 | 888 | 1745 | 0 | 0 | 0 | 0 | 0 | 0 | 0 | 0 | 0 | 0 | 0 | 0 | 0 | 0 | 0 | |
| 1203 | F | 39-82 | | 723 | 713 | 0 | 0 | 727 | 897 | 0 | 0 | 1 | 0 | 0 | 0 | 0 | 0 | 0 | 0 | 0 | 0 | 1 | 0 | 0 | |
| 1204 | F | 19-38 | | 24059 | 35639 | 87 | 1 | 20440 | 26235 | 9 | 1 | 1 | 0 | 1 | 0 | 1 | 1 | 1 | 0 | 1 | 1 | 1 | 1 | 0 | |
| 1205 | F | 19-38 | | 1725 | 1169 | 0 | 0 | 1439 | 500 | 0 | 0 | 0 | 0 | 0 | 0 | 0 | 0 | 0 | 0 | 0 | 0 | 0 | 0 | 0 | |
| 1206 | F | 19-38 | | 906 | 3886 | 0 | 0 | 219 | 244 | 0 | 0 | 1 | 1 | 1 | 1 | 1 | 1 | 0 | 0 | 0 | 0 | 0 | 1 | 0 | |
| 1207 | F | 19-38 | | 1017 | 1100 | 0 | 0 | #N/A | #N/A | #N/A | #N/A | 0 | 0 | 0 | 0 | 0 | 0 | 0 | 0 | 0 | 0 | 0 | 0 | 0 | |
| 1208 | F | 19-38 | | 791 | 691 | 0 | 0 | 425 | 439 | 0 | 0 | 0 | 0 | 0 | 0 | 0 | 0 | 0 | 0 | 0 | 0 | 0 | 0 | 0 | |
| 1209 | F | 19-38 | | 28142 | 30725 | 58 | 1 | 17772 | 25040 | 41 | 1 | 1 | 1 | 0 | 1 | 0 | 1 | 0 | 0 | 0 | 1 | 0 | 0 | 1 | 1 |
| 1210 | F | 39-82 | | 1608 | 867 | 0 | 0 | 1213 | 1156 | 0 | 0 | 1 | 0 | 1 | 0 | 0 | 0 | 0 | 0 | 0 | 0 | 0 | 0 | 0 | |
| 1211 | F | 39-82 | | 1096 | 549 | 0 | 0 | 1977 | 1140 | 0 | 0 | 1 | 1 | 1 | 1 | 1 | 0 | 1 | 0 | 0 | 0 | 1 | 0 | 0 | |
| 1212 | F | 39-82 | | 1908 | 1178 | 0 | 0 | 1155 | 1896 | 0 | 0 | 0 | 0 | 0 | 0 | 0 | 0 | 0 | 0 | 0 | 0 | 0 | 0 | 0 | |
| 1213 | F | 19-38 | IgM positive | 1010 | 710 | 0 | 0 | 856 | 3913 | 0 | 0 | 1 | 1 | 1 | 1 | 1 | 1 | 0 | 0 | 1 | 1 | 0 | 0 | 0 | |
| 1214 | F | 39-82 | | 921 | 1271 | 0 | 0 | 1502 | 1358 | 0 | 0 | 1 | 0 | 0 | 0 | 0 | 0 | 0 | 0 | 1 | 0 | 1 | 0 | 0 | |
| 1215 | F | 39-82 | | 898 | 1228 | 0 | 0 | 3356 | 1742 | 0 | 0 | 0 | 0 | 0 | 0 | 0 | 0 | 0 | 0 | 0 | 0 | 0 | 0 | 0 | |
| 1216 | F | 19-38 | | 2132 | 2685 | 0 | 0 | 2807 | 1239 | 0 | 0 | 0 | 0 | 0 | 0 | 0 | 0 | 0 | 0 | 0 | 0 | 0 | 0 | 0 | |
| 1217 | F | 39-82 | | 594 | 1216 | 0 | 0 | 690 | 1018 | 0 | 0 | 0 | 0 | 0 | 0 | 0 | 0 | 0 | 0 | 0 | 0 | 0 | 0 | 0 | |
| 1218 | F | 19-38 | | 870 | 640 | 0 | 0 | 2442 | 1285 | 0 | 0 | 1 | 0 | 1 | 0 | 0 | 0 | 0 | 0 | 1 | 0 | 1 | 0 | 0 | |
| 1219 | F | 39-82 | | 951 | 387 | 0 | 0 | #N/A | #N/A | #N/A | #N/A | 1 | 1 | 1 | 0 | 0 | 0 | 0 | 0 | 0 | 0 | 0 | 0 | 0 | |
| 1220 | F | 39-82 | | 935 | 967 | 0 | 0 | 1146 | 346 | 0 | 0 | 1 | 0 | 1 | 0 | 0 | 0 | 0 | 0 | 0 | 0 | 1 | 1 | 0 | |
| 1221 | F | 19-38 | | 1115 | 710 | 0 | 0 | 1101 | 597 | 0 | 0 | 0 | 0 | 0 | 0 | 0 | 0 | 0 | 0 | 0 | 0 | 0 | 0 | 0 | |
| 1222 | H | 39-82 | | 1345 | 422 | 0 | 0 | 1123 | 564 | 0 | 0 | 0 | 0 | 0 | 0 | 0 | 0 | 0 | 0 | 0 | 0 | 0 | 0 | 0 | |
| 1223 | F | 19-38 | | 2171 | 1624 | 0 | 0 | #N/A | #N/A | #N/A | #N/A | 1 | 0 | 0 | 0 | 0 | 0 | 0 | 0 | 0 | 0 | 0 | 1 | 0 | |
| 1224 | F | 39-82 | | 962 | 815 | 0 | 0 | 836 | 816 | 0 | 0 | 1 | 0 | 1 | 0 | 1 | 0 | 0 | 0 | 0 | 0 | 0 | 0 | 0 | |
| 1225 | F | 39-82 | | 5071 | 864 | 0 | 0 | 5589 | 1140 | 0 | 0 | 1 | 0 | 0 | 0 | 0 | 0 | 0 | 0 | 1 | 0 | 0 | 0 | 0 | |
| 1226 | H | 39-82 | | 715 | 488 | 0 | 0 | #N/A | #N/A | #N/A | #N/A | 1 | 0 | 1 | 0 | 0 | 0 | 1 | 1 | 0 | 0 | 0 | 0 | 0 | |
| 1227 | F | 39-82 | | 1190 | 520 | 0 | 0 | 1470 | 534 | 0 | 0 | 1 | 0 | 1 | 0 | 0 | 0 | 0 | 0 | 0 | 0 | 1 | 0 | 0 | |
| 1228 | F | 39-82 | | 930 | 608 | 0 | 0 | 717 | 1139 | 0 | 0 | 0 | 0 | 0 | 0 | 0 | 0 | 0 | 0 | 0 | 0 | 0 | 0 | 0 | |
| 1229 | H | 39-82 | | 2071 | 1207 | 0 | 0 | 2435 | 1063 | 0 | 0 | 1 | 0 | 1 | 1 | 1 | 1 | 0 | 0 | 0 | 0 | 1 | 1 | 0 | |
| 1230 | F | 39-82 | | 977 | 1281 | 0 | 0 | #N/A | #N/A | #N/A | #N/A | 1 | 0 | 1 | 0 | 0 | 0 | 0 | 0 | 0 | 0 | 0 | 0 | 0 | |
| 1231 | H | 19-38 | | 1154 | 3423 | 0 | 0 | 1217 | 866 | 0 | 0 | 0 | 0 | 0 | 0 | 0 | 0 | 0 | 0 | 0 | 0 | 0 | 0 | 0 | |
| 1232 | H | 39-82 | | 862 | 574 | 0 | 0 | 1297 | 537 | 0 | 0 | 0 | 0 | 0 | 0 | 0 | 0 | 0 | 0 | 0 | 0 | 0 | 0 | 0 | |
| 1233 | H | 19-38 | | 1688 | 2317 | 0 | 0 | 1982 | 1803 | 0 | 0 | 0 | 0 | 0 | 0 | 0 | 0 | 0 | 0 | 0 | 0 | 0 | 0 | 0 | |
| 1234 | F | 19-38 | | 2563 | 1021 | 0 | 0 | 2533 | 1321 | 0 | 0 | 1 | 1 | 0 | 0 | 0 | 0 | 0 | 1 | 1 | 0 | 0 | 1 | 0 | |
| 1236 | F | 19-38 | | 1494 | 1572 | 0 | 0 | #N/A | #N/A | #N/A | #N/A | 1 | 0 | 1 | 1 | 0 | 0 | 0 | 0 | 0 | 0 | 1 | 0 | 0 | |
| 1237 | F | 39-82 | | 579 | 660 | 0 | 0 | 914 | 386 | 0 | 0 | 0 | 0 | 0 | 0 | 0 | 0 | 0 | 0 | 0 | 0 | 0 | 0 | 0 | |
| 1238 | F | 39-82 | | 737 | 452 | 0 | 0 | 1810 | 1223 | 0 | 0 | 1 | 0 | 1 | 0 | 1 | 0 | 1 | 0 | 0 | 0 | 1 | 0 | 0 | |
| 1239 | F | 39-82 | | 2140 | 1770 | 0 | 0 | 1673 | 1791 | 0 | 0 | 1 | 0 | 1 | 0 | 0 | 0 | 1 | 0 | 0 | 0 | 0 | 0 | 0 | |
| 1240 | F | 39-82 | | 571 | 882 | 0 | 0 | #N/A | #N/A | #N/A | #N/A | 0 | 0 | 0 | 0 | 0 | 0 | 0 | 0 | 0 | 0 | 0 | 0 | 0 | |
| 1241 | H | 19-38 | | 967 | 1466 | 0 | 0 | 913 | 1052 | 0 | 0 | 0 | 0 | 0 | 0 | 0 | 0 | 0 | 0 | 0 | 0 | 0 | 0 | 0 | |
| 1242 | F | 19-38 | | 1267 | 491 | 0 | 0 | 1382 | 900 | 0 | 0 | 0 | 0 | 0 | 0 | 0 | 0 | 0 | 0 | 0 | 0 | 0 | 0 | 0 | |

| | | | | | | | | | | | | | | | | | | | | | | | | | |
|---|---|---|---|---|---|---|---|---|---|---|---|---|---|---|---|---|---|---|---|---|---|---|---|---|---|
| 1243 | F | 19-38 | | 2989 | 979 | 0 | 0 | 1516 | 961 | 0 | 0 | 0 | 0 | 0 | 0 | 0 | 0 | 0 | 0 | 0 | 0 | 0 | 0 | 0 | |
| 1244 | H | 19-38 | | 6883 | 1388 | 0 | 0 | 4840 | 1618 | 0 | 0 | 0 | 0 | 0 | 0 | 0 | 0 | 0 | 0 | 0 | 0 | 0 | 0 | 0 | |
| 1245 | F | 39-82 | | 40770 | 96495 | 89 | 1 | #N/A | #N/A | #N/A | #N/A | 1 | 0 | 1 | 0 | 0 | 0 | 0 | 0 | 0 | 1 | 0 | 0 | 0 | |
| 1246 | F | 19-38 | | 1229 | 1513 | 0 | 0 | 282 | 676 | 0 | 0 | 0 | 0 | 0 | 0 | 0 | 0 | 0 | 0 | 0 | 0 | 0 | 0 | 0 | |
| 1247 | F | 39-82 | | 3609 | 2604 | 0 | 0 | 1748 | 2382 | 0 | 0 | 1 | 1 | 0 | 1 | 1 | 1 | 0 | 0 | 1 | 0 | 1 | 1 | 0 | |
| 1248 | F | 39-82 | | 2020 | 636 | 0 | 0 | 1648 | 512 | 0 | 0 | 0 | 0 | 0 | 0 | 0 | 0 | 0 | 0 | 0 | 0 | 0 | 0 | 0 | |
| 1249 | F | 39-82 | | 788 | 612 | 0 | 0 | 1087 | 6602 | 0 | 0 | 0 | 0 | 0 | 0 | 0 | 0 | 0 | 0 | 0 | 0 | 0 | 0 | 0 | |
| 1250 | H | 39-82 | | 1955 | 1318 | 0 | 0 | #N/A | #N/A | #N/A | #N/A | 0 | 0 | 0 | 0 | 0 | 0 | 0 | 0 | 0 | 0 | 0 | 0 | 0 | |
| 1251 | F | 19-38 | | 1726 | 2748 | 0 | 0 | 1084 | 1499 | 0 | 0 | 0 | 0 | 0 | 0 | 0 | 0 | 0 | 0 | 0 | 0 | 0 | 0 | 0 | |
| 1252 | F | 39-82 | | 1891 | 719 | 0 | 0 | 7344 | 1111 | 0 | 0 | 1 | 1 | 1 | 0 | 0 | 0 | 1 | 0 | 1 | 0 | 0 | 0 | 1 | 0 |
| 1253 | F | 39-82 | | 2410 | 703 | 0 | 0 | 1928 | 433 | 0 | 0 | 0 | 0 | 0 | 0 | 0 | 0 | 0 | 0 | 0 | 0 | 0 | 0 | 0 | |
| 1254 | F | 39-82 | | 2802 | 1676 | 0 | 0 | 4532 | 2535 | 0 | 0 | 0 | 0 | 0 | 0 | 0 | 0 | 0 | 0 | 0 | 0 | 0 | 0 | 0 | |
| 1256 | F | 19-38 | | 2210 | 3941 | 0 | 0 | #N/A | #N/A | #N/A | #N/A | 1 | 1 | 1 | 1 | 1 | 1 | 1 | 1 | 1 | 0 | 1 | 1 | 0 | |
| 1257 | F | 39-82 | | 722 | 364 | 0 | 0 | #N/A | #N/A | #N/A | #N/A | 1 | 0 | 0 | 0 | 0 | 0 | 0 | 0 | 1 | 0 | 0 | 0 | 0 | |
| 1258 | F | 19-38 | | 3790 | 536 | 0 | 0 | 1412 | 432 | 0 | 0 | 0 | 0 | 0 | 0 | 0 | 0 | 0 | 0 | 0 | 0 | 0 | 0 | 0 | |
| 1259 | F | 19-38 | | 48158 | 50204 | 63 | 1 | 36746 | 55614 | 5 | 1 | 1 | 0 | 1 | 1 | 0 | 0 | 1 | 0 | 1 | 1 | 0 | 0 | 0 | |
| 1260 | H | 39-82 | | 1242 | 1107 | 0 | 0 | #N/A | #N/A | #N/A | #N/A | 1 | 0 | 0 | 1 | 0 | 0 | 0 | 0 | 0 | 0 | 1 | 0 | 0 | |
| 1261 | H | 39-82 | | 2698 | 774 | 0 | 0 | #N/A | #N/A | #N/A | #N/A | 1 | 0 | 1 | 0 | 0 | 0 | 0 | 0 | 0 | 0 | 0 | 0 | 0 | |
| 1262 | F | 19-38 | | 831 | 879 | 0 | 0 | #N/A | #N/A | #N/A | #N/A | 1 | 1 | 1 | 0 | 1 | 1 | 1 | 0 | 0 | 0 | 1 | 0 | 0 | |
| 1263 | H | 39-82 | | 5018 | 4830 | 0 | 0 | 4802 | 2656 | 0 | 0 | 1 | 0 | 1 | 0 | 0 | 0 | 1 | 0 | 1 | 0 | 1 | 0 | 1 | 0 |
| 1264 | F | 39-82 | | 4802 | 831 | 0 | 0 | #N/A | #N/A | #N/A | #N/A | 0 | 0 | 0 | 0 | 0 | 0 | 0 | 0 | 0 | 0 | 0 | 0 | 0 | |
| 1265 | H | 19-38 | | 21195 | 837 | 0 | 1 | 23145 | 675 | 0 | 1 | 0 | 0 | 0 | 0 | 0 | 0 | 0 | 0 | 0 | 0 | 0 | 0 | 0 | |
| 1266 | F | 19-38 | | 3888 | 3097 | 0 | 0 | #N/A | #N/A | #N/A | #N/A | 1 | 0 | 1 | 0 | 0 | 0 | 0 | 0 | 0 | 0 | 1 | 0 | 1 | 0 |
| 1267 | F | 39-82 | | 1925 | 1723 | 0 | 0 | 1516 | 1529 | 0 | 0 | 1 | 0 | 0 | 0 | 0 | 0 | 0 | 1 | 0 | 0 | 1 | 1 | 0 | |
| 1268 | F | 39-82 | | 1807 | 940 | 0 | 0 | 1355 | 985 | 0 | 0 | 1 | 1 | 1 | 0 | 0 | 0 | 1 | 1 | 1 | 0 | 1 | 0 | 0 | |
| 1269 | F | 39-82 | | 2174 | 1473 | 0 | 0 | #N/A | #N/A | #N/A | #N/A | 0 | 0 | 0 | 0 | 0 | 0 | 0 | 0 | 0 | 0 | 0 | 0 | 0 | |
| 1270 | F | 39-82 | | 7687 | 16100 | 24 | 1 | 3547 | 20631 | 0 | 1 | 0 | 0 | 0 | 0 | 0 | 0 | 0 | 0 | 0 | 0 | 0 | 0 | 0 | |
| 1271 | F | 39-82 | | 5795 | 770 | 0 | 0 | 353 | 211 | 0 | 0 | 1 | 0 | 1 | 0 | 0 | 0 | 0 | 0 | 0 | 0 | 0 | 0 | 0 | |
| 1272 | F | 39-82 | | 32776 | 19149 | 22 | 1 | 28180 | 23517 | 0 | 1 | 1 | 0 | 0 | 0 | 0 | 0 | 1 | 1 | 0 | 1 | 1 | 0 | 0 | |
| 1273 | F | 19-38 | | 1073 | 628 | 0 | 0 | 1624 | 828 | 0 | 0 | 1 | 1 | 1 | 1 | 0 | 0 | 0 | 0 | 0 | 0 | 0 | 0 | 0 | |
| 1274 | F | 19-38 | | 102969 | 60383 | 39 | 1 | 90843 | 42251 | 3 | 1 | 1 | 1 | 1 | 1 | 1 | 1 | 1 | 0 | 1 | 1 | 1 | 1 | 0 | |
| 1275 | F | 19-38 | | 1069 | 752 | 0 | 0 | 904 | 380 | 0 | 0 | 0 | 0 | 0 | 0 | 0 | 0 | 0 | 0 | 0 | 0 | 0 | 0 | 0 | |
| 1276 | F | 39-82 | | 1487 | 944 | 0 | 0 | 1414 | 328 | 0 | 0 | 1 | 0 | 1 | 0 | 0 | 0 | 0 | 0 | 1 | 0 | 1 | 0 | 0 | |
| 1277 | F | 39-82 | | 2920 | 598 | 0 | 0 | 2105 | 487 | 0 | 0 | 0 | 0 | 0 | 0 | 0 | 0 | 0 | 0 | 0 | 0 | 0 | 0 | 0 | |
| 1278 | F | 19-38 | | 662 | 1297 | 0 | 0 | 665 | 2208 | 0 | 0 | 1 | 0 | 0 | 0 | 0 | 0 | 1 | 0 | 0 | 0 | 1 | 0 | 0 | |
| 1279 | F | 39-82 | | 1445 | 1219 | 0 | 0 | 1171 | 1147 | 0 | 0 | 1 | 1 | 1 | 1 | 0 | 1 | 1 | 1 | 1 | 0 | 1 | 0 | 1 | |
| 1280 | H | 39-82 | | 3428 | 1602 | 0 | 0 | 2586 | 1150 | 0 | 0 | 1 | 0 | 0 | 0 | 0 | 0 | 0 | 0 | 0 | 0 | 0 | 1 | 0 | |
| 1281 | F | 19-38 | | 390 | 292 | 0 | 0 | #N/A | #N/A | #N/A | #N/A | 1 | 1 | 1 | 1 | 0 | 0 | 0 | 0 | 0 | 0 | 1 | 1 | 0 | |
| 1282 | H | 19-38 | | 1776 | 718 | 0 | 0 | #N/A | #N/A | #N/A | #N/A | 1 | 0 | 1 | 1 | 0 | 0 | 0 | 0 | 1 | 0 | 0 | 0 | 0 | |
| 1283 | F | 19-38 | | 1069 | 767 | 0 | 0 | #N/A | #N/A | #N/A | #N/A | 1 | 0 | 1 | 0 | 1 | 1 | 1 | 0 | 0 | 0 | 1 | 0 | 1 | 0 |
| 1284 | F | 39-82 | | 48535 | 44325 | 63 | 1 | #N/A | #N/A | #N/A | #N/A | 1 | 0 | 0 | 0 | 0 | 0 | 0 | 0 | 0 | 0 | 1 | 0 | 0 | |
| 1285 | F | 39-82 | | 1143 | 1085 | 0 | 0 | 437 | 472 | 0 | 0 | 0 | 0 | 0 | 1 | 0 | 0 | 0 | 0 | 0 | 0 | 0 | 0 | 0 | |
| 1286 | H | 39-82 | | 171457 | 148866 | 92 | 1 | 156992 | 82591 | 13 | 1 | 1 | 1 | 1 | 1 | 1 | 1 | 0 | 0 | 0 | 0 | 1 | 1 | 1 | 0 |
| 1287 | F | 19-38 | | 943 | 1119 | 0 | 0 | 457 | 571 | 0 | 0 | 1 | 1 | 1 | 1 | 0 | 0 | 0 | 0 | 1 | 0 | 0 | 0 | 0 | |
| 1288 | F | 19-38 | | 7260 | 2042 | 0 | 0 | 3800 | 2062 | 0 | 0 | 1 | 1 | 1 | 1 | 1 | 1 | 0 | 0 | 0 | 0 | 1 | 0 | 0 | |
| 1289 | F | 39-82 | | 3125 | 665 | 0 | 0 | 2666 | 609 | 0 | 0 | 0 | 0 | 0 | 0 | 0 | 0 | 0 | 0 | 0 | 0 | 0 | 0 | 0 | |
| 1290 | M | 19-38 | | 2803 | 4965 | 0 | 0 | #N/A | #N/A | #N/A | #N/A | 1 | 1 | 1 | 0 | 1 | 0 | 1 | 0 | 0 | 0 | 1 | 0 | 0 | |
| 1291 | F | 19-38 | | 935 | 602 | 0 | 0 | 998 | 1076 | 0 | 0 | 0 | 0 | 0 | 0 | 0 | 0 | 0 | 0 | 0 | 0 | 0 | 0 | 0 | |
| 1292 | F | 19-38 | | 554 | 1125 | 0 | 0 | 500 | 515 | 0 | 0 | 1 | 0 | 0 | 0 | 0 | 0 | 0 | 1 | 0 | 0 | 0 | 0 | 0 | |
| 1293 | M | 19-38 | | 2411 | 667 | 0 | 0 | #N/A | #N/A | #N/A | #N/A | 0 | 0 | 0 | 0 | 0 | 0 | 0 | 0 | 0 | 0 | 0 | 0 | 0 | |
| 1294 | F | 19-38 | | 6194 | 1418 | 0 | 0 | 2927 | 928 | 0 | 0 | 0 | 0 | 0 | 0 | 0 | 0 | 0 | 0 | 0 | 0 | 0 | 0 | 0 | |
| 1295 | F | 39-82 | negative | 483 | 457 | 0 | 0 | #N/A | #N/A | #N/A | #N/A | 1 | 0 | 1 | 0 | 1 | 0 | 1 | 0 | 1 | 1 | 1 | 1 | 1 | 0 |
| 1296 | H | 39-82 | | 626 | 2766 | 0 | 0 | 1152 | 1146 | 0 | 0 | 1 | 0 | 1 | 0 | 0 | 0 | 0 | 0 | 0 | 0 | 0 | 0 | 0 | |
| 1297 | F | 19-38 | | 667 | 867 | 0 | 0 | #N/A | #N/A | #N/A | #N/A | 1 | 0 | 1 | 0 | 0 | 0 | 0 | 0 | 0 | 0 | 1 | 0 | 0 | |
| 1298 | F | 39-82 | | 901 | 889 | 0 | 0 | 682 | 406 | 0 | 0 | 1 | 0 | 1 | 0 | 0 | 0 | 0 | 0 | 0 | 0 | 0 | 0 | 0 | |
| 1299 | M | 39-82 | | 6391 | 1465 | 0 | 0 | 4356 | 2767 | 0 | 0 | 1 | 0 | 1 | 0 | 0 | 0 | 0 | 0 | 1 | 0 | 0 | 0 | 0 | |
| 1301 | F | 19-38 | | 556 | 689 | 0 | 0 | 1034 | 966 | 0 | 0 | 0 | 0 | 0 | 0 | 0 | 0 | 0 | 0 | 0 | 0 | 0 | 0 | 0 | |
| 1302 | F | 19-38 | | 789 | 482 | 0 | 0 | 1397 | 494 | 0 | 0 | 1 | 0 | 1 | 0 | 1 | 1 | 1 | 0 | 0 | 0 | 0 | 1 | 0 | |
| 1303 | F | 39-82 | | 1965 | 3329 | 0 | 0 | #N/A | #N/A | #N/A | #N/A | 1 | 0 | 1 | 0 | 0 | 0 | 0 | 0 | 0 | 0 | 1 | 0 | 0 | |
| 1304 | M | 39-82 | | 2309 | 804 | 0 | 0 | 3710 | 6016 | 0 | 0 | 1 | 1 | 0 | 1 | 0 | 0 | 0 | 0 | 1 | 0 | 0 | 0 | 0 | |
| 1305 | F | 19-38 | | 2005 | 757 | 0 | 0 | 3364 | 632 | 0 | 0 | 0 | 0 | 0 | 0 | 0 | 0 | 0 | 0 | 0 | 0 | 0 | 0 | 0 | |
| 1306 | M | 39-82 | | 378 | 818 | 0 | 0 | 909 | 1037 | 0 | 0 | 0 | 0 | 0 | 0 | 0 | 0 | 0 | 0 | 0 | 0 | 0 | 0 | 0 | |
| 1307 | M | 39-82 | | 590 | 889 | 0 | 0 | #N/A | #N/A | #N/A | #N/A | 0 | 0 | 0 | 0 | 0 | 0 | 0 | 0 | 0 | 0 | 0 | 0 | 1 | 0 |
| 1308 | F | 19-38 | | 907 | 1113 | 0 | 0 | #N/A | #N/A | #N/A | #N/A | 0 | 0 | 0 | 0 | 0 | 0 | 0 | 0 | 0 | 0 | 0 | 0 | 0 | |
| 1309 | M | 19-38 | | 3541 | 5451 | 0 | 0 | 1372 | 4784 | 0 | 0 | 0 | 0 | 0 | 0 | 0 | 0 | 0 | 0 | 0 | 0 | 0 | 0 | 0 | |
| 1310 | F | 39-82 | | 849 | 2165 | 0 | 0 | 210 | 178 | 0 | 0 | 1 | 0 | 1 | 0 | 0 | 0 | 0 | 0 | 0 | 0 | 1 | 0 | 0 | |
| 1311 | F | 39-82 | | 1223 | 427 | 0 | 0 | #N/A | #N/A | #N/A | #N/A | 1 | 0 | 1 | 0 | 1 | 1 | 1 | 0 | 0 | 0 | 1 | 1 | 0 | |
| 1312 | F | 19-38 | | 967 | 555 | 0 | 0 | 722 | 603 | 0 | 0 | 0 | 0 | 0 | 0 | 0 | 0 | 0 | 0 | 0 | 0 | 0 | 0 | 0 | |
| 1313 | F | 19-38 | | 1581 | 16092 | 0 | 1 | 928 | 1225 | 0 | 0 | 0 | 0 | 0 | 0 | 0 | 0 | 0 | 0 | 0 | 0 | 0 | 0 | 0 | |
| 1314 | F | 19-38 | | 1790 | 2156 | 0 | 0 | #N/A | #N/A | #N/A | #N/A | 1 | 1 | 1 | 1 | 0 | 1 | 1 | 0 | 1 | 0 | 1 | 0 | 0 | |
| 1315 | F | 19-38 | | 1532 | 441 | 0 | 0 | #N/A | #N/A | #N/A | #N/A | 1 | 1 | 1 | 1 | 1 | 1 | 1 | 0 | 1 | 0 | 1 | 0 | 0 | |
| 1316 | F | 39-82 | | 2744 | 1156 | 0 | 0 | 1212 | 1859 | 0 | 0 | 1 | 0 | 1 | 0 | 0 | 0 | 0 | 0 | 1 | 0 | 1 | 0 | 0 | |
| 1317 | F | 19-38 | | 730 | 929 | 0 | 0 | 768 | 392 | 0 | 0 | 1 | 0 | 0 | 0 | 0 | 0 | 0 | 0 | 1 | 0 | 1 | 0 | 0 | |
| 1318 | F | 39-82 | | 1201 | 1986 | 0 | 0 | 1453 | 1406 | 0 | 0 | 0 | 0 | 0 | 0 | 0 | 0 | 0 | 0 | 0 | 0 | 0 | 0 | 0 | |
| 1319 | F | 39-82 | | 435 | 730 | 0 | 0 | 304 | 331 | 0 | 0 | 1 | 0 | 0 | 0 | 0 | 0 | 0 | 0 | 0 | 0 | 1 | 0 | 0 | |
| 1320 | F | 19-38 | | 612 | 666 | 0 | 0 | 472 | 363 | 0 | 0 | 1 | 0 | 1 | 0 | 0 | 0 | 0 | 0 | 0 | 0 | 1 | 0 | 0 | |
| 1321 | F | 19-38 | | 767 | 848 | 0 | 0 | 998 | 739 | 0 | 0 | 0 | 0 | 0 | 0 | 0 | 0 | 0 | 0 | 0 | 0 | 0 | 0 | 0 | |

| | | | | | | | | | | | | | | | | | | | | | | | | | |
|---|---|---|---|---|---|---|---|---|---|---|---|---|---|---|---|---|---|---|---|---|---|---|---|---|---|
| 1322 | F | 39-82 | | 518 | 1910 | 0 | 0 | 608 | 979 | 0 | 0 | 0 | 0 | 0 | 0 | 0 | 0 | 0 | 0 | 0 | 0 | 0 | 0 | 0 | |
| 1323 | F | 19-38 | | 8781 | 1003 | 0 | 0 | 6454 | 1049 | 0 | 0 | 1 | 1 | 1 | 1 | 1 | 1 | 0 | 0 | 0 | 0 | 0 | 1 | 0 | |
| 1324 | F | 19-38 | | 1070 | 345 | 0 | 0 | 1225 | 298 | 0 | 0 | 1 | 0 | 0 | 0 | 0 | 0 | 1 | 0 | 1 | 0 | 0 | 0 | 0 | |
| 1325 | F | 19-38 | | 777 | 660 | 0 | 0 | 1807 | 1182 | 0 | 0 | 1 | 1 | 0 | 0 | 0 | 0 | 0 | 0 | 0 | 0 | 1 | 0 | 0 | |
| 1326 | F | 19-38 | | 1511 | 6556 | 0 | 0 | 2355 | 6419 | 0 | 0 | 1 | 0 | 1 | 0 | 0 | 0 | 0 | 0 | 0 | 0 | 1 | 0 | 0 | |
| 1327 | F | 19-38 | | 83295 | 31479 | 65 | 1 | 53925 | 21510 | 8 | 1 | 1 | 1 | 1 | 0 | 0 | 0 | 1 | 0 | 0 | 0 | 1 | 0 | 1 | 1 |
| 1328 | H | 39-82 | | 150031 | 232350 | 93 | 1 | #N/A | #N/A | #N/A | #N/A | 1 | 1 | 1 | 1 | 0 | 0 | 0 | 0 | 0 | 0 | 1 | 1 | 1 | 1 |
| 1329 | H | 19-38 | | 1320 | 952 | 0 | 0 | 697 | 1005 | 0 | 0 | 1 | 1 | 1 | 1 | 0 | 0 | 1 | 0 | 1 | 0 | 1 | 0 | 0 | |
| 1330 | F | 39-82 | | 745 | 1180 | 0 | 0 | 1751 | 303 | 0 | 0 | 1 | 0 | 0 | 0 | 0 | 0 | 0 | 0 | 1 | 0 | 0 | 0 | 0 | |
| 1331 | H | 19-38 | | 1305 | 1066 | 0 | 0 | 493 | 390 | 0 | 0 | 1 | 0 | 1 | 1 | 0 | 0 | 0 | 0 | 1 | 0 | 0 | 0 | 0 | |
| 1332 | F | 19-38 | | 3513 | 1993 | 0 | 0 | #N/A | #N/A | #N/A | #N/A | 0 | 0 | 0 | 0 | 0 | 0 | 0 | 0 | 0 | 0 | 0 | 0 | 0 | |
| 1333 | F | 39-82 | | 2087 | 1151 | 0 | 0 | #N/A | #N/A | #N/A | #N/A | 0 | 0 | 0 | 0 | 0 | 0 | 0 | 0 | 0 | 0 | 0 | 0 | 0 | |
| 1334 | F | 19-38 | | 1145 | 1071 | 0 | 0 | 916 | 388 | 0 | 0 | 0 | 0 | 0 | 0 | 0 | 0 | 0 | 0 | 0 | 0 | 0 | 0 | 0 | |
| 1335 | H | 39-82 | | 2738 | 1380 | 0 | 0 | 883 | 1394 | 0 | 0 | 0 | 0 | 0 | 0 | 0 | 0 | 0 | 0 | 0 | 0 | 0 | 0 | 0 | |
| 1336 | F | 39-82 | | 616 | 1440 | 0 | 0 | #N/A | #N/A | #N/A | #N/A | 0 | 0 | 0 | 0 | 0 | 0 | 0 | 0 | 0 | 0 | 0 | 0 | 0 | |
| 1337 | H | 39-82 | | 4118 | 3923 | 0 | 0 | #N/A | #N/A | #N/A | #N/A | 0 | 0 | 0 | 0 | 0 | 0 | 0 | 0 | 0 | 0 | 0 | 0 | 0 | |
| 1338 | F | 39-82 | | 1318 | 524 | 0 | 0 | 1316 | 1073 | 0 | 0 | 0 | 0 | 0 | 0 | 0 | 0 | 0 | 0 | 0 | 0 | 0 | 0 | 0 | |
| 1340 | H | 39-82 | | 559 | 753 | 0 | 0 | 365 | 497 | 0 | 0 | 1 | 0 | 1 | 0 | 1 | 0 | 1 | 0 | 0 | 0 | 1 | 0 | 0 | |
| 1341 | H | 39-82 | | 671 | 823 | 0 | 0 | 412 | 716 | 0 | 0 | 1 | 0 | 1 | 0 | 1 | 0 | 1 | 0 | 1 | 0 | 0 | 0 | 0 | |
| 1342 | F | 19-38 | | 2904 | 1416 | 0 | 0 | 2031 | 990 | 0 | 0 | 0 | 0 | 0 | 0 | 0 | 0 | 0 | 0 | 0 | 0 | 0 | 0 | 0 | |
| 1343 | H | 39-82 | | 976 | 5323 | 0 | 0 | 1688 | 5338 | 0 | 0 | 1 | 0 | 1 | 0 | 0 | 0 | 0 | 0 | 0 | 0 | 0 | 0 | 0 | |
| 1344 | F | 39-82 | | 9250 | 1600 | 0 | 0 | 9465 | 1440 | 0 | 0 | 1 | 0 | 1 | 1 | 0 | 0 | 0 | 0 | 1 | 0 | 1 | 0 | 0 | |
| 1345 | F | 19-38 | | 1399 | 761 | 0 | 0 | 1996 | 1151 | 0 | 0 | 0 | 0 | 0 | 0 | 0 | 0 | 0 | 0 | 0 | 0 | 0 | 0 | 0 | |
| 1346 | F | 39-82 | | 1315 | 1005 | 0 | 0 | #N/A | #N/A | #N/A | #N/A | 1 | 0 | 1 | 0 | 0 | 0 | 1 | 0 | 0 | 0 | 1 | 0 | 0 | |
| 1348 | F | 39-82 | | 3225 | 3437 | 0 | 0 | #N/A | #N/A | #N/A | #N/A | 1 | 0 | 1 | 0 | 1 | 0 | 1 | 0 | 0 | 0 | 1 | 0 | 0 | |
| 1349 | F | 19-38 | | 2667 | 1373 | 0 | 0 | 2192 | 1559 | 0 | 0 | 1 | 1 | 1 | 0 | 0 | 0 | 0 | 0 | 1 | 0 | 1 | 0 | 0 | |
| 1350 | F | 39-82 | | 2929 | 1179 | 0 | 0 | 4445 | 1088 | 0 | 0 | 1 | 0 | 0 | 1 | 0 | 0 | 0 | 1 | 1 | 0 | 0 | 0 | 0 | |
| 1351 | F | 39-82 | | 1676 | 533 | 0 | 0 | 1317 | 722 | 0 | 0 | 0 | 0 | 0 | 0 | 0 | 0 | 0 | 0 | 0 | 0 | 0 | 0 | 0 | |
| 1352 | F | 19-38 | | 881 | 1433 | 0 | 0 | #N/A | #N/A | #N/A | #N/A | 0 | 0 | 0 | 0 | 0 | 0 | 0 | 0 | 0 | 0 | 0 | 0 | 0 | |
| 1353 | F | 19-38 | IgM positive | 1315 | 1344 | 0 | 0 | #N/A | #N/A | #N/A | #N/A | 1 | 1 | 1 | 1 | 1 | 1 | 1 | 0 | 1 | 1 | 1 | 0 | 0 | |
| 1354 | F | 39-82 | | 2008 | 3857 | 0 | 0 | 4674 | 3957 | 0 | 0 | 0 | 0 | 0 | 0 | 0 | 0 | 0 | 0 | 0 | 0 | 0 | 0 | 0 | |
| 1355 | F | 39-82 | | 1454 | 1928 | 0 | 0 | 2928 | 597 | 0 | 0 | 0 | 0 | 0 | 0 | 0 | 0 | 0 | 0 | 0 | 0 | 0 | 0 | 0 | |
| 1356 | F | 19-38 | | 2342 | 588 | 0 | 0 | #N/A | #N/A | #N/A | #N/A | 0 | 0 | 0 | 0 | 0 | 0 | 0 | 0 | 0 | 0 | 0 | 0 | 0 | |
| 1357 | F | 39-82 | | 3140 | 2343 | 0 | 0 | 7611 | 1137 | 0 | 0 | 0 | 0 | 0 | 0 | 0 | 0 | 0 | 0 | 0 | 0 | 0 | 0 | 0 | |
| 1358 | F | 19-38 | | 1460 | 497 | 0 | 0 | #N/A | #N/A | #N/A | #N/A | 0 | 0 | 0 | 0 | 0 | 0 | 0 | 0 | 0 | 0 | 0 | 0 | 0 | |
| 1359 | F | 19-38 | | 1617 | 1198 | 0 | 0 | 1631 | 1448 | 0 | 0 | 1 | 0 | 1 | 0 | 0 | 0 | 0 | 0 | 1 | 0 | 1 | 0 | 0 | |
| 1360 | F | 39-82 | | 908 | 743 | 0 | 0 | 1032 | 701 | 0 | 0 | 1 | 0 | 1 | 0 | 0 | 0 | 0 | 0 | 0 | 0 | 1 | 0 | 0 | |
| 1361 | F | 39-82 | IgG/IgM positi | 5320 | 19044 | 14 | 1 | 703 | 1487 | 0 | 0 | 1 | 1 | 1 | 1 | 0 | 1 | 1 | 0 | 1 | 1 | 1 | 0 | 0 | |
| 1362 | M | 19-38 | | 1257 | 526 | 0 | 0 | 136 | 198 | 0 | 0 | 0 | 0 | 0 | 0 | 0 | 0 | 0 | 0 | 0 | 0 | 0 | 0 | 0 | |
| 1363 | M | 39-82 | | 861 | 949 | 0 | 0 | #N/A | #N/A | #N/A | #N/A | 0 | 0 | 0 | 0 | 0 | 0 | 0 | 0 | 0 | 0 | 0 | 0 | 0 | |
| 1364 | F | 19-38 | | 2826 | 770 | 0 | 0 | 1577 | 753 | 0 | 0 | 1 | 0 | 1 | 1 | 0 | 0 | 0 | 0 | 0 | 0 | 1 | 0 | 0 | |
| 1365 | F | 19-38 | | 938 | 556 | 0 | 0 | 456 | 573 | 0 | 0 | 1 | 0 | 1 | 1 | 0 | 0 | 0 | 0 | 0 | 0 | 1 | 0 | 0 | |
| 1366 | M | 39-82 | | 1681 | 427 | 0 | 0 | 3206 | 1015 | 0 | 0 | 0 | 0 | 0 | 0 | 0 | 0 | 0 | 0 | 0 | 0 | 0 | 0 | 0 | |
| 1367 | F | 19-38 | | 1041 | 1030 | 0 | 0 | 715 | 1317 | 0 | 0 | 0 | 0 | 0 | 0 | 0 | 0 | 0 | 0 | 0 | 0 | 0 | 0 | 0 | |
| 1368 | M | 39-82 | | 895 | 521 | 0 | 0 | #N/A | #N/A | #N/A | #N/A | 1 | 0 | 0 | 0 | 1 | 0 | 0 | 0 | 1 | 0 | 1 | 1 | 0 | |
| 1369 | F | 19-38 | | 2105 | 234 | 0 | 0 | #N/A | #N/A | #N/A | #N/A | 1 | 0 | 1 | 1 | 0 | 0 | 0 | 0 | 0 | 0 | 1 | 1 | 1 | 0 |
| 1370 | M | 39-82 | | 1152 | 813 | 0 | 0 | 1722 | 1526 | 0 | 0 | 1 | 0 | 1 | 1 | 0 | 1 | 0 | 1 | 0 | 1 | 1 | 1 | 0 | |
| 1371 | F | 19-38 | | 886 | 581 | 0 | 0 | #N/A | #N/A | #N/A | #N/A | 1 | 0 | 1 | 0 | 0 | 0 | 0 | 0 | 1 | 0 | 1 | 0 | 0 | |
| 1373 | F | 19-38 | | 564 | 291 | 0 | 0 | 527 | 296 | 0 | 0 | 0 | 0 | 0 | 0 | 0 | 0 | 0 | 0 | 0 | 0 | 0 | 0 | 1 | 0 |
| 1374 | F | 19-38 | | 100194 | 158808 | 95 | 1 | 64147 | 99839 | 90 | 1 | 1 | 1 | 1 | 0 | 1 | 0 | 1 | 0 | 1 | 0 | 1 | 0 | 1 | 0 |
| 1375 | F | 19-38 | | 1150 | 997 | 0 | 0 | 961 | 284 | 0 | 0 | 1 | 0 | 0 | 0 | 0 | 0 | 0 | 0 | 1 | 0 | 0 | 0 | 1 | 0 |
| 1376 | F | 39-82 | | 8096 | 1094 | 0 | 0 | 8370 | 1058 | 0 | 0 | 1 | 0 | 1 | 0 | 0 | 0 | 0 | 0 | 0 | 0 | 0 | 0 | 0 | |
| 1377 | F | 19-38 | | 870 | 611 | 0 | 0 | #N/A | #N/A | #N/A | #N/A | 1 | 0 | 0 | 0 | 0 | 1 | 1 | 0 | 0 | 0 | 0 | 0 | 0 | |
| 1378 | F | 39-82 | | 1966 | 1029 | 0 | 0 | 1377 | 1257 | 0 | 0 | 0 | 0 | 0 | 0 | 0 | 0 | 0 | 0 | 0 | 0 | 0 | 0 | 0 | |
| 1379 | F | 19-38 | | 47011 | 19546 | 35 | 1 | 30724 | 19352 | 12 | 1 | 1 | 0 | 0 | 0 | 0 | 0 | 0 | 0 | 1 | 0 | 0 | 0 | 0 | |
| 1380 | F | 39-82 | | 1132 | 975 | 0 | 0 | 1460 | 1117 | 0 | 0 | 0 | 0 | 0 | 0 | 0 | 0 | 0 | 0 | 0 | 0 | 0 | 0 | 0 | |
| 1381 | F | 39-82 | | 1220 | 732 | 0 | 0 | #N/A | #N/A | #N/A | #N/A | 0 | 0 | 0 | 0 | 0 | 0 | 0 | 0 | 0 | 0 | 0 | 0 | 0 | |
| 1382 | F | 39-82 | | 577 | 1707 | 0 | 0 | 604 | 1402 | 0 | 0 | 0 | 0 | 0 | 0 | 0 | 0 | 0 | 0 | 0 | 0 | 0 | 0 | 0 | |
| 1383 | F | 19-38 | | 2396 | 604 | 0 | 0 | 3814 | 542 | 0 | 0 | 0 | 0 | 0 | 0 | 0 | 0 | 0 | 0 | 0 | 0 | 0 | 0 | 0 | |
| 1384 | F | 19-38 | | 551 | 788 | 0 | 0 | 602 | 1360 | 0 | 0 | 1 | 1 | 1 | 1 | 1 | 1 | 1 | 0 | 1 | 0 | 1 | 1 | 0 | |
| 1385 | F | 39-82 | | 115951 | 103763 | 73 | 1 | #N/A | #N/A | #N/A | #N/A | 1 | 0 | 0 | 0 | 0 | 0 | 0 | 0 | 0 | 0 | 1 | 0 | 0 | |
| 1386 | M | 19-38 | | 1287 | 998 | 0 | 0 | #N/A | #N/A | #N/A | #N/A | 0 | 0 | 0 | 0 | 0 | 0 | 0 | 0 | 0 | 0 | 0 | 0 | 0 | |
| 1387 | F | 19-38 | | 801 | 455 | 0 | 0 | #N/A | #N/A | #N/A | #N/A | 0 | 0 | 0 | 0 | 0 | 0 | 0 | 0 | 0 | 0 | 0 | 0 | 0 | |
| 1388 | M | 39-82 | | 2464 | 857 | 0 | 0 | 4407 | 638 | 0 | 0 | 0 | 0 | 0 | 0 | 0 | 0 | 0 | 0 | 0 | 0 | 0 | 0 | 0 | |
| 1389 | M | 39-82 | | 2228 | 790 | 0 | 0 | 3296 | 1035 | 0 | 0 | 0 | 0 | 0 | 0 | 0 | 0 | 0 | 0 | 0 | 0 | 0 | 0 | 0 | |
| 1390 | M | 39-82 | | 5969 | 572 | 0 | 0 | 6341 | 318 | 0 | 0 | 0 | 0 | 0 | 0 | 0 | 0 | 0 | 0 | 0 | 0 | 0 | 0 | 0 | |
| 1391 | F | 19-38 | | 1438 | 604 | 0 | 0 | 4070 | 968 | 0 | 0 | 0 | 0 | 0 | 0 | 0 | 0 | 0 | 0 | 0 | 0 | 0 | 0 | 0 | |
| 1392 | F | 19-38 | | 482 | 1163 | 0 | 0 | 1105 | 1577 | 0 | 0 | 0 | 0 | 0 | 0 | 0 | 0 | 0 | 0 | 0 | 0 | 0 | 0 | 0 | |
| 1393 | F | 19-38 | | 853 | 481 | 0 | 0 | #N/A | #N/A | #N/A | #N/A | 0 | 0 | 0 | 0 | 0 | 0 | 0 | 0 | 0 | 0 | 0 | 0 | 0 | |
| 1394 | M | 39-82 | | 2170 | 1189 | 0 | 0 | 1697 | 2924 | 0 | 0 | 0 | 0 | 0 | 0 | 0 | 0 | 0 | 0 | 0 | 0 | 0 | 0 | 0 | |
| 1395 | M | 39-82 | | 4457 | 314 | 0 | 0 | 5838 | 844 | 0 | 0 | 1 | 0 | 1 | 1 | 0 | 0 | 1 | 0 | 1 | 0 | 1 | 0 | 0 | |
| 1396 | M | 19-38 | | 2341 | 1242 | 0 | 0 | 4102 | 2710 | 0 | 0 | 0 | 0 | 0 | 0 | 0 | 0 | 0 | 0 | 0 | 0 | 0 | 0 | 0 | |
| 1397 | F | 19-38 | | 809 | 821 | 0 | 0 | 760 | 298 | 0 | 0 | 0 | 0 | 0 | 1 | 0 | 0 | 0 | 0 | 0 | 0 | 0 | 0 | 0 | |
| 1398 | M | 19-38 | | 1046 | 765 | 0 | 0 | #N/A | #N/A | #N/A | #N/A | 0 | 0 | 0 | 0 | 0 | 0 | 0 | 0 | 0 | 0 | 0 | 0 | 0 | |
| 1400 | F | 39-82 | | 1951 | 1376 | 0 | 0 | 1648 | 688 | 0 | 0 | 1 | 0 | 1 | 0 | 0 | 0 | 1 | 1 | 0 | 0 | 0 | 1 | 0 | |
| 1401 | M | 39-82 | | 2647 | 1050 | 0 | 0 | 2104 | 726 | 0 | 0 | 1 | 1 | 1 | 1 | 0 | 0 | 1 | 0 | 0 | 0 | 1 | 0 | 0 | |
| 1402 | M | 39-82 | IgG positive | 565 | 446 | 0 | 0 | 672 | 746 | 0 | 0 | 1 | 0 | 1 | 0 | 0 | 0 | 1 | 0 | 0 | 1 | 1 | 1 | 0 | |

| | | | | | | | | | | | | | | | | | | | | | | | | | |
|---|---|---|---|---|---|---|---|---|---|---|---|---|---|---|---|---|---|---|---|---|---|---|---|---|---|
| 1403 | M | 19-38 | | 1778 | 896 | 0 | 0 | #N/A | #N/A | #N/A | #N/A | 0 | 0 | 0 | 1 | 0 | 0 | 0 | 0 | 0 | 0 | 0 | 0 | 0 | |
| 1404 | F | 39-82 | | 1228 | 526 | 0 | 0 | 1548 | 868 | 0 | 0 | 0 | 0 | 0 | 0 | 0 | 0 | 0 | 0 | 0 | 0 | 0 | 0 | 0 | |
| 1405 | F | 19-38 | | 942 | 686 | 0 | 0 | #N/A | #N/A | #N/A | #N/A | 0 | 0 | 0 | 0 | 0 | 0 | 0 | 0 | 0 | 0 | 0 | 0 | 0 | |
| 1406 | F | 19-38 | | 1336 | 2529 | 0 | 0 | #N/A | #N/A | #N/A | #N/A | 1 | 0 | 1 | 0 | 0 | 0 | 0 | 0 | 1 | 0 | 1 | 0 | 0 | |
| 1407 | F | 19-38 | | 848 | 1597 | 0 | 0 | #N/A | #N/A | #N/A | #N/A | 0 | 0 | 0 | 0 | 0 | 0 | 0 | 0 | 0 | 0 | 0 | 0 | 0 | |
| 1408 | F | 39-82 | | 1674 | 1999 | 0 | 0 | 2200 | 354 | 0 | 0 | 0 | 0 | 0 | 0 | 0 | 0 | 0 | 0 | 0 | 0 | 0 | 0 | 0 | |
| 1409 | H | 19-38 | IgG/IgM positi | 8379 | 19961 | 25 | 1 | 7699 | 1052 | 24 | 1 | 1 | 0 | 0 | 0 | 0 | 0 | 0 | 0 | 0 | 1 | 0 | 0 | 0 | |
| 1410 | F | 19-38 | | 1380 | 437 | 0 | 0 | #N/A | #N/A | #N/A | #N/A | 1 | 0 | 1 | 1 | 0 | 0 | 0 | 0 | 0 | 0 | 1 | 0 | 0 | |
| 1411 | F | 19-38 | | 754 | 529 | 0 | 0 | 518 | 1866 | 0 | 0 | 0 | 0 | 0 | 0 | 0 | 0 | 0 | 0 | 0 | 0 | 0 | 0 | 0 | |
| 1412 | F | 39-82 | | 1839 | 553 | 0 | 0 | 1821 | 1145 | 0 | 0 | 1 | 1 | 1 | 1 | 0 | 0 | 0 | 0 | 1 | 0 | 1 | 0 | 0 | |
| 1413 | H | 19-38 | | 2334 | 928 | 0 | 0 | 1699 | 921 | 0 | 0 | 0 | 0 | 0 | 0 | 0 | 0 | 0 | 0 | 0 | 0 | 0 | 0 | 0 | |
| 1414 | F | 39-82 | | 997 | 589 | 0 | 0 | 1211 | 947 | 0 | 0 | 0 | 0 | 0 | 0 | 0 | 0 | 0 | 0 | 0 | 0 | 0 | 0 | 0 | |
| 1415 | F | 19-38 | | 8040 | 743 | 0 | 0 | 7520 | 638 | 0 | 0 | 0 | 0 | 0 | 0 | 0 | 0 | 0 | 0 | 0 | 0 | 0 | 0 | 0 | |
| 1416 | F | 19-38 | | 1469 | 1865 | 0 | 0 | 4390 | 975 | 0 | 0 | 0 | 0 | 0 | 0 | 0 | 0 | 0 | 0 | 0 | 0 | 0 | 0 | 0 | |
| 1417 | F | 19-38 | | 1056 | 600 | 0 | 0 | #N/A | #N/A | #N/A | #N/A | 0 | 0 | 0 | 0 | 0 | 0 | 0 | 0 | 0 | 0 | 0 | 0 | 0 | |
| 1418 | F | 19-38 | | 2863 | 489 | 0 | 0 | #N/A | #N/A | #N/A | #N/A | 0 | 0 | 0 | 0 | 0 | 0 | 0 | 0 | 0 | 0 | 0 | 0 | 0 | |
| 1419 | F | 19-38 | | 3610 | 6229 | 0 | 0 | #N/A | #N/A | #N/A | #N/A | 0 | 0 | 0 | 0 | 0 | 0 | 0 | 0 | 0 | 0 | 0 | 0 | 0 | |
| 1420 | F | 19-38 | | 5730 | 2207 | 0 | 0 | #N/A | #N/A | #N/A | #N/A | 1 | 0 | 1 | 0 | 0 | 0 | 0 | 0 | 0 | 0 | 1 | 1 | 1 | |
| 1421 | F | 19-38 | | 2173 | 789 | 0 | 0 | 617 | 456 | 0 | 0 | 1 | 0 | 1 | 0 | 0 | 0 | 0 | 0 | 0 | 0 | 1 | 0 | 0 | |
| 1422 | F | 19-38 | | 2253 | 9304 | 0 | 1 | 388 | 389 | 0 | 0 | 1 | 1 | 1 | 1 | 0 | 1 | 0 | 0 | 1 | 0 | 0 | 0 | 0 | |
| 1423 | F | 19-38 | | 4353 | 16339 | 3 | 1 | 477 | 829 | 0 | 0 | 0 | 0 | 0 | 0 | 0 | 0 | 0 | 0 | 0 | 0 | 0 | 0 | 0 | |
| 1424 | F | 39-82 | | 3323 | 1548 | 0 | 0 | 2302 | 940 | 0 | 0 | 0 | 0 | 0 | 0 | 0 | 0 | 0 | 0 | 0 | 0 | 0 | 0 | 0 | |
| 1425 | F | 39-82 | | 1374 | 760 | 0 | 0 | 508 | 316 | 0 | 0 | 1 | 0 | 1 | 0 | 0 | 0 | 0 | 0 | 0 | 0 | 0 | 0 | 1 | |
| 1426 | M | 19-38 | negative | 799 | 827 | 0 | 0 | 1321 | 787 | 0 | 0 | 1 | 0 | 1 | 1 | 1 | 1 | 0 | 0 | 1 | 1 | 0 | 0 | 0 | |
| 1427 | F | 39-82 | | 1395 | 643 | 0 | 0 | 1505 | 1341 | 0 | 0 | 1 | 0 | 1 | 0 | 0 | 0 | 0 | 0 | 0 | 0 | 0 | 0 | 0 | |
| 1428 | F | 39-82 | | 1531 | 1526 | 0 | 0 | #N/A | #N/A | #N/A | #N/A | 0 | 0 | 0 | 1 | 0 | 0 | 0 | 0 | 0 | 0 | 0 | 0 | 0 | |
| 1429 | M | 39-82 | | 2205 | 808 | 57 | 1 | #N/A | #N/A | #N/A | #N/A | 1 | 0 | 1 | 0 | 0 | 0 | 1 | 0 | 0 | 0 | 1 | 0 | 0 | |
| 1430 | F | 19-38 | | 156644 | 189949 | 96 | 1 | 125371 | 362460 | 175 | 1 | 1 | 1 | 1 | 1 | 1 | 1 | 1 | 0 | 1 | 1 | 1 | 1 | 1 | 1 |
| 1431 | M | 19-38 | | 1723 | 1744 | 0 | 0 | 1166 | 1630 | 0 | 0 | 1 | 0 | 1 | 1 | 0 | 1 | 0 | 0 | 1 | 0 | 1 | 1 | 0 | |
| 1432 | F | 19-38 | | 5499 | 3589 | 0 | 0 | 1927 | 694 | 0 | 0 | 1 | 0 | 0 | 0 | 0 | 0 | 0 | 0 | 1 | 0 | 0 | 0 | 0 | |
| 1433 | F | 39-82 | | 1392 | 752 | 0 | 0 | 1038 | 469 | 0 | 0 | 0 | 0 | 0 | 0 | 0 | 0 | 0 | 0 | 0 | 0 | 0 | 0 | 0 | |
| 1434 | F | 39-82 | | 4352 | 1114 | 0 | 0 | 3865 | 836 | 0 | 0 | 0 | 0 | 0 | 0 | 0 | 0 | 0 | 0 | 0 | 0 | 0 | 0 | 0 | |
| 1435 | F | 39-82 | | 1086 | 763 | 0 | 0 | 381 | 179 | 0 | 0 | 0 | 0 | 0 | 0 | 0 | 0 | 0 | 0 | 0 | 0 | 0 | 0 | 0 | |
| 1436 | F | 19-38 | | 2071 | 885 | 0 | 0 | #N/A | #N/A | #N/A | #N/A | 0 | 0 | 0 | 0 | 0 | 0 | 0 | 0 | 0 | 0 | 0 | 0 | 0 | |
| 1437 | F | 19-38 | | 1836 | 1058 | 0 | 0 | #N/A | #N/A | #N/A | #N/A | 1 | 1 | 0 | 0 | 1 | 0 | 0 | 0 | 0 | 0 | 0 | 0 | 1 | 0 |
| 1438 | M | 19-38 | negative | 1174 | 779 | 0 | 0 | #N/A | #N/A | #N/A | #N/A | 1 | 0 | 1 | 1 | 0 | 0 | 0 | 0 | 1 | 1 | 1 | 0 | 0 | |
| 1439 | F | 19-38 | | 1026 | 838 | 0 | 0 | 508 | 628 | 0 | 0 | 0 | 0 | 0 | 0 | 0 | 0 | 0 | 0 | 0 | 0 | 0 | 0 | 0 | |
| 1440 | F | 19-38 | | 2166 | 854 | 0 | 0 | 2183 | 622 | 0 | 0 | 0 | 0 | 0 | 0 | 0 | 0 | 0 | 0 | 0 | 0 | 0 | 0 | 0 | |
| 1441 | F | 19-38 | | 2494 | 608 | 0 | 0 | 2022 | 784 | 0 | 0 | 0 | 0 | 0 | 0 | 0 | 0 | 0 | 0 | 0 | 0 | 0 | 0 | 0 | |
| 1442 | F | 19-38 | | 2207 | 3764 | 0 | 0 | 1192 | 3471 | 0 | 0 | 1 | 0 | 1 | 0 | 0 | 0 | 1 | 0 | 1 | 0 | 1 | 0 | 1 | 0 |
| 1443 | F | 19-38 | | 1749 | 701 | 0 | 0 | 858 | 398 | 0 | 0 | 1 | 1 | 1 | 0 | 0 | 0 | 1 | 0 | 0 | 0 | 1 | 0 | 0 | |
| 1444 | M | 39-82 | | 22893 | 30496 | 61 | 1 | 17617 | 24477 | 0 | 1 | 1 | 0 | 1 | 0 | 0 | 0 | 1 | 1 | 0 | 1 | 1 | 1 | 1 | |
| 1445 | F | 39-82 | | 1217 | 849 | 0 | 0 | 1164 | 435 | 0 | 0 | 0 | 0 | 0 | 0 | 0 | 0 | 0 | 0 | 0 | 0 | 0 | 0 | 0 | |
| 1446 | F | 39-82 | | 1769 | 1093 | 0 | 0 | 980 | 765 | 0 | 0 | 0 | 0 | 0 | 0 | 0 | 0 | 0 | 0 | 0 | 0 | 0 | 0 | 0 | |
| 1447 | F | 39-82 | | 1893 | 1271 | 0 | 0 | 2019 | 621 | 0 | 0 | 0 | 0 | 0 | 0 | 0 | 0 | 0 | 0 | 0 | 0 | 0 | 0 | 0 | |
| 1448 | F | 19-38 | | 1200 | 901 | 0 | 0 | #N/A | #N/A | #N/A | #N/A | 1 | 1 | 1 | 1 | 0 | 0 | 1 | 0 | 1 | 0 | 1 | 0 | 0 | |
| 1449 | H | 39-82 | | 1524 | 911 | 0 | 0 | 389 | 310 | 0 | 0 | 1 | 1 | 1 | 0 | 1 | 1 | 1 | 0 | 0 | 0 | 1 | 0 | 1 | 0 |
| 1450 | H | 19-38 | | 1358 | 464 | 0 | 0 | 1015 | 735 | 0 | 0 | 0 | 0 | 0 | 0 | 0 | 0 | 0 | 0 | 0 | 0 | 0 | 0 | 0 | |
| 1451 | F | 39-82 | | 1189 | 714 | 0 | 0 | 1147 | 501 | 0 | 0 | 0 | 0 | 0 | 0 | 0 | 0 | 0 | 0 | 0 | 0 | 0 | 0 | 0 | |
| 1452 | H | 19-38 | | 1433 | 1114 | 0 | 0 | 804 | 808 | 0 | 0 | 0 | 0 | 0 | 0 | 0 | 0 | 0 | 0 | 0 | 0 | 0 | 0 | 0 | |
| 1453 | F | 39-82 | | 1534 | 675 | 0 | 0 | 1404 | 269 | 0 | 0 | 0 | 0 | 0 | 0 | 0 | 0 | 0 | 0 | 0 | 0 | 0 | 0 | 0 | |
| 1454 | H | 19-38 | IgG positive | 2444 | 3110 | 0 | 0 | #N/A | #N/A | #N/A | #N/A | 1 | 0 | 0 | 1 | 0 | 0 | 0 | 0 | 1 | 1 | 1 | 0 | 1 | 0 |
| 1455 | F | 39-82 | | 1025 | 3828 | 0 | 0 | 156 | 281 | 0 | 0 | 0 | 0 | 0 | 0 | 0 | 0 | 0 | 0 | 0 | 0 | 0 | 0 | 0 | |
| 1456 | F | 19-38 | | 1292 | 782 | 0 | 0 | #N/A | #N/A | #N/A | #N/A | 0 | 0 | 0 | 1 | 0 | 0 | 0 | 0 | 0 | 0 | 0 | 0 | 1 | 0 |
| 1457 | F | 19-38 | | 1829 | 4828 | 0 | 0 | 654 | 349 | 0 | 0 | 0 | 0 | 0 | 0 | 0 | 0 | 0 | 0 | 0 | 0 | 0 | 0 | 0 | |
| 1458 | H | 39-82 | | 1302 | 780 | 0 | 0 | 299 | 502 | 0 | 0 | 0 | 0 | 0 | 0 | 0 | 0 | 0 | 0 | 0 | 0 | 0 | 0 | 0 | |
| 1459 | F | 19-38 | | 2157 | 2086 | 0 | 0 | 1326 | 2305 | 0 | 0 | 1 | 0 | 1 | 0 | 0 | 0 | 0 | 0 | 0 | 0 | 1 | 0 | 0 | |
| 1460 | F | 19-38 | | 3046 | 565 | 0 | 0 | 1151 | 512 | 0 | 0 | 1 | 1 | 1 | 1 | 1 | 1 | 1 | 0 | 1 | 0 | 0 | 0 | 0 | |
| 1461 | H | 39-82 | | 1508 | 3871 | 0 | 0 | 1056 | 1895 | 0 | 0 | 0 | 0 | 0 | 0 | 0 | 0 | 0 | 0 | 0 | 0 | 0 | 0 | 0 | |
| 1462 | F | 39-82 | | 1108 | 1153 | 0 | 0 | 605 | 302 | 0 | 0 | 1 | 0 | 0 | 0 | 0 | 0 | 0 | 0 | 0 | 0 | 1 | 0 | 0 | |
| 1463 | F | 19-38 | | 3129 | 805 | 0 | 0 | #N/A | #N/A | #N/A | #N/A | 1 | 0 | 0 | 1 | 0 | 0 | 0 | 0 | 1 | 0 | 1 | 0 | 1 | 0 |
| 1464 | F | 19-38 | | 1192 | 2491 | 0 | 0 | #N/A | #N/A | #N/A | #N/A | 1 | 0 | 1 | 1 | 0 | 0 | 0 | 0 | 1 | 0 | 1 | 0 | 0 | |
| 1465 | F | 19-38 | | 595 | 911 | 0 | 0 | 485 | 903 | 0 | 0 | 0 | 0 | 0 | 0 | 0 | 0 | 0 | 0 | 0 | 0 | 0 | 0 | 0 | |
| 1466 | F | 39-82 | | 412 | 395 | 70 | 1 | 833 | 1222 | 0 | 0 | 1 | 0 | 1 | 1 | 0 | 0 | 0 | 0 | 0 | 0 | 0 | 0 | 0 | |
| 1467 | F | 19-38 | | 1184 | 532 | 0 | 0 | 910 | 552 | 0 | 0 | 0 | 0 | 0 | 0 | 0 | 0 | 0 | 0 | 0 | 0 | 0 | 0 | 0 | |
| 1469 | M | 39-82 | | 609 | 442 | 0 | 0 | #N/A | #N/A | #N/A | #N/A | 1 | 1 | 1 | 0 | 0 | 0 | 1 | 0 | 0 | 0 | 0 | 0 | 0 | |
| 1470 | F | 39-82 | | 1057 | 409 | 0 | 0 | 1582 | 727 | 0 | 0 | 0 | 0 | 0 | 0 | 0 | 0 | 0 | 0 | 0 | 0 | 0 | 0 | 0 | |
| 1471 | F | 19-38 | | 1070 | 1537 | 0 | 0 | 748 | 3695 | 0 | 0 | 0 | 0 | 0 | 0 | 0 | 0 | 0 | 0 | 0 | 0 | 0 | 0 | 0 | |
| 1472 | F | 19-38 | | 2789 | 1456 | 0 | 0 | #N/A | #N/A | #N/A | #N/A | 0 | 0 | 0 | 0 | 0 | 0 | 0 | 0 | 0 | 0 | 0 | 0 | 0 | |
| 1473 | F | 39-82 | | 2144 | 1625 | 0 | 0 | 840 | 402 | 0 | 0 | 1 | 0 | 1 | 0 | 0 | 0 | 0 | 1 | 0 | 0 | 0 | 0 | 0 | |
| 1474 | M | 19-38 | | 5481 | 3505 | 0 | 0 | 2307 | 506 | 0 | 0 | 1 | 0 | 1 | 0 | 0 | 0 | 0 | 0 | 0 | 0 | 0 | 0 | 0 | |
| 1475 | F | 19-38 | | 1384 | 986 | 0 | 0 | #N/A | #N/A | #N/A | #N/A | 0 | 0 | 0 | 0 | 0 | 0 | 0 | 0 | 0 | 0 | 0 | 0 | 0 | |
| 1476 | F | 39-82 | | 2326 | 4125 | 0 | 0 | #N/A | #N/A | #N/A | #N/A | 1 | 1 | 1 | 0 | 0 | 0 | 0 | 0 | 0 | 0 | 0 | 0 | 0 | |
| 1477 | M | 39-82 | | 39393 | 66215 | 95 | 1 | #N/A | #N/A | #N/A | #N/A | 1 | 0 | 1 | 0 | 0 | 0 | 1 | 0 | 0 | 1 | 0 | 0 | 0 | |
| 1478 | F | 19-38 | | 3511 | 2745 | 0 | 0 | 2002 | 1569 | 0 | 0 | 1 | 0 | 1 | 0 | 0 | 0 | 0 | 0 | 0 | 0 | 0 | 0 | 0 | |
| 1479 | F | 19-38 | | 736 | 532 | 0 | 0 | 433 | 254 | 0 | 0 | 1 | 0 | 1 | 1 | 0 | 1 | 0 | 0 | 0 | 0 | 1 | 0 | 0 | |
| 1480 | M | 39-82 | | 3241 | 1781 | 0 | 0 | #N/A | #N/A | #N/A | #N/A | 0 | 0 | 0 | 0 | 0 | 0 | 0 | 0 | 0 | 0 | 0 | 0 | 0 | |

| | | | | | | | | | | | | | | | | | | | | | | | | | |
|---|---|---|---|---|---|---|---|---|---|---|---|---|---|---|---|---|---|---|---|---|---|---|---|---|---|
| 1481 | M | 39-82 | | 1313 | 2350 | 0 | 0 | 701 | 373 | 0 | 0 | 0 | 0 | 0 | 0 | 0 | 0 | 0 | 0 | 0 | 0 | 0 | 0 | 0 | |
| 1482 | M | 39-82 | | 1308 | 1416 | 0 | 0 | #N/A | #N/A | #N/A | #N/A | 1 | 0 | 1 | 1 | 0 | 0 | 0 | 0 | 1 | 0 | 0 | 0 | 0 | |
| 1483 | F | 19-38 | | 997 | 1408 | 0 | 0 | 407 | 834 | 0 | 0 | 1 | 0 | 1 | 1 | 0 | 0 | 0 | 0 | 0 | 0 | 1 | 0 | 0 | |
| 1484 | F | 19-38 | | 576 | 1857 | 0 | 0 | 520 | 492 | 0 | 0 | 1 | 1 | 1 | 0 | 0 | 0 | 0 | 0 | 1 | 0 | 1 | 0 | 0 | |
| 1485 | F | 19-38 | | 840 | 1273 | 0 | 0 | #N/A | #N/A | #N/A | #N/A | 1 | 1 | 1 | 0 | 1 | 0 | 1 | 0 | 1 | 0 | 1 | 0 | 1 | 0 |
| 1486 | F | 19-38 | | 924 | 2190 | 0 | 0 | 5510 | 871 | 0 | 0 | 1 | 1 | 1 | 0 | 0 | 0 | 0 | 0 | 0 | 0 | 1 | 0 | 0 | |
| 1487 | F | 19-38 | | 1794 | 1473 | 0 | 0 | 1196 | 745 | 0 | 0 | 0 | 0 | 0 | 0 | 0 | 0 | 0 | 0 | 0 | 0 | 0 | 0 | 0 | |
| 1488 | F | 19-38 | | 2170 | 2162 | 0 | 0 | 2946 | 2346 | 0 | 0 | 1 | 1 | 1 | 0 | 0 | 0 | 1 | 0 | 0 | 0 | 1 | 0 | 0 | |
| 1489 | F | 39-82 | | 2187 | 1323 | 0 | 0 | 856 | 564 | 0 | 0 | 1 | 0 | 1 | 1 | 1 | 0 | 0 | 0 | 1 | 0 | 1 | 1 | 0 | |
| 1490 | F | 19-38 | | 11837 | 915 | 0 | 1 | 10177 | 1634 | 0 | 0 | 1 | 1 | 1 | 1 | 0 | 0 | 0 | 0 | 1 | 0 | 0 | 0 | 1 | 0 |
| 1491 | M | 19-38 | | 4668 | 2023 | 0 | 0 | 2817 | 390 | 0 | 0 | 0 | 0 | 0 | 0 | 0 | 0 | 0 | 0 | 0 | 0 | 0 | 0 | 0 | |
| 1492 | M | 19-38 | | 845 | 623 | 0 | 0 | 394 | 695 | 0 | 0 | 0 | 0 | 0 | 0 | 0 | 0 | 0 | 0 | 0 | 0 | 0 | 0 | 0 | |
| 1493 | F | 19-38 | | 1366 | 3256 | 0 | 0 | 1045 | 2201 | 0 | 0 | 1 | 0 | 1 | 0 | 0 | 0 | 0 | 0 | 1 | 0 | 0 | 0 | 0 | |
| 1494 | M | 19-38 | | 16399 | 1045 | 0 | 1 | 12909 | 926 | 0 | 1 | 0 | 0 | 0 | 1 | 0 | 0 | 0 | 0 | 0 | 0 | 0 | 0 | 0 | |
| 1495 | F | 39-82 | | 1780 | 2341 | 0 | 0 | 1161 | 794 | 0 | 0 | 1 | 0 | 1 | 0 | 0 | 0 | 0 | 0 | 0 | 0 | 0 | 0 | 1 | 0 |
| 1496 | F | 39-82 | | 1638 | 1014 | 0 | 0 | 1273 | 957 | 0 | 0 | 1 | 0 | 1 | 0 | 0 | 0 | 0 | 0 | 0 | 0 | 0 | 0 | 0 | |
| 1497 | F | 39-82 | | 1302 | 1125 | 0 | 0 | 1156 | 757 | 0 | 0 | 1 | 1 | 0 | 0 | 0 | 0 | 0 | 0 | 0 | 0 | 1 | 0 | 0 | |
| 1498 | F | 19-38 | | 2635 | 1640 | 0 | 0 | 1380 | 931 | 0 | 0 | 1 | 0 | 0 | 0 | 0 | 0 | 0 | 0 | 0 | 0 | 1 | 1 | 0 | |
| 1499 | F | 39-82 | | 1619 | 1089 | 0 | 0 | 1224 | 341 | 0 | 0 | 0 | 0 | 0 | 0 | 0 | 0 | 0 | 0 | 0 | 0 | 0 | 0 | 0 | |
| 1500 | F | 39-82 | | 1920 | 1272 | 0 | 0 | 1314 | 926 | 0 | 0 | 1 | 0 | 1 | 0 | 0 | 0 | 1 | 0 | 0 | 0 | 1 | 0 | 0 | |
| 1501 | F | 39-82 | | 91751 | 207447 | 98 | 1 | 65074 | 170311 | 99 | 1 | 1 | 1 | 1 | 1 | 1 | 0 | 0 | 0 | 1 | 1 | 1 | 1 | 0 | |
| 1502 | F | 39-82 | | 3564 | 1132 | 0 | 0 | 3445 | 552 | 0 | 0 | 1 | 0 | 0 | 1 | 1 | 1 | 0 | 0 | 0 | 0 | 0 | 0 | 0 | |
| 1503 | F | 39-82 | | 1599 | 955 | 0 | 0 | #N/A | #N/A | #N/A | #N/A | 1 | 1 | 1 | 0 | 0 | 0 | 1 | 0 | 1 | 0 | 1 | 0 | 0 | |
| 1504 | F | 39-82 | | 1503 | 1387 | 0 | 0 | 907 | 1095 | 0 | 0 | 1 | 0 | 0 | 0 | 0 | 0 | 0 | 0 | 1 | 0 | 0 | 0 | 0 | |
| 1505 | F | 19-38 | | 1382 | 1382 | 0 | 0 | #N/A | #N/A | #N/A | #N/A | 0 | 0 | 0 | 0 | 0 | 0 | 0 | 0 | 0 | 0 | 0 | 0 | 0 | |
| 1506 | F | 39-82 | | 2035 | 1416 | 0 | 0 | 1052 | 462 | 0 | 0 | 0 | 0 | 0 | 0 | 0 | 0 | 0 | 0 | 0 | 0 | 0 | 0 | 0 | |
| 1507 | F | 39-82 | | 3593 | 2385 | 0 | 0 | 3243 | 660 | 0 | 0 | 0 | 0 | 0 | 0 | 0 | 0 | 0 | 0 | 0 | 0 | 0 | 0 | 0 | |
| 1508 | F | 19-38 | | 1046 | 1064 | 0 | 0 | 662 | 361 | 0 | 0 | 0 | 0 | 0 | 0 | 0 | 0 | 0 | 0 | 0 | 0 | 0 | 0 | 0 | |
| 1509 | F | 19-38 | | 1039 | 1796 | 0 | 0 | 698 | 251 | 0 | 0 | 0 | 0 | 0 | 0 | 0 | 0 | 0 | 0 | 0 | 0 | 0 | 0 | 0 | |
| 1510 | F | 19-38 | | 1211 | 682 | 0 | 0 | 602 | 384 | 0 | 0 | 0 | 0 | 0 | 0 | 0 | 0 | 0 | 0 | 0 | 0 | 0 | 0 | 0 | |
| 1511 | H | 39-82 | | 2038 | 1989 | 0 | 0 | 1702 | 1076 | 0 | 0 | 0 | 0 | 0 | 0 | 0 | 0 | 0 | 0 | 0 | 0 | 0 | 0 | 0 | |
| 1512 | F | 39-82 | | 7751 | 2316 | 0 | 0 | 8891 | 839 | 0 | 0 | 1 | 0 | 1 | 1 | 1 | 0 | 0 | 0 | 1 | 0 | 1 | 1 | 0 | |
| 1513 | F | 39-82 | | 879 | 871 | 0 | 0 | 588 | 434 | 0 | 0 | 0 | 0 | 0 | 0 | 0 | 0 | 0 | 0 | 0 | 0 | 0 | 0 | 0 | |
| 1514 | F | 19-38 | | 1924 | 3846 | 0 | 0 | 804 | 1637 | 0 | 0 | 1 | 0 | 1 | 0 | 0 | 0 | 0 | 0 | 1 | 0 | 1 | 0 | 0 | |
| 1515 | F | 19-38 | | 2023 | 1986 | 0 | 0 | #N/A | #N/A | #N/A | #N/A | 1 | 0 | 0 | 0 | 0 | 0 | 1 | 0 | 1 | 0 | 1 | 0 | 0 | |
| 1516 | F | 39-82 | | 3011 | 2265 | 85 | 1 | #N/A | #N/A | #N/A | #N/A | 0 | 0 | 0 | 0 | 0 | 0 | 0 | 0 | 0 | 0 | 0 | 0 | 1 | 0 |
| 1517 | F | 39-82 | | 1487 | 1252 | 0 | 0 | 483 | 284 | 0 | 0 | 0 | 0 | 0 | 0 | 0 | 0 | 0 | 0 | 0 | 0 | 0 | 0 | 0 | |
| 1518 | M | 19-38 | | 5433 | 4326 | 0 | 0 | 3634 | 2944 | 0 | 0 | 0 | 0 | 0 | 0 | 0 | 0 | 0 | 0 | 0 | 0 | 0 | 0 | 0 | |
| 1519 | F | 19-38 | | 1790 | 1784 | 0 | 0 | 816 | 620 | 0 | 0 | 0 | 0 | 0 | 0 | 0 | 0 | 0 | 0 | 0 | 0 | 0 | 0 | 0 | |
| 1520 | F | 39-82 | | 1323 | 1054 | 0 | 0 | 463 | 413 | 0 | 0 | 0 | 0 | 0 | 0 | 0 | 0 | 0 | 0 | 0 | 0 | 0 | 0 | 0 | |
| 1521 | M | 39-82 | | 3931 | 1692 | 0 | 0 | #N/A | #N/A | #N/A | #N/A | 0 | 0 | 0 | 0 | 0 | 0 | 0 | 0 | 0 | 0 | 0 | 0 | 0 | |
| 1522 | F | 39-82 | | 1962 | 1511 | 0 | 0 | 1445 | 774 | 0 | 0 | 1 | 0 | 0 | 0 | 0 | 0 | 0 | 1 | 0 | 0 | 0 | 0 | 0 | |
| 1523 | M | 39-82 | IgG/IgM positi | 10393 | 29095 | 87 | 1 | #N/A | #N/A | #N/A | #N/A | 1 | 1 | 1 | 0 | 0 | 0 | 1 | 0 | 1 | 1 | 1 | 0 | 0 | |
| 1524 | F | 19-38 | | 1419 | 1159 | 0 | 0 | 888 | 394 | 0 | 0 | 0 | 0 | 0 | 0 | 0 | 0 | 0 | 0 | 0 | 0 | 0 | 0 | 0 | |
| 1525 | F | 39-82 | | 2387 | 1858 | 0 | 0 | 1407 | 983 | 0 | 0 | 0 | 0 | 0 | 0 | 0 | 0 | 0 | 0 | 0 | 0 | 0 | 0 | 0 | |
| 1526 | F | 19-38 | | 1739 | 1591 | 0 | 0 | 1331 | 1440 | 0 | 0 | 1 | 1 | 1 | 1 | 0 | 0 | 0 | 0 | 0 | 0 | 1 | 0 | 0 | |
| 1527 | H | 39-82 | | 2452 | 2792 | 0 | 0 | #N/A | #N/A | #N/A | #N/A | 0 | 0 | 0 | 0 | 0 | 0 | 0 | 0 | 0 | 0 | 0 | 0 | 0 | |
| 1528 | F | 19-38 | | 4689 | 1515 | 0 | 0 | 3160 | 617 | 0 | 0 | 0 | 0 | 0 | 0 | 0 | 0 | 0 | 0 | 0 | 0 | 0 | 0 | 0 | |
| 1529 | F | 39-82 | | 1874 | 1371 | 0 | 0 | 1165 | 788 | 0 | 0 | 0 | 0 | 0 | 0 | 0 | 0 | 0 | 0 | 0 | 0 | 0 | 0 | 0 | |
| 1530 | F | 39-82 | | 1147 | 1257 | 0 | 0 | 552 | 559 | 0 | 0 | 1 | 0 | 1 | 0 | 1 | 1 | 0 | 0 | 0 | 0 | 0 | 0 | 0 | |
| 1531 | F | 39-82 | | 60609 | 45523 | 80 | 1 | 54156 | 256378 | 6 | 1 | 0 | 0 | 0 | 0 | 0 | 0 | 0 | 0 | 0 | 0 | 0 | 0 | 0 | |
| 1532 | F | 19-38 | | 1783 | 1482 | 0 | 0 | 474 | 542 | 0 | 0 | 1 | 1 | 1 | 1 | 1 | 1 | 0 | 0 | 0 | 0 | 1 | 0 | 0 | |
| 1533 | H | 19-38 | | 5689 | 1565 | 0 | 0 | #N/A | #N/A | #N/A | #N/A | 1 | 0 | 1 | 0 | 0 | 0 | 0 | 0 | 0 | 0 | 1 | 0 | 0 | |
| 1534 | F | 19-38 | | 1290 | 838 | 0 | 0 | 427 | 213 | 0 | 0 | 1 | 1 | 1 | 0 | 0 | 0 | 0 | 0 | 0 | 0 | 1 | 0 | 0 | |
| 1535 | F | 19-38 | negative | 2626 | 1697 | 0 | 0 | #N/A | #N/A | #N/A | #N/A | 1 | 1 | 1 | 1 | 1 | 1 | 1 | 0 | 0 | 1 | 1 | 0 | 0 | |
| 1537 | F | 39-82 | | 2607 | 4857 | 0 | 0 | 1803 | 2331 | 0 | 0 | 0 | 0 | 0 | 0 | 0 | 0 | 0 | 0 | 0 | 0 | 0 | 0 | 0 | |
| 1538 | F | 19-38 | | 964 | 1903 | 0 | 0 | 888 | 779 | 0 | 0 | 0 | 0 | 0 | 0 | 0 | 0 | 0 | 0 | 0 | 0 | 0 | 0 | 0 | |
| 1539 | F | 19-38 | | 1017 | 1880 | 0 | 0 | 1289 | 940 | 0 | 0 | 0 | 0 | 0 | 0 | 0 | 0 | 0 | 0 | 0 | 0 | 0 | 0 | 0 | |
| 1540 | F | 19-38 | | 8586 | 4780 | 0 | 0 | 4574 | 4443 | 0 | 0 | 0 | 0 | 0 | 0 | 0 | 0 | 0 | 0 | 0 | 0 | 0 | 0 | 0 | |
| 1541 | F | 19-38 | | 2121 | 867 | 0 | 0 | 713 | 263 | 0 | 0 | 1 | 0 | 0 | 0 | 0 | 0 | 0 | 0 | 0 | 0 | 1 | 0 | 0 | |
| 1542 | F | 19-38 | | 1626 | 1257 | 0 | 0 | 695 | 1006 | 0 | 0 | 1 | 0 | 0 | 1 | 0 | 0 | 0 | 0 | 0 | 0 | 1 | 0 | 0 | |
| 1543 | M | 39-82 | | 1313 | 826 | 0 | 0 | #N/A | #N/A | #N/A | #N/A | 0 | 0 | 0 | 0 | 0 | 0 | 0 | 0 | 0 | 0 | 0 | 0 | 0 | |
| 1544 | M | 19-38 | | 1194 | 2471 | 0 | 0 | #N/A | #N/A | #N/A | #N/A | 0 | 0 | 0 | 0 | 0 | 0 | 0 | 0 | 0 | 0 | 0 | 0 | 0 | |
| 1545 | M | 39-82 | | 1529 | 6027 | 0 | 0 | 830 | 5688 | 0 | 0 | 1 | 1 | 1 | 1 | 0 | 1 | 1 | 1 | 1 | 0 | 1 | 0 | 0 | |
| 1546 | M | 19-38 | | 1577 | 1248 | 0 | 0 | #N/A | #N/A | #N/A | #N/A | 0 | 0 | 0 | 0 | 0 | 0 | 0 | 0 | 0 | 0 | 0 | 0 | 0 | |
| 1548 | F | 19-38 | | 934 | 1661 | 0 | 0 | 4052 | 2004 | 34 | 1 | 0 | 0 | 0 | 0 | 0 | 0 | 0 | 0 | 0 | 0 | 0 | 0 | 1 | |
| 1549 | F | 39-82 | | 2304 | 1070 | 0 | 0 | 1729 | 432 | 0 | 0 | 1 | 0 | 1 | 0 | 0 | 0 | 1 | 0 | 0 | 0 | 0 | 0 | 0 | |
| 1550 | F | 39-82 | | 3826 | 1096 | 0 | 0 | 1480 | 459 | 0 | 0 | 1 | 0 | 1 | 0 | 0 | 0 | 0 | 0 | 0 | 0 | 1 | 1 | 0 | |
| 1551 | M | 39-82 | | 2895 | 1103 | 0 | 0 | #N/A | #N/A | #N/A | #N/A | 1 | 0 | 1 | 0 | 0 | 0 | 1 | 0 | 0 | 0 | 0 | 0 | 0 | |
| 1553 | F | 39-82 | | 2041 | 5955 | 0 | 0 | #N/A | #N/A | #N/A | #N/A | 1 | 1 | 1 | 1 | 1 | 1 | 1 | 0 | 1 | 0 | 1 | 0 | 1 | 0 |
| 1555 | F | 39-82 | | 2360 | 1006 | 0 | 0 | 1509 | 287 | 0 | 0 | 0 | 0 | 0 | 0 | 0 | 0 | 0 | 0 | 0 | 0 | 0 | 0 | 0 | |
| 1556 | F | 39-82 | | 2362 | 652 | 0 | 0 | 1596 | 1736 | 0 | 0 | 0 | 0 | 0 | 0 | 0 | 0 | 0 | 0 | 0 | 0 | 0 | 0 | 0 | |
| 1557 | F | 39-82 | | 1318 | 1770 | 0 | 0 | #N/A | #N/A | #N/A | #N/A | 0 | 0 | 0 | 0 | 0 | 0 | 0 | 0 | 0 | 0 | 0 | 0 | 0 | |
| 1558 | F | 19-38 | | 1658 | 1001 | 0 | 0 | 1246 | 365 | 0 | 0 | 1 | 0 | 0 | 0 | 0 | 0 | 1 | 0 | 0 | 0 | 1 | 0 | 0 | |
| 1559 | F | 39-82 | | 6736 | 12144 | 0 | 1 | 6542 | 795 | 0 | 0 | 1 | 1 | 1 | 0 | 0 | 0 | 1 | 0 | 0 | 0 | 1 | 1 | 0 | |
| 1560 | F | 39-82 | | 1705 | 5652 | 0 | 0 | 1599 | 5904 | 0 | 0 | 1 | 0 | 1 | 1 | 1 | 1 | 1 | 0 | 1 | 0 | 1 | 1 | 0 | |
| 1561 | M | 19-38 | | 3710 | 4344 | 0 | 0 | 3265 | 1274 | 0 | 0 | 0 | 0 | 0 | 0 | 0 | 0 | 0 | 0 | 0 | 0 | 0 | 0 | 0 | |

| | | | | | | | | | | | | | | | | | | | | | | | | |
|---|---|---|---|---|---|---|---|---|---|---|---|---|---|---|---|---|---|---|---|---|---|---|---|---|
| 1562 | M | 19-38 | 1918 | 1001 | 0 | 0 | 666 | 289 | 0 | 0 | 1 | 0 | 1 | 0 | 0 | 0 | 0 | 0 | 0 | 0 | 0 | 0 | 0 | |
| 1563 | F | 39-82 | 1825 | 1982 | 0 | 0 | 779 | 1031 | 0 | 0 | 1 | 0 | 1 | 0 | 0 | 0 | 0 | 0 | 0 | 0 | 1 | 0 | 0 | |
| 1564 | M | 39-82 | 2548 | 3177 | 0 | 0 | 684 | 1313 | 0 | 0 | 1 | 0 | 1 | 0 | 0 | 0 | 1 | 0 | 0 | 0 | 1 | 0 | 0 | |
| 1565 | M | 39-82 | 8887 | 1361 | 0 | 0 | 5284 | 660 | 0 | 0 | 1 | 0 | 1 | 1 | 0 | 0 | 0 | 0 | 0 | 0 | 1 | 0 | 0 | |
| 1566 | F | 39-82 | 1231 | 1206 | 0 | 0 | 620 | 698 | 0 | 0 | 0 | 0 | 0 | 0 | 0 | 0 | 0 | 0 | 0 | 0 | 0 | 0 | 0 | |
| 1567 | M | 39-82 | 4850 | 1063 | 0 | 0 | 1976 | 1227 | 0 | 0 | 1 | 0 | 1 | 0 | 0 | 0 | 0 | 0 | 0 | 0 | 1 | 0 | 0 | |
| 1568 | M | 19-38 | 1126 | 1213 | 0 | 0 | 195 | 681 | 0 | 0 | 1 | 0 | 1 | 0 | 0 | 0 | 0 | 0 | 0 | 0 | 1 | 1 | 0 | |
| 1569 | F | 39-82 | 2430 | 1574 | 0 | 0 | 1113 | 287 | 0 | 0 | 0 | 0 | 0 | 0 | 0 | 0 | 0 | 0 | 0 | 0 | 0 | 0 | 0 | |
| 1570 | F | 19-38 | 1103 | 1855 | 0 | 0 | #N/A | #N/A | #N/A | #N/A | 1 | 0 | 1 | 0 | 0 | 0 | 0 | 0 | 0 | 0 | 1 | 0 | 0 | |
| 1571 | F | 39-82 | 1710 | 826 | 0 | 0 | 1728 | 424 | 0 | 0 | 0 | 0 | 0 | 0 | 0 | 0 | 0 | 0 | 0 | 0 | 0 | 0 | 0 | |
| 1572 | F | 19-38 | 892 | 1470 | 0 | 0 | #N/A | #N/A | #N/A | #N/A | 1 | 0 | 0 | 1 | 0 | 0 | 0 | 0 | 0 | 0 | 1 | 0 | 0 | |
| 1573 | F | 39-82 | 1498 | 1270 | 0 | 0 | 1096 | 616 | 0 | 0 | 1 | 0 | 0 | 0 | 0 | 0 | 0 | 0 | 0 | 0 | 1 | 0 | 0 | |
| 1574 | F | 19-38 | 1201 | 3922 | 0 | 0 | 427 | 2181 | 0 | 0 | 0 | 0 | 0 | 0 | 0 | 0 | 0 | 0 | 0 | 0 | 0 | 0 | 0 | |
| 1575 | F | 39-82 | 4003 | 1857 | 0 | 0 | 3500 | 1154 | 0 | 0 | 0 | 0 | 0 | 0 | 0 | 0 | 0 | 0 | 0 | 0 | 0 | 0 | 0 | |
| 1576 | F | 39-82 | 3555 | 1282 | 0 | 0 | #N/A | #N/A | #N/A | #N/A | 1 | 0 | 1 | 0 | 0 | 0 | 0 | 0 | 0 | 0 | 0 | 0 | 1 | 0 |
| 1577 | F | 39-82 | 2305 | 1883 | 0 | 0 | 1441 | 1013 | 0 | 0 | 0 | 0 | 0 | 0 | 0 | 0 | 0 | 0 | 0 | 0 | 0 | 0 | 0 | |
| 1578 | F | 39-82 | 1345 | 1982 | 0 | 0 | 679 | 996 | 0 | 0 | 1 | 1 | 1 | 0 | 0 | 0 | 0 | 0 | 0 | 0 | 1 | 0 | 0 | |
| 1579 | H | 19-38 | 1577 | 1647 | 0 | 0 | #N/A | #N/A | #N/A | #N/A | 0 | 0 | 0 | 0 | 0 | 0 | 0 | 0 | 0 | 0 | 0 | 0 | 0 | |
| 1580 | F | 19-38 | 1623 | 1384 | 0 | 0 | #N/A | #N/A | #N/A | #N/A | 0 | 0 | 0 | 0 | 0 | 0 | 0 | 0 | 0 | 0 | 0 | 0 | 0 | |
| 1581 | F | 39-82 | 3259 | 1764 | 0 | 0 | #N/A | #N/A | #N/A | #N/A | 0 | 0 | 0 | 0 | 0 | 0 | 0 | 0 | 0 | 0 | 0 | 0 | 0 | |
| 1582 | H | 19-38 | 4502 | 1695 | 0 | 0 | 2361 | 2352 | 0 | 0 | 0 | 0 | 0 | 0 | 0 | 0 | 0 | 0 | 0 | 0 | 0 | 0 | 0 | |
| 1583 | F | 19-38 | 4260 | 2713 | 0 | 0 | #N/A | #N/A | #N/A | #N/A | 1 | 0 | 1 | 0 | 0 | 0 | 1 | 0 | 0 | 0 | 1 | 1 | 0 | |
| 1584 | F | 39-82 | 1197 | 720 | 0 | 0 | 536 | 211 | 0 | 0 | 0 | 0 | 0 | 0 | 0 | 0 | 0 | 0 | 0 | 0 | 0 | 0 | 0 | |
| 1585 | F | 19-38 | 1676 | 1967 | 0 | 0 | #N/A | #N/A | #N/A | #N/A | 0 | 0 | 0 | 0 | 0 | 0 | 0 | 0 | 0 | 0 | 0 | 0 | 0 | |
| 1586 | F | 19-38 | 6673 | 2388 | 0 | 0 | #N/A | #N/A | #N/A | #N/A | 1 | 1 | 1 | 1 | 1 | 0 | 1 | 0 | 1 | 0 | 0 | 0 | 0 | |
| 1587 | F | 19-38 | 3120 | 1553 | 0 | 0 | 4170 | 943 | 0 | 0 | 0 | 0 | 0 | 0 | 0 | 0 | 0 | 0 | 0 | 0 | 0 | 0 | 0 | |
| 1588 | F | 19-38 | 2337 | 10538 | 0 | 1 | 2357 | 1020 | 0 | 0 | 0 | 0 | 0 | 0 | 0 | 0 | 0 | 0 | 0 | 0 | 0 | 0 | 0 | |
| 1589 | F | 39-82 | 4932 | 832 | 0 | 0 | 3548 | 176 | 0 | 0 | 0 | 0 | 0 | 0 | 0 | 0 | 0 | 0 | 0 | 0 | 0 | 0 | 0 | |
| 1590 | F | 39-82 | 1467 | 3128 | 0 | 0 | 836 | 2189 | 0 | 0 | 1 | 1 | 0 | 0 | 0 | 0 | 0 | 0 | 1 | 0 | 1 | 0 | 0 | |
| 1591 | M | 19-38 | 1437 | 4198 | 0 | 0 | #N/A | #N/A | #N/A | #N/A | 0 | 0 | 0 | 0 | 0 | 0 | 0 | 0 | 0 | 0 | 0 | 0 | 0 | |
| 1592 | F | 39-82 | 1180 | 1398 | 0 | 0 | 960 | 1560 | 0 | 0 | 1 | 0 | 1 | 1 | 0 | 1 | 1 | 0 | 1 | 0 | 1 | 0 | 1 | 0 |
| 1593 | F | 19-38 | 9858 | 865 | 0 | 0 | 9041 | 427 | 0 | 0 | 0 | 0 | 0 | 0 | 0 | 0 | 0 | 0 | 0 | 0 | 0 | 0 | 0 | |
| 1594 | F | 39-82 | 2070 | 1419 | 0 | 0 | 1154 | 437 | 0 | 0 | 0 | 0 | 0 | 0 | 0 | 0 | 0 | 0 | 0 | 0 | 0 | 0 | 0 | |
| 1595 | M | 39-82 | 1197 | 1446 | 0 | 0 | 471 | 751 | 0 | 0 | 0 | 0 | 0 | 0 | 0 | 0 | 0 | 0 | 0 | 0 | 0 | 0 | 0 | |
| 1596 | F | 19-38 | 2068 | 1685 | 0 | 0 | 4489 | 973 | 0 | 0 | 1 | 0 | 1 | 0 | 0 | 0 | 0 | 1 | 0 | 0 | 0 | 0 | 0 | |
| 1597 | F | 19-38 | 2876 | 6306 | 0 | 0 | #N/A | #N/A | #N/A | #N/A | 1 | 0 | 0 | 0 | 0 | 0 | 1 | 0 | 0 | 0 | 0 | 0 | 1 | |
| 1598 | M | 39-82 | 212526 | 324261 | 96 | 1 | 168511 | 119701 | 87 | 1 | 1 | 0 | 1 | 0 | 0 | 0 | 0 | 0 | 0 | 0 | 0 | 0 | 0 | |
| 1599 | F | 19-38 | 3086 | 877 | 0 | 0 | #N/A | #N/A | #N/A | #N/A | 0 | 0 | 0 | 0 | 0 | 0 | 0 | 0 | 0 | 0 | 0 | 0 | 0 | |
| 1600 | F | 39-82 | 2187 | 859 | 0 | 0 | 1432 | 1105 | 0 | 0 | 0 | 0 | 0 | 0 | 0 | 0 | 0 | 0 | 0 | 0 | 0 | 0 | 0 | |
| 1601 | F | 39-82 | 1591 | 1698 | 0 | 0 | #N/A | #N/A | #N/A | #N/A | 1 | 0 | 0 | 0 | 0 | 0 | 0 | 0 | 0 | 0 | 1 | 1 | 1 | 0 |
| 1602 | F | 19-38 | 1021 | 1228 | 0 | 0 | #N/A | #N/A | #N/A | #N/A | 1 | 0 | 0 | 1 | 0 | 0 | 0 | 0 | 1 | 0 | 0 | 0 | 1 | 0 |
| 1603 | F | 19-38 | 2940 | 2267 | 0 | 0 | #N/A | #N/A | #N/A | #N/A | 1 | 0 | 1 | 0 | 0 | 1 | 0 | 0 | 0 | 0 | 1 | 0 | 0 | |
| 1604 | H | 39-82 | 1235 | 810 | 0 | 0 | #N/A | #N/A | #N/A | #N/A | 1 | 0 | 0 | 1 | 0 | 0 | 1 | 0 | 0 | 0 | 0 | 0 | 0 | |
| 1605 | F | 19-38 | 4397 | 2766 | 0 | 0 | #N/A | #N/A | #N/A | #N/A | 0 | 0 | 0 | 0 | 0 | 0 | 0 | 0 | 0 | 0 | 0 | 0 | 0 | |
| 1606 | F | 19-38 | 2503 | 1752 | 0 | 0 | #N/A | #N/A | #N/A | #N/A | 1 | 0 | 1 | 0 | 1 | 1 | 1 | 0 | 0 | 0 | 0 | 1 | 0 | |
| 1607 | F | 19-38 | 973 | 1731 | 0 | 0 | #N/A | #N/A | #N/A | #N/A | 0 | 0 | 0 | 0 | 0 | 0 | 0 | 0 | 0 | 0 | 0 | 0 | 0 | |
| 1608 | F | 19-38 | 3409 | 1686 | 0 | 0 | 2375 | 853 | 0 | 0 | 1 | 0 | 1 | 0 | 0 | 0 | 0 | 0 | 0 | 0 | 0 | 1 | 0 | |
| 1609 | F | 39-82 | 1540 | 1790 | 0 | 0 | #N/A | #N/A | #N/A | #N/A | 0 | 0 | 0 | 0 | 0 | 0 | 0 | 0 | 0 | 0 | 0 | 0 | 0 | |
| 1610 | F | 39-82 | 7652 | 2020 | 0 | 0 | 6396 | 997 | 0 | 0 | 1 | 0 | 1 | 0 | 0 | 0 | 0 | 0 | 0 | 0 | 0 | 1 | 0 | |
| 1611 | F | 19-38 | 2570 | 1411 | 0 | 0 | 1324 | 551 | 0 | 0 | 0 | 0 | 0 | 0 | 0 | 0 | 0 | 0 | 0 | 0 | 0 | 0 | 0 | |
| 1612 | H | 39-82 | 2384 | 1545 | 0 | 0 | 1741 | 1394 | 0 | 0 | 1 | 0 | 1 | 0 | 0 | 0 | 0 | 0 | 0 | 0 | 0 | 0 | 0 | |
| 1613 | H | 39-82 | 18869 | 40418 | 56 | 1 | 12459 | 36352 | 21 | 1 | 1 | 1 | 0 | 0 | 0 | 0 | 1 | 0 | 0 | 1 | 0 | 1 | 0 | |
| 1614 | F | 19-38 | 2030 | 3004 | 0 | 0 | 1664 | 2885 | 0 | 0 | 1 | 1 | 1 | 0 | 0 | 0 | 0 | 0 | 0 | 0 | 0 | 0 | 0 | |
| 1615 | F | 19-38 | 1240 | 834 | 0 | 0 | #N/A | #N/A | #N/A | #N/A | 0 | 0 | 0 | 0 | 0 | 0 | 0 | 0 | 0 | 0 | 0 | 0 | 0 | |
| 1616 | F | 19-38 | 997 | 1142 | 0 | 0 | #N/A | #N/A | #N/A | #N/A | 1 | 1 | 0 | 0 | 0 | 0 | 0 | 0 | 1 | 0 | 1 | 0 | 0 | |
| 1617 | F | 19-38 | 9432 | 668 | 0 | 0 | 11396 | 800 | 0 | 1 | 0 | 0 | 0 | 0 | 0 | 0 | 0 | 0 | 0 | 0 | 0 | 0 | 0 | |
| 1618 | F | 19-38 | 1231 | 2946 | 0 | 0 | #N/A | #N/A | #N/A | #N/A | 1 | 0 | 1 | 0 | 0 | 0 | 0 | 0 | 1 | 0 | 1 | 0 | 0 | |
| 1619 | F | 19-38 | 2141 | 1870 | 0 | 0 | 1303 | 824 | 0 | 0 | 0 | 0 | 0 | 0 | 0 | 0 | 0 | 0 | 0 | 0 | 0 | 0 | 0 | |
| 1620 | M | 39-82 | 1487 | 3304 | 0 | 0 | #N/A | #N/A | #N/A | #N/A | 1 | 0 | 1 | 0 | 0 | 0 | 0 | 0 | 0 | 0 | 1 | 0 | 0 | |
| 1621 | F | 19-38 | 2583 | 1395 | 0 | 0 | 2118 | 2975 | 0 | 0 | 1 | 0 | 0 | 1 | 0 | 0 | 0 | 0 | 0 | 0 | 0 | 1 | 0 | |
| 1622 | F | 19-38 | 1964 | 830 | 0 | 0 | 1683 | 804 | 0 | 0 | 1 | 0 | 1 | 0 | 0 | 0 | 0 | 0 | 0 | 0 | 0 | 0 | 0 | |
| 1623 | F | 19-38 | 1772 | 1600 | 0 | 0 | 1274 | 1058 | 0 | 0 | 0 | 0 | 0 | 0 | 0 | 0 | 0 | 0 | 0 | 0 | 0 | 0 | 0 | |
| 1624 | F | 39-82 | 735 | 528 | 0 | 0 | #N/A | #N/A | #N/A | #N/A | 1 | 1 | 1 | 1 | 1 | 1 | 1 | 0 | 0 | 0 | 1 | 0 | 0 | |
| 1625 | M | 19-38 | 2044 | 1362 | 0 | 0 | 2456 | 692 | 0 | 0 | 1 | 1 | 0 | 0 | 0 | 0 | 0 | 0 | 0 | 0 | 0 | 0 | 1 | 0 |
| 1626 | F | 19-38 | 1377 | 828 | 0 | 0 | 692 | 411 | 0 | 0 | 1 | 0 | 0 | 1 | 0 | 0 | 0 | 0 | 1 | 0 | 1 | 0 | 0 | |
| 1627 | M | 39-82 | 2488 | 3136 | 0 | 0 | 1605 | 2393 | 0 | 0 | 0 | 0 | 0 | 0 | 0 | 0 | 0 | 0 | 0 | 0 | 0 | 0 | 0 | |
| 1628 | F | 19-38 | 1523 | 1396 | 0 | 0 | #N/A | #N/A | #N/A | #N/A | 0 | 0 | 0 | 0 | 0 | 0 | 0 | 0 | 0 | 0 | 0 | 0 | 0 | |
| 1629 | F | 39-82 | 1631 | 2638 | 0 | 0 | 769 | 978 | 0 | 0 | 1 | 0 | 0 | 0 | 0 | 0 | 0 | 0 | 0 | 0 | 0 | 1 | 0 | |
| 1630 | F | 19-38 | 8773 | 1841 | 0 | 0 | 12895 | 1437 | 0 | 1 | 0 | 0 | 0 | 0 | 0 | 0 | 0 | 0 | 0 | 0 | 0 | 0 | 0 | |
| 1631 | H | 39-82 | 1725 | 1883 | 0 | 0 | 1208 | 1012 | 0 | 0 | 1 | 0 | 1 | 1 | 1 | 0 | 1 | 0 | 0 | 0 | 1 | 0 | 0 | |
| 1632 | F | 39-82 | 4738 | 960 | 0 | 0 | 4549 | 576 | 0 | 0 | 0 | 0 | 0 | 0 | 0 | 0 | 0 | 0 | 0 | 0 | 0 | 0 | 0 | |
| 1633 | F | 39-82 | 947 | 4108 | 0 | 0 | #N/A | #N/A | #N/A | #N/A | 0 | 0 | 0 | 0 | 0 | 0 | 0 | 0 | 0 | 0 | 0 | 0 | 0 | |
| 1634 | F | 19-38 | 913 | 1783 | 0 | 0 | 622 | 645 | 0 | 0 | 1 | 0 | 0 | 0 | 0 | 0 | 0 | 0 | 0 | 0 | 1 | 0 | 0 | |
| 1635 | M | 19-38 | 2297 | 1168 | 0 | 0 | 1423 | 434 | 0 | 0 | 1 | 0 | 1 | 1 | 0 | 0 | 0 | 0 | 1 | 0 | 1 | 0 | 0 | |
| 1636 | M | 39-82 | 2155 | 1199 | 0 | 0 | 1216 | 465 | 0 | 0 | 1 | 0 | 1 | 0 | 0 | 0 | 1 | 0 | 0 | 0 | 1 | 0 | 0 | |
| 1637 | F | 19-38 | 2663 | 1369 | 0 | 0 | #N/A | #N/A | #N/A | #N/A | 0 | 0 | 0 | 0 | 0 | 0 | 0 | 0 | 0 | 0 | 0 | 0 | 0 | |
| 1638 | M | 39-82 | 2689 | 4310 | 0 | 0 | #N/A | #N/A | #N/A | #N/A | 0 | 0 | 0 | 0 | 0 | 0 | 0 | 0 | 0 | 0 | 0 | 0 | 0 | |

| | | | | | | | | | | | | | | | | | | | | | | | | | |
|---|---|---|---|---|---|---|---|---|---|---|---|---|---|---|---|---|---|---|---|---|---|---|---|---|---|
| 1639 | F | 19-38 | | 4103 | 1121 | 0 | 0 | 2485 | 561 | 0 | 0 | 1 | 0 | 0 | 0 | 0 | 0 | 0 | 0 | 0 | 0 | 1 | 0 | 0 | |
| 1640 | F | 19-38 | negative | 1923 | 1178 | 0 | 0 | 855 | 480 | 0 | 0 | 1 | 0 | 1 | 1 | 0 | 0 | 1 | 0 | 0 | 1 | 1 | 0 | 0 | |
| 1641 | M | 39-82 | | 1270 | 1993 | 0 | 0 | 957 | 871 | 0 | 0 | 0 | 0 | 0 | 0 | 0 | 0 | 0 | 0 | 0 | 0 | 0 | 0 | 0 | |
| 1642 | F | 39-82 | | 1602 | 1130 | 0 | 0 | 627 | 548 | 0 | 0 | 1 | 0 | 1 | 0 | 1 | 1 | 1 | 0 | 0 | 0 | 1 | 1 | 0 | |
| 1643 | F | 19-38 | | 4371 | 1358 | 0 | 0 | #N/A | #N/A | #N/A | #N/A | 0 | 0 | 0 | 0 | 0 | 0 | 0 | 0 | 0 | 0 | 0 | 0 | 0 | |
| 1644 | H | 19-38 | | 2543 | 2233 | 0 | 0 | #N/A | #N/A | #N/A | #N/A | 0 | 0 | 0 | 0 | 0 | 0 | 0 | 0 | 0 | 0 | 0 | 0 | 0 | |
| 1645 | H | 19-38 | | 1449 | 1147 | 0 | 0 | #N/A | #N/A | #N/A | #N/A | 0 | 0 | 0 | 0 | 0 | 0 | 0 | 0 | 0 | 0 | 0 | 0 | 0 | |
| 1646 | H | 39-82 | | 1676 | 1250 | 0 | 0 | #N/A | #N/A | #N/A | #N/A | 1 | 0 | 1 | 0 | 0 | 0 | 0 | 0 | 0 | 0 | 0 | 0 | 0 | |
| 1647 | F | 19-38 | | 1775 | 1277 | 0 | 0 | 1186 | 902 | 0 | 0 | 0 | 0 | 0 | 0 | 0 | 0 | 0 | 0 | 0 | 0 | 0 | 0 | 0 | |
| 1648 | M | 19-38 | | 1109 | 1030 | 0 | 0 | #N/A | #N/A | #N/A | #N/A | 1 | 0 | 0 | 0 | 0 | 1 | 0 | 0 | 0 | 0 | 0 | 0 | 0 | |
| 1649 | M | 19-38 | | 1451 | 3003 | 0 | 0 | #N/A | #N/A | #N/A | #N/A | 1 | 0 | 0 | 0 | 0 | 0 | 0 | 0 | 0 | 0 | 1 | 0 | 0 | |
| 1650 | F | 19-38 | | 4330 | 1585 | 0 | 0 | #N/A | #N/A | #N/A | #N/A | 0 | 0 | 0 | 0 | 0 | 0 | 0 | 0 | 0 | 0 | 0 | 0 | 0 | |
| 1651 | M | 19-38 | | 5918 | 1305 | 0 | 0 | #N/A | #N/A | #N/A | #N/A | 1 | 0 | 1 | 0 | 0 | 0 | 1 | 0 | 0 | 0 | 1 | 0 | 0 | |
| 1652 | M | 19-38 | | 1759 | 2654 | 0 | 0 | #N/A | #N/A | #N/A | #N/A | 0 | 0 | 0 | 0 | 0 | 0 | 0 | 0 | 0 | 0 | 0 | 0 | 0 | |
| 1653 | F | 39-82 | | 1015 | 830 | 0 | 0 | 329 | 215 | 0 | 0 | 1 | 1 | 1 | 1 | 0 | 0 | 0 | 1 | 0 | 0 | 1 | 0 | 0 | |
| 1654 | F | 19-38 | | 3159 | 1417 | 0 | 0 | 2968 | 1013 | 0 | 0 | 0 | 0 | 0 | 0 | 0 | 0 | 0 | 0 | 0 | 0 | 0 | 0 | 0 | |
| 1655 | F | 39-82 | | 2355 | 1623 | 0 | 0 | 1558 | 786 | 0 | 0 | 1 | 0 | 0 | 0 | 0 | 0 | 0 | 0 | 1 | 0 | 1 | 0 | 0 | |
| 1656 | F | 39-82 | | 2164 | 1490 | 0 | 0 | 1244 | 485 | 0 | 0 | 1 | 1 | 0 | 1 | 0 | 0 | 0 | 0 | 1 | 0 | 0 | 0 | 0 | |
| 1657 | M | 19-38 | | 2140 | 3886 | 0 | 0 | 697 | 1402 | 0 | 0 | 0 | 0 | 0 | 0 | 0 | 0 | 0 | 0 | 0 | 0 | 0 | 0 | 0 | |
| 1658 | M | 19-38 | | 1704 | 1588 | 0 | 0 | 672 | 545 | 0 | 0 | 0 | 0 | 0 | 1 | 0 | 0 | 0 | 0 | 0 | 0 | 0 | 0 | 0 | |
| 1659 | M | 19-38 | | 6269 | 1277 | 0 | 0 | 2887 | 568 | 0 | 0 | 0 | 0 | 0 | 0 | 0 | 0 | 0 | 0 | 0 | 0 | 0 | 0 | 0 | |
| 1660 | F | 19-38 | | 1698 | 1273 | 0 | 0 | #N/A | #N/A | #N/A | #N/A | 0 | 0 | 0 | 0 | 0 | 0 | 0 | 0 | 0 | 0 | 0 | 0 | 0 | |
| 1661 | F | 19-38 | | 2602 | 3331 | 0 | 0 | #N/A | #N/A | #N/A | #N/A | 1 | 0 | 1 | 0 | 0 | 0 | 0 | 0 | 0 | 0 | 0 | 0 | 0 | |
| 1662 | F | 19-38 | | 16597 | 622 | 0 | 1 | 11847 | 1402 | 0 | 1 | 0 | 0 | 0 | 0 | 0 | 0 | 0 | 0 | 0 | 0 | 0 | 0 | 0 | |
| 1663 | F | 39-82 | | 8977 | 2865 | 0 | 0 | #N/A | #N/A | #N/A | #N/A | 1 | 0 | 1 | 0 | 0 | 0 | 0 | 0 | 0 | 0 | 1 | 0 | 1 | 0 |
| 1664 | F | 19-38 | | 1197 | 2543 | 0 | 0 | #N/A | #N/A | #N/A | #N/A | 1 | 0 | 0 | 0 | 0 | 0 | 1 | 0 | 0 | 0 | 1 | 0 | 0 | |
| 1665 | F | 19-38 | | 1429 | 873 | 0 | 0 | #N/A | #N/A | #N/A | #N/A | 1 | 1 | 1 | 1 | 1 | 0 | 0 | 0 | 0 | 0 | 0 | 0 | 0 | |
| 1666 | F | 19-38 | | 3348 | 1098 | 0 | 0 | #N/A | #N/A | #N/A | #N/A | 0 | 0 | 0 | 0 | 0 | 0 | 0 | 0 | 0 | 0 | 0 | 0 | 0 | |
| 1667 | F | 19-38 | | 1835 | 2152 | 0 | 0 | #N/A | #N/A | #N/A | #N/A | 0 | 0 | 0 | 0 | 0 | 0 | 0 | 0 | 0 | 0 | 0 | 0 | 1 | 0 |
| 1668 | F | 39-82 | | 1888 | 1658 | 0 | 0 | #N/A | #N/A | #N/A | #N/A | 1 | 0 | 1 | 0 | 1 | 0 | 1 | 0 | 0 | 0 | 1 | 0 | 0 | |
| 1669 | F | 39-82 | | 637 | 7531 | 0 | 0 | 528 | 3346 | 0 | 0 | 0 | 0 | 0 | 0 | 0 | 0 | 0 | 0 | 0 | 0 | 0 | 0 | 0 | |
| 1670 | F | 19-38 | | 1702 | 1349 | 0 | 0 | #N/A | #N/A | #N/A | #N/A | 0 | 0 | 0 | 0 | 0 | 0 | 0 | 0 | 0 | 0 | 0 | 0 | 0 | |
| 1671 | M | 19-38 | | 2193 | 1872 | 0 | 0 | 1451 | 1485 | 0 | 0 | 0 | 0 | 0 | 0 | 0 | 0 | 0 | 0 | 0 | 0 | 0 | 0 | 0 | |
| 1672 | M | 19-38 | | 1270 | 1677 | 0 | 0 | 977 | 934 | 0 | 0 | 1 | 0 | 1 | 0 | 0 | 0 | 1 | 0 | 0 | 0 | 1 | 1 | 0 | |
| 1673 | F | 39-82 | | 1577 | 703 | 0 | 0 | #N/A | #N/A | #N/A | #N/A | 1 | 0 | 0 | 0 | 0 | 0 | 0 | 0 | 0 | 0 | 1 | 0 | 0 | |
| 1674 | F | 39-82 | | 3659 | 744 | 0 | 0 | 4002 | 1029 | 0 | 0 | 0 | 0 | 0 | 0 | 0 | 0 | 0 | 0 | 0 | 0 | 0 | 0 | 0 | |
| 1675 | F | 39-82 | | 1144 | 759 | 0 | 0 | 1005 | 799 | 0 | 0 | 1 | 0 | 1 | 0 | 1 | 0 | 0 | 0 | 0 | 0 | 0 | 0 | 0 | |
| 1676 | F | 39-82 | | 209358 | 207754 | 98 | 1 | #N/A | #N/A | #N/A | #N/A | 1 | 0 | 1 | 0 | 1 | 1 | 0 | 0 | 0 | 1 | 1 | 1 | 1 | 0 |
| 1677 | F | 19-38 | | 3204 | 1022 | 0 | 0 | #N/A | #N/A | #N/A | #N/A | 1 | 1 | 0 | 0 | 0 | 0 | 0 | 0 | 0 | 0 | 0 | 0 | 1 | 0 |
| 1678 | H | 39-82 | | 3185 | 2338 | 0 | 0 | 1445 | 632 | 0 | 0 | 1 | 0 | 1 | 1 | 1 | 0 | 1 | 0 | 1 | 0 | 1 | 1 | 0 | |
| 1679 | F | 39-82 | | 1097 | 674 | 0 | 0 | #N/A | #N/A | #N/A | #N/A | 0 | 0 | 0 | 1 | 0 | 0 | 0 | 0 | 0 | 0 | 0 | 0 | 0 | |
| 1680 | F | 19-38 | | 2952 | 1460 | 0 | 0 | 2427 | 1078 | 0 | 0 | 0 | 0 | 0 | 0 | 0 | 0 | 0 | 0 | 0 | 0 | 0 | 0 | 0 | |
| 1681 | M | 19-38 | | 3179 | 1532 | 0 | 0 | #N/A | #N/A | #N/A | #N/A | 1 | 0 | 0 | 1 | 0 | 0 | 0 | 1 | 1 | 0 | 0 | 1 | 0 | |
| 1682 | F | 19-38 | | 2322 | 2398 | 0 | 0 | 1640 | 1065 | 0 | 0 | 1 | 0 | 1 | 1 | 0 | 0 | 0 | 0 | 1 | 0 | 1 | 0 | 0 | |
| 1683 | F | 19-38 | | 1286 | 2805 | 0 | 0 | #N/A | #N/A | #N/A | #N/A | 0 | 0 | 0 | 0 | 0 | 0 | 0 | 0 | 0 | 0 | 0 | 0 | 0 | |
| 1684 | H | 19-38 | | 2742 | 5027 | 0 | 0 | 1557 | 343 | 0 | 0 | 1 | 1 | 1 | 0 | 1 | 1 | 1 | 0 | 1 | 0 | 1 | 0 | 0 | |
| 1685 | H | 39-82 | | 1246 | 879 | 0 | 0 | #N/A | #N/A | #N/A | #N/A | 1 | 0 | 1 | 1 | 0 | 0 | 0 | 0 | 0 | 0 | 0 | 0 | 0 | |
| 1686 | H | 39-82 | | 1266 | 4286 | 0 | 0 | #N/A | #N/A | #N/A | #N/A | 1 | 1 | 1 | 1 | 0 | 0 | 0 | 0 | 0 | 0 | 0 | 0 | 0 | |
| 1687 | F | 19-38 | | 806 | 818 | 0 | 0 | 761 | 1226 | 0 | 0 | 0 | 0 | 0 | 0 | 0 | 0 | 0 | 0 | 0 | 0 | 0 | 0 | 0 | |
| 1688 | F | 39-82 | | 1230 | 3329 | 0 | 0 | 2709 | 1460 | 0 | 0 | 1 | 0 | 0 | 0 | 0 | 0 | 0 | 0 | 1 | 0 | 0 | 0 | 0 | |
| 1689 | F | 39-82 | | 5850 | 901 | 0 | 0 | 5558 | 1116 | 0 | 0 | 0 | 0 | 0 | 0 | 0 | 0 | 0 | 0 | 0 | 0 | 0 | 0 | 0 | |
| 1690 | F | 19-38 | | 1599 | 1314 | 0 | 0 | #N/A | #N/A | #N/A | #N/A | 0 | 0 | 0 | 0 | 0 | 0 | 0 | 0 | 0 | 0 | 0 | 0 | 0 | |
| 1691 | F | 19-38 | | 2969 | 6473 | 0 | 0 | #N/A | #N/A | #N/A | #N/A | 1 | 0 | 1 | 0 | 0 | 0 | 0 | 0 | 0 | 0 | 1 | 0 | 0 | |
| 1692 | M | 39-82 | | 2005 | 1150 | 0 | 0 | #N/A | #N/A | #N/A | #N/A | 1 | 0 | 1 | 0 | 0 | 0 | 0 | 0 | 1 | 0 | 1 | 1 | 0 | |
| 1693 | F | 19-38 | | 5495 | 2166 | 0 | 0 | 3351 | 585 | 0 | 0 | 0 | 0 | 0 | 0 | 0 | 0 | 0 | 0 | 0 | 0 | 0 | 0 | 0 | |
| 1694 | H | 39-82 | | 2025 | 991 | 0 | 0 | #N/A | #N/A | #N/A | #N/A | 1 | 0 | 1 | 1 | 0 | 1 | 0 | 0 | 1 | 0 | 1 | 0 | 0 | |
| 1695 | F | 39-82 | | 1639 | 6345 | 0 | 0 | #N/A | #N/A | #N/A | #N/A | 1 | 0 | 1 | 0 | 0 | 0 | 0 | 0 | 0 | 0 | 0 | 0 | 0 | |
| 1696 | H | 19-38 | | 2344 | 2037 | 0 | 0 | #N/A | #N/A | #N/A | #N/A | 1 | 1 | 1 | 0 | 0 | 0 | 1 | 0 | 0 | 0 | 0 | 0 | 0 | |
| 1697 | F | 19-38 | | 1896 | 6350 | 0 | 0 | #N/A | #N/A | #N/A | #N/A | 1 | 0 | 1 | 1 | 0 | 1 | 0 | 0 | 1 | 1 | 1 | 0 | 0 | |
| 1698 | F | 39-82 | | 2643 | 646 | 0 | 0 | #N/A | #N/A | #N/A | #N/A | 0 | 0 | 0 | 0 | 0 | 0 | 0 | 0 | 0 | 0 | 0 | 0 | 0 | |
| 1699 | F | 39-82 | | 1660 | 1118 | 0 | 0 | 1297 | 937 | 0 | 0 | 0 | 0 | 0 | 0 | 0 | 0 | 0 | 0 | 0 | 0 | 0 | 0 | 0 | |
| 1700 | F | 19-38 | | 3815 | 1248 | 0 | 0 | #N/A | #N/A | #N/A | #N/A | 1 | 0 | 1 | 0 | 1 | 0 | 0 | 0 | 0 | 0 | 1 | 1 | 0 | |
| 1701 | F | 39-82 | | 1977 | 690 | 0 | 0 | 1042 | 720 | 0 | 0 | 0 | 0 | 0 | 0 | 0 | 0 | 0 | 0 | 0 | 0 | 0 | 0 | 0 | |
| 1702 | F | 39-82 | | 2144 | 1114 | 0 | 0 | 929 | 823 | 0 | 0 | 0 | 0 | 0 | 0 | 0 | 0 | 0 | 0 | 0 | 0 | 0 | 0 | 0 | |
| 1703 | M | 39-82 | | 4548 | 1244 | 0 | 0 | 3349 | 891 | 0 | 0 | 1 | 1 | 0 | 1 | 0 | 0 | 0 | 0 | 1 | 0 | 0 | 0 | 0 | |
| 1704 | F | 39-82 | | 1091 | 1021 | 0 | 0 | 812 | 692 | 0 | 0 | 0 | 0 | 0 | 0 | 0 | 0 | 0 | 0 | 0 | 0 | 0 | 0 | 0 | |
| 1705 | F | 19-38 | | 3436 | 2515 | 5 | 1 | 2169 | 881 | 0 | 0 | 0 | 0 | 0 | 0 | 0 | 0 | 0 | 0 | 0 | 0 | 0 | 0 | 0 | |
| 1706 | F | 39-82 | | 2378 | 611 | 0 | 0 | 1544 | 4277 | 0 | 0 | 0 | 0 | 0 | 0 | 0 | 0 | 0 | 0 | 0 | 0 | 0 | 0 | 0 | |
| 1707 | F | 39-82 | | 9131 | 408 | 0 | 0 | 9074 | 763 | 0 | 0 | 0 | 0 | 0 | 0 | 0 | 0 | 0 | 0 | 0 | 0 | 0 | 0 | 0 | |
| 1708 | F | 19-38 | | 3032 | 3086 | 0 | 0 | 1520 | 746 | 0 | 0 | 1 | 0 | 0 | 1 | 0 | 1 | 0 | 0 | 1 | 0 | 1 | 0 | 0 | |
| 1709 | F | 19-38 | | 1561 | 949 | 0 | 0 | #N/A | #N/A | #N/A | #N/A | 0 | 0 | 0 | 0 | 0 | 0 | 0 | 0 | 0 | 0 | 0 | 0 | 0 | |
| 1710 | H | 39-82 | | 952 | 897 | 0 | 0 | 952 | 897 | 0 | 0 | 0 | 0 | 0 | 0 | 0 | 0 | 0 | 0 | 0 | 0 | 0 | 0 | 0 | |
| 1711 | F | 19-38 | | 1194 | 4126 | 0 | 0 | #N/A | #N/A | #N/A | #N/A | 1 | 0 | 1 | 1 | 0 | 0 | 0 | 0 | 1 | 0 | 1 | 0 | 0 | |
| 1712 | F | 19-38 | | 2512 | 847 | 0 | 0 | 2155 | 258 | 0 | 0 | 0 | 0 | 0 | 0 | 0 | 0 | 0 | 0 | 0 | 0 | 0 | 0 | 0 | |
| 1713 | F | 19-38 | | 94968 | 49050 | 75 | 1 | #N/A | #N/A | #N/A | #N/A | 1 | 1 | 1 | 1 | 0 | 0 | 1 | 0 | 0 | 1 | 1 | 1 | 1 | 1 |
| 1714 | F | 19-38 | | 512 | 3583 | 0 | 0 | #N/A | #N/A | #N/A | #N/A | 1 | 0 | 0 | 0 | 0 | 0 | 0 | 0 | 0 | 0 | 1 | 0 | 1 | 0 |
| 1715 | H | 19-38 | | 2981 | 1157 | 0 | 0 | 1463 | 1552 | 0 | 0 | 0 | 0 | 0 | 1 | 0 | 0 | 0 | 0 | 0 | 0 | 0 | 0 | 0 | |

| | | | | | | | | | | | | | | | | | | | | | | | | |
|---|---|---|---|---|---|---|---|---|---|---|---|---|---|---|---|---|---|---|---|---|---|---|---|---|
| 1716 | F | 39-82 | 1552 | 1107 | 0 | 0 | 1798 | 497 | 0 | 0 | 1 | 1 | 0 | 0 | 1 | 1 | 1 | 0 | 0 | 0 | 0 | 0 | 0 | |
| 1717 | F | 19-38 | 1555 | 644 | 0 | 0 | #N/A | #N/A | #N/A | #N/A | 0 | 0 | 0 | 0 | 0 | 0 | 0 | 0 | 0 | 0 | 0 | 0 | 0 | |
| 1718 | H | 19-38 | 1585 | 758 | 0 | 0 | #N/A | #N/A | #N/A | #N/A | 1 | 0 | 1 | 1 | 0 | 0 | 0 | 0 | 1 | 0 | 0 | 0 | 0 | |
| 1719 | F | 39-82 | 4847 | 687 | 0 | 0 | #N/A | #N/A | #N/A | #N/A | 1 | 0 | 0 | 0 | 0 | 0 | 0 | 0 | 0 | 0 | 1 | 0 | 0 | |
| 1720 | F | 39-82 | 1489 | 923 | 0 | 0 | #N/A | #N/A | #N/A | #N/A | 1 | 0 | 1 | 0 | 0 | 0 | 0 | 0 | 0 | 0 | 1 | 0 | 0 | |
| 1721 | F | 39-82 | 1407 | 2930 | 0 | 0 | #N/A | #N/A | #N/A | #N/A | 1 | 0 | 0 | 1 | 0 | 0 | 1 | 0 | 0 | 0 | 0 | 0 | 0 | |
| 1722 | F | 39-82 | 1926 | 1069 | 0 | 0 | 1645 | 909 | 0 | 0 | 1 | 0 | 0 | 0 | 0 | 0 | 0 | 0 | 0 | 0 | 1 | 0 | 0 | |
| 1723 | F | 19-38 | 2822 | 4121 | 0 | 0 | #N/A | #N/A | #N/A | #N/A | 0 | 0 | 0 | 0 | 0 | 0 | 0 | 0 | 0 | 0 | 0 | 0 | 0 | |
| 1724 | H | 39-82 | 3455 | 1093 | 0 | 0 | #N/A | #N/A | #N/A | #N/A | 1 | 0 | 1 | 1 | 1 | 1 | 0 | 0 | 0 | 0 | 0 | 0 | 0 | |
| 1725 | H | 19-38 | 1286 | 650 | 0 | 0 | 1128 | 201 | 0 | 0 | 1 | 1 | 1 | 1 | 1 | 1 | 1 | 0 | 1 | 1 | 1 | 1 | 0 | |
| 1726 | F | 19-38 | 2833 | 799 | 0 | 0 | #N/A | #N/A | #N/A | #N/A | 0 | 0 | 0 | 0 | 0 | 0 | 0 | 0 | 0 | 0 | 0 | 0 | 0 | |
| 1727 | F | 19-38 | 1102 | 1394 | 0 | 0 | #N/A | #N/A | #N/A | #N/A | 1 | 1 | 1 | 0 | 0 | 0 | 0 | 0 | 0 | 0 | 0 | 1 | 0 | |
| 1728 | F | 39-82 | 1372 | 821 | 0 | 0 | #N/A | #N/A | #N/A | #N/A | 1 | 0 | 1 | 0 | 0 | 1 | 0 | 0 | 0 | 0 | 0 | 0 | 0 | |
| 1729 | F | 39-82 | 814 | 602 | 0 | 0 | 816 | 598 | 0 | 0 | 0 | 0 | 0 | 0 | 0 | 0 | 0 | 0 | 0 | 0 | 0 | 0 | 0 | |
| 1730 | F | 19-38 | 1031 | 1200 | 0 | 0 | #N/A | #N/A | #N/A | #N/A | 1 | 0 | 0 | 0 | 0 | 1 | 0 | 0 | 0 | 0 | 0 | 0 | 0 | |
| 1731 | F | 19-38 | 15197 | 945 | 0 | 1 | 13166 | 966 | 0 | 1 | 0 | 0 | 0 | 0 | 0 | 0 | 0 | 0 | 0 | 0 | 0 | 0 | 0 | |
| 1732 | F | 39-82 | 1909 | 1130 | 0 | 0 | #N/A | #N/A | #N/A | #N/A | 1 | 0 | 1 | 0 | 0 | 0 | 1 | 0 | 0 | 0 | 1 | 1 | 0 | |
| 1733 | F | 19-38 | 1759 | 1858 | 0 | 0 | 1759 | 1858 | 0 | 0 | 1 | 0 | 1 | 1 | 0 | 0 | 0 | 0 | 0 | 0 | 1 | 0 | 0 | |
| 1734 | H | 19-38 | 2441 | 1365 | 0 | 0 | #N/A | #N/A | #N/A | #N/A | 1 | 0 | 1 | 0 | 1 | 0 | 0 | 0 | 0 | 0 | 0 | 0 | 0 | |
| 1735 | F | 39-82 | 2360 | 2634 | 0 | 0 | 21842 | 3911 | 50 | 1 | 0 | 0 | 0 | 0 | 0 | 0 | 0 | 0 | 0 | 0 | 0 | 0 | 0 | |
| 1736 | F | 39-82 | 1450 | 624 | 0 | 0 | #N/A | #N/A | #N/A | #N/A | 1 | 0 | 1 | 0 | 0 | 0 | 0 | 0 | 0 | 0 | 0 | 1 | 0 | |
| 1737 | F | 39-82 | 3719 | 792 | 0 | 0 | #N/A | #N/A | #N/A | #N/A | 1 | 0 | 1 | 0 | 0 | 0 | 0 | 0 | 0 | 0 | 0 | 0 | 0 | |
| 1738 | F | 19-38 | 1476 | 2185 | 0 | 0 | #N/A | #N/A | #N/A | #N/A | 0 | 0 | 0 | 0 | 0 | 0 | 0 | 0 | 0 | 0 | 0 | 0 | 0 | |
| 1739 | F | 39-82 | 735 | 361 | 0 | 0 | #N/A | #N/A | #N/A | #N/A | 1 | 1 | 1 | 0 | 0 | 0 | 0 | 0 | 0 | 0 | 1 | 1 | 0 | |
| 1740 | H | 19-38 | 2095 | 1152 | 0 | 0 | #N/A | #N/A | #N/A | #N/A | 1 | 0 | 0 | 0 | 0 | 0 | 0 | 0 | 0 | 0 | 0 | 1 | 0 | |
| 1741 | F | 39-82 | 4433 | 2395 | 0 | 0 | #N/A | #N/A | #N/A | #N/A | 0 | 0 | 0 | 0 | 0 | 0 | 0 | 0 | 0 | 0 | 0 | 0 | 0 | |
| 1742 | F | 39-82 | 2302 | 574 | 0 | 0 | #N/A | #N/A | #N/A | #N/A | 1 | 1 | 0 | 1 | 0 | 0 | 0 | 0 | 1 | 0 | 1 | 0 | 0 | |
| 1743 | F | 19-38 | 5071 | 2372 | 0 | 0 | #N/A | #N/A | #N/A | #N/A | 0 | 0 | 0 | 0 | 0 | 0 | 0 | 0 | 0 | 0 | 0 | 0 | 0 | |
| 1744 | F | 19-38 | 2458 | 5132 | 0 | 0 | #N/A | #N/A | #N/A | #N/A | 0 | 0 | 0 | 0 | 0 | 0 | 0 | 0 | 0 | 0 | 0 | 0 | 0 | |
| 1745 | F | 39-82 | 793 | 538 | 0 | 0 | 793 | 538 | 0 | 0 | 0 | 0 | 0 | 0 | 0 | 0 | 0 | 0 | 0 | 0 | 0 | 0 | 0 | |
| 1746 | F | 39-82 | 6162 | 1369 | 0 | 0 | 5197 | 573 | 0 | 0 | 0 | 0 | 0 | 0 | 0 | 0 | 0 | 0 | 0 | 0 | 0 | 0 | 0 | |
| 1747 | F | 19-38 | 918 | 654 | 0 | 0 | 654 | 453 | 0 | 0 | 0 | 0 | 0 | 0 | 0 | 0 | 0 | 0 | 0 | 0 | 0 | 0 | 0 | |
| 1748 | H | 39-82 | 880 | 909 | 2 | 1 | #N/A | #N/A | #N/A | #N/A | 0 | 0 | 0 | 0 | 0 | 0 | 0 | 0 | 0 | 0 | 0 | 0 | 0 | |
| 1749 | F | 19-38 | 6531 | 6851 | 0 | 0 | 3645 | 2466 | 0 | 0 | 0 | 0 | 0 | 0 | 0 | 0 | 0 | 0 | 0 | 0 | 0 | 0 | 0 | |
| 1750 | M | 19-38 | 2310 | 812 | 0 | 0 | 1741 | 1684 | 0 | 0 | 1 | 0 | 0 | 0 | 0 | 0 | 0 | 0 | 1 | 0 | 0 | 0 | 0 | |
| 1751 | F | 19-38 | 765 | 1219 | 0 | 0 | #N/A | #N/A | #N/A | #N/A | 0 | 0 | 0 | 0 | 0 | 0 | 0 | 0 | 0 | 0 | 0 | 0 | 0 | |
| 1752 | F | 39-82 | 1878 | 1009 | 0 | 0 | #N/A | #N/A | #N/A | #N/A | 1 | 0 | 1 | 0 | 0 | 0 | 0 | 1 | 0 | 0 | 0 | 0 | 0 | |
| 1753 | M | 19-38 | 2335 | 2636 | 0 | 0 | 1429 | 671 | 0 | 0 | 1 | 0 | 1 | 1 | 0 | 0 | 1 | 0 | 0 | 0 | 0 | 0 | 0 | |
| 1754 | M | 19-38 | 3161 | 2548 | 0 | 0 | #N/A | #N/A | #N/A | #N/A | 0 | 0 | 0 | 0 | 0 | 0 | 0 | 0 | 0 | 0 | 0 | 0 | 0 | |
| 1755 | F | 19-38 | 13314 | 19847 | 0 | 1 | 16825 | 1243 | 0 | 1 | 1 | 0 | 0 | 0 | 0 | 0 | 0 | 0 | 1 | 0 | 0 | 0 | 0 | |
| 1756 | F | 19-38 | 1261 | 772 | 0 | 0 | #N/A | #N/A | #N/A | #N/A | 0 | 0 | 0 | 0 | 0 | 0 | 0 | 0 | 0 | 0 | 0 | 0 | 0 | |
| 1757 | F | 19-38 | 1491 | 1366 | 0 | 0 | #N/A | #N/A | #N/A | #N/A | 0 | 0 | 0 | 0 | 0 | 0 | 0 | 0 | 0 | 0 | 0 | 0 | 0 | |
| 1758 | F | 19-38 | 5211 | 2211 | 0 | 0 | #N/A | #N/A | #N/A | #N/A | 0 | 0 | 0 | 0 | 0 | 0 | 0 | 0 | 0 | 0 | 0 | 0 | 0 | |
| 1759 | F | 19-38 | 3979 | 1075 | 0 | 0 | #N/A | #N/A | #N/A | #N/A | 1 | 0 | 0 | 0 | 0 | 0 | 0 | 0 | 0 | 1 | 1 | 1 | 0 | |
| 1761 | F | 39-82 | 1963 | 2325 | 0 | 0 | 2155 | 5597 | 0 | 0 | 0 | 0 | 0 | 1 | 0 | 0 | 0 | 0 | 0 | 0 | 0 | 0 | 0 | |
| 1762 | F | 19-38 | 2976 | 2366 | 60 | 1 | #N/A | #N/A | #N/A | #N/A | 0 | 0 | 0 | 0 | 0 | 0 | 0 | 0 | 0 | 0 | 0 | 0 | 0 | |
| 1763 | M | 19-38 | 19495 | 7734 | 0 | 1 | #N/A | #N/A | #N/A | #N/A | 1 | 0 | 1 | 1 | 1 | 1 | 0 | 0 | 1 | 0 | 1 | 1 | 0 | |
| 1764 | F | 39-82 | 3579 | 199 | 0 | 0 | 3593 | 876 | 0 | 0 | 1 | 1 | 0 | 0 | 0 | 0 | 0 | 0 | 1 | 0 | 0 | 0 | 0 | |
| 1765 | F | 19-38 | 2989 | 5090 | 0 | 0 | 1311 | 804 | 0 | 0 | 1 | 0 | 1 | 1 | 0 | 1 | 0 | 0 | 0 | 0 | 0 | 0 | 0 | |
| 1766 | F | 39-82 | 9640 | 940 | 0 | 0 | #N/A | #N/A | #N/A | #N/A | 1 | 0 | 1 | 0 | 0 | 0 | 0 | 0 | 0 | 0 | 0 | 0 | 0 | |
| 1767 | F | 19-38 | 963 | 808 | 23 | 1 | 564 | 1368 | 0 | 0 | 0 | 0 | 0 | 0 | 0 | 0 | 0 | 0 | 0 | 0 | 0 | 0 | 0 | |
| 1768 | F | 39-82 | 1824 | 629 | 0 | 0 | 2274 | 368 | 0 | 0 | 0 | 0 | 0 | 0 | 0 | 0 | 0 | 0 | 0 | 0 | 0 | 0 | 0 | |
| 1769 | F | 19-38 | 766 | 509 | 0 | 0 | #N/A | #N/A | #N/A | #N/A | 0 | 0 | 0 | 0 | 0 | 0 | 0 | 0 | 0 | 0 | 0 | 0 | 0 | |
| 1770 | H | 39-82 | 2655 | 2056 | 0 | 0 | #N/A | #N/A | #N/A | #N/A | 0 | 0 | 0 | 0 | 0 | 0 | 0 | 0 | 0 | 0 | 0 | 0 | 0 | |
| 1771 | F | 19-38 | 2204 | 607 | 8 | 1 | 1661 | 2331 | 0 | 0 | 1 | 0 | 0 | 1 | 0 | 0 | 0 | 1 | 1 | 0 | 0 | 0 | 0 | |
| 1772 | F | 19-38 | 1018 | 607 | 0 | 0 | 604 | 1342 | 0 | 0 | 0 | 0 | 0 | 0 | 0 | 0 | 0 | 0 | 0 | 0 | 0 | 0 | 0 | |
| 1773 | F | 39-82 | 1263 | 1388 | 0 | 0 | 911 | 796 | 0 | 0 | 0 | 0 | 0 | 0 | 0 | 0 | 0 | 0 | 0 | 0 | 0 | 0 | 0 | |
| 1774 | F | 19-38 | 969 | 771 | 21 | 1 | #N/A | #N/A | #N/A | #N/A | 0 | 0 | 0 | 0 | 0 | 0 | 0 | 0 | 0 | 0 | 0 | 0 | 0 | |
| 1775 | M | 39-82 | 944 | 748 | 0 | 0 | #N/A | #N/A | #N/A | #N/A | 1 | 0 | 0 | 0 | 0 | 0 | 0 | 0 | 0 | 0 | 1 | 0 | 0 | |
| 1776 | F | 19-38 | 1661 | 532 | 0 | 0 | #N/A | #N/A | #N/A | #N/A | 1 | 0 | 0 | 0 | 0 | 0 | 0 | 0 | 0 | 0 | 0 | 1 | 1 | 0 |
| 1777 | M | 39-82 | 30080 | 61882 | 0 | 1 | 28467 | 16522 | 0 | 1 | 0 | 0 | 0 | 0 | 0 | 0 | 0 | 0 | 0 | 0 | 0 | 0 | 0 | |
| 1778 | F | 19-38 | 2373 | 2732 | 0 | 0 | #N/A | #N/A | #N/A | #N/A | 0 | 0 | 0 | 0 | 0 | 0 | 0 | 0 | 0 | 0 | 0 | 0 | 0 | |
| 1779 | F | 39-82 | 1664 | 1550 | 0 | 0 | #N/A | #N/A | #N/A | #N/A | 0 | 0 | 0 | 1 | 0 | 0 | 0 | 0 | 0 | 0 | 0 | 0 | 0 | |
| 1781 | F | 19-38 | 950 | 1675 | 0 | 0 | #N/A | #N/A | #N/A | #N/A | 1 | 0 | 1 | 0 | 0 | 0 | 0 | 0 | 0 | 0 | 0 | 0 | 0 | |
| 1782 | M | 39-82 | 2782 | 524 | 0 | 0 | 2929 | 400 | 0 | 0 | 1 | 0 | 1 | 0 | 0 | 0 | 0 | 0 | 0 | 0 | 0 | 0 | 1 | 0 |
| 1783 | F | 39-82 | 1717 | 1330 | 0 | 0 | #N/A | #N/A | #N/A | #N/A | 1 | 0 | 1 | 1 | 1 | 0 | 0 | 0 | 1 | 0 | 0 | 1 | 0 | |
| 1784 | F | 19-38 | 1857 | 1199 | 0 | 0 | 2107 | 1030 | 0 | 0 | 0 | 0 | 0 | 0 | 0 | 0 | 0 | 0 | 0 | 0 | 0 | 0 | 0 | |
| 1785 | F | 39-82 | 793 | 638 | 0 | 0 | #N/A | #N/A | #N/A | #N/A | 1 | 0 | 0 | 0 | 0 | 0 | 0 | 0 | 0 | 0 | 1 | 0 | 0 | |
| 1786 | F | 19-38 | 842 | 508 | 4 | 1 | #N/A | #N/A | #N/A | #N/A | 0 | 0 | 0 | 0 | 0 | 0 | 0 | 0 | 0 | 0 | 0 | 0 | 0 | |
| 1787 | F | 19-38 | 2102 | 2044 | 0 | 0 | #N/A | #N/A | #N/A | #N/A | 1 | 0 | 0 | 0 | 0 | 0 | 0 | 0 | 1 | 0 | 1 | 1 | 0 | |
| 1788 | F | 39-82 | 893 | 1218 | 0 | 0 | 1079 | 1386 | 0 | 0 | 1 | 0 | 1 | 0 | 0 | 0 | 0 | 0 | 0 | 0 | 1 | 0 | 0 | |
| 1789 | F | 19-38 | 6249 | 1006 | 0 | 0 | #N/A | #N/A | #N/A | #N/A | 0 | 0 | 0 | 0 | 0 | 0 | 0 | 0 | 0 | 0 | 0 | 0 | 0 | |
| 1790 | F | 19-38 | 2080 | 2651 | 0 | 0 | 1141 | 605 | 0 | 0 | 0 | 0 | 0 | 0 | 0 | 0 | 0 | 0 | 0 | 0 | 0 | 0 | 0 | |
| 1791 | F | 39-82 | 1165 | 1325 | 0 | 0 | #N/A | #N/A | #N/A | #N/A | 0 | 0 | 0 | 0 | 0 | 0 | 0 | 0 | 0 | 0 | 0 | 0 | 0 | |
| 1792 | F | 19-38 | 1885 | 1290 | 0 | 0 | #N/A | #N/A | #N/A | #N/A | 1 | 1 | 1 | 1 | 1 | 1 | 0 | 0 | 0 | 1 | 0 | 1 | 1 | 0 |
| 1793 | F | 19-38 | 2831 | 2247 | 0 | 0 | 2332 | 2609 | 0 | 0 | 0 | 0 | 0 | 0 | 0 | 0 | 0 | 0 | 0 | 0 | 0 | 0 | 0 | |
| 1794 | F | 39-82 | 341 | 911 | 0 | 0 | #N/A | #N/A | #N/A | #N/A | 0 | 0 | 0 | 0 | 0 | 0 | 0 | 0 | 0 | 0 | 0 | 0 | 0 | |

| | | | | | | | | | | | | | | | | | | | | | | | | | |
|---|---|---|---|---|---|---|---|---|---|---|---|---|---|---|---|---|---|---|---|---|---|---|---|---|---|
| 1795 | F | | 39-82 | 4603 | 1516 | 0 | 0 | 3721 | 1119 | 0 | 0 | 0 | 0 | 0 | 0 | 0 | 0 | 0 | 0 | 0 | 0 | 0 | 0 | 0 | |
| 1796 | F | | 19-38 | 1947 | 1254 | 0 | 0 | 973 | 921 | 0 | 0 | 0 | 0 | 0 | 0 | 0 | 0 | 0 | 0 | 0 | 0 | 0 | 0 | 0 | |
| 1798 | F | | 19-38 | 1909 | 1119 | 0 | 0 | #N/A | #N/A | #N/A | #N/A | 0 | 0 | 0 | 0 | 0 | 0 | 0 | 0 | 0 | 0 | 0 | 0 | 0 | |
| 1799 | M | | 39-82 | 1462 | 1366 | 0 | 0 | #N/A | #N/A | #N/A | #N/A | 0 | 0 | 0 | 0 | 0 | 0 | 0 | 0 | 0 | 0 | 0 | 0 | 0 | |
| 1800 | M | | 39-82 | 4152 | 1374 | 0 | 0 | 3320 | 3867 | 0 | 0 | 0 | 0 | 0 | 0 | 0 | 0 | 0 | 0 | 0 | 0 | 0 | 0 | 0 | |
| 1801 | F | | 39-82 | 965 | 584 | 0 | 0 | #N/A | #N/A | #N/A | #N/A | 0 | 0 | 0 | 0 | 0 | 0 | 0 | 0 | 0 | 0 | 0 | 0 | 0 | |
| 1802 | F | | 19-38 | 5572 | 927 | 0 | 0 | #N/A | #N/A | #N/A | #N/A | 1 | 1 | 1 | 0 | 1 | 1 | 1 | 0 | 0 | 0 | 1 | 0 | 0 | |
| 1803 | F | | 19-38 | 4545 | 2424 | 0 | 0 | 1302 | 2038 | 0 | 0 | 0 | 0 | 0 | 0 | 0 | 0 | 0 | 0 | 0 | 0 | 0 | 0 | 0 | |
| 1804 | M | | 19-38 | 1452 | 819 | 0 | 0 | #N/A | #N/A | #N/A | #N/A | 0 | 0 | 0 | 0 | 0 | 0 | 0 | 0 | 0 | 0 | 0 | 0 | 0 | |
| 1805 | F | | 39-82 | 3579 | 610 | 0 | 0 | #N/A | #N/A | #N/A | #N/A | 1 | 1 | 0 | 1 | 0 | 1 | 0 | 0 | 0 | 0 | 1 | 0 | 1 | 0 |
| 1806 | F | | 39-82 | 1858 | 780 | 0 | 0 | #N/A | #N/A | #N/A | #N/A | 0 | 0 | 0 | 0 | 0 | 0 | 0 | 0 | 0 | 0 | 0 | 0 | 0 | |
| 1807 | F | | 19-38 | 2358 | 1076 | 0 | 0 | #N/A | #N/A | #N/A | #N/A | 0 | 0 | 0 | 0 | 0 | 0 | 0 | 0 | 0 | 0 | 0 | 0 | 0 | |
| 1808 | F | | 19-38 | 1201 | 863 | 0 | 0 | #N/A | #N/A | #N/A | #N/A | 1 | 0 | 0 | 0 | 0 | 0 | 0 | 0 | 1 | 0 | 0 | 0 | 0 | |
| 1809 | M | | 39-82 | 1079 | 834 | 0 | 0 | #N/A | #N/A | #N/A | #N/A | 0 | 0 | 0 | 0 | 0 | 0 | 0 | 0 | 0 | 0 | 0 | 0 | 0 | |
| 1810 | F | | 39-82 | 1905 | 674 | 0 | 0 | #N/A | #N/A | #N/A | #N/A | 0 | 0 | 0 | 0 | 0 | 0 | 0 | 0 | 0 | 0 | 0 | 0 | 0 | |
| 1811 | F | | 39-82 | 20343 | 88142 | 78 | 1 | #N/A | #N/A | #N/A | #N/A | 0 | 0 | 0 | 0 | 0 | 0 | 0 | 0 | 0 | 0 | 0 | 0 | 0 | |
| 1813 | F | | 39-82 | 1442 | 1279 | 0 | 0 | #N/A | #N/A | #N/A | #N/A | 0 | 0 | 0 | 0 | 0 | 0 | 0 | 0 | 0 | 0 | 0 | 0 | 0 | |
| 1814 | F | | 19-38 | 5055 | 2051 | 0 | 0 | #N/A | #N/A | #N/A | #N/A | 1 | 0 | 1 | 1 | 0 | 1 | 0 | 0 | 1 | 0 | 1 | 0 | 0 | |
| 1815 | F | | 39-82 | 3248 | 695 | 0 | 0 | #N/A | #N/A | #N/A | #N/A | 0 | 0 | 0 | 0 | 0 | 0 | 0 | 0 | 0 | 0 | 0 | 0 | 0 | |
| 1816 | F | | 19-38 | 1333 | 1757 | 0 | 0 | #N/A | #N/A | #N/A | #N/A | 1 | 1 | 1 | 1 | 0 | 0 | 1 | 0 | 1 | 0 | 1 | 1 | 0 | |
| 1817 | F | | 19-38 | 8275 | 1945 | 0 | 0 | #N/A | #N/A | #N/A | #N/A | 0 | 0 | 0 | 0 | 0 | 0 | 0 | 0 | 0 | 0 | 0 | 0 | 0 | |
| 1818 | M | | 19-38 | 2256 | 615 | 0 | 0 | #N/A | #N/A | #N/A | #N/A | 0 | 0 | 0 | 0 | 0 | 0 | 0 | 0 | 0 | 0 | 0 | 0 | 0 | |
| 1819 | F | | 39-82 | 2332 | 779 | 0 | 0 | #N/A | #N/A | #N/A | #N/A | 1 | 0 | 1 | 1 | 1 | 1 | 0 | 0 | 1 | 0 | 1 | 0 | 0 | |
| 1821 | M | | 19-38 | 3505 | 1009 | 0 | 0 | 2710 | 1023 | 0 | 0 | 1 | 0 | 0 | 0 | 0 | 0 | 0 | 0 | 0 | 0 | 1 | 0 | 0 | |
| 1822 | F | | 39-82 | 2857 | 562 | 0 | 0 | 2055 | 828 | 0 | 0 | 1 | 0 | 1 | 0 | 0 | 0 | 0 | 0 | 0 | 0 | 1 | 0 | 0 | |
| 1823 | M | | 39-82 | 2455 | 1336 | 0 | 0 | 875 | 850 | 0 | 0 | 0 | 0 | 0 | 0 | 0 | 0 | 0 | 0 | 0 | 0 | 0 | 0 | 0 | |
| 1824 | F | | 19-38 | 5219 | 2054 | 0 | 0 | 1791 | 1219 | 0 | 0 | 1 | 1 | 1 | 1 | 0 | 0 | 1 | 0 | 1 | 1 | 1 | 0 | 0 | |
| 1825 | M | | 19-38 | 1878 | 1346 | 0 | 0 | 1296 | 809 | 0 | 0 | 0 | 0 | 0 | 0 | 0 | 0 | 0 | 0 | 0 | 0 | 0 | 0 | 0 | |
| 1826 | M | | 19-38 | 39738 | 42636 | 66 | 1 | #N/A | #N/A | #N/A | #N/A | 0 | 0 | 0 | 0 | 0 | 0 | 0 | 0 | 0 | 0 | 0 | 0 | 0 | |
| 1827 | F | | 39-82 | 1536 | 743 | 0 | 0 | 335 | 598 | 0 | 0 | 0 | 0 | 0 | 0 | 0 | 0 | 0 | 0 | 0 | 0 | 0 | 0 | 0 | |
| 1828 | F | | 39-82 | 2036 | 618 | 0 | 0 | 1201 | 359 | 0 | 0 | 0 | 0 | 0 | 0 | 0 | 0 | 0 | 0 | 0 | 0 | 0 | 0 | 0 | |
| 1829 | F | | 19-38 | 1635 | 849 | 0 | 0 | 2902 | 645 | 0 | 0 | 1 | 0 | 1 | 0 | 0 | 1 | 0 | 0 | 0 | 0 | 0 | 0 | 0 | |
| 1830 | F | | 19-38 | 2410 | 3918 | 0 | 0 | #N/A | #N/A | #N/A | #N/A | 0 | 0 | 0 | 0 | 0 | 0 | 0 | 0 | 0 | 0 | 0 | 0 | 0 | |
| 1831 | F | | 39-82 | 26610 | 38755 | 42 | 1 | 11298 | 22681 | 71 | 1 | 0 | 0 | 0 | 0 | 0 | 0 | 0 | 0 | 0 | 0 | 0 | 0 | 0 | |
| 1832 | F | | 39-82 | 1529 | 1087 | 0 | 0 | 1141 | 955 | 0 | 0 | 0 | 0 | 0 | 0 | 0 | 0 | 0 | 0 | 0 | 0 | 0 | 0 | 0 | |
| 1833 | F | | 19-38 | 2005 | 979 | 0 | 0 | #N/A | #N/A | #N/A | #N/A | 0 | 0 | 0 | 0 | 0 | 0 | 0 | 0 | 0 | 0 | 0 | 0 | 0 | |
| 1834 | F | | 19-38 | 1763 | 2414 | 0 | 0 | #N/A | #N/A | #N/A | #N/A | 0 | 0 | 0 | 0 | 0 | 0 | 0 | 0 | 0 | 0 | 0 | 0 | 0 | |
| 1835 | F | | 39-82 | 605 | 378 | 0 | 0 | #N/A | #N/A | #N/A | #N/A | 1 | 0 | 1 | 1 | 0 | 0 | 0 | 0 | 0 | 0 | 0 | 0 | 0 | |
| 1836 | F | | 39-82 | 1179 | 1580 | 0 | 0 | #N/A | #N/A | #N/A | #N/A | 0 | 0 | 0 | 0 | 0 | 0 | 0 | 0 | 0 | 0 | 0 | 0 | 0 | |
| 1837 | F | | 39-82 | 115579 | 95007 | 89 | 1 | 77282 | 100489 | 98 | 1 | 1 | 1 | 1 | 1 | 1 | 1 | 1 | 1 | 1 | 0 | 1 | 1 | 1 | 1 |
| 1838 | M | | 19-38 | 2089 | 2757 | 0 | 0 | #N/A | #N/A | #N/A | #N/A | 1 | 1 | 1 | 1 | 1 | 1 | 1 | 0 | 0 | 0 | 1 | 0 | 1 | 0 |
| 1839 | M | | 39-82 | 52576 | 72075 | 91 | 1 | 28948 | 46858 | 88 | 1 | 1 | 1 | 1 | 1 | 1 | 0 | 1 | 0 | 1 | 0 | 1 | 0 | 0 | |
| 1840 | F | | 19-38 | 1716 | 908 | 0 | 0 | 755 | 501 | 0 | 0 | 0 | 0 | 0 | 0 | 0 | 0 | 0 | 0 | 0 | 0 | 0 | 0 | 0 | |
| 1841 | F | | 39-82 | 1028 | 844 | 0 | 0 | 334 | 1013 | 0 | 0 | 1 | 1 | 1 | 1 | 1 | 1 | 0 | 0 | 0 | 0 | 0 | 0 | 0 | |
| 1842 | M | | 19-38 | 8566 | 708 | 0 | 0 | #N/A | #N/A | #N/A | #N/A | 0 | 0 | 0 | 0 | 0 | 0 | 0 | 0 | 0 | 0 | 0 | 0 | 0 | |
| 1843 | F | | 19-38 | 2036 | 1588 | 0 | 0 | #N/A | #N/A | #N/A | #N/A | 1 | 1 | 1 | 0 | 0 | 0 | 1 | 0 | 1 | 0 | 1 | 0 | 0 | |
| 1844 | M | | 19-38 | 3084 | 764 | 0 | 0 | #N/A | #N/A | #N/A | #N/A | 1 | 0 | 1 | 0 | 0 | 0 | 1 | 0 | 0 | 0 | 1 | 1 | 0 | |
| 1845 | F | | 39-82 | 1492 | 711 | 0 | 0 | #N/A | #N/A | #N/A | #N/A | 1 | 0 | 1 | 0 | 0 | 0 | 0 | 0 | 0 | 0 | 1 | 0 | 0 | |
| 1846 | M | | 19-38 | 2026 | 1519 | 0 | 0 | #N/A | #N/A | #N/A | #N/A | 1 | 1 | 1 | 1 | 0 | 0 | 0 | 0 | 0 | 0 | 1 | 1 | 0 | |
| 1847 | F | | 39-82 | 3571 | 1887 | 0 | 0 | #N/A | #N/A | #N/A | #N/A | 1 | 1 | 0 | 0 | 0 | 1 | 1 | 0 | 0 | 0 | 1 | 1 | 0 | |
| 1848 | M | | 19-38 | 1682 | 946 | 0 | 0 | #N/A | #N/A | #N/A | #N/A | 0 | 0 | 0 | 0 | 0 | 0 | 0 | 0 | 0 | 0 | 0 | 0 | 0 | |
| 1849 | F | | 39-82 | 64923 | 51790 | 75 | 1 | #N/A | #N/A | #N/A | #N/A | 0 | 0 | 0 | 0 | 0 | 0 | 0 | 0 | 0 | 0 | 0 | 0 | 0 | |
| 1850 | F | | 19-38 | 1399 | 1222 | 0 | 0 | #N/A | #N/A | #N/A | #N/A | 0 | 0 | 0 | 0 | 0 | 0 | 0 | 0 | 0 | 0 | 0 | 0 | 0 | |
| 1852 | F | | 19-38 | 1455 | 874 | 0 | 0 | #N/A | #N/A | #N/A | #N/A | 0 | 0 | 0 | 0 | 0 | 0 | 0 | 0 | 0 | 0 | 0 | 0 | 0 | |
| 1853 | F | | 19-38 | 2899 | 902 | 0 | 0 | #N/A | #N/A | #N/A | #N/A | 0 | 0 | 0 | 0 | 0 | 0 | 0 | 0 | 0 | 0 | 0 | 0 | 0 | |
| 1854 | | #N/A | 19-38 | 2524 | 1485 | 0 | 0 | #N/A | #N/A | #N/A | #N/A | 0 | 0 | 0 | 0 | 0 | 0 | 0 | 0 | 0 | 0 | 0 | 0 | 0 | |
| 1857 | M | | 19-38 | 1663 | 908 | 0 | 0 | #N/A | #N/A | #N/A | #N/A | 0 | 0 | 0 | 0 | 0 | 0 | 0 | 0 | 0 | 0 | 0 | 0 | 0 | |
| 1858 | f | | 39-82 | 1555 | 825 | 0 | 0 | #N/A | #N/A | #N/A | #N/A | 1 | 1 | 1 | 1 | 1 | 0 | 1 | 0 | 1 | 0 | 1 | 0 | 0 | |
| 1859 | F | | 39-82 | 2312 | 1596 | 0 | 0 | 1671 | 917 | 0 | 0 | 0 | 0 | 0 | 0 | 0 | 0 | 0 | 0 | 0 | 0 | 0 | 0 | 0 | |
| 1860 | f | | 19-38 | 1774 | 796 | 0 | 0 | #N/A | #N/A | #N/A | #N/A | 0 | 0 | 0 | 0 | 0 | 0 | 0 | 0 | 0 | 0 | 0 | 0 | 0 | |
| 1861 | F | | 39-82 | 2422 | 760 | 0 | 0 | 1513 | 1140 | 0 | 0 | 0 | 0 | 0 | 0 | 0 | 0 | 0 | 0 | 0 | 0 | 0 | 0 | 0 | |
| 1862 | F | | 19-38 | 2897 | 1000 | 0 | 0 | 1363 | 2177 | 0 | 0 | 0 | 0 | 0 | 0 | 0 | 0 | 0 | 0 | 0 | 0 | 0 | 0 | 0 | |
| 1863 | F | | 19-38 | 729 | 576 | 0 | 0 | 693 | 438 | 0 | 0 | 0 | 0 | 0 | 0 | 0 | 0 | 0 | 0 | 0 | 0 | 0 | 0 | 1 | 0 |
| 1864 | F | | 39-82 | 1220 | 1714 | 0 | 0 | #N/A | #N/A | #N/A | #N/A | 1 | 0 | 0 | 0 | 0 | 0 | 0 | 0 | 0 | 0 | 1 | 0 | 0 | |
| 1865 | M | | 19-38 | 4946 | 902 | 0 | 0 | 2965 | 1391 | 0 | 0 | 1 | 1 | 1 | 0 | 0 | 0 | 1 | 0 | 0 | 1 | 0 | 0 | 0 | |
| 1866 | F | | 19-38 | 2460 | 1598 | 0 | 0 | #N/A | #N/A | #N/A | #N/A | 0 | 0 | 0 | 0 | 0 | 0 | 0 | 0 | 0 | 0 | 0 | 0 | 0 | |
| 1867 | F | | 39-82 | 4163 | 1696 | 0 | 0 | #N/A | #N/A | #N/A | #N/A | 0 | 0 | 0 | 0 | 0 | 0 | 0 | 0 | 0 | 0 | 0 | 0 | 1 | |
| 1868 | F | | 19-38 | 5881 | 2070 | 0 | 0 | 6291 | 716 | 0 | 0 | 0 | 0 | 0 | 0 | 0 | 0 | 0 | 0 | 0 | 0 | 0 | 0 | 0 | |
| 1869 | F | | 19-38 | 2463 | 5442 | 0 | 0 | #N/A | #N/A | #N/A | #N/A | 0 | 0 | 0 | 0 | 0 | 0 | 0 | 0 | 0 | 0 | 0 | 0 | 0 | |
| 1870 | F | | 19-38 | 1805 | 735 | 0 | 0 | 1503 | 988 | 0 | 0 | 0 | 0 | 0 | 0 | 0 | 0 | 0 | 0 | 0 | 0 | 0 | 0 | 0 | |
| 1871 | M | | 39-82 | 1552 | 3253 | 0 | 0 | #N/A | #N/A | #N/A | #N/A | 0 | 0 | 0 | 0 | 0 | 0 | 0 | 0 | 0 | 0 | 0 | 0 | 0 | |
| 1872 | F | | 39-82 | 346 | 422 | 0 | 0 | 216 | 535 | 0 | 0 | 0 | 0 | 0 | 0 | 0 | 0 | 0 | 0 | 0 | 0 | 0 | 0 | 0 | |
| 1873 | M | | 19-38 | 4139 | 1111 | 0 | 0 | 4933 | 708 | 0 | 0 | 0 | 0 | 0 | 0 | 0 | 0 | 0 | 0 | 0 | 0 | 0 | 0 | 0 | |
| 1874 | M | | 19-38 | 540 | 605 | 0 | 0 | #N/A | #N/A | #N/A | #N/A | 0 | 0 | 0 | 0 | 0 | 0 | 0 | 0 | 0 | 0 | 0 | 0 | 0 | |
| 1875 | F | | 39-82 | 63956 | 145562 | 40 | 1 | 19125 | 187957 | 97 | 1 | 0 | 0 | 0 | 0 | 0 | 0 | 0 | 0 | 0 | 0 | 0 | 0 | 1 | 1 |
| 1876 | M | | 39-82 | 1609 | 2048 | 0 | 0 | #N/A | #N/A | #N/A | #N/A | 0 | 0 | 0 | 0 | 0 | 0 | 0 | 0 | 0 | 0 | 0 | 0 | 0 | |
| 1877 | F | | 39-82 | 5450 | 825 | 0 | 0 | #N/A | #N/A | #N/A | #N/A | 0 | 0 | 0 | 0 | 0 | 0 | 0 | 0 | 0 | 0 | 0 | 0 | 0 | |

| | | | | | | | | | | | | | | | | | | | | | | | |
|---|---|---|---|---|---|---|---|---|---|---|---|---|---|---|---|---|---|---|---|---|---|---|---|
| 1878 | F | 19-38 | 1806 | 577 | 0 | 0 | #N/A | #N/A | #N/A | #N/A | 0 | 0 | 0 | 0 | 0 | 0 | 0 | 0 | 0 | 0 | 0 | 0 | 0 |
| 1879 | F | 39-82 | 1064 | 348 | 0 | 0 | #N/A | #N/A | #N/A | #N/A | 0 | 0 | 0 | 0 | 0 | 0 | 0 | 0 | 0 | 0 | 0 | 0 | 0 |
| 1880 | M | 39-82 | 895 | 429 | 0 | 0 | #N/A | #N/A | #N/A | #N/A | 0 | 0 | 0 | 0 | 0 | 0 | 0 | 0 | 0 | 0 | 0 | 0 | 0 |
| 1881 | M | 19-38 | 923 | 306 | 0 | 0 | #N/A | #N/A | #N/A | #N/A | 0 | 0 | 0 | 0 | 0 | 0 | 0 | 0 | 0 | 0 | 0 | 0 | 0 |
| 1882 | M | 39-82 | 860 | 392 | 0 | 0 | #N/A | #N/A | #N/A | #N/A | 0 | 0 | 0 | 0 | 0 | 0 | 0 | 0 | 0 | 0 | 0 | 0 | 0 |
| 1883 | F | 39-82 | 756 | 1102 | 0 | 0 | #N/A | #N/A | #N/A | #N/A | 0 | 0 | 0 | 0 | 0 | 0 | 0 | 0 | 0 | 0 | 0 | 0 | 0 |
| 1884 | F | 39-82 | 1154 | 413 | 0 | 0 | #N/A | #N/A | #N/A | #N/A | 0 | 0 | 0 | 0 | 0 | 0 | 0 | 0 | 0 | 0 | 0 | 0 | 0 |
| 1885 | F | 19-38 | 1195 | 1338 | 0 | 0 | #N/A | #N/A | #N/A | #N/A | 1 | 0 | 1 | 0 | 0 | 0 | 0 | 0 | 0 | 0 | 1 | 0 | 0 |

0
